# The Yale Department of Medicine COVID-19 Data Explorer and Repository (DOM-CovX): An Innovative Approach to Promoting Collaborative Scholarship During a Pandemic

**DOI:** 10.1101/2021.08.23.21262241

**Authors:** Tanima Arora, Michael Simonov, Jameel Alausa, Labeebah Subair, Brett Gerber, Andrew Nguyen, Allen Hsiao, Richard Hintz, Yu Yamamoto, Robert Soufer, Gary Desir, Francis Perry Wilson, Merceditas Villanueva

## Abstract

**Background:** The COVID-19 pandemic has led to an explosion of research publications spanning epidemiology, basic and clinical science. While a digital revolution has allowed for open access to large datasets enabling real-time tracking of the epidemic, detailed, locally-specific clinical data has been less readily accessible to a broad range of academic faculty and their trainees. This perpetuates the separation of the primary missions of clinically-focused and primary research faculty resulting in lost opportunities for improved understanding of the local epidemic; expansion of the scope of scholarship; limitation of the diversity of the research pool; lack of creation of initiatives for growth and dissemination of research skills needed for the training of the next generation of clinicians and faculty.

**Objectives:** Create a common, easily accessible and up-to-date database that would promote access to local COVID-19 clinical data, thereby increasing efficiency, streamlining and democratizing the research enterprise. By providing a robust dataset, a broad range of researchers (faculty, trainees) and clinicians are encouraged to explore and collaborate on novel clinically relevant research questions.

**Methods:** We constructed a research platform called the Yale Department of Medicine COVID-19 Explorer and Repository (DOM-CovX), to house cleaned, highly granular, de-identified, continually-updated data from over 7,000 patients hospitalized with COVID-19 (1/2020-present) across the Yale New Haven Health System. This included a front-end user interface for simple data visualization of aggregate data and more detailed clinical datasets for researchers after a review board process. The goal is to promote access to local COVID-19 clinical data, thereby increasing efficiency, streamlining and democratizing the research enterprise.

**Expected Outcomes:**

1. Accelerate generation of new knowledge and increase scholarly productivity with particular local relevance
2. Improve the institutional academic climate by:
  a. Broadening research scope
  b. Expanding research capability to more diverse group of stakeholders including clinical and research-based faculty and trainees
  c. Enhancing interdepartmental collaborations

**Conclusions:** The DOM-CovX Data Explorer and Repository have great potential to increase academic productivity. By providing an accessible tool for simple data analysis and access to a consistently updated, standardized and large-scale dataset, it overcomes barriers for a wide variety of researchers. Beyond academic productivity, this innovative approach represents an opportunity to improve the institutional climate by fostering collaboration, diversity of scholarly pursuits and expanding medical education. It provides a novel approach that can be expanded to other diseases beyond COVID 19.

## INTRODUCTION

Coronavirus disease 2019 (COVID-19), the disease caused by the Severe Acute Respiratory Syndrome Coronavirus 2(SARS-CoV-2), emerged as a global pandemic that has challenged a broad range of researchers to rapidly develop a new understanding of epidemiology, basic and clinical science to implement novel approaches for prevention and care. There has been an explosion of articles both in the pre-print and peer-reviewed literature as evidenced by over 35,000 PubMed indexed publications within the first 6 months of the pandemic.

The initial reports on COVID-19 from China, describing clinical presentation, diagnosis, and mortality risk factors, were multi-author collaborations from hospitals and academic centers ^1-4^. Subsequent reports included other research and public health stakeholders. Clinical series in the U.S. evolved to include larger multicenter patient cohorts from multiple academic centers and large health systems ^5 6^. This focus on large databases has led to the exclusion of many clinically-intensive front-line clinicians from the research and scholarly enterprise. Many of these are academic physicians whose roles and career paths include their own scholarly productivity and training of fellows, residents and students. During the pandemic, they lacked dedicated time and resources to do scholarly work and were unable to compete with their colleagues with primary research roles. These faculty and their trainees have the unique opportunity to generate research questions based on first-hand observations, but without institutional support, these questions go uninvestigated. Other primary research faculty created proprietary clinical databases to answer research question relevant to their field but the lack of cross-fertilization with front-line clinicians and colleagues in other disciplines compromises the depth of analysis. This affects the institutional climate, already stressed by the pandemic, in that the energy and enthusiasm from diverse faculty and trainees are not harnessed for the advancement of knowledge; sadly, there is disengagement and a sense of lack of institutional investment in their academic futures.

Our own local COVID-19 response plan at Yale University School of Medicine (YSM) started in March 2020 and was organized by leadership at YSM and Yale New Haven Health System (YNHHS). The detailed orchestration of efforts involved close interplay between front-line clinicians (academic faculty and hospitalist staff) and colleagues experienced in clinical data analysis. Other clinical faculty and trainees who were making key clinical observations expressed a desire to produce scholarly work but were hampered by lack of a readily available and comprehensive clinical data repository. Various groups tried to create their own databases with manual data extraction from the electronic health record, but clinical workload did not allow for timely data management and led to duplication of data collection efforts.

Feedback to leadership from a diverse group of faculty members in the Department of Medicine (DOM) revealed that this lack of access to clinical data resulted in front-line clinicians feeling excluded from the research enterprise. Thus, DOM leadership and a sub-group of faculty (clinicians, clinical researchers and informatics specialists) met to discuss different approaches to this problem. Ultimately, the group agreed on a DOM-supported innovative approach to creating a real-time, clinically comprehensive, common clinical data repository on patients admitted for COVID 19 dating from the start of the pandemic. In this report, we describe the creation and dissemination of Department of Medicine COVID-19 Explorer and Repository (DOM-CovX) which is envisioned to serve as a starting point for clinical queries that could lead to projects ranging from quality improvement to scholarly efforts. We also describe how this innovation serves to “democratize” the research enterprise by engaging diverse researchers and trainees and promoting a broader scope of scholarship activities.

## METHODS

### Construction of the DOM-CovX Data Repository

YNHHS encompasses 5 hospitals located across Connecticut and Rhode Island with a combined capacity for 2681 inpatient beds. The participating hospitals work together with the Yale University School of Medicine (YSM) which oversees both clinical and non-clinical faculty. All sites use Epic (Verona, WI) as the joint electronic health record (EHR) for documentation of clinical care issues.

Data on COVID-19-positive hospitalized patients were extracted from the EHR using Clarity. The cohort was defined as hospitalizations where the patient’s first positive COVID-19 test was 14 days preceding admission to time of discharge. This definition was chosen to be sufficiently sensitive to capture individuals who may have been tested as an outpatient and then presented for hospitalization 1-2 weeks following diagnosis, while also sufficiently specific to exclude patients with recurrent positive swabs, which may not signify new infection but rather non-viable virus with continued PCR positivity. COVID-19 testing within the YNHHS was done by nasopharyngeal (NP) swab testing via PCR.

Data across several key domains were extracted: demographics, past medical history, laboratory values during hospitalization, vital signs, medications, imaging, procedures (e.g. intubation), and outcomes (e.g. death, length of stay, patient disposition). Given the time-varying nature of several data domains, summary statistics were constructed to limit the computational size of the dataset and provide a reasonable data file that the broader research community could use for basic statistical analyses. To compress each hospitalization to ‘one-row-per-patient’, summary statistics were calculated for the time-varying data domains with continuous values, which included vital signs and labs. Summary statistics included minimum, maximum, mean, median, 25^th^ and 75^th^ percentiles, standard deviation. Medications and procedures were reported as ‘ever’ and ‘never’, e.g. if a patient ever received tocilizumab during hospitalization, the variable would read as a ‘1’ and ‘0’ otherwise.

Data were de-identified to remove protected health information as per the HIPAA Security Rule. All timestamps more specific than year were removed including admission and discharge times, timing of vital signs, laboratory measurements, procedures, or medication administrations. Ages greater than 90 were set to 90 as per the HIPAA Security Rule.

### Accessing the DOM CovX Data Explorer and Repository

The process for accessing the DOM-CovX Data Explorer (the interactive platform) and/or the Data Repository (master dataset) is shown in **Figure 1**. Researchers who want to pursue further study are afforded an online form **(Supplement 1)** to request specific data elements from the DOM-CovX repository as well as information on the population, exposure, outcome of interest and hypothesis being tested. A core DOM-CovX team consisting of various representatives from the clinical faculty, informatics department, data scientists, statisticians and research coordinators reviews the application and assists in the process. A pre-approved HIC number and an Institutional Review Board (IRB) Protocol is required to be uploaded at the time of data request. If the team recognizes any potential for collaboration between departments and research groups doing similar or related projects, a dialogue would be facilitated.

**Figure 1:**
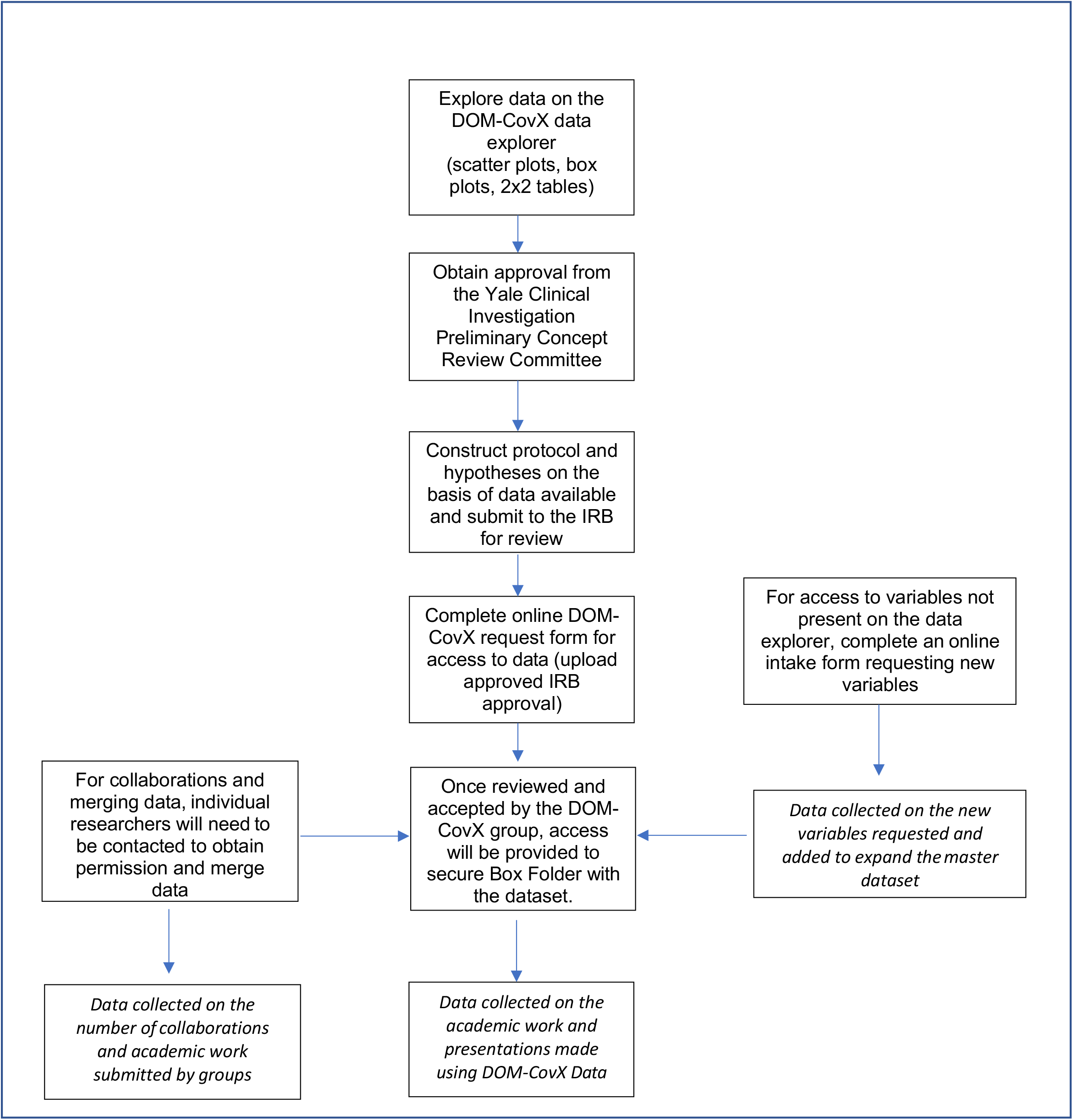
Flowchart showing process of data request, acquisition and distribution, with relevant outcomes and data elements planned to be collected by the DOM-CovX management team.

### Data Distribution

Once the application is approved by the core DOM-CovX team, data is distributed to researchers through Secure Box, a HIPAA-compliant cloud-based file management system used at Yale for data distribution. Data is left in raw format and is used by researchers on a variety of statistical software packages for analysis. Biostatistical analysis is left to the individual researchers. This could include importation to other data collection and analytical systems such as REDCap, a browser based, metadata-driven software to collect and store data for clinical and translational research^7,8^. Researchers have the option of requesting additional data elements from the DOM-CovX team as well as engaging in manual data extraction for fields that were not amenable to computerized data transfer; these additional data elements will be added to the master dataset. A master-file for all request and data provisions is kept and monitored to determine any potential for facilitating collaborations. The DOM-CovX team also tracks resulting presentations, publications and other academic productivity.

### Front-Facing Web Interface for Data Exploration (DOM-CovX Explorer)

A unique feature is the DOM-CovX Explorer which is a web application designed to provide investigators a front-end interface to view and manipulate variables, see summary statistics, as well as receive additional information about cohort design and data requests. It was constructed using R Shiny (R Foundation for Statistical Computing, Vienna, Austria) to Statistical visualizations included single variable analyses, scatter plots, box plots, as well as 2×2 tables as shown in **Figure 2**. Yale researchers can openly explore the data to assess if they are interested in further in-depth study that requires access to the Data Repository.

**Figure 2:**
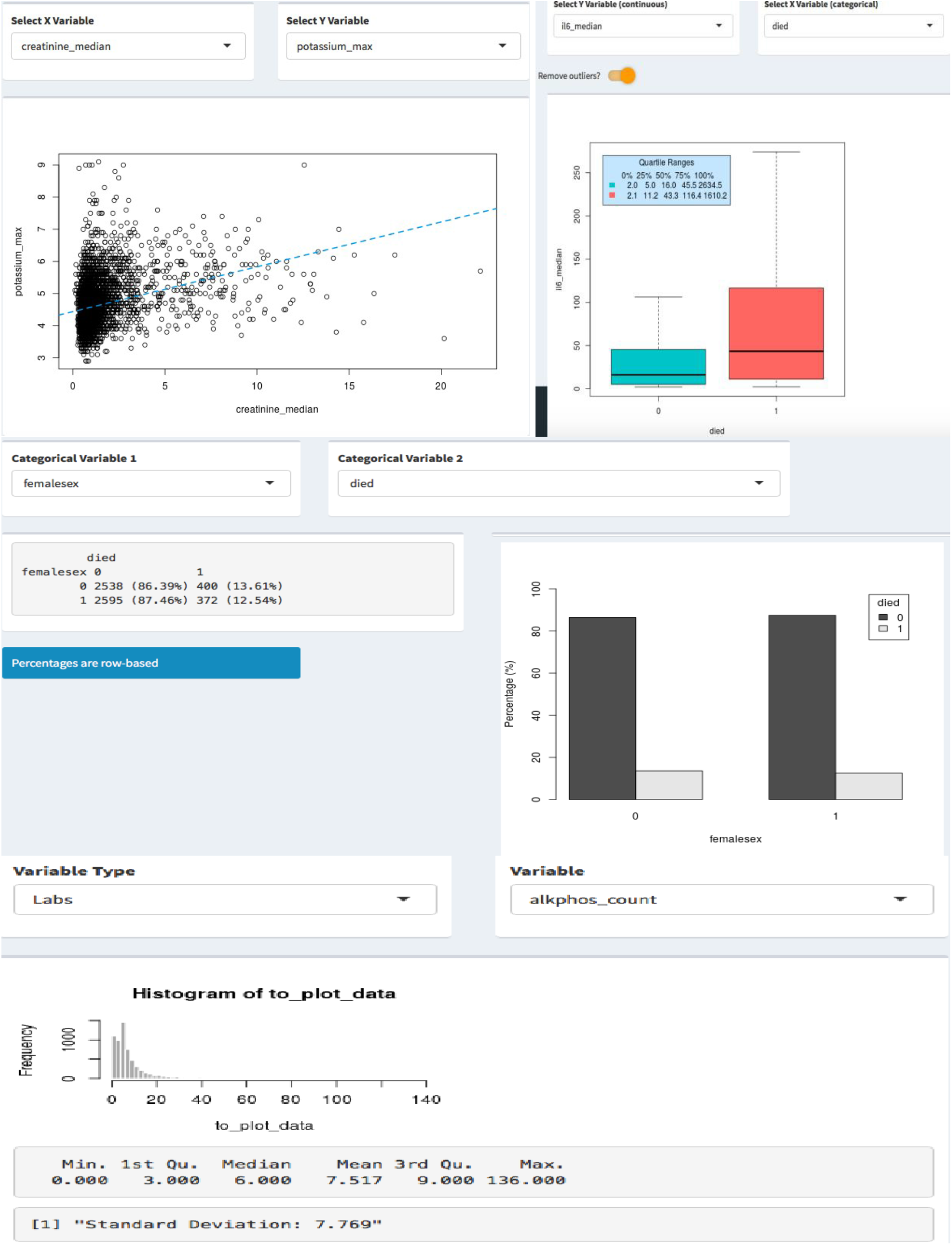
The DOM-CovX Data Explorer (web interface) showing different functions to find correlations between distinct variables and aid in hypothesis generation.

## EXPECTED AND PRELIMINARY OUTCOMES

Open access data is not novel; especially during the COVID-19 pandemic, there has been increasing availability of such data. For example, clinicians and researchers can access online data trackers from Johns Hopkins, the CDC COVID data tracker, Clinicaltrials.gov, and local health departments. These databases offer information on case trends, demographics, vaccinations, and other outcomes; however, specific clinically relevant data are often lacking. Larger national databases such as N3C created and maintained by NIH are available but access can be restricted. Local proprietary databases such as the COVID-19-specific Johns Hopkins Crown Registry (JH-CROWN) uses a precision medicine analytics platform which aims to distribute data to institutional investigators^9,10^. Yale researchers have also created large scale data repositories for COVID including an alternative COVID clinical data repository curated through the Yale Observational Medical Outcomes Partnership (OMOP) ^11^. However, these repositories are primarily accessible to large research teams and can present logistic barriers for other faculty and trainees to access. We believe that the DOM-CovX Data Explorer and Repository offer the following innovative and distinct advantages:

### 1) Accelerate generation of new knowledge and increase scholarly productivity with particular local relevance

Specific features that promote this outcome include:

#### a) Comprehensive clinical data

The DOM-CovX dataset contains more than 3000 clinically relevant data elements, including from demographics, laboratory, past medical history, vital signs, medications. The dataset covers the vast majority of electronic health record data variables, includes specifics of disease stages (e.g. acute kidney injury stage 1, 2, 3) and distribution of data elements (e.g. mean, mode, median, standard deviation and inter-quartile ranges) and thus caters to a wide array of clinicians and researchers.

#### b) User-friendly interface

The DOM-CovX Data Explorer (web-interface) was created to offer openly accessible data to support hypothesis generation and facilitate grant and manuscript assembly. Variables within the DOM-CovX data can be easily searched for, and associations compared using the single variable summary, scatter plot, box plot and 2×2 table functions. A detailed data dictionary **(Supplement 2)** is available to facilitate understanding and transparency of data.

#### c) Easy, free and quick access

The DOM-CovX is easily accessed via the webpage which also contains a guide for explaining how to request data, in addition to providing a downloadable Institutional Review Board (IRB) template for guidance and rapid approval. Significantly, the DOM-CovX database is completely free to all Yale faculty and trainees who may otherwise lack independent grant funding and staff to pursue data repository creation.

#### d) Rapidly evolving dataset with customization options

With a vision to initiate and promote collaborations, the DOM-CovX allows and encourages researchers to contribute their own data elements, whether through an automated data extraction or chart review. The database is regularly updated in a biweekly fashion. Since inception (July 2020), the dataset has expanded to include 7,147 inpatient encounters, with 3,071 variables across 7 different clinical domains.

### 2) Improve the institutional academic climate by

#### a) Broadening research scope

The DOM-CovX provides a centralized, uniformly-populated resource that serves as a starting point for research. The types of questions asked can be broad especially as hypothesis testing is promoted by using the Explorer function. The comprehensiveness of data fields can spawn hypotheses that could involve interactions between clinician observers and basic scientists. Different subspecialties within the DOM have already used the database to address relevant questions such as: (1) the impact of obesity and diabetes ^12^; (2) the association of acute kidney injury (AKI) with Covid-19^13^; (3) the role of neutrophil activation signature in predicting critical illness and mortality in Covid-19^14^; (4) the association of HIV with COVID outcomes^15^.

#### b) Expanding research capability to more diverse group of stakeholders

The DOM-CovX is designed to be inclusive of a broad range of researchers. The simplicity of data request and acquisition, and encouragement to collaborate with established faculty allow early career trainees to receive mentorship. The DOM-CovX core team identifies researchers in need of biostatics support and connects them to appropriate resources. We envision that this effort will lead to future educational and training opportunities to expand research capability.

#### c) Enhancing interdepartmental collaborations

The DOM-CovX data access process can provide more collaborative opportunities as the application process can identify researchers interested in similar projects. Often, faculty are not aware that other expertise exists outside their immediate circles that could expand the depth and scope of their inquiry. Multiple collaborations have already been made, bringing together researchers from Cardiology, Nephrology, Endocrinology and Hematology/Oncology. Despite being active for less than a year, 5 manuscripts have already been published using DOM-CovX data^13,14,16-18^, with 5 abstracts submitted and 4 grant submissions, of which 2 have been funded.

## LIMITATIONS

Analytic tools such as statistical analysis, have not yet been built into the workflow. Future goals will incorporate access to fundamental analytic tools and statisticians as well as improved training and mentoring of clinician-educators and clinicians to engage in fundamental understanding of research analytics as well as to improve data management skills. Another limitation is that the DOM-CovX database is limited to local hospitalization data and excludes outpatient and subsequent follow-up data. The database focuses only on clinical data and does not lend itself to studies requiring a biorepository. The database and web interface are not public -facing, but efforts in this direction have commenced.

## CONCLUSIONS

We believe the DOM-CovX Data Explorer and Repository have great potential to increase academic productivity and our preliminary experience bears this out. By providing an accessible tool for simple data analysis and access to a consistently updated, standardized and large-scale dataset, it overcomes barriers for a wide variety of researchers. We believe that this innovative approach represents an opportunity to improve the institutional climate by fostering collaboration and diversity of scholarly pursuits. The application process and use of DOM-CovX are currently operational and being expanded. We anticipate that as the process evolves, there will be a growing impact on medical education. Specifically, the rapidly changing academic landscape requires that curricula provide the intellectual currency for all trainees and faculty to critically understand the explosion of clinical and research literature including the optimal use of data and recognition of its limitations. Inequities of data access may result in a more divided group of learners with narrower expectations and rewards. We believe that the DOM Cov-X can bridge this intellectual divide and expand the scope and quality of medical education.

## Supporting information

Supplement 1. DOM CovX Request Form

Supplement 2. DOM CovX Data Dictionary

## Data Availability

Any data referred to in this manuscript will be made available upon request

## Notes

### Competing Interest Statement

The authors have declared no competing interest.

### Funding Statement

No external funding was received for this project

### Author Declarations

The Yale University HIC Board provided exemption for this study

