## Supplement 1. DOM CovX Request Form for "The Yale Department of Medicine COVID-19 Data Explorer and Repository (DOM-CovX): An Innovative Approach to Promoting Collaborative Scholarship During a Pandemic"

### DOMCOVX CTRA Request Form

Start of Block: Default Question Block

Q16 **DOM-COVX Data Request Form**

Q1 Title of Research Proposal

________________________________________________________________

Q2 First name of Principle Investigator

________________________________________________________________

Q3 Last name of Principle Investigator

________________________________________________________________

Q4 NetID of Principle Investigator

________________________________________________________________

Q31 Email of Principle Investigator

________________________________________________________________

Q6 Would you like to add any Co-Investigators?

○ Yes (1)

○ No (2)

---

*Display This Question:*

*If Would you like to add any Co-Investigators? = Yes*

Q7 Name of Co-Investigator

________________________________________________________________

________________________________________________________________

________________________________________________________________

________________________________________________________________

________________________________________________________________

---

*Display This Question:*

*If If Name of Co-Investigator Text Response Is Displayed*

Q25 NetID of Co-Investigator

________________________________________________________________

---

*Display This Question:*

*If If Name of Co-Investigator Text Response Is Not Empty*

Q17 Would you like to add another Co-Investigator?

○ Yes (1)

○ No (2)

---

*Display This Question:*

*If Would you like to add another Co-Investigator? = Yes*

Q18 Name of Co-Investigator

________________________________________________________________

*Display This Question:*

*If If Name of Co-Investigator  Text Response Is Displayed*

Q26 NetID of Co-Investigator

________________________________________________________________

Q10 Population(s) of Interest
(e.g. patients with diabetes who underwent intubation)

*Currently, the DOM-CovX only contains data on individuals hospitalised with PCR-proven SARS-COV2 infection at one of the six YNHH-affiliated hospitals.*

________________________________________________________________

Q8 Exposure(s) of Interest
(e.g. metformin)

________________________________________________________________

Q9 Outcome(s) of Interest
(e.g. death)

________________________________________________________________

Q11 Hypothesis and/or Statement of Intent
(e.g. metformin confers protection in diabetes with severe respiratory compromise from COVID)

________________________________________________________________

Q12 Approved project HIC #

*Please note that no data will be available without valid IRB approval or exemption.*

________________________________________________________________

Q29 Please upload your IRB-approved protocol

Q13 Will your team require biostatistics support?

- ○ Yes (1)
- ○ No (2)

Q14 Will your team require data including PHI? (e.g. patient identifiers such as MRNs, CSNs, zip codes etc.)

*Please note that PHI data may take longer to process and may require additional review.*

- ○ Yes (1)
- ○ No (2)

*Display This Question:*

*If Will your team require data including PHI? (e.g. patient identifiers such as MRNs, CSNs, zip code... = Yes*

Q27 Does your team have access to a secure workstation managed by Yale/YNHH for receiving data with PHI?

- Yes (1)
- No (2)

---

Q15 From the following sub-categories, please select ALL relevant to your data needs.

- Demographics (1)
- Comorbidities (2)
- Vitals (3)
- Orders/Procedures (4)
- Medications (5)
- Laboratory Results (6)

---

Q21 Will the data you are requesting be combined with novel data you or your collaborators have generated as part of a research project ?

- Yes (1)
- No (2)

---

*Display This Question:*

*If Will the data you are requesting be combined with novel data you or your collaborators have gener... = Yes*

Q22 Will you be willing to integrate your novel dataset into the larger DOM-CovX Dataset? (Please note that all your contributed data will require your express approval prior to being distributed to other investigators).

◯ Yes (1)

◯ No (2)

---

*Display This Question:*

*If Will you be willing to integrate your novel dataset into the larger DOM-CovX Dataset? (Please not... = Yes*

Q23 Please list the novel elements you have collected that can be added the DOM-CovX Data Repository.

________________________________________________________________

________________________________________________________________

________________________________________________________________

________________________________________________________________

________________________________________________________________

---

Q30 Will data you receive by DOM-CovX team be sent, used, or analyzed by any non-Yale institutions, companies, contractors or non-Yale affiliated individuals?

◯ Yes (1)

◯ No (2)

---

Q19 Would you like to provide any additional notes/comments?

________________________________________________________________

**End of Block: Default Question Block**
