## Supplement 2. DOM CovX Data Dictionary for "The Yale Department of Medicine COVID-19 Data Explorer and Repository (DOM-CovX): An Innovative Approach to Promoting Collaborative Scholarship During a Pandemic"

Supplement 3 -DOM-CovX Data Dictionary

| variable_name | pretty_name | data_type | short_description | units_continuous_only | variable_domain | deprecated | allowed_values | key_outcome | validated | detailed_note | final_domain | questions_notes |
|---|---|---|---|---|---|---|---|---|---|---|---|---|
| **abogroupinga** | Blood Type (A) | categorical | ABO Blood Group test reads blood type A for this patient | | Demographics | 0 | | | | | | |
| **abogroupingab** | Blood Type (AB) | categorical | ABO Blood Group test reads blood type AB for this patient | | Demographics | 0 | | | | | | |
| **abogroupingb** | Blood Type (B) | categorical | ABO Blood Group test reads blood type B for this patient | | Demographics | 0 | | | | | | |
| **abogroupingo** | Blood Type (O) | categorical | ABO Blood Group test reads blood type O for this patient | | Demographics | 0 | | | | | | |
| **age** | Age | continuous | Age | Years | Demographics | 0 | | | | Age at admission | demographics | |
| **bmi** | Body Mass Index | continuous | Body Mass Index | kg/m2 | Demographics | 0 | | | | weight(kg)/height(m)^2 | demographics | |
| **died** | Died | categorical | Death during hospitalization | | Demographics | 0 | | 1 | | discharge disposition=""""Expired"""" | location | |
| **discharge_dispsn** | Discharge Disposition | categorical | Discharge disposition | | Demographics | 0 | | | | | demographics | |
| **femalesex** | Female | categorical | Female sex | | Demographics | 0 | | | | sex==""""female"""" | demographics | |
| **first_pos_covid_test** | First Positive Covid Test | categorical | First Positive Covid Test Recorded for this patient | | Demographics | 0 | | | | Not in demographics within Dom_Covx | | |
| **height** | Height | continuous | Height | Cm | Demographics | 0 | | | | | demographics | |
| **latino** | Latino | categorical | Latino Identity | | Demographics | 0 | | | | | demographics | |
| **los** | Length of Stay | continuous | Length of Hospitalization | days | Demographics | 0 | | | | | location | |
| **race** | Race | categorical | Patient's racial Identity | | Demographics | 0 | | | | | demographics | |
| **rhtype** | Rhesus Factor Blood Test | categorical | Rhesus Factor Blood Test results for this patient | | Demographics | 0 | | | | | | |
| **weight** | Weight | continuous | Patient weight | kg | Demographics | 0 | | | | | demographics | |
| **emergency_room** | Emergency Room | categorical | Admitted to Emergency Room during admission | | Hospitalization Variables | 0 | | | | | location | |
| **hospital** | Hospital | categorical | Hospitalized | | Hospitalization Variables | 0 | | | | | location | |
| **los_icu** | Length of Stay ICU | continuous | Length of stay in ICU | days | Hospitalization Variables | 0 | | | | | location | |
| **medical_icu** | Medical ICU | categorical | Admitted to Medical ICU during admission | | Hospitalization Variables | 0 | | | | | location | |
| **medical_surgical_icu** | Medical Surgical ICU | categorical | Admitted to Medical Surgical ICU during admission | | Hospitalization Variables | 0 | | | | | location | |
| **medical_surgical_ward** | Medical Surgical Ward | categorical | Admitted to Medical Surgical Ward during admission | | Hospitalization Variables | 0 | | | | | location | |
| **medical_ward** | Medical Ward | categorical | Admitted to Medical Ward during admission | | Hospitalization Variables | 0 | | | | | location | |
| **surgical_icu** | Surgical ICU | categorical | Admitted to Surgical ICU during admission | | Hospitalization Variables | 0 | | | | | location | |
| **surgical_ward** | Surgical Ward | categorical | Admitted to Surgical Ward during admission | | Hospitalization Variables | 0 | | | | | location | |
| **a1glob_25** | Alpha 1 Globulin (25th Percentile) | continuous | Alpha 1 Globulin reading at the 25th percentile of this patient's measurements | g/L | Labs | 0 | | | | | labs | |
| **a1glob_75** | Alpha 1 Globulin (75th Percentile) | continuous | Alpha 1 Globulin reading at the 75th percentile of this patient's measurements | g/L | Labs | 0 |  |  |  |  | labs |  |
| **a1glob_count** | Alpha 1 Globulin (Count) | continuous | Count of alpha 1 globulin measurements for this patient | Number of measurements | Labs | 0 | | | | | labs | |
| **a1glob_first** | Alpha 1 Globulin (First) | continuous | First alpha 1 globulin reading for this patient's hospitalization | g/L | Labs | 0 | | | | | labs | |
| **a1glob_last** | Alpha 1 Globulin (Last) | continuous | Last alpha 1 globulin reading for this patient's hospitalization | g/L | Labs | 0 | | | | | labs | |
| **a1glob_max** | Alpha 1 Globulin (Max) | continuous | Maximum alpha 1 globulin reading for this patient's hospitalization | g/L | Labs | 0 | | | | | labs | |
| **a1glob_mean** | Alpha 1 Globulin (Mean) | continuous | Average alpha 1 globulin reading for this patient's hospitalization | g/L | Labs | 0 | | | | | labs | |
| **a1glob_median** | Alpha 1 Globulin (Median) | continuous | Median alpha 1 globulinreading for this patient's hospitalization | g/L | Labs | 0 | | | | | labs | |
| **a1glob_min** | Alpha 1 Globulin (Minimum) | continuous | Minimum alpha 1 globulin reading for this patient's hospitalization | g/L | Labs | 0 | | | | | labs | |
| **a1glob_sd** | Alpha 1 Globulin (SD) | continuous | Standard deviation of the alpha 1 globulin readings during this patient's hospitalization | g/L | Labs | 0 |  |  |  |  | labs |  |
| **a2glob_25** | Alpha 2 Globulin (25th Percentile) | continuous | Alpha 2 Globulin reading at the 25th percentile of this patient's measurements | g/L | Labs | 0 | | | | | labs | |
| **a2glob_75** | Alpha 2 Globulin (75th Percentile) | continuous | Alpha 2 Globulin reading at the 75th percentile of this patient's measurements | g/L | Labs | 0 |  |  |  |  | labs |  |
| **a2glob_count** | Alpha 2 Globulin (Count) | continuous | Count of alpha 2 globulin measurements for this patient | Number of measurements | Labs | 0 | | | | | labs | |
| **a2glob_first** | Alpha 2 Globulin (First) | continuous | First alpha 2 globulin reading for this patient's hospitalization | g/L | Labs | 0 | | | | | labs | |
| **a2glob_last** | Alpha 2 Globulin (Last) | continuous | Last alpha 2 globulin reading for this patient's hospitalization | g/L | Labs | 0 | | | | | labs | |
| **a2glob_max** | Alpha 2 Globulin (Max) | continuous | Maximum alpha 2 globulin reading for this patient's hospitalization | g/L | Labs | 0 | | | | | labs | |
| **a2glob_mean** | Alpha 2 Globulin (Mean) | continuous | Average alpha 2 globulin reading for this patient's hospitalization | g/L | Labs | 0 | | | | | labs | |
| **a2glob_median** | Alpha 2 Globulin (Median) | continuous | Median alpha 2 globulinreading for this patient's hospitalization | g/L | Labs | 0 | | | | | labs | |
| **a2glob_min** | Alpha 2 Globulin (Minimum) | continuous | Minimum alpha 2 globulin reading for this patient's hospitalization | g/L | Labs | 0 | | | | | labs | |
| **a2glob_sd** | Alpha 2 Globulin (SD) | continuous | Standard deviation of the alpha 2 globulin readings during this patient's hospitalization | g/L | Labs | 0 |  |  |  |  | labs |  |
| **a2plasma_25** | | continuous | | | Labs | 0 | | | | | labs | |
| **a2plasma_75** | | continuous | | | Labs | 0 | | | | | labs | |
| **a2plasma_count** | | continuous | | | Labs | 0 | | | | | labs | |
| **a2plasma_first** | | continuous | | | Labs | 0 | | | | | labs | |
| **a2plasma_last** | | continuous | | | Labs | 0 | | | | | labs | |
| **a2plasma_max** | | continuous | | | Labs | 0 | | | | | labs | |
| **a2plasma_mean** | | continuous | | | Labs | 0 | | | | | labs | |
| **a2plasma_median** | | continuous | | | Labs | 0 | | | | | labs | |
| **a2plasma_min** | | continuous | | | Labs | 0 | | | | | labs | |
| **a2plasma_sd** | | continuous | | | Labs | 0 | | | | | labs | |
| **abogrouping_first** | Blood Type (ABO) (First) | categorical | First ABO Blood Group Test for this patient's hospitalization | | Labs | 0 | | | | | labs | |
| **abogrouping_last** | Blood Type (ABO) (Last) | categorical | Last ABO Blood Group Test for this patient's hospitalization | | Labs | 0 | | | | | labs | |
| **acelevel_25** | Angiotensin Converting Enzyme Levels (25th Percentile) | continuous | Angiotensin converting enzyme levels in the blood at the 25th percentile of this patient's measurements | ?L | Labs | 0 |  |  |  |  | labs |  |
| **acelevel_75** | Angiotensin Converting Enzyme Levels (75th Percentile) | continuous | Angiotensin converting enzyme levels in the blood at the 75th percentile of this patient's measurements | ?L | Labs | 0 | | | | | labs | |
| **acelevel_count** | Angiotensin Converting Enzyme Levels (Count) | continuous | Count of angiotensin converting enzyme blood measurements of this patient | Number of measurements | Labs | 0 | | | | | labs | |
| **acelevel_first** | Angiotensin Converting Enzyme Levels (First) | continuous | First angiotensin converting enzyme blood measurement for this patient's hospitalization | ?L | Labs | 0 | | | | | labs | |
| **acelevel_last** | Angiotensin Converting Enzyme Levels (Last) | continuous | Last Angiotensin converting enzyme blood measurement for this patient's hospitalization | ?L | Labs | 0 | | | | | labs | |
| **acelevel_max** | Angiotensin Converting Enzyme Levels (Max) | continuous | Maximum angiotensin converting enzyme blood measurement for this patient's hospitalization | ?L | Labs | 0 | | | | | labs | |
| **acelevel_mean** | Angiotensin Converting Enzyme Levels (Mean) | continuous | Average angiotensin converting enzyme blood measurement for this patient's hospitalization | ?L | Labs | 0 | | | | | labs | |
| **acelevel_median** | Angiotensin Converting Enzyme Levels (Median) | continuous | Median angiotensin converting enzyme blood measurement for this patient's hospitalization | ?L | Labs | 0 | | | | | labs | |
| **acelevel_min** | Angiotensin Converting Enzyme Levels (Min) | continuous | Minimum angiotensin converting enzyme blood measurement for this patient's hospitalization | ?L | Labs | 0 | | | | | labs | |
| **acelevel_sd** | Angiotensin Converting Enzyme Levels (SD) | continuous | Standard deviation of the angiotensin converting enzyme blood measurements during this patient's hosp | ?L | Labs | 0 | | | | | labs | |
| **acetaminophen_25** | Acetaminophen Level (25th Percentile) | continuous | Acetaminophen blood level at the 25th percentile of this patient's measurements | mcg/mL | Labs | 0 | | | | | labs | |
| **acetaminophen_75** | Acetaminophen Level (75th Percentile) | continuous | Acetaminophen blood level at the 75th percentile of this patient's measurements | mcg/mL | Labs | 0 | | | | | labs | |
| **acetaminophen_count** | Acetaminophen Level (Count) | continuous | Count of acetaminophen blood level measurements for this patient | Number of measurements | Labs | 0 | | | | | labs | |
| **acetaminophen_first** | Acetaminophen Level (First) | continuous | First acetaminophen blood level measurement for this patient's hospitalization | mcg/mL | Labs | 0 | | | | | labs | |
| **acetaminophen_last** | Acetaminophen Level (Last) | continuous | Last acetaminophen blood level measurement for this patient's hospitalization | mcg/mL | Labs | 0 | | | | | labs | |
| **acetaminophen_max** | Acetaminophen Level (Max) | continuous | Maximum acetaminophen blood level for this patient's hospitalization | mcg/mL | Labs | 0 | | | | | labs | |
| **acetaminophen_mean** | Acetaminophen Level (Mean) | continuous | Average acetaminophen blood level for this patient's hospitalization | mcg/mL | Labs | 0 | | | | | labs | |
| **acetaminophen_median** | Acetaminophen Level (Median) | continuous | Median acetaminophen blood level for this patient's hospitalization | mcg/mL | Labs | 0 | | | | | labs | |
| **acetaminophen_min** | Acetaminophen Level (Min) | continuous | Minimum acetaminophen blood level for this patient's hospitalization | mcg/mL | Labs | 0 | | | | | labs | |
| **acetaminophen_sd** | Acetaminophen Level (SD) | continuous | Standard deviation of the acetaminophen levels during this patient's hospitalization | mcg/mL | Labs | 0 | | | | | labs | |
| **acetoneblood_25** | Acetone Blood Level (25th Percentile) | continuous | Acetone blood level at the 25th percentile of this patient's measurements | mcg/mL | Labs | 0 | | | | | labs | |
| **acetoneblood_75** | Acetone Blood Level (75th Percentile) | continuous | Acetone blood level at the 75th percentile of this patient's measurements | mcg/mL | Labs | 0 | | | | | labs | |
| **acetoneblood_count** | Acetone Blood Level (Count) | continuous | Count of acetone blood level measurements for this patient | Number of measurements | Labs | 0 | | | | | labs | |
| **acetoneblood_first** | Acetone Blood Level (First) | continuous | First acetone blood level reading for this patient's hospitalization | mcg/mL | Labs | 0 | | | | | labs | |
| **acetoneblood_last** | Acetone Blood Level (Last) | continuous | Last acetone blood level reading for this patient's hospitalization | mcg/mL | Labs | 0 | | | | | labs | |
| **acetoneblood_max** | Acetone Blood Level (Max) | continuous | Maximum acetone blood level for this patient's hospitalization | mcg/mL | Labs | 0 | | | | | labs | |
| **acetoneblood_mean** | Acetone Blood Level (Mean) | continuous | Average acetone blood level for this patient's hospitalization | mcg/mL | Labs | 0 | | | | | labs | |
| **acetoneblood_median** | Acetone Blood Level (Median) | continuous | Median acetone blood level for this patient's hospitalization | mcg/mL | Labs | 0 | | | | | labs | |
| **acetoneblood_min** | Acetone Blood Level (Min) | continuous | Minimum acetone blood level for this patient's hospitalization | mcg/mL | Labs | 0 | | | | | labs | |
| **acetoneblood_sd** | Acetone Blood Level (SD) | continuous | Standard deviation of the acetone blood level measurements during this patient's hospitalization | mcg/mL | Labs | 0 | | | | | labs | |
| **acratiourine_25** | Urine Albumin-Creatinine Ratio (25th Percentile) | continuous | Urine Albumin-Creatinine ratio at the 25th percentile of this patient's measurements | mg | Labs | 0 | | | | | labs | |
| **acratiourine_75** | Urine Albumin-Creatinine Ratio (75th Percentile) | continuous | Urine Albumin-Creatinine ratio at the 75th percentile of this patient's measurements | mg | Labs | 0 |  |  |  |  | labs |  |
| **acratiourine_count** | Urine Albumin-Creatinine Ratio (Count) | continuous | Count of urine Albumin-Creatinine ratios for this patient | Number of measurements | Labs | 0 |  |  |  |  | labs |  |
| **acratiourine_first** | Urine Albumin-Creatinine Ratio (First) | continuous | First urine Albumin-Creatinine ratio measurement for this patient's hospitalization | mg | Labs | 0 | | | | | labs | |
| **acratiourine_last** | Urine Albumin-Creatinine Ratio (Last) | continuous | Last urine Albumin-Creatinine ratio measurement for this patient's hospitalization | mg | Labs | 0 | | | | | labs | |
| **acratiourine_max** | Urine Albumin-Creatinine Ratio (Max) | continuous | Maximum urine Albumin-Creatinine ratio for this patient's hospitalization | mg | Labs | 0 | | | | | labs | |
| **acratiourine_mean** | Urine Albumin-Creatinine Ratio (Mean) | continuous | Average urine Albumin-Creatinine ratio for this patient's hospitalization | mg | Labs | 0 |  |  |  |  | labs |  |
| **acratiourine_median** | Urine Albumin-Creatinine Ratio (Median) | continuous | Median urine albumin creatinine ratio t for this patient's hospitalization | mg | Labs | 0 | | | | | labs | |
| **acratiourine_min** | Albumin Creatinine Ratio (Min) | continuous | Minimum urine albumin creatinine ratio for this patient's hospitalization | mg | Labs | 0 | | | | | labs | |
| **acratiourine_sd** | Albumin Creatinine Ratio (SD) | continuous | Standard deviation of the urine albumin creatinine ratio measurements during this patient's hospitalizatio | mg | Labs | 0 | | | | | labs | |
| **act_25** | Activated Clotting Time Level (25th Percentile) | continuous | Activated clotting times at the 25th percentile of this patient's measurements | Seconds | Labs | 0 | | | | | labs | |
| **act_75** | Activated Clotting Time Level (75th Percentile) | continuous | Activated clotting time at the 75th percentile of this patient's measurements | Seconds | Labs | 0 | | | | | labs | |
| **act_count** | Activated Clotting Time Level (Count) | continuous | Count of activated clotting times for this patient | Number of measurements | Labs | 0 |  |  |  |  | labs |  |
| **act_first** | Activated Clotting Time Level (First) | continuous | First activated clotting time for this patient's hospitalization | Seconds | Labs | 0 | | | | | labs | |
| **act_last** | Activated Clotting Time Level (Last) | continuous | Last activated clotting time for this patient's hospitalization | Seconds | Labs | 0 | | | | | labs | |
| **act_max** | Activated Clotting Time Level (Max) | continuous | Maximum activated clotting time for this patient's hospitalization | Seconds | Labs | 0 | | | | | labs | |
| **act_mean** | Activated Clotting Time Level (Mean) | continuous | Average activated clotting time for this patient's hospitalization | Seconds | Labs | 0 | | | | | labs | |
| **act_median** | Activated Clotting Time Level (Median) | continuous | Median activated clotting time for this patient's hospitalization | Seconds | Labs | 0 | | | | | labs | |
| **act_min** | Activated Clotting Time Level (Min) | continuous | Minimum activated clotting time for this patient's hospitalization | Seconds | Labs | 0 | | | | | labs | |
| **act_sd** | Activated Clotting Time Level (SD) | continuous | Standard deviation of the activated clotting times during this patient's hospitalization | Seconds | Labs | 0 | | | | | labs | |
| **adenovirus_first** | | | | | Labs | 0 | | | | | labs | |
| **adenovirus_last** | | | | | Labs | 0 | | | | | labs | |
| **afp_25** | Alpha-fetoprotein (25th Percentile) | continuous | Alpha-fetoprotein level at the 25th percentile of this patient's measurements | ng/mL | Labs | 0 | | | | | labs | |
| **afp_75** | Alpha-fetoprotein (75th Percentile) | continuous | Alpha-fetoprotein level at the 75th percentile of this patient's measurements | ng/mL | Labs | 0 | | | | | labs | |
| **afp_count** | Alpha-fetoprotein (Count) | continuous | Count of alpha-fetoprotein measurements for this patient | Number of measurements | Labs | 0 | | | | | labs | |
| **afp_first** | Alpha-fetoprotein (First) | continuous | First alpha-fetoprotein measurement for this patient's hospitalization | ng/mL | Labs | 0 | | | | | labs | |
| **afp_last** | Alpha-fetoprotein (Last) | continuous | Last alpha-fetoprotein measurement for this patient's hospitalization | ng/mL | Labs | 0 | | | | | labs | |
| **afp_max** | Alpha-fetoprotein (MaAlpha-fetoprotein) | continuous | Maximum alpha-fetoprotein measurement for this patient's hospitalization | ng/mL | Labs | 0 | | | | | labs | |
| **afp_mean** | Alpha-fetoprotein (Mean) | continuous | Average alpha-fetoprotein measurement for this patient's hospitalization | ng/mL | Labs | 0 | | | | | labs | |
| **afp_median** | Alpha-fetoprotein (Median) | continuous | Median alpha-fetoprotein measurement for this patient's hospitalization | ng/mL | Labs | 0 | | | | | labs | |
| **afp_min** | Alpha-fetoprotein (Min) | continuous | Minimum alpha-fetoprotein measurement for this patient's hospitalization | ng/mL | Labs | 0 | | | | | labs | |
| **afp_sd** | Alpha-fetoprotein (SD) | continuous | Standard deviation of the alpha-fetoprotein measurements during this patient's hospitalization | ng/mL | Labs | 0 | | | | | labs | |
| **ag_levels_25** | Aminoglycoside Levels (25th Percentile) | continuous | Aminoglycoside levels at the 25th percentile of this patient's measurements | µg/mL | Labs | 0 | | | | | labs | |
| **ag_levels_75** | Aminoglycoside Levels (75th Percentile) | continuous | Aminoglycoside levels at the 75th percentile of this patient's measurements | µg/mL | Labs | 0 | | | | | labs | |

| | | | | | | | | | | | |
|---|---|---|---|---|---|---|---|---|---|---|---|
| **ag_levels_count** | Aminoglycoside Levels (Count) | continuous | Count of aminoglycoside measurements for this patient | Number of measurements | Labs | 0 | | | | | labs |
| **ag_levels_first** | Aminoglycoside Levels (First) | continuous | First aminoglycoside measurement for this patient's hospitalization | µg/mL | Labs | 0 | | | | | labs |
| **ag_levels_last** | Aminoglycoside Levels (Last) | continuous | Last aminoglycoside measurement for this patient's hospitalization | µg/mL | Labs | 0 |  |  |  |  | labs |
| **ag_levels_max** | Aminoglycoside Levels (Max) | continuous | Maximum aminoglycoside level for this patient's hospitalization | µg/mL | Labs | 0 | | | | | labs |
| **ag_levels_mean** | Aminoglycoside Levels (Mean) | continuous | Average aminoglycoside level for this patient's hospitalization | µg/mL | Labs | 0 | | | | | labs |
| **ag_levels_median** | Aminoglycoside Levels (Median) | continuous | Median aminoglycoside level for this patient's hospitalization | µg/mL | Labs | 0 | | | | | labs |
| **ag_levels_min** | Aminoglycoside Levels (Min) | continuous | Minimum aminoglycoside levels for this patient's hospitalization | µg/mL | Labs | 0 | | | | | labs |
| **ag_levels_sd** | Aminoglycoside Levels (SD) | continuous | Standard deviation of the aminoglycoside measurements during this patient's hospitalization | µg/mL | Labs | 0 |  |  |  |  | labs |
| **agratio_25** | Albumin-Globulin Ratio (25th Percentile) | continuous | Albumin-Globulin ratio at the 25th percentile of this patient's measurements | | Labs | 0 | | | | | labs |
| **agratio_75** | Albumin-Globulin Ratio (75th Percentile) | continuous | Albumin-Globulin ratio at the 75th percentile of this patient's measurements | | Labs | 0 | | | | | labs |
| **agratio_count** | Albumin-Globulin Ratio (Count) | continuous | Count of Albumin-Globulin ratio measurements for this patient | Number of measurements | Labs | 0 | | | | | labs |
| **agratio_first** | Albumin-Globulin Ratio (First) | continuous | First Albumin-Globulin ratio measurement for this patient's hospitalization | | Labs | 0 | | | | | labs |
| **agratio_last** | Albumin-Globulin Ratio (Last) | continuous | Last Albumin-Globulin ratio measurement for this patient's hospitalization | | Labs | 0 | | | | | labs |
| **agratio_max** | Albumin-Globulin Ratio (Max) | continuous | Maximum Albumin-Globulin ratio measurement for this patient's hospitalization | | Labs | 0 | | | | | labs |
| **agratio_mean** | Albumin-Globulin Ratio (Mean) | continuous | Average Albumin-Globulin ratio measurement for this patient's hospitalization | | Labs | 0 | | | | | labs |
| **agratio_median** | Albumin-Globulin Ratio (Median) | continuous | Median Albumin-Globulin ratio measurement for this patient's hospitalization | | Labs | 0 | | | | | labs |
| **agratio_min** | Albumin-Globulin Ratio (Min) | continuous | Minimum Albumin-Globulin ratio measurement for this patient's hospitalization | | Labs | 0 | | | | | labs |
| **agratio_sd** | Albumin-Globulin Ratio (SD) | continuous | Standard deviation of the Albumin-Globulin ratio measurements during this patient's hospitalization | | Labs | 0 | | | | | labs |
| **aki_stage1** | | | | | Labs | 0 | | | | | |
| **aki_stage2** | | | | | Labs | 0 | | | | | |
| **aki_stage3** | | | | | Labs | 0 | | | | | |
| **albumin_25** | Albumin (25th Percentile) | continuous | Albumin level at the 25th percentile of this patient's measurements | g/dL | Labs | 0 | | | | | labs |
| **albumin_75** | Albumin (75th Percentile) | continuous | Albumin level at the 75th percentile of this patient's measurements | g/dL | Labs | 0 | | | | | labs |
| **albumin_count** | Albumin (Count) | continuous | Count of albumin level measurements for this patient | Number of measurements | Labs | 0 | | | | | labs |
| **albumin_first** | Albumin (First) | continuous | First albumin measurement for this patient's hospitalization | g/dL | Labs | 0 | | | | | labs |
| **albumin_last** | Albumin (Last) | continuous | Last albumin measurement for this patient's hospitalization | g/dL | Labs | 0 | | | | | labs |
| **albumin_max** | Albumin (Max) | continuous | Maximum albumin level measurement for this patient's hospitalization | g/dL | Labs | 0 | | | | | labs |
| **albumin_mean** | Albumin (Mean) | continuous | Average albumin level measurement for this patient's hospitalization | g/dL | Labs | 0 | | | | | labs |
| **albumin_median** | Albumin (Median) | continuous | Median albumin level measurement for this patient's hospitalization | g/dL | Labs | 0 | | | | | labs |
| **albumin_min** | Albumin (Min) | continuous | Minimum albumin level measurement for this patient's hospitalization | g/dL | Labs | 0 | | | | | labs |
| **albumin_sd** | Albumin (SD) | continuous | Standard deviation of the albumin level measurements during this patient's hospitalization | g/dL | Labs | 0 | | | | | labs |
| **albuminurine_25** | Albumin Urine Level (25th Percentile) | continuous | Albumin urine at the 25th percentile of this patient's measurements | mg | Labs | 0 |  |  |  |  | labs |
| **albuminurine_75** | Albumin Urine Level (75th Percentile) | continuous | Albumin urine at the 75th percentile of this patient's measurements | mg | Labs | 0 |  |  |  |  | labs |
| **albuminurine_count** | Albumin Urine Level (Count) | continuous | Count of albumin urine measurements for this patient | Number of measurements | Labs | 0 |  |  |  |  | labs |
| **albuminurine_first** | Albumin Urine Level (First) | continuous | First albumin urine measurement for this patient's hospitalization | mg | Labs | 0 |  |  |  |  | labs |
| **albuminurine_last** | Albumin Urine Level (Last) | continuous | Last albumin urine measurement for this patient's hospitalization | mg | Labs | 0 |  |  |  |  | labs |
| **albuminurine_max** | Albumin Urine Level (Max ) | continuous | Maximum albumin urine measurement for this patient's hospitalization | mg | Labs | 0 |  |  |  |  | labs |
| **albuminurine_mean** | Albumin Urine Level (Mean) | continuous | Average albumin urine measurement for this patient's hospitalization | mg | Labs | 0 |  |  |  |  | labs |
| **albuminurine_median** | Albumin Urine Level (Median) | continuous | Median albumin urine measurement for this patient's hospitalization | mg | Labs | 0 |  |  |  |  | labs |
| **albuminurine_min** | Albumin Urine Level (Min) | continuous | Minimum albumin urine measurement for this patient's hospitalization | mg | Labs | 0 |  |  |  |  | labs |
| **albuminurine_sd** | Albumin Urine Level (SD) | continuous | Standard deviation of the albumin urine measurements during this patient's hospitalization | mg | Labs | 0 |  |  |  |  | labs |
| **alkphos_25** | Alkaline Phosphatase (25th Percentile) | continuous | Alkaline phosphatase amount at the 25th percentile of this patient's measurements | IU/L | Labs | 0 | | | | | labs |
| **alkphos_75** | Alkaline Phosphatase (75th Percentile) | continuous | Alkaline phosphatase amount at the 75th percentile of this patient's measurements | IU/L | Labs | 0 | | | | | labs |
| **alkphos_count** | Alkaline Phosphatase (Count) | continuous | Count of alkaline phosphatase measurements for this patient | Number of measurements | Labs | 0 | | | | | labs |
| **alkphos_first** | Alkaline Phosphatase (First) | continuous | First alkaline phosphatase measurement for this patient's hospitalization | IU/L | Labs | 0 | | | | | labs |
| **alkphos_last** | Alkaline Phosphatase (Last) | continuous | Last alkaline phosphatase measurement for this patient's hospitalization | IU/L | Labs | 0 | | | | | labs |
| **alkphos_max** | Alkaline Phosphatase (Max) | continuous | Maximum alkaline phosphatase measurement for this patient's hospitalization | IU/L | Labs | 0 | | | | | labs |
| **alkphos_mean** | Alkaline Phosphatase (Mean) | continuous | Average alkaline phosphatase measurement for this patient's hospitalization | IU/L | Labs | 0 | | | | | labs |
| **alkphos_median** | Alkaline Phosphatase (Median) | continuous | Median alkaline phosphatase measurement for this patient's hospitalization | IU/L | Labs | 0 | | | | | labs |
| **alkphos_min** | Alkaline Phosphatase (Min) | continuous | Minimum alkaline phosphatase measurement for this patient's hospitalization | IU/L | Labs | 0 | | | | | labs |
| **alkphos_sd** | Alkaline Phosphatase (SD) | continuous | Standard deviation of the alkaline phosphatase measurements during this patient's hospitalization | IU/L | Labs | 0 | | | | | labs |
| **alt_25** | Alanine Aminotransferease (25th Percentile) | continuous | Alanine aminotransferease levels at the 25th percentile of this patient's measurements | U/L | Labs | 0 | | | | | labs |
| **alt_75** | Alanine Aminotransferease (75th Percentile) | continuous | Alanine aminotransferease levels at the 75th percentile of this patient's measurements | U/L | Labs | 0 | | | | | labs |
| **alt_count** | Alanine Aminotransferease (Count) | continuous | Count of alanine aminotransferease measurements for this patient | Number of measurements | Labs | 0 |  |  |  |  | labs |
| **alt_first** | Alanine Aminotransferease (First) | continuous | First alanine aminotransferease measurement for this patient's hospitalization | U/L | Labs | 0 | | | | | labs |
| **alt_last** | Alanine Aminotransferease (Last) | continuous | Last alanine aminotransferease measurement for this patient's hospitalization | U/L | Labs | 0 | | | | | labs |
| **alt_max** | Alanine Aminotransferease (Max) | continuous | Maximum alanine aminotransferease measurement for this patient's hospitalization | U/L | Labs | 0 | | | | | labs |
| **alt_mean** | Alanine Aminotransferease (Mean) | continuous | Average alanine aminotransferease measurement for this patient's hospitalization | U/L | Labs | 0 | | | | | labs |
| **alt_median** | Alanine Aminotransferease (Median) | continuous | Median alanine aminotransferease measurement for this patient's hospitalization | U/L | Labs | 0 | | | | | labs |
| **alt_min** | Alanine Aminotransferease (Min) | continuous | Minimum alanine amino transferease levels measurement for this patient's hospitalization | U/L | Labs | 0 | | | | | labs |
| **alt_sd** | Alanine Aminotransferease (SD) | continuous | Standard deviation of the alanine aminotransferease measurements during this patient's hospitalization | U/L | Labs | 0 | | | | | labs |
| **amikacinlevel_25** | Amikacin level (25th Percentile) | continuous | Amikacin level at the 25th percentile of this patient's measurement | mcg/mL | Labs | 0 | | | | | |
| **amikacinlevel_75** | Amikacin level (75th Percentile) | continuous | Amikacin level at the 75th percentile of this patient's measurement | mcg/mL | Labs | 0 | | | | | |
| **amikacinlevel_count** | Amikacin Level (Count) | continuous | Count of amikacin measurements for this patient | Number of measurements | Labs | 0 | | | | | labs |
| **amikacinlevel_first** | Amikacin level (First) | continuous | First amikacin level measurement for this patient's hospitalization | mcg/mL | Labs | 0 | | | | | |
| **amikacinlevel_last** | Amikacin level (Last) | continuous | Last amikacin level measurement for this patient's hospitalization | mcg/mL | Labs | 0 | | | | | |
| **amikacinlevel_max** | Amikacin level (MaAmikacin levelimum) | continuous | Maximum amikacin level measurement for this patient's hospitalization | mcg/mL | Labs | 0 | | | | | |
| **amikacinlevel_mean** | Amikacin level (Mean) | continuous | Average amikacin level measurement for this patient's hospitalization | mcg/mL | Labs | 0 | | | | | |
| **amikacinlevel_median** | Amikacin level (Median) | continuous | Median amikacin level measurement for this patient's hospitalization | mcg/mL | Labs | 0 | | | | | |
| **amikacinlevel_min** | Amikacin level (Minimum) | continuous | Minimum amikacin level measurement for this patient's hospitalization | mcg/mL | Labs | 0 | | | | | |
| **amikacinlevel_sd** | Amikacin level (Standard Deviation) | continuous | Standard deviation of the amikacin level measurements during this patient's hospitalization | mcg/mL | Labs | 0 | | | | | |
| **ammonia_25** | Ammonia Level (25th Percentile) | continuous | Ammonia level at the 25th percentile of this patient's measurements | µ/dL | Labs | 0 | | | | | labs |
| **ammonia_75** | Ammonia Level (75th Percentile) | continuous | Ammonia level at the 75th percentile of this patient's measurements | µ/dL | Labs | 0 | | | | | labs |
| **ammonia_count** | Ammonia Level (Count) | continuous | Count of ammonia measurements for this patient | Number of measurements | Labs | 0 | | | | | labs |
| **ammonia_first** | Ammonia Level (First) | continuous | First ammonia measurement for this patient's hospitalization | µ/dL | Labs | 0 | | | | | labs |
| **ammonia_last** | Ammonia Level (Last) | continuous | Last ammonia measurement for this patient's hospitalization | µ/dL | Labs | 0 | | | | | labs |
| **ammonia_max** | Ammonia Level (Max) | continuous | Maximum ammonia measurement for this patient's hospitalization | µ/dL | Labs | 0 | | | | | labs |
| **ammonia_mean** | Ammonia Level (Mean) | continuous | Average ammonia measurement for this patient's hospitalization | µ/dL | Labs | 0 | | | | | labs |
| **ammonia_median** | Ammonia Level (Median) | continuous | Median ammonia measurement for this patient's hospitalization | µ/dL | Labs | 0 | | | | | labs |
| **ammonia_min** | Ammonia Level (Min) | continuous | Minimum ammonia measurement for this patient's hospitalization | µ/dL | Labs | 0 | | | | | labs |
| **ammonia_sd** | Ammonia Level (SD) | continuous | Standard deviation of the y measurements during this patient's hospitalization | µ/dL | Labs | 0 | | | | | labs |
| **amylase_25** | Amylase Level (25th Percentile) | continuous | Amylase level at the 25th percentile of this patient's measurements | U/L | Labs | 0 | | | | | labs |
| **amylase_75** | Amylase Level (75th Percentile) | continuous | amylase level at the 75th percentile of this patient's measurements | U/L | Labs | 0 | | | | | labs |
| **amylase_count** | Amylase Level (Count) | continuous | Count of amylase measurements for this patient | Number of measurements | Labs | 0 | | | | | labs |
| **amylase_first** | Amylase Level (First) | continuous | First amylase measurement for this patient's hospitalization | U/L | Labs | 0 | | | | | labs |
| **amylase_last** | Amylase Level (Last) | continuous | Last amylase measurement for this patient's hospitalization | U/L | Labs | 0 | | | | | labs |
| **amylase_max** | Amylase Level (Max) | continuous | Maximum amylase measurement for this patient's hospitalization | U/L | Labs | 0 | | | | | labs |
| **amylase_mean** | Amylase Level (Mean) | continuous | Average amylase measurement for this patient's hospitalization | U/L | Labs | 0 | | | | | labs |
| **amylase_median** | Amylase Level (Median) | continuous | Median amylase measurement for this patient's hospitalization | U/L | Labs | 0 | | | | | labs |
| **amylase_min** | Amylase Level (Min) | continuous | Minimum amylase measurement for this patient's hospitalization | U/L | Labs | 0 | | | | | labs |
| **amylase_sd** | Amylase Level (SD) | continuous | Standard deviation of the amylase measurements during this patient's hospitalization | U/L | Labs | 0 | | | | | labs |
| **ana_first** | Anti Nuclear Antibodies (First) | categorical | Results of the first anti nuclear antibodies test during this patient's hospitalization | Titer | Labs | 0 | | | | | labs |
| **ana_last** | Anti Nuclear Antibodies (Last) | categorical | Results of the last anti nuclear antibodies test during this patient's hospitalization | Titer | Labs | 0 | | | | | labs |
| **anapattern_first** | Anti Nuclear Antibodies Pattern (First) | categorical | Results of the first anti nuclear antibodies pattern test during this patient's hospitalization | Titer | Labs | 0 | | | | | labs |
| **anapattern_last** | Anti Nuclear Antibodies Pattern (Last) | categorical | Results of the last anti nuclear antibodies pattern test during this patient's hospitalization | Titer | Labs | 0 | | | | | labs |
| **anatiter_first** | Anti Nuclear Antibodies Titer (First) | categorical | Results of the first anti nuclear antibodies titer test during this patient's hospitalization | Titer | Labs | 0 | | | | | labs |
| **anatiter_last** | Anti Nuclear Antibodies Titer (Last) | categorical | Results of the last anti nuclear antibodies titer test during this patient's hospitalization | Titer | Labs | 0 | | | | | labs |
| **ancascreen_first** | Anti-Neutrophilic Cytoplasmic Autoantibody Screen (First | categorical | Results of the first anti neutrophilic cytoplasmic autoantibody screen test during this patient's hospitaliza | Titer | Labs | 0 | | | | | labs |
| **ancascreen_last** | Anti-Neutrophilic Cytoplasmic Autoantibody Screen (Last | categorical | Results of the last anti neutrophilic cytoplasmic autoantibody screen test during this patient's hospitaliza | Titer | Labs | 0 | | | | | labs |
| **ancatiter_first** | Anti-Neutrophilic Cytoplasmic Autoantibody Titer (First) | categorical | Results of the first anti neutrophilic cytoplasmic autoantibody titer test during this patient's hospitalizatio | Titer | Labs | 0 | | | | | labs |
| **ancatiter_last** | Anti-Neutrophilic Cytoplasmic Autoantibody Titer (Last) | categorical | Results of the last anti neutrophilic cytoplasmic autoantibody titer test during this patient's hospitalizatio | Titer | Labs | 0 | | | | | labs |
| **aniongap_25** | Anion Gap (25th Percentile) | continuous | Anion gap at the 25th percentile of this patient's measurements | mEq/L | Labs | 0 | | | | | labs |
| **aniongap_75** | Anion Gap (75th Percentile) | continuous | Anion gap at the 75th percentile of this patient's measurements | mEq/L | Labs | 0 | | | | | labs |
| **aniongap_count** | Anion Gap (Count) | continuous | Count of anion gap measurements for this patient | Number of measurements | Labs | 0 | | | | | labs |
| **aniongap_first** | Anion Gap (First) | continuous | First anion gap measurement for this patient's hospitalization | mEq/L | Labs | 0 | | | | | labs |
| **aniongap_last** | Anion Gap (Last) | continuous | Last anion gap measurement for this patient's hospitalization | mEq/L | Labs | 0 | | | | | labs |
| **aniongap_max** | Anion Gap (Max) | continuous | Maximum anion gap measurement for this patient's hospitalization | mEq/L | Labs | 0 | | | | | labs |
| **aniongap_mean** | Anion Gap (Mean) | continuous | Average anion gap measurement for this patient's hospitalization | mEq/L | Labs | 0 | | | | | labs |
| **aniongap_median** | Anion Gap (Median) | continuous | Median anion gap measurement for this patient's hospitalization | mEq/L | Labs | 0 | | | | | labs |
| **aniongap_min** | Anion Gap (Min) | continuous | Minimum anion gap measurement for this patient's hospitalization | mEq/L | Labs | 0 | | | | | labs |
| **aniongap_sd** | Anion Gap (SD) | continuous | Standard deviation of the anion gap measurements during this patient's hospitalization | mEq/L | Labs | 0 | | | | | labs |
| **antiscl_first** | Anti-scleroderma (First) | categorical | Results of the first Anti-scleroderma test during this patient's hospitalization | mEq/L | Labs | 0 | | | | | labs |
| **antiscl_last** | Anti-scleroderma (Last) | categorical | Results of the last Anti-scleroderma test during this patient's hospitalization | mEq/L | Labs | 0 | | | | | labs |
| **ast_25** | Aspartate Aminotransferase Test (25th Percentile) | continuous | Aspartate Aminotransferase Test at the 25th percentile of this patient's measurements | U/L | Labs | 0 | | | | | labs |
| **ast_75** | Aspartate Aminotransferase Test (75th Percentile) | continuous | Aspartate Aminotransferase Test at the 75th percentile of this patient's measurements | U/L | Labs | 0 | | | | | labs |
| **ast_count** | Aspartate Aminotransferase Test (Count) | continuous | Count of aspartate Aminotransferase Test measurements for this patient | Number of measurements | Labs | 0 | | | | | labs |
| **ast_first** | Aspartate Aminotransferase Test (First) | continuous | First aspartate Aminotransferase Test measurement for this patient's hospitalization | U/L | Labs | 0 | | | | | labs |
| **ast_last** | Aspartate Aminotransferase Test (Last) | continuous | Last aspartate Aminotransferase Test measurement for this patient's hospitalization | U/L | Labs | 0 | | | | | labs |
| **ast_max** | Aspartate Aminotransferase Test (Max) | continuous | Maximum aspartate Aminotransferase Test measurement for this patient's hospitalization | U/L | Labs | 0 | | | | | labs |
| **ast_mean** | Aspartate Aminotransferase Test (Mean) | continuous | Average aspartate Aminotransferase Test measurement for this patient's hospitalization | U/L | Labs | 0 | | | | | labs |
| **ast_median** | Aspartate Aminotransferase Test (Median) | continuous | Median aspartate Aminotransferase Test measurement for this patient's hospitalization | U/L | Labs | 0 | | | | | labs |
| **ast_min** | Aspartate Aminotransferase Test (Min) | continuous | Minimum aspartate Aminotransferase Test measurement for this patient's hospitalization | U/L | Labs | 0 | | | | | labs |
| **ast_sd** | Aspartate Aminotransferase Test (SD) | continuous | Standard deviation of the aspartate Aminotransferase Test measurements during this patient's hospitaliz | U/L | Labs | 0 | | | | | labs |
| **atyplymph_25** | Atypical Lymphocites Blood Test (25th Percentile) | continuous | Atypical Lymphocites Blood Test at the 25th percentile of this patient's measurements | µL | Labs | 0 | | | | | |

| | | | | | | | | | | | |
|---|---|---|---|---|---|---|---|---|---|---|---|
| **atyplymph_75** | Atypical Lymphocites Blood Test (75th Percentile) | continuous | Atypical Lymphocites Blood Test at the 75th percentile of this patient's measurements | μL | Labs | 0 | | | | | |
| **atyplymph_count** | Atypical Lymphocites Blood Test (Count) | continuous | Count of Atypical Lymphocites Blood Test measurements for this patient | Number of measurements | Labs | 0 | | | | | |
| **atyplymph_first** | Atypical Lymphocites Blood Test (First) | continuous | First Atypical Lymphocites Blood Test measurement for this patient's hospitalization | μL | Labs | 0 | | | | | |
| **atyplymph_last** | Atypical Lymphocites Blood Test (Last) | continuous | Last Atypical Lymphocites Blood Test measurement for this patient's hospitalization | μL | Labs | 0 | | | | | |
| **atyplymph_max** | Atypical Lymphocites Blood Test (Max) | continuous | Maximum Atypical Lymphocites Blood Test measurement for this patient's hospitalization | μL | Labs | 0 | | | | | |
| **atyplymph_mean** | Atypical Lymphocites Blood Test (Mean) | continuous | Average Atypical Lymphocites Blood Test measurement for this patient's hospitalization | μL | Labs | 0 | | | | | |
| **atyplymph_median** | Atypical Lymphocites Blood Test (Median) | continuous | Median Atypical Lymphocites Blood Test measurement for this patient's hospitalization | μL | Labs | 0 | | | | | |
| **atyplymph_min** | Atypical Lymphocites Blood Test (Min) | continuous | Minimum Atypical Lymphocites Blood Test measurement for this patient's hospitalization | μL | Labs | 0 | | | | | |
| **atyplymph_sd** | Atypical Lymphocites Blood Test (SD) | continuous | Standard deviation of the Atypical Lymphocites Blood Test measurements during this patient's hospitaliz | μL | Labs | 0 | | | | | |
| **b12_25** | Vitamin B12 (Cobalamin) (25th Percentile) | continuous | Vitamin B (Cobalamin) at the 25th percentile of this patient's measurements | pg/mL | Labs | 0 | | | | | labs |
| **b12_75** | Vitamin B12 (Cobalamin) (75th Percentile) | continuous | Vitamin B (Cobalamin) at the 75th percentile of this patient's measurements | pg/mL | Labs | 0 | | | | | labs |
| **b12_count** | Vitamin B12 (Cobalamin) (Count) | continuous | Count of Vitamin B (Cobalamin) measurements for this patient | Number of measurements | Labs | 0 | | | | | labs |
| **b12_first** | Vitamin B12 (Cobalamin) (First) | continuous | First Vitamin B (Cobalamin) measurement for this patient's hospitalization | pg/mL | Labs | 0 | | | | | labs |
| **b12_last** | Vitamin B12 (Cobalamin) (Last) | continuous | Last Vitamin B (Cobalamin) measurement for this patient's hospitalization | pg/mL | Labs | 0 | | | | | labs |
| **b12_max** | Vitamin B12 (Cobalamin) (Max) | continuous | Maximum Vitamin B (Cobalamin) measurement for this patient's hospitalization | pg/mL | Labs | 0 | | | | | labs |
| **b12_mean** | Vitamin B12 (Cobalamin) (Mean) | continuous | Average Vitamin B (Cobalamin) measurement for this patient's hospitalization | pg/mL | Labs | 0 | | | | | labs |
| **b12_median** | Vitamin B12 (Cobalamin) (Median) | continuous | Median Vitamin B (Cobalamin) measurement for this patient's hospitalization | pg/mL | Labs | 0 | | | | | labs |
| **b12_min** | Vitamin B12 (Cobalamin) (Min) | continuous | Minimum Vitamin B (Cobalamin) measurement for this patient's hospitalization | pg/mL | Labs | 0 | | | | | labs |
| **b12_sd** | Vitamin B12 (Cobalamin) (SD) | continuous | Standard deviation of the Vitamin B (Cobalamin) measurements during this patient's hospitalization | pg/mL | Labs | 0 | | | | | labs |
| **bands_25** | Band Neutrophil (25th Percentile) | continuous | Band Neutrophil at the 25th percentile of this patient's measurements | per. mm3 | Labs | 0 | | | | | labs |
| **bands_75** | Band Neutrophil (75th Percentile) | continuous | Band Neutrophil at the 75th percentile of this patient's measurements | per. mm3 | Labs | 0 | | | | | labs |
| **bands_count** | Band Neutrophil (Count) | continuous | Count of band neutrophil measurements for this patient | Number of measurements | Labs | 0 | | | | | labs |
| **bands_first** | Band Neutrophil (First) | continuous | First band neutrophil measurement for this patient's hospitalization | per. mm3 | Labs | 0 | | | | | labs |
| **bands_last** | Band Neutrophil (Last) | continuous | Last band neutrophil measurement for this patient's hospitalization | per. mm3 | Labs | 0 | | | | | labs |
| **bands_max** | Band Neutrophil (Max) | continuous | Maximum band neutrophil measurement for this patient's hospitalization | per. mm3 | Labs | 0 | | | | | labs |
| **bands_mean** | Band Neutrophil (Mean) | continuous | Average band neutrophil measurement for this patient's hospitalization | per. mm3 | Labs | 0 | | | | | labs |
| **bands_median** | Band Neutrophil (Median) | continuous | Median band neutrophil measurement for this patient's hospitalization | per. mm3 | Labs | 0 | | | | | labs |
| **bands_min** | Band Neutrophil (Min) | continuous | Minimum band neutrophil measurement for this patient's hospitalization | per. mm3 | Labs | 0 | | | | | labs |
| **bands_sd** | Band Neutrophil (SD) | continuous | Standard deviation of the band neutrophil measurements during this patient's hospitalization | per. mm3 | Labs | 0 | | | | | labs |
| **baseex_25** | Base Excess (25th Percentile) | continuous | Base Excess at the 25th percentile of this patient's measurements | mEq/L | Labs | 0 | | | | | labs |
| **baseex_75** | Base Excess (75th Percentile) | continuous | Base Excess at the 75th percentile of this patient's measurements | mEq/L | Labs | 0 | | | | | labs |
| **baseex_count** | Base Excess (Count) | continuous | Count of base excess measurements for this patient | Number of measurements | Labs | 0 | | | | | labs |
| **baseex_first** | Base Excess (First) | continuous | First base excess measurement for this patient's hospitalization | mEq/L | Labs | 0 | | | | | labs |
| **baseex_last** | Base Excess (Last) | continuous | Last base excess measurement for this patient's hospitalization | mEq/L | Labs | 0 | | | | | labs |
| **baseex_max** | Base Excess (Max) | continuous | Maximum base excess measurement for this patient's hospitalization | mEq/L | Labs | 0 | | | | | labs |
| **baseex_mean** | Base Excess (Mean) | continuous | Average base excess measurement for this patient's hospitalization | mEq/L | Labs | 0 | | | | | labs |
| **baseex_median** | Base Excess (Median) | continuous | Median base excess measurement for this patient's hospitalization | mEq/L | Labs | 0 | | | | | labs |
| **baseex_min** | Base Excess (Min) | continuous | Minimum base excess measurement for this patient's hospitalization | mEq/L | Labs | 0 | | | | | labs |
| **baseex_sd** | Base Excess (SD) | continuous | Standard deviation of the base excess measurements during this patient's hospitalization | mEq/L | Labs | 0 | | | | | labs |
| **baseline_egfr** | Baseline eGFR | continuous | Baseline Estimated Glomerular Filtration Rate measured for this patient | mL/min./m2 | Labs | 0 | | | | | |
| **basophil_abs_25** | Basophil Absolute (25th Percentile) | continuous | Basophil Absolute at the 25th percentile of this patient's measurements | k/ul | Labs | 0 | | | | | labs |
| **basophil_abs_75** | Basophil Absolute (75th Percentile) | continuous | Basophil Absolute at the 75th percentile of this patient's measurements | k/ul | Labs | 0 | | | | | labs |
| **basophil_abs_count** | Basophil Absolute (Count) | continuous | Count of basophil absolute measurements for this patient | Number of measurements | Labs | 0 | | | | | labs |
| **basophil_abs_first** | Basophil Absolute (First) | continuous | First basophil absolute measurement for this patient's hospitalization | k/ul | Labs | 0 | | | | | labs |
| **basophil_abs_last** | Basophil Absolute (Last) | continuous | Last basophil absolute measurement for this patient's hospitalization | k/ul | Labs | 0 | | | | | labs |
| **basophil_abs_max** | Basophil Absolute (Max) | continuous | Maximum basophil absolute measurement for this patient's hospitalization | k/ul | Labs | 0 | | | | | labs |
| **basophil_abs_mean** | Basophil Absolute (Mean) | continuous | Average basophil absolute measurement for this patient's hospitalization | k/ul | Labs | 0 | | | | | labs |
| **basophil_abs_median** | Basophil Absolute (Median) | continuous | Median basophil absolute measurement for this patient's hospitalization | k/ul | Labs | 0 | | | | | labs |
| **basophil_abs_min** | Basophil Absolute (Min) | continuous | Minimum basophil absolute measurement for this patient's hospitalization | k/ul | Labs | 0 | | | | | labs |
| **basophil_abs_sd** | Basophil Absolute (SD) | continuous | Standard deviation of the basophil absolute measurements during this patient's hospitalization | k/ul | Labs | 0 | | | | | labs |
| **basophil_percent_25** | Basophil Percent (25th Percentile) | continuous | Basophil Percent at the 25th percentile of this patient's measurements | % | Labs | 0 | | | | | labs |
| **basophil_percent_75** | Basophil Percent (75th Percentile) | continuous | Basophil Percent at the 75th percentile of this patient's measurements | % | Labs | 0 | | | | | labs |
| **basophil_percent_count** | Basophil Percent (Count) | continuous | Count of basophil percent measurements for this patient | Number of measurements | Labs | 0 | | | | | labs |
| **basophil_percent_first** | Basophil Percent (First) | continuous | First basophil percent measurement for this patient's hospitalization | % | Labs | 0 | | | | | labs |
| **basophil_percent_last** | Basophil Percent (Last) | continuous | Last basophil percent measurement for this patient's hospitalization | % | Labs | 0 | | | | | labs |
| **basophil_percent_max** | Basophil Percent (Max) | continuous | Maximum basophil percent measurement for this patient's hospitalization | % | Labs | 0 | | | | | labs |
| **basophil_percent_mean** | Basophil Percent (Mean) | continuous | Average basophil percent measurement for this patient's hospitalization | % | Labs | 0 | | | | | labs |
| **basophil_percent_media** | Basophil Percent (Median) | continuous | Median basophil percent measurement for this patient's hospitalization | % | Labs | 0 | | | | | labs |
| **basophil_percent_min** | Basophil Percent (Min) | continuous | Minimum basophil percent measurement for this patient's hospitalization | % | Labs | 0 | | | | | labs |
| **basophil_percent_sd** | Basophil Percent (SD) | continuous | Standard deviation of the basophil percent measurements during this patient's hospitalization | % | Labs | 0 | | | | | labs |
| **benzourine_first** | Benzodiazepine Urine (First) | categorical | Results of the first Benzodiazepine Urine test during this patient's hospitalization | | Labs | 0 | | | | | labs |
| **benzourine_last** | Benzodiazepine Urine (Last) | categorical | Results of the last Benzodiazepine Urine test during this patient's hospitalization | | Labs | 0 | | | | | labs |
| **betaglob_25** | Beta Globulin (25th Percentile) | continuous | Beta globulin level at the 25th percentile of this patient's measurements | g/dL | Labs | 0 | | | | | labs |
| **betaglob_75** | Beta Globulin (75th Percentile) | continuous | Beta globulin level at the 75th percentile of this patient's measurements | g/dL | Labs | 0 | | | | | labs |
| **betaglob_count** | Beta Globulin (Count) | continuous | Count of beta globulin measurements for this patient | Number of measurements | Labs | 0 | | | | | labs |
| **betaglob_first** | Beta Globulin (First) | continuous | First beta globulin measurement for this patient's hospitalization | g/dL | Labs | 0 | | | | | labs |
| **betaglob_last** | Beta Globulin (Last) | continuous | Last beta globulin measurement for this patient's hospitalization | g/dL | Labs | 0 | | | | | labs |
| **betaglob_max** | Beta Globulin (Max) | continuous | Maximum beta globulin measurement for this patient's hospitalization | g/dL | Labs | 0 | | | | | labs |
| **betaglob_mean** | Beta Globulin (Mean) | continuous | Average beta globulin measurement for this patient's hospitalization | g/dL | Labs | 0 | | | | | labs |
| **betaglob_median** | Beta Globulin (Median) | continuous | Median beta globulin measurement for this patient's hospitalization | g/dL | Labs | 0 | | | | | labs |
| **betaglob_min** | Beta Globulin (Min) | continuous | Minimum beta globulin measurement for this patient's hospitalization | g/dL | Labs | 0 | | | | | labs |
| **betaglob_sd** | Beta Globulin (SD) | continuous | Standard deviation of the beta globulin measurements during this patient's hospitalization | g/dL | Labs | 0 | | | | | labs |
| **bhb_25** | Beta-hydroxybutyrate (25th Percentile) | continuous | Beta-hydroxybutyrate levels at the 25th percentile of this patient's measurements | mmol/L | Labs | 0 | | | | | labs |
| **bhb_75** | Beta-hydroxybutyrate (75th Percentile) | continuous | Beta-hydroxybutyrate levels at the 75th percentile of this patient's measurements | mmol/L | Labs | 0 | | | | | labs |
| **bhb_count** | Beta-hydroxybutyrate (Count) | continuous | Count of beta-hydroxybutyrate measurements for this patient | Number of measurements | Labs | 0 | | | | | labs |
| **bhb_first** | Beta-hydroxybutyrate (First) | continuous | First beta-hydroxybutyrate measurement for this patient's hospitalization | mmol/L | Labs | 0 | | | | | labs |
| **bhb_last** | Beta-hydroxybutyrate (Last) | continuous | Last beta-hydroxybutyrate measurement for this patient's hospitalization | mmol/L | Labs | 0 | | | | | labs |
| **bhb_max** | Beta-hydroxybutyrate (Max) | continuous | Maximum beta-hydroxybutyrate measurement for this patient's hospitalization | mmol/L | Labs | 0 | | | | | labs |
| **bhb_mean** | Beta-hydroxybutyrate (Mean) | continuous | Average beta-hydroxybutyrate measurement for this patient's hospitalization | mmol/L | Labs | 0 | | | | | labs |
| **bhb_median** | Beta-hydroxybutyrate (Median) | continuous | Median beta-hydroxybutyrate measurement for this patient's hospitalization | mmol/L | Labs | 0 | | | | | labs |
| **bhb_min** | Beta-hydroxybutyrate (Min) | continuous | Minimum beta-hydroxybutyrate measurement for this patient's hospitalization | mmol/L | Labs | 0 | | | | | labs |
| **bhb_sd** | Beta-hydroxybutyrate (SD) | continuous | Standard deviation of the beta-hydroxybutyrate measurements during this patient's hospitalization | mmol/L | Labs | 0 | | | | | labs |
| **bicarbonate_25** | Bicarbonate (25th Percentile) | continuous | Bicarbonate level at the 25th percentile of this patient's measurements | mEq/L | Labs | 0 | | | | | labs |
| **bicarbonate_75** | Bicarbonate (75th Percentile) | continuous | Bicarbonate level at the 75th percentile of this patient's measurements | mEq/L | Labs | 0 | | | | | labs |
| **bicarbonate_count** | Bicarbonate (Count) | continuous | Count of bicarbonate level measurements for this patient | Number of measurements | Labs | 0 | | | | | labs |
| **bicarbonate_first** | Bicarbonate (First) | continuous | First bicarbonate level measurement for this patient's hospitalization | mEq/L | Labs | 0 | | | | | labs |
| **bicarbonate_last** | Bicarbonate (Last) | continuous | Last bicarbonate level measurement for this patient's hospitalization | mEq/L | Labs | 0 | | | | | labs |
| **bicarbonate_max** | Bicarbonate (Max) | continuous | Maximum bicarbonate level measurement for this patient's hospitalization | mEq/L | Labs | 0 | | | | | labs |
| **bicarbonate_mean** | Bicarbonate (Mean) | continuous | Average bicarbonate level measurement for this patient's hospitalization | mEq/L | Labs | 0 | | | | | labs |
| **bicarbonate_median** | Bicarbonate (Median) | continuous | Median bicarbonate level measurement for this patient's hospitalization | mEq/L | Labs | 0 | | | | | labs |
| **bicarbonate_min** | Bicarbonate (Min) | continuous | Minimum bicarbonate level measurement for this patient's hospitalization | mEq/L | Labs | 0 | | | | | labs |
| **bicarbonate_sd** | Bicarbonate (SD) | continuous | Standard deviation of the bicarbonate level measurements during this patient's hospitalization | mEq/L | Labs | 0 | | | | | labs |
| **bilidirect_25** | Direct Bilirubin (25th Percentile) | continuous | Direct Bilirubin level at the 25th percentile of this patient's measurements | | Labs | 0 | | | | | labs |
| **bilidirect_75** | Direct Bilirubin (75th Percentile) | continuous | Direct Bilirubin level at the 75th percentile of this patient's measurements | | Labs | 0 | | | | | labs |
| **bilidirect_count** | Direct Bilirubin (Count) | continuous | Count of Direct Bilirubin measurements for this patient | Number of measurements | Labs | 0 | | | | | labs |
| **bilidirect_first** | Direct Bilirubin (First) | continuous | First Direct Bilirubin measurement for this patient's hospitalization | | Labs | 0 | | | | | labs |
| **bilidirect_last** | Direct Bilirubin (Last) | continuous | Last Direct Bilirubin measurement for this patient's hospitalization | | Labs | 0 | | | | | labs |
| **bilidirect_max** | Direct Bilirubin (Max) | continuous | Maximum Direct Bilirubin level measurement for this patient's hospitalization | | Labs | 0 | | | | | labs |
| **bilidirect_mean** | Direct Bilirubin (Mean) | continuous | Average Direct Bilirubin level measurement for this patient's hospitalization | | Labs | 0 | | | | | labs |
| **bilidirect_median** | Direct Bilirubin (Median) | continuous | Median Direct Bilirubin level measurement for this patient's hospitalization | | Labs | 0 | | | | | labs |
| **bilidirect_min** | Direct Bilirubin (Min) | continuous | Minimum Direct Bilirubin level measurement for this patient's hospitalization | | Labs | 0 | | | | | labs |
| **bilidirect_sd** | Direct Bilirubin (SD) | continuous | Standard deviation of the Direct Bilirubin level measurements during this patient's hospitalization | | Labs | 0 | | | | | labs |
| **biliindirect_25** | | | | | Labs | 0 | | | | | |
| **biliindirect_75** | | | | | Labs | 0 | | | | | |
| **biliindirect_count** | | | | | Labs | 0 | | | | | |
| **biliindirect_first** | | | | | Labs | 0 | | | | | |
| **biliindirect_last** | | | | | Labs | 0 | | | | | |
| **biliindirect_max** | | | | | Labs | 0 | | | | | |
| **biliindirect_mean** | | | | | Labs | 0 | | | | | |
| **biliindirect_median** | | | | | Labs | 0 | | | | | |
| **biliindirect_min** | | | | | Labs | 0 | | | | | |
| **biliindirect_sd** | | | | | Labs | 0 | | | | | |
| **bilitotal_25** | Total Bilirubin (25th Percentile) | continuous | Total bilirubin at the 25th percentile of this patient's measurements | mg/dL | Labs | 0 | | | | | labs |
| **bilitotal_75** | Total Bilirubin (75th Percentile) | continuous | Total bilirubin at the 75th percentile of this patient's measurements | mg/dL | Labs | 0 | | | | | labs |
| **bilitotal_count** | Total Bilirubin (Count) | continuous | Count of total bilirubin measurements for this patient | Number of measurements | Labs | 0 | | | | | labs |
| **bilitotal_first** | Total Bilirubin (First) | continuous | First bilirubin (total) measurement for this patient's hospitalization | mg/dL | Labs | 0 | | | | | labs |
| **bilitotal_last** | Total Bilirubin (Last) | continuous | Last bilirubin (total) measurement for this patient's hospitalization | mg/dL | Labs | 0 | | | | | labs |
| **bilitotal_max** | Total Bilirubin (Max) | continuous | Maximum total bilirubin measurement for this patient's hospitalization | mg/dL | Labs | 0 | | | | | labs |
| **bilitotal_mean** | Total Bilirubin (Mean) | continuous | Average total bilirubin measurement for this patient's hospitalization | mg/dL | Labs | 0 | | | | | labs |
| **bilitotal_median** | Total Bilirubin (Median) | continuous | Median total bilirubin measurement for this patient's hospitalization | mg/dL | Labs | 0 | | | | | labs |
| **bilitotal_min** | Total Bilirubin (Min) | continuous | Minimum total bilirubin measurement for this patient's hospitalization | mg/dL | Labs | 0 | | | | | labs |
| **bilitotal_sd** | Total Bilirubin (SD) | continuous | Standard deviation of the total bilirubin measurements during this patient's hospitalization | mg/dL | Labs | 0 | | | | | labs |
| **bkplasma_first** | BK Plasma (First) | categorical | Results of the first BK virus in blood plasma test during this patient's hospitalization | | Labs | 0 | | | | | labs |
| **bkplasma_last** | BK Plasma (Last) | categorical | Results of the last BK virus in blood plasma test during this patient's hospitalization | | Labs | 0 | | | | | labs |

| | | | | | | | | | | | |
|---|---|---|---|---|---|---|---|---|---|---|---|
| **bkquant_first** | BK Virus Quantitative (First) | categorical | Results of the first BK virus Quantitative test during this patient's hospitalization | | Labs | 0 | | | | | |
| **bkquant_last** | BK Virus Quantitative (Last) | categorical | Results of the last BK virus Quantitative test during this patient's hospitalization | | Labs | 0 | | | | | |
| **bkquantlog_first** | BK Virus Quantitative Log (First) | categorical | Results of the first BK virus Quantitative log during this patient's hospitalization | | Labs | 0 | | | | | |
| **bkquantlog_last** | BK Virus Quantitative Log (Last) | categorical | Results of the last BK virus Quantitative log during this patient's hospitalization | | Labs | 0 | | | | | |
| **bkurine_first** | BK Urine (First) | categorical | Results of the first BK virus in urine test during this patient's hospitalization | | Labs | 0 | | | | | labs |
| **bkurine_last** | BK Urine (Last) | categorical | Results of the last BK virus in urine test during this patient's hospitalization | | Labs | 0 | | | | | labs |
| **bkurinelog_first** | BK Urine Log (First) | categorical | Results of the first BK virus in urine log during this patient's hospitalization | | Labs | 0 | | | | | |
| **bkurinelog_last** | BK Urine Log (Last) | categorical | Results of the last BK virus in urine log during this patient's hospitalization | | Labs | 0 | | | | | |
| **blastpercent_25** | Blast Percent (25th Percentile) | continuous | Blast percent at the 25th percentile of this patient's measurements | % | Labs | 0 | | | | | labs |
| **blastpercent_75** | Blast Percent (75th Percentile) | continuous | Blast percent at the 75th percentile of this patient's measurements | % | Labs | 0 | | | | | labs |
| **blastpercent_count** | Blast Percent (Count) | continuous | Count of blast percent measurements for this patient | Number of measurements | Labs | 0 | | | | | labs |
| **blastpercent_first** | Blast Percent (First) | continuous | First blast percent measurement for this patient's hospitalization | % | Labs | 0 | | | | | labs |
| **blastpercent_last** | Blast Percent (Last) | continuous | Last blast percent measurement for this patient's hospitalization | % | Labs | 0 | | | | | labs |
| **blastpercent_max** | Blast Percent (Max) | continuous | Maximum blast percent measurement for this patient's hospitalization | % | Labs | 0 | | | | | labs |
| **blastpercent_mean** | Blast Percent (Mean) | continuous | Average blast percent measurement for this patient's hospitalization | % | Labs | 0 | | | | | labs |
| **blastpercent_median** | Blast Percent (Median) | continuous | Median blast percent measurement for this patient's hospitalization | % | Labs | 0 | | | | | labs |
| **blastpercent_min** | Blast Percent (Min) | continuous | Minimum blast percent measurement for this patient's hospitalization | % | Labs | 0 | | | | | labs |
| **blastpercent_sd** | Blast Percent (SD) | continuous | Standard deviation of the blast percent measurements during this patient's hospitalization | % | Labs | 0 | | | | | labs |
| **bloodcult_first** | Blood Culture (First) | categorical | Results of the first blood culture test during this patient's hospitalization | | Labs | 0 | | | | | labs |
| **bloodcult_last** | Blood Culture (Last) | categorical | Results of the last blood culture test during this patient's hospitalization | | Labs | 0 | | | | | labs |
| **bloodstool_first** | Blood Stool (First) | categorical | Results of the first blood stool test during this patient's hospitalization | | Labs | 0 | | | | | labs |
| **bloodstool_last** | Blood Stool (Last) | categorical | Results of the last blood stool test during this patient's hospitalization | | Labs | 0 | | | | | labs |
| **bnp_25** | B-Type Natriuretic Peptide (25th Percentile) | continuous | BNP level at the 25th percentile of this patient's measurements | pg/ml | Labs | 0 | | | | | labs |
| **bnp_75** | B-Type Natriuretic Peptide (75th Percentile) | continuous | BNP level at the 75th percentile of this patient's measurements | pg/ml | Labs | 0 | | | | | labs |
| **bnp_count** | B-Type Natriuretic Peptide (Count) | continuous | Count of BNP level measurements for this patient | Number of measurements | Labs | 0 | | | | | labs |
| **bnp_first** | B-Type Natriuretic Peptide (First) | continuous | First BNP level measurement for this patient's hospitalization | pg/ml | Labs | 0 | | | | | labs |
| **bnp_last** | B-Type Natriuretic Peptide (Last) | continuous | Last BNP level measurement for this patient's hospitalization | pg/ml | Labs | 0 | | | | | labs |
| **bnp_max** | B-Type Natriuretic Peptide (Max) | continuous | Maximum BNP level measurement for this patient's hospitalization | pg/ml | Labs | 0 | | | | | labs |
| **bnp_mean** | B-Type Natriuretic Peptide (Mean) | continuous | Average BNP level measurement for this patient's hospitalization | pg/ml | Labs | 0 | | | | | labs |
| **bnp_median** | B-Type Natriuretic Peptide (Median) | continuous | Median BNP level measurement for this patient's hospitalization | pg/ml | Labs | 0 | | | | | labs |
| **bnp_min** | B-Type Natriuretic Peptide (Min) | continuous | Minimum BNP level measurement for this patient's hospitalization | pg/ml | Labs | 0 | | | | | labs |
| **bnp_sd** | B-Type Natriuretic Peptide (SD) | continuous | Standard deviation of the BNP level measurements during this patient's hospitalization | pg/ml | Labs | 0 | | | | | labs |
| **bnppoc_first** | Point of Care BNP (First) | categorical | First Point of Care BNP (B-type Natriuretic Peptide) during this patient's hospitalization | | Labs | 0 | | | | | labs |
| **bnppoc_last** | Point of Care BNP (Last) | categorical | Last Point of Care BNP (B-type Natriuretic Peptide) during this patient's hospitalization | | Labs | 0 | | | | | labs |
| **bun_25** | Blood Urea Nitrogen (25th Percentile) | continuous | Blood urea nitrogen level at the 25th percentile of this patient's measurements | mg/dL | Labs | 0 | | | | | labs |
| **bun_75** | Blood Urea Nitrogen (75th Percentile) | continuous | Blood urea nitrogen level at the 75th percentile of this patient's measurements | mg/dL | Labs | 0 | | | | | labs |
| **bun_count** | Blood Urea Nitrogen (Count) | continuous | Count of blood urea nitrogen level measurements for this patient | Number of measurements | Labs | 0 | | | | | labs |
| **bun_first** | Blood Urea Nitrogen (First) | continuous | First blood urea nitrogen level measurement for this patient's hospitalization | mg/dL | Labs | 0 | | | | | labs |
| **bun_last** | Blood Urea Nitrogen (Last) | continuous | Last blood urea nitrogen level measurement for this patient's hospitalization | mg/dL | Labs | 0 | | | | | labs |
| **bun_max** | Blood Urea Nitrogen (Max) | continuous | Maximum blood urea nitrogen level measurement for this patient's hospitalization | mg/dL | Labs | 0 | | | | | labs |
| **bun_mean** | Blood Urea Nitrogen (Mean) | continuous | Average blood urea nitrogen level measurement for this patient's hospitalization | mg/dL | Labs | 0 | | | | | labs |
| **bun_median** | Blood Urea Nitrogen (Median) | continuous | Median blood urea nitrogen level measurement for this patient's hospitalization | mg/dL | Labs | 0 | | | | | labs |
| **bun_min** | Blood Urea Nitrogen (Min) | continuous | Minimum blood urea nitrogen level measurement for this patient's hospitalization | mg/dL | Labs | 0 | | | | | labs |
| **bun_sd** | Blood Urea Nitrogen (SD) | continuous | Standard deviation of the blood urea nitrogen level measurements during this patient's hospitalization | mg/dL | Labs | 0 | | | | | labs |
| **buncreatratio_25** | BUN to Creatinine Ratio (25th Percentile) | continuous | BUN to creatinine ratio at the 25th percentile of this patient's measurements | | Labs | 0 | | | | | labs |
| **buncreatratio_75** | BUN to Creatinine Ratio (75th Percentile) | continuous | BUN to creatinine ratio at the 75th percentile of this patient's measurements | | Labs | 0 | | | | | labs |
| **buncreatratio_count** | BUN to Creatinine Ratio (Count) | continuous | Count of BUN to creatinine ratio measurements for this patient | Number of measurements | Labs | 0 | | | | | labs |
| **buncreatratio_first** | BUN to Creatinine Ratio (First) | continuous | First BUN to creatinine ratio measurement for this patient's hospitalization | | Labs | 0 | | | | | labs |
| **buncreatratio_last** | BUN to Creatinine Ratio (Last) | continuous | Last BUN to creatinine ratio measurement for this patient's hospitalization | | Labs | 0 | | | | | labs |
| **buncreatratio_max** | BUN to Creatinine Ratio (Max) | continuous | Maximum BUN to creatinine ratio measurement for this patient's hospitalization | | Labs | 0 | | | | | labs |
| **buncreatratio_mean** | BUN to Creatinine Ratio (Mean) | continuous | Average BUN to creatinine ratio measurement for this patient's hospitalization | | Labs | 0 | | | | | labs |
| **buncreatratio_median** | BUN to Creatinine Ratio (Median) | continuous | Median BUN to creatinine ratio measurement for this patient's hospitalization | | Labs | 0 | | | | | labs |
| **buncreatratio_min** | BUN to Creatinine Ratio (Min) | continuous | Minimum BUN to creatinine ratio measurement for this patient's hospitalization | | Labs | 0 | | | | | labs |
| **buncreatratio_sd** | BUN to Creatinine Ratio (SD) | continuous | Standard deviation of the BUN to creatinine ratio measurements during this patient's hospitalization | | Labs | 0 | | | | | labs |
| **c3complement_25** | Complement C3 (25th Percentile) | continuous | Complement c3 level at the 25th percentile of this patient's measurements | mg/dL | Labs | 0 | | | | | labs |
| **c3complement_75** | Complement C3 (75th Percentile) | continuous | Complement c3 level at the 75th percentile of this patient's measurements | mg/dL | Labs | 0 | | | | | labs |
| **c3complement_count** | Complement C3 (Count) | continuous | Count of complement c3 level measurements for this patient | Number of measurements | Labs | 0 | | | | | labs |
| **c3complement_first** | Complement C3 (First) | continuous | First complement c3 level measurement for this patient's hospitalization | mg/dL | Labs | 0 | | | | | labs |
| **c3complement_last** | Complement C3 (Last) | continuous | Last complement c3 level measurement for this patient's hospitalization | mg/dL | Labs | 0 | | | | | labs |
| **c3complement_max** | Complement C3 (Max) | continuous | Maximum complement c3 level measurement for this patient's hospitalization | mg/dL | Labs | 0 | | | | | labs |
| **c3complement_mean** | Complement C3 (Mean) | continuous | Average complement c3 level measurement for this patient's hospitalization | mg/dL | Labs | 0 | | | | | labs |
| **c3complement_median** | Complement C3 (Median) | continuous | Median complement c3 level measurement for this patient's hospitalization | mg/dL | Labs | 0 | | | | | labs |
| **c3complement_min** | Complement C3 (Min) | continuous | Minimum complement c3 level measurement for this patient's hospitalization | mg/dL | Labs | 0 | | | | | labs |
| **c3complement_sd** | Complement C3 (SD) | continuous | Standard deviation of the complement c3 level measurements during this patient's hospitalization | mg/dL | Labs | 0 | | | | | labs |
| **c3nephritic_first** | C3 Nephritic Factor (First) | categorical | First C3 Nephritic Factor measurement for this patient's hospitalization | | Labs | 0 | | | | | |
| **c3nephritic_last** | C3 Nephritic Factor (Last) | categorical | Last C3 Nephritic Factor measurement for this patient's hospitalization | | Labs | 0 | | | | | |
| **c4complement_25** | Complement C4 (25th Percentile) | continuous | Complement c4 level at the 25th percentile of this patient's measurements | mg/dL | Labs | 0 | | | | | labs |
| **c4complement_75** | Complement C4 (75th Percentile) | continuous | Complement c4 level at the 75th percentile of this patient's measurements | mg/dL | Labs | 0 | | | | | labs |
| **c4complement_count** | Complement C4 (Count) | continuous | Count of complement c4 level measurements for this patient | Number of measurements | Labs | 0 | | | | | labs |
| **c4complement_first** | Complement C4 (First) | continuous | First complement c4 level measurement for this patient's hospitalization | mg/dL | Labs | 0 | | | | | labs |
| **c4complement_last** | Complement C4 (Last) | continuous | Last complement c4 level measurement for this patient's hospitalization | mg/dL | Labs | 0 | | | | | labs |
| **c4complement_max** | Complement C4 (Max) | continuous | Maximum complement c4 level measurement for this patient's hospitalization | mg/dL | Labs | 0 | | | | | labs |
| **c4complement_mean** | Complement C4 (Mean) | continuous | Average complement c4 level measurement for this patient's hospitalization | mg/dL | Labs | 0 | | | | | labs |
| **c4complement_median** | Complement C4 (Median) | continuous | Median complement c4 level measurement for this patient's hospitalization | mg/dL | Labs | 0 | | | | | labs |
| **c4complement_min** | Complement C4 (Min) | continuous | Minimum complement c4 level measurement for this patient's hospitalization | mg/dL | Labs | 0 | | | | | labs |
| **c4complement_sd** | Complement C4 (SD) | continuous | Standard deviation of the complement c4 level measurements during this patient's hospitalization | mg/dL | Labs | 0 | | | | | labs |
| **ca125_25** | Cancer Antigen 125 (25th Percentile) | continuous | Cancer antigen 125 level at the 25th percentile of this patient's measurements | U/mL | Labs | 0 | | | | | labs |
| **ca125_75** | Cancer Antigen 125 (75th Percentile) | continuous | Cancer antigen 125 level at the 75th percentile of this patient's measurements | U/mL | Labs | 0 | | | | | labs |
| **ca125_count** | Cancer Antigen 125 (Count) | continuous | Count of cancer antigen 125 level measurements for this patient | Number of measurements | Labs | 0 | | | | | labs |
| **ca125_first** | Cancer Antigen 125 (First) | continuous | First cancer antigen 125 level measurement for this patient's hospitalization | U/mL | Labs | 0 | | | | | labs |
| **ca125_last** | Cancer Antigen 125 (Last) | continuous | Last cancer antigen 125 level measurement for this patient's hospitalization | U/mL | Labs | 0 | | | | | labs |
| **ca125_max** | Cancer Antigen 125 (Max) | continuous | Maximum cancer antigen 125 level measurement for this patient's hospitalization | U/mL | Labs | 0 | | | | | labs |
| **ca125_mean** | Cancer Antigen 125 (Mean) | continuous | Average cancer antigen 125 level measurement for this patient's hospitalization | U/mL | Labs | 0 | | | | | labs |
| **ca125_median** | Cancer Antigen 125 (Median) | continuous | Median cancer antigen 125 level measurement for this patient's hospitalization | U/mL | Labs | 0 | | | | | labs |
| **ca125_min** | Cancer Antigen 125 (Min) | continuous | Minimum cancer antigen 125 level measurement for this patient's hospitalization | U/mL | Labs | 0 | | | | | labs |
| **ca125_sd** | Cancer Antigen 125 (SD) | continuous | Standard deviation of the cancer antigen 125 level measurements during this patient's hospitalization | U/mL | Labs | 0 | | | | | labs |
| **ca199_25** | CA19-9 tumor marker (25th Percentile) | continuous | CA19-9 tumor marker level at the 25th percentile of this patient's measurements | U/mL | Labs | 0 | | | | | labs |
| **ca199_75** | CA19-9 tumor marker (75th Percentile) | continuous | CA19-9 tumor marker level at the 75th percentile of this patient's measurements | U/mL | Labs | 0 | | | | | labs |
| **ca199_count** | CA19-9 tumor marker (Count) | continuous | Count of CA19-9 tumor marker level measurements for this patient | Number of measurements | Labs | 0 | | | | | labs |
| **ca199_first** | CA19-9 tumor marker (First) | continuous | First CA19-9 tumor marker level measurement for this patient's hospitalization | U/mL | Labs | 0 | | | | | labs |
| **ca199_last** | CA19-9 tumor marker (Last) | continuous | Last CA19-9 tumor marker level measurement for this patient's hospitalization | U/mL | Labs | 0 | | | | | labs |
| **ca199_max** | CA19-9 tumor marker (Max) | continuous | Maximum CA19-9 tumor marker level measurement for this patient's hospitalization | U/mL | Labs | 0 | | | | | labs |
| **ca199_mean** | CA19-9 tumor marker (Mean) | continuous | Average CA19-9 tumor marker level measurement for this patient's hospitalization | U/mL | Labs | 0 | | | | | labs |
| **ca199_median** | CA19-9 tumor marker (Median) | continuous | Median CA19-9 tumor marker level measurement for this patient's hospitalization | U/mL | Labs | 0 | | | | | labs |
| **ca199_min** | CA19-9 tumor marker (Min) | continuous | Minimum CA19-9 tumor marker level measurement for this patient's hospitalization | U/mL | Labs | 0 | | | | | labs |
| **ca199_sd** | CA19-9 tumor marker (SD) | continuous | Standard deviation of the CA19-9 tumor marker level measurements during this patient's hospitalization | U/mL | Labs | 0 | | | | | labs |
| **ca2729_25** | Cancer Antigen 27-29 (25th Percentile) | continuous | CA 27-29 level at the 25th percentile of this patient's measurements | U/mL | Labs | 0 | | | | | labs |
| **ca2729_75** | Cancer Antigen 27-29 (75th Percentile) | continuous | CA 27-29 level at the 75th percentile of this patient's measurements | U/mL | Labs | 0 | | | | | labs |
| **ca2729_count** | Cancer Antigen 27-29 (Count) | continuous | Count of CA 27-29 level measurements for this patient | Number of measurements | Labs | 0 | | | | | labs |
| **ca2729_first** | Cancer Antigen 27-29 (First) | continuous | First CA 27-29 level measurement for this patient's hospitalization | U/mL | Labs | 0 | | | | | labs |
| **ca2729_last** | Cancer Antigen 27-29 (Last) | continuous | Last CA 27-29 level measurement for this patient's hospitalization | U/mL | Labs | 0 | | | | | labs |
| **ca2729_max** | Cancer Antigen 27-29 (Max) | continuous | Maximum CA 27-29 level measurement for this patient's hospitalization | U/mL | Labs | 0 | | | | | labs |
| **ca2729_mean** | Cancer Antigen 27-29 (Mean) | continuous | Average CA 27-29 level measurement for this patient's hospitalization | U/mL | Labs | 0 | | | | | labs |
| **ca2729_median** | Cancer Antigen 27-29 (Median) | continuous | Median CA 27-29 level measurement for this patient's hospitalization | U/mL | Labs | 0 | | | | | labs |
| **ca2729_min** | Cancer Antigen 27-29 (Min) | continuous | Minimum CA 27-29 level measurement for this patient's hospitalization | U/mL | Labs | 0 | | | | | labs |
| **ca2729_sd** | Cancer Antigen 27-29 (SD) | continuous | Standard deviation of the CA 27-29 level measurements during this patient's hospitalization | U/mL | Labs | 0 | | | | | labs |
| **cacaineurine_first** | | | | | Labs | 0 | | | | | |
| **cacaineurine_last** | | | | | Labs | 0 | | | | | |
| **calcium_25** | Calcium (25th Percentile) | continuous | Calcium level at the 25th percentile of this patient's measurements | mg/dL | Labs | 0 | | | | | labs |
| **calcium_75** | Calcium (75th Percentile) | continuous | Calcium level at the 75th percentile of this patient's measurements | mg/dL | Labs | 0 | | | | | labs |
| **calcium_count** | Calcium (Count) | continuous | Count of calcium level measurements for this patient | Number of measurements | Labs | 0 | | | | | labs |
| **calcium_first** | Calcium (First) | continuous | First calcium level measurement for this patient's hospitalization | mg/dL | Labs | 0 | | | | | labs |
| **calcium_last** | Calcium (Last) | continuous | Last calcium level measurement for this patient's hospitalization | mg/dL | Labs | 0 | | | | | labs |
| **calcium_max** | Calcium (Max) | continuous | Maximum calcium level measurement for this patient's hospitalization | mg/dL | Labs | 0 | | | | | labs |
| **calcium_mean** | Calcium (Mean) | continuous | Average calcium level measurement for this patient's hospitalization | mg/dL | Labs | 0 | | | | | labs |
| **calcium_median** | Calcium (Median) | continuous | Median calcium level measurement for this patient's hospitalization | mg/dL | Labs | 0 | | | | | labs |
| **calcium_min** | Calcium (Min) | continuous | Minimum calcium level measurement for this patient's hospitalization | mg/dL | Labs | 0 | | | | | labs |
| **calcium_sd** | Calcium (SD) | continuous | Standard deviation of the calcium level measurements during this patient's hospitalization | mg/dL | Labs | 0 | | | | | labs |
| **carboxyhgb_25** | Carboxyhemoglobin (25th Percentile) | continuous | Carboxyhemoglobin at the 25th percentile of this patient's measurements | % | Labs | 0 | | | | | labs |
| **carboxyhgb_75** | Carboxyhemoglobin (75th Percentile) | continuous | Carboxyhemoglobin at the 75th percentile of this patient's measurements | % | Labs | 0 | | | | | labs |
| **carboxyhgb_count** | Carboxyhemoglobin (Count) | continuous | Count of carboxyhemoglobin measurements for this patient | Number of measurements | Labs | 0 | | | | | labs |
| **carboxyhgb_first** | Carboxyhemoglobin (First) | continuous | First carboxyhemoglobin measurement for this patient's hospitalization | % | Labs | 0 | | | | | labs |
| **carboxyhgb_last** | Carboxyhemoglobin (Last) | continuous | Last carboxyhemoglobin measurement for this patient's hospitalization | % | Labs | 0 | | | | | labs |
| **carboxyhgb_max** | Carboxyhemoglobin (Max) | continuous | Maximum carboxyhemoglobin measurement for this patient's hospitalization | % | Labs | 0 | | | | | labs |

| | | | | | | | | | | | |
|---|---|---|---|---|---|---|---|---|---|---|---|
| **carboxyhgb_mean** | Carboxyhemoglobin (Mean) | continuous | Average carboxyhemoglobin measurement for this patient's hospitalization | % | Labs | 0 | | | | | labs |
| **carboxyhgb_median** | Carboxyhemoglobin (Median) | continuous | Median carboxyhemoglobin measurement for this patient's hospitalization | % | Labs | 0 | | | | | labs |
| **carboxyhgb_min** | Carboxyhemoglobin (Min) | continuous | Minimum carboxyhemoglobin measurement for this patient's hospitalization | % | Labs | 0 | | | | | labs |
| **carboxyhgb_sd** | Carboxyhemoglobin (SD) | continuous | Standard deviation of the carboxyhemoglobin measurements during this patient's hospitalization | % | Labs | 0 | | | | | labs |
| **cbdurine_first** | Common Bile Duct Urine (First) | categorical | Results of the first Common Bile Duct in urine test during this patient's hospitalization | | Labs | 0 | | | | | labs |
| **cbdurine_last** | Common Bile Duct Urine (Last) | categorical | Results of the last Common Bile Duct in urine test during this patient's hospitalization | | Labs | 0 | | | | | labs |
| **cdfcytoassay_first** | Clostridium difficile Cyto-assay (First) | categorical | Results of the first Clostridium difficile in Cyto-assay during this patient's hospitalization | | Labs | 0 | | | | | labs |
| **cdfcytoassay_last** | Clostridium difficile Cyto-assay (Last) | categorical | Results of the last Clostridium difficile in Cyto-assay during this patient's hospitalization | | Labs | 0 | | | | | labs |
| **cdfgdhantigen_first** | Clostridium difficile glutamate dehydrogenase Antigen (Fi | categorical | Results of the first Clostridium difficile glutamate dehydrogenase Antigen test during this patient's hospitalization | | Labs | 0 | | | | | labs |
| **cdfgdhantigen_last** | Clostridium difficile glutamate dehydrogenase Antigen (La | categorical | Results of the last Clostridium difficile glutamate dehydrogenase Antigen test during this patient's hospitalization | | Labs | 0 | | | | | labs |
| **cdfpcr_first** | Clostridium difficile PCR (First) | categorical | Results of the first Clostridium difficile PCR during this patient's hospitalization | | Labs | 0 | | | | | labs |
| **cdfpcr_last** | Clostridium difficile PCR (Last) | categorical | Results of the last Clostridium difficile PCR during this patient's hospitalization | | Labs | 0 | | | | | labs |
| **cdfrapidtoxin_first** | Clostridium difficile Rapid Toxin (First) | categorical | Results of the first Clostridium difficile Rapid Toxin test during this patient's hospitalization | | Labs | 0 | | | | | labs |
| **cdfrapidtoxin_last** | Clostridium difficile Rapid Toxin (Last) | categorical | Results of the last Clostridium difficile Rapid Toxin test during this patient's hospitalization | | Labs | 0 | | | | | labs |
| **cea_25** | Carcinoembryonic Antigen (25th Percentile) | continuous | Carcinoembryonic antigen at the 25th percentile of this patient's measurements | ng/mL | Labs | 0 | | | | | labs |
| **cea_75** | Carcinoembryonic Antigen (75th Percentile) | continuous | Carcinoembryonic antigen at the 75th percentile of this patient's measurements | ng/mL | Labs | 0 | | | | | labs |
| **cea_count** | Carcinoembryonic Antigen (Count) | continuous | Count of carcinoembryonic antigen measurements for this patient | Number of measurements | Labs | 0 | | | | | labs |
| **cea_first** | Carcinoembryonic Antigen (First) | continuous | First carcinoembryonic antigen measurement for this patient's hospitalization | ng/mL | Labs | 0 | | | | | labs |
| **cea_last** | Carcinoembryonic Antigen (Last) | continuous | Last carcinoembryonic antigen measurement for this patient's hospitalization | ng/mL | Labs | 0 | | | | | labs |
| **cea_max** | Carcinoembryonic Antigen (Max) | continuous | Maximum carcinoembryonic antigen measurement for this patient's hospitalization | ng/mL | Labs | 0 | | | | | labs |
| **cea_mean** | Carcinoembryonic Antigen (Mean) | continuous | Average carcinoembryonic antigen measurement for this patient's hospitalization | ng/mL | Labs | 0 | | | | | labs |
| **cea_median** | Carcinoembryonic Antigen (Median) | continuous | Median carcinoembryonic antigen measurement for this patient's hospitalization | ng/mL | Labs | 0 | | | | | labs |
| **cea_min** | Carcinoembryonic Antigen (Min) | continuous | Minimum carcinoembryonic antigen measurement for this patient's hospitalization | ng/mL | Labs | 0 | | | | | labs |
| **cea_sd** | Carcinoembryonic Antigen (SD) | continuous | Standard deviation of the carcinoembryonic antigen measurements during this patient's hospitalization | ng/mL | Labs | 0 | | | | | labs |
| **chloride_25** | Chloride (25th Percentile) | continuous | Chloride at the 25th percentile of this patient's measurements | mEq/L | Labs | 0 | | | | | labs |
| **chloride_75** | Chloride (75th Percentile) | continuous | Chloride at the 75th percentile of this patient's measurements | mEq/L | Labs | 0 | | | | | labs |
| **chloride_count** | Chloride (Count) | continuous | Count of chloride measurements for this patient | Number of measurements | Labs | 0 | | | | | labs |
| **chloride_first** | Chloride (First) | continuous | First chloride measurement for this patient's hospitalization | mEq/L | Labs | 0 | | | | | labs |
| **chloride_last** | Chloride (Last) | continuous | Last chloride measurement for this patient's hospitalization | mEq/L | Labs | 0 | | | | | labs |
| **chloride_max** | Chloride (Max) | continuous | Maximum chloride measurement for this patient's hospitalization | mEq/L | Labs | 0 | | | | | labs |
| **chloride_mean** | Chloride (Mean) | continuous | Average chloride measurement for this patient's hospitalization | mEq/L | Labs | 0 | | | | | labs |
| **chloride_median** | Chloride (Median) | continuous | Median chloride measurement for this patient's hospitalization | mEq/L | Labs | 0 | | | | | labs |
| **chloride_min** | Chloride (Min) | continuous | Minimum chloride measurement for this patient's hospitalization | mEq/L | Labs | 0 | | | | | labs |
| **chloride_sd** | Chloride (SD) | continuous | Standard deviation of the chloride measurements during this patient's hospitalization | mEq/L | Labs | 0 | | | | | labs |
| **chlorideurine_25** | Chloride Urine (25th Percentile) | continuous | Chloride Urine at the 25th percentile of this patient's measurements | mEq/L | Labs | 0 | | | | | labs |
| **chlorideurine_75** | Chloride Urine (75th Percentile) | continuous | Chloride Urine at the 75th percentile of this patient's measurements | mEq/L | Labs | 0 | | | | | labs |
| **chlorideurine_count** | Chloride Urine (Count) | continuous | Count of chloride urine measurements for this patient | Number of measurements | Labs | 0 | | | | | labs |
| **chlorideurine_first** | Chloride Urine (First) | continuous | First chloride urine measurement for this patient's hospitalization | mEq/L | Labs | 0 | | | | | labs |
| **chlorideurine_last** | Chloride Urine (Last) | continuous | Last chloride urine measurement for this patient's hospitalization | mEq/L | Labs | 0 | | | | | labs |
| **chlorideurine_max** | Chloride Urine (Max) | continuous | Maximum chloride urine measurement for this patient's hospitalization | mEq/L | Labs | 0 | | | | | labs |
| **chlorideurine_mean** | Chloride Urine (Mean) | continuous | Average chloride urine measurement for this patient's hospitalization | mEq/L | Labs | 0 | | | | | labs |
| **chlorideurine_median** | Chloride Urine (Median) | continuous | Median chloride urine measurement for this patient's hospitalization | mEq/L | Labs | 0 | | | | | labs |
| **chlorideurine_min** | Chloride Urine (Min) | continuous | Minimum chloride urine measurement for this patient's hospitalization | mEq/L | Labs | 0 | | | | | labs |
| **chlorideurine_sd** | Chloride Urine (SD) | continuous | Standard deviation of the chloride urine measurements during this patient's hospitalization | mEq/L | Labs | 0 | | | | | labs |
| **cholesterol_25** | Cholesterol (25th Percentile) | continuous | Cholesterol at the 25th percentile of this patient's measurements | mg/dL | Labs | 0 | | | | | labs |
| **cholesterol_75** | Cholesterol (75th Percentile) | continuous | Cholesterol at the 75th percentile of this patient's measurements | mg/dL | Labs | 0 | | | | | labs |
| **cholesterol_count** | Cholesterol (Count) | continuous | Count of cholesterol measurements for this patient | Number of measurements | Labs | 0 | | | | | labs |
| **cholesterol_first** | Cholesterol (First) | continuous | First cholesterol measurement for this patient's hospitalization | mg/dL | Labs | 0 | | | | | labs |
| **cholesterol_last** | Cholesterol (Last) | continuous | Last cholesterol measurement for this patient's hospitalization | mg/dL | Labs | 0 | | | | | labs |
| **cholesterol_max** | Cholesterol (Max) | continuous | Maximum cholesterol measurement for this patient's hospitalization | mg/dL | Labs | 0 | | | | | labs |
| **cholesterol_mean** | Cholesterol (Mean) | continuous | Average cholesterol measurement for this patient's hospitalization | mg/dL | Labs | 0 | | | | | labs |
| **cholesterol_median** | Cholesterol (Median) | continuous | Median cholesterol measurement for this patient's hospitalization | mg/dL | Labs | 0 | | | | | labs |
| **cholesterol_min** | Cholesterol (Min) | continuous | Minimum cholesterol measurement for this patient's hospitalization | mg/dL | Labs | 0 | | | | | labs |
| **cholesterol_sd** | Cholesterol (SD) | continuous | Standard deviation of the cholesterol measurements during this patient's hospitalization | mg/dL | Labs | 0 | | | | | labs |
| **cholesterolhdl_25** | Cholesterol HDL (25th Percentile) | continuous | Cholesterol HDL at the 25th percentile of this patient's measurements | mg/dL | Labs | 0 | | | | | labs |
| **cholesterolhdl_75** | Cholesterol HDL (75th Percentile) | continuous | Cholesterol HDL at the 75th percentile of this patient's measurements | mg/dL | Labs | 0 | | | | | labs |
| **cholesterolhdl_count** | Cholesterol HDL (Count) | continuous | Count of cholesterol HDL measurements for this patient | Number of measurements | Labs | 0 | | | | | labs |
| **cholesterolhdl_first** | Cholesterol HDL (First) | continuous | First cholesterol HDL measurement for this patient's hospitalization | mg/dL | Labs | 0 | | | | | labs |
| **cholesterolhdl_last** | Cholesterol HDL (Last) | continuous | Last cholesterol HDL measurement for this patient's hospitalization | mg/dL | Labs | 0 | | | | | labs |
| **cholesterolhdl_max** | Cholesterol HDL (Max) | continuous | Maximum cholesterol HDL measurement for this patient's hospitalization | mg/dL | Labs | 0 | | | | | labs |
| **cholesterolhdl_mean** | Cholesterol HDL (Mean) | continuous | Average cholesterol HDL measurement for this patient's hospitalization | mg/dL | Labs | 0 | | | | | labs |
| **cholesterolhdl_median** | Cholesterol HDL (Median) | continuous | Median cholesterol HDL measurement for this patient's hospitalization | mg/dL | Labs | 0 | | | | | labs |
| **cholesterolhdl_min** | Cholesterol HDL (Min) | continuous | Minimum cholesterol HDL measurement for this patient's hospitalization | mg/dL | Labs | 0 | | | | | labs |
| **cholesterolhdl_sd** | Cholesterol HDL (SD) | continuous | Standard deviation of the cholesterol HDL measurements during this patient's hospitalization | mg/dL | Labs | 0 | | | | | labs |
| **cholesterolldl_25** | Cholesterol LDL (25th Percentile) | continuous | Cholesterol LDL at the 25th percentile of this patient's measurements | mg/dL | Labs | 0 | | | | | labs |
| **cholesterolldl_75** | Cholesterol LDL (75th Percentile) | continuous | Cholesterol LDL at the 75th percentile of this patient's measurements | mg/dL | Labs | 0 | | | | | labs |
| **cholesterolldl_count** | Cholesterol LDL (Count) | continuous | Count of cholesterol LDL measurements for this patient | Number of measurements | Labs | 0 | | | | | labs |
| **cholesterolldl_first** | Cholesterol LDL (First) | continuous | First cholesterol LDL measurement for this patient's hospitalization | mg/dL | Labs | 0 | | | | | labs |
| **cholesterolldl_last** | Cholesterol LDL (Last) | continuous | Last cholesterol LDL measurement for this patient's hospitalization | mg/dL | Labs | 0 | | | | | labs |
| **cholesterolldl_max** | Cholesterol LDL (Max) | continuous | Maximum cholesterol LDL measurement for this patient's hospitalization | mg/dL | Labs | 0 | | | | | labs |
| **cholesterolldl_mean** | Cholesterol LDL (Mean) | continuous | Average cholesterol LDL measurement for this patient's hospitalization | mg/dL | Labs | 0 | | | | | labs |
| **cholesterolldl_median** | Cholesterol LDL (Median) | continuous | Median cholesterol LDL measurement for this patient's hospitalization | mg/dL | Labs | 0 | | | | | labs |
| **cholesterolldl_min** | Cholesterol LDL (Min) | continuous | Minimum cholesterol LDL measurement for this patient's hospitalization | mg/dL | Labs | 0 | | | | | labs |
| **cholesterolldl_sd** | Cholesterol LDL (SD) | continuous | Standard deviation of the cholesterol LDL measurements during this patient's hospitalization | mg/dL | Labs | 0 | | | | | labs |
| **ckmb_25** | Creatine Kinase-MB (25th Percentile) | continuous | Creatine Kinase-MB at the 25th percentile of this patient's measurements | IU/L | Labs | 0 | | | | | labs |
| **ckmb_75** | Creatine Kinase-MB (75th Percentile) | continuous | Creatine Kinase-MB at the 75th percentile of this patient's measurements | IU/L | Labs | 0 | | | | | labs |
| **ckmb_count** | Creatine Kinase-MB (Count) | continuous | Count of creatine kinase-MB measurements for this patient | Number of measurements | Labs | 0 | | | | | labs |
| **ckmb_first** | Creatine Kinase-MB (First) | continuous | First creatine kinase-MB measurement for this patient's hospitalization | IU/L | Labs | 0 | | | | | labs |
| **ckmb_last** | Creatine Kinase-MB (Last) | continuous | Last creatine kinase-MB measurement for this patient's hospitalization | IU/L | Labs | 0 | | | | | labs |
| **ckmb_max** | Creatine Kinase-MB (Max) | continuous | Maximum creatine kinase-MB measurement for this patient's hospitalization | IU/L | Labs | 0 | | | | | labs |
| **ckmb_mean** | Creatine Kinase-MB (Mean) | continuous | Average creatine kinase-MB measurement for this patient's hospitalization | IU/L | Labs | 0 | | | | | labs |
| **ckmb_median** | Creatine Kinase-MB (Median) | continuous | Median creatine kinase-MB measurement for this patient's hospitalization | IU/L | Labs | 0 | | | | | labs |
| **ckmb_min** | Creatine Kinase-MB (Min) | continuous | Minimum creatine kinase-MB measurement for this patient's hospitalization | IU/L | Labs | 0 | | | | | labs |
| **ckmb_sd** | Creatine Kinase-MB (SD) | continuous | Standard deviation of the creatine kinase-MB measurements during this patient's hospitalization | IU/L | Labs | 0 | | | | | labs |
| **cktotal_25** | Creatine Kinase Total (25th Percentile) | continuous | Creatine Kinase Total at the 25th percentile of this patient's measurements | IU/L | Labs | 0 | | | | | labs |
| **cktotal_75** | Creatine Kinase Total (75th Percentile) | continuous | Creatine Kinase Total at the 75th percentile of this patient's measurements | IU/L | Labs | 0 | | | | | labs |
| **cktotal_count** | Creatine Kinase Total (Count) | continuous | Count of creatine kinase total measurements for this patient | Number of measurements | Labs | 0 | | | | | labs |
| **cktotal_first** | Creatine Kinase Total (First) | continuous | First creatine kinase total measurement for this patient's hospitalization | IU/L | Labs | 0 | | | | | labs |
| **cktotal_last** | Creatine Kinase Total (Last) | continuous | Last creatine kinase total measurement for this patient's hospitalization | IU/L | Labs | 0 | | | | | labs |
| **cktotal_max** | Creatine Kinase Total (Max) | continuous | Maximum creatine kinase total measurement for this patient's hospitalization | IU/L | Labs | 0 | | | | | labs |
| **cktotal_mean** | Creatine Kinase Total (Mean) | continuous | Average creatine kinase total measurement for this patient's hospitalization | IU/L | Labs | 0 | | | | | labs |
| **cktotal_median** | Creatine Kinase Total (Median) | continuous | Median creatine kinase total measurement for this patient's hospitalization | IU/L | Labs | 0 | | | | | labs |
| **cktotal_min** | Creatine Kinase Total (Min) | continuous | Minimum creatine kinase total measurement for this patient's hospitalization | IU/L | Labs | 0 | | | | | labs |
| **cktotal_sd** | Creatine Kinase Total (SD) | continuous | Standard deviation of the creatine kinase total measurements during this patient's hospitalization | IU/L | Labs | 0 | | | | | labs |
| **cmvlog_25** | Cytomegalovirus (25th Percentile) | continuous | Cytomegalovirus at the 25th percentile of this patient's measurements | AI | Labs | 0 | | | | | labs |
| **cmvlog_75** | Cytomegalovirus (75th Percentile) | continuous | Cytomegalovirus at the 75th percentile of this patient's measurements | AI | Labs | 0 | | | | | labs |
| **cmvlog_count** | Cytomegalovirus (Count) | continuous | Count of cytomegalovirus measurements for this patient | Number of measurements | Labs | 0 | | | | | labs |
| **cmvlog_first** | Cytomegalovirus (First) | continuous | First cytomegalovirus measurement for this patient's hospitalization | AI | Labs | 0 | | | | | labs |
| **cmvlog_last** | Cytomegalovirus (Last) | continuous | Last cytomegalovirus measurement for this patient's hospitalization | AI | Labs | 0 | | | | | labs |
| **cmvlog_max** | Cytomegalovirus (Max) | continuous | Maximum cytomegalovirus measurement for this patient's hospitalization | AI | Labs | 0 | | | | | labs |
| **cmvlog_mean** | Cytomegalovirus (Mean) | continuous | Average cytomegalovirus measurement for this patient's hospitalization | AI | Labs | 0 | | | | | labs |
| **cmvlog_median** | Cytomegalovirus (Median) | continuous | Median cytomegalovirus measurement for this patient's hospitalization | AI | Labs | 0 | | | | | labs |
| **cmvlog_min** | Cytomegalovirus (Min) | continuous | Minimum cytomegalovirus measurement for this patient's hospitalization | AI | Labs | 0 | | | | | labs |
| **cmvlog_sd** | Cytomegalovirus (SD) | continuous | Standard deviation of the cytomegalovirus measurements during this patient's hospitalization | AI | Labs | 0 | | | | | labs |
| **cmvpcr_first** | Cytomegalovirus PCR (First) | categorical | Results of the first Cytomegalovirus PCR during this patient's hospitalization | AI | Labs | 0 | | | | | labs |
| **cmvpcr_last** | Cytomegalovirus PCR (Last) | categorical | Results of the last Cytomegalovirus PCR during this patient's hospitalization | AI | Labs | 0 | | | | | labs |
| **cocaineurine_first** | Cocaine Urine (First) | categorical | Results of the first Cocaine Urine Test during this patient's hospitalization | ng/mL | Labs | 0 | | | | | labs |
| **cocaineurine_last** | Cocaine Urine (Last) | categorical | Results of the last Cocaine Urine Test during this patient's hospitalization | ng/mL | Labs | 0 | | | | | labs |
| **community_aki** | Community Acquired Kidney Injury | categorical | Community Acquired Kidney Injury prior to admission | | Labs | 0 | | | | | aki_flag |
| **community_aki_stage** | Community Acquired Kidney Injury Stage | categorical | Community Acquired Kidney Injury Stage diagnosis for this patient | | Labs | 0 | | | | | |
| **cortisol_25** | Cortisol (25th Percentile) | continuous | Cortisol at the 25th percentile of this patient's measurements | µg/dL | Labs | 0 | | | | | labs |
| **cortisol_75** | Cortisol (75th Percentile) | continuous | Cortisol at the 75th percentile of this patient's measurements | µg/dL | Labs | 0 | | | | | labs |
| **cortisol_count** | Cortisol (Count) | continuous | Count of cortisol measurements for this patient | Number of measurements | Labs | 0 | | | | | labs |
| **cortisol_first** | Cortisol (First) | continuous | First cortisol measurement for this patient's hospitalization | µg/dL | Labs | 0 | | | | | labs |
| **cortisol_last** | Cortisol (Last) | continuous | Last cortisol measurement for this patient's hospitalization | µg/dL | Labs | 0 | | | | | labs |
| **cortisol_max** | Cortisol (Max) | continuous | Maximum cortisol measurement for this patient's hospitalization | µg/dL | Labs | 0 | | | | | labs |
| **cortisol_mean** | Cortisol (Mean) | continuous | Average cortisol measurement for this patient's hospitalization | µg/dL | Labs | 0 | | | | | labs |
| **cortisol_median** | Cortisol (Median) | continuous | Median cortisol measurement for this patient's hospitalization | µg/dL | Labs | 0 | | | | | labs |
| **cortisol_min** | Cortisol (Min) | continuous | Minimum cortisol measurement for this patient's hospitalization | µg/dL | Labs | 0 | | | | | labs |
| **cortisol_sd** | Cortisol (SD) | continuous | Standard deviation of the cortisol measurements during this patient's hospitalization | µg/dL | Labs | 0 | | | | | labs |
| **covid** | COVID-19 | categorical | Patient has been diagnosed with COVID during their hospitalization | | Labs | 0 | | | | | labs |
| **covid_25** | Covid (25th Percentile) | continuous | Covid at the 25th percentile of this patient's measurements | | Labs | 0 | | | | | labs |
| **covid_75** | Covid (75th Percentile) | continuous | Covid at the 75th percentile of this patient's measurements | | Labs | 0 | | | | | labs |
| **covid_count** | Covid (Count) | continuous | Count of covid measurements for this patient | Number of measurements | Labs | 0 | | | | | labs |

| | | | | | | | | | | | |
|---|---|---|---|---|---|---|---|---|---|---|---|
| **covid_first** | Covid (First) | continuous | First covid measurement for this patient's hospitalization | | Labs | 0 | | | | | labs |
| **covid_last** | Covid (Last) | continuous | Last covid measurement for this patient's hospitalization | | Labs | 0 | | | | | labs |
| **covid_max** | Covid (Max) | continuous | Maximum covid measurement for this patient's hospitalization | | Labs | 0 | | | | | labs |
| **covid_mean** | Covid (Mean) | continuous | Average covid measurement for this patient's hospitalization | | Labs | 0 | | | | | labs |
| **covid_median** | Covid (Median) | categorical | Median covid measurement for this patient's hospitalization | | Labs | 0 | | | | | labs |
| **covid_min** | Covid (Min) | continuous | Minimum covid measurement for this patient's hospitalization | | Labs | 0 | | | | | labs |
| **covid_sd** | Covid (SD) | continuous | Standard deviation of the covid measurements during this patient's hospitalization | | Labs | 0 | | | | | labs |
| **covidab_first** | Covid AB (First) | continuous | First covid AB measurement for this patient's hospitalization | | Labs | 0 | | | | | labs |
| **covidab_last** | Covid AB (Last) | continuous | Last covid AB measurement for this patient's hospitalization | | Labs | 0 | | | | | labs |
| **covidtest_25** | Covid Test (25th Percentile) | continuous | Covid Test at the 25th percentile of this patient's measurements | | Labs | 0 | | | | | |
| **covidtest_75** | Covid Test (75th Percentile) | continuous | Covid Test at the 75th percentile of this patient's measurements | | Labs | 0 | | | | | |
| **covidtest_count** | Covid Test (Count) | continuous | Count of covid test measurements for this patient | | Labs | 0 | | | | | |
| **covidtest_first** | Covid Test (First) | continuous | First covid test measurement for this patient's hospitalization | | Labs | 0 | | | | | labs |
| **covidtest_last** | Covid Test (Last) | continuous | Last covid test measurement for this patient's hospitalization | | Labs | 0 | | | | | labs |
| **covidtest_max** | Covid Test (Max) | continuous | Maximum covid test measurement for this patient's hospitalization | | Labs | 0 | | | | | |
| **covidtest_mean** | Covid Test (Mean) | continuous | Average covid test measurement for this patient's hospitalization | | Labs | 0 | | | | | |
| **covidtest_median** | Covid Test (Median) | categorical | Median covid test measurement for this patient's hospitalization | | Labs | 0 | | | | | |
| **covidtest_min** | Covid Test (Minimum) | continuous | Minimum covid test measurement for this patient's hospitalization | | Labs | 0 | | | | | |
| **covidtest_sd** | Covid Test (SD) | continuous | Standard deviation of the covid test measurements during this patient's hospitalization | | Labs | 0 | | | | | |
| **cpeptide_first** | C-Peptide (First) | categorical | First C-Peptide test measurement for this patient's hospitalization | | Labs | 0 | | | | | |
| **cpeptide_last** | C-Peptide (Last) | categorical | Last C-Peptide test measurement for this patient's hospitalization | | Labs | 0 | | | | | |
| **creatclearance_25** | Creatinine Clearance (25th Percentile) | continuous | Creatinine Clearance at the 25th percentile of this patient's measurements | mL/min | Labs | 0 | | | | | labs |
| **creatclearance_75** | Creatinine Clearance (75th Percentile) | continuous | Creatinine Clearance at the 75th percentile of this patient's measurements | mL/min | Labs | 0 | | | | | labs |
| **creatclearance_count** | Creatinine Clearance (Count) | continuous | Count of creatinine clearance measurements for this patient | Number of measurements | Labs | 0 | | | | | labs |
| **creatclearance_first** | Creatinine Clearance (First) | continuous | First creatinine clearance measurement for this patient's hospitalization | mL/min | Labs | 0 | | | | | labs |
| **creatclearance_last** | Creatinine Clearance (Last) | continuous | Last creatinine clearance measurement for this patient's hospitalization | mL/min | Labs | 0 | | | | | labs |
| **creatclearance_max** | Creatinine Clearance (Max) | continuous | Maximum creatinine clearance measurement for this patient's hospitalization | mL/min | Labs | 0 | | | | | labs |
| **creatclearance_mean** | Creatinine Clearance (Mean) | continuous | Average creatinine clearance measurement for this patient's hospitalization | mL/min | Labs | 0 | | | | | labs |
| **creatclearance_median** | Creatinine Clearance (Median) | continuous | Median creatinine clearance measurement for this patient's hospitalization | mL/min | Labs | 0 | | | | | labs |
| **creatclearance_min** | Creatinine Clearance (Min) | continuous | Minimum creatinine clearance measurement for this patient's hospitalization | mL/min | Labs | 0 | | | | | labs |
| **creatclearance_sd** | Creatinine Clearance (SD) | continuous | Standard deviation of the creatinine clearance measurements during this patient's hospitalization | mL/min | Labs | 0 | | | | | labs |
| **creatinine24hour_25** | Creatinine 24 Hour (25th Percentile) | continuous | Creatinine 24 Hour at the 25th percentile of this patient's measurements | mg/24 hr. | Labs | 0 | | | | | labs |
| **creatinine24hour_75** | Creatinine 24 Hour (75th Percentile) | continuous | Creatinine 24 Hour at the 75th percentile of this patient's measurements | mg/24 hr. | Labs | 0 | | | | | labs |
| **creatinine24hour_count** | Creatinine 24 Hour (Count) | continuous | Count of creatinine 24 hour measurements for this patient | Number of measurements | Labs | 0 | | | | | labs |
| **creatinine24hour_first** | Creatinine 24 Hour (First) | continuous | First creatinine 24 hour measurement for this patient's hospitalization | mg/24 hr. | Labs | 0 | | | | | labs |
| **creatinine24hour_last** | Creatinine 24 Hour (Last) | continuous | Last creatinine 24 hour measurement for this patient's hospitalization | mg/24 hr. | Labs | 0 | | | | | labs |
| **creatinine24hour_max** | Creatinine 24 Hour (Max) | continuous | Maximum creatinine 24 hour measurement for this patient's hospitalization | mg/24 hr. | Labs | 0 | | | | | labs |
| **creatinine24hour_mean** | Creatinine 24 Hour (Mean) | continuous | Average creatinine 24 hour measurement for this patient's hospitalization | mg/24 hr. | Labs | 0 | | | | | labs |
| **creatinine24hour_media** | Creatinine 24 Hour (Median) | continuous | Median creatinine 24 hour measurement for this patient's hospitalization | mg/24 hr. | Labs | 0 | | | | | labs |
| **creatinine24hour_min** | Creatinine 24 Hour (Min) | continuous | Minimum creatinine 24 hour measurement for this patient's hospitalization | mg/24 hr. | Labs | 0 | | | | | labs |
| **creatinine24hour_sd** | Creatinine 24 Hour (SD) | continuous | Standard deviation of the creatinine 24 hour measurements during this patient's hospitalization | mg/24 hr. | Labs | 0 | | | | | labs |
| **creatinine24hoururine_2** | Creatinine 24 Hour Urine (25th Percentile) | continuous | Creatinine 24 Hour Urine at the 25th percentile of this patient's measurements | mg/24 hr. | Labs | 0 | | | | | labs |
| **creatinine24hoururine_7** | Creatinine 24 Hour Urine (75th Percentile) | continuous | Creatinine 24 Hour Urine at the 75th percentile of this patient's measurements | mg/24 hr. | Labs | 0 | | | | | labs |
| **creatinine24hoururine_c** | Creatinine 24 Hour Urine (Count) | continuous | Count of creatinine 24 hour urine measurements for this patient | Number of measurements | Labs | 0 | | | | | labs |
| **creatinine24hoururine_f** | Creatinine 24 Hour Urine (First) | continuous | First creatinine 24 hour urine measurement for this patient's hospitalization | mg/24 hr. | Labs | 0 | | | | | labs |
| **creatinine24hoururine_l** | Creatinine 24 Hour Urine (Last) | continuous | Last creatinine 24 hour urine measurement for this patient's hospitalization | mg/24 hr. | Labs | 0 | | | | | labs |
| **creatinine24hoururine_m** | Creatinine 24 Hour Urine (Max) | continuous | Maximum creatinine 24 hour urine measurement for this patient's hospitalization | mg/24 hr. | Labs | 0 | | | | | labs |
| **creatinine24hoururine_m** | Creatinine 24 Hour Urine (Mean) | continuous | Average creatinine 24 hour urine measurement for this patient's hospitalization | mg/24 hr. | Labs | 0 | | | | | labs |
| **creatinine24hoururine_m** | Creatinine 24 Hour Urine (Median) | continuous | Median creatinine 24 hour urine measurement for this patient's hospitalization | mg/24 hr. | Labs | 0 | | | | | labs |
| **creatinine24hoururine_m** | Creatinine 24 Hour Urine (Min) | continuous | Minimum creatinine 24 hour urine measurement for this patient's hospitalization | mg/24 hr. | Labs | 0 | | | | | labs |
| **creatinine24hoururine_s** | Creatinine 24 Hour Urine (SD) | continuous | Standard deviation of the creatinine 24 hour urine measurements during this patient's hospitalization | mg/24 hr. | Labs | 0 | | | | | labs |
| **creatinine_25** | Creatinine (25th Percentile) | continuous | Creatinine at the 25th percentile of this patient's measurements | ?mol/L | Labs | 0 | | | | | labs |
| **creatinine_75** | Creatinine (75th Percentile) | continuous | Creatinine at the 75th percentile of this patient's measurements | ?mol/L | Labs | 0 | | | | | labs |
| **creatinine_count** | Creatinine (Count) | continuous | Count of creatinine measurements for this patient | Number of measurements | Labs | 0 | | | | | labs |
| **creatinine_first** | Creatinine (First) | continuous | First creatinine measurement for this patient's hospitalization | ?mol/L | Labs | 0 | | | | | labs |
| **creatinine_last** | Creatinine (Last) | continuous | Last creatinine measurement for this patient's hospitalization | ?mol/L | Labs | 0 | | | | | labs |
| **creatinine_max** | Creatinine (Max) | continuous | Maximum creatinine measurement for this patient's hospitalization | ?mol/L | Labs | 0 | | | | | labs |
| **creatinine_mean** | Creatinine (Mean) | continuous | Average creatinine measurement for this patient's hospitalization | ?mol/L | Labs | 0 | | | | | labs |
| **creatinine_median** | Creatinine (Median) | continuous | Median creatinine measurement for this patient's hospitalization | ?mol/L | Labs | 0 | | | | | labs |
| **creatinine_min** | Creatinine (Min) | continuous | Minimum creatinine measurement for this patient's hospitalization | ?mol/L | Labs | 0 | | | | | labs |
| **creatinine_sd** | Creatinine (SD) | continuous | Standard deviation of the creatinine measurements during this patient's hospitalization | ?mol/L | Labs | 0 | | | | | labs |
| **creatininefluid_25** | Creatinine Fluid (25th Percentile) | continuous | Creatinine Fluid at the 25th percentile of this patient's measurements | mg/dL | Labs | 0 | | | | | labs |
| **creatininefluid_75** | Creatinine Fluid (75th Percentile) | continuous | Creatinine Fluid at the 75th percentile of this patient's measurements | mg/dL | Labs | 0 | | | | | labs |
| **creatininefluid_count** | Creatinine Fluid (Count) | continuous | Count of creatinine fluid measurements for this patient | Number of measurements | Labs | 0 | | | | | labs |
| **creatininefluid_first** | Creatinine Fluid (First) | continuous | First creatinine fluid measurement for this patient's hospitalization | mg/dL | Labs | 0 | | | | | labs |
| **creatininefluid_last** | Creatinine Fluid (Last) | continuous | Last creatinine fluid measurement for this patient's hospitalization | mg/dL | Labs | 0 | | | | | labs |
| **creatininefluid_max** | Creatinine Fluid (Max) | continuous | Maximum creatinine fluid measurement for this patient's hospitalization | mg/dL | Labs | 0 | | | | | labs |
| **creatininefluid_mean** | Creatinine Fluid (Mean) | continuous | Average creatinine fluid measurement for this patient's hospitalization | mg/dL | Labs | 0 | | | | | labs |
| **creatininefluid_median** | Creatinine Fluid (Median) | continuous | Median creatinine fluid measurement for this patient's hospitalization | mg/dL | Labs | 0 | | | | | labs |
| **creatininefluid_min** | Creatinine Fluid (Min) | continuous | Minimum creatinine fluid measurement for this patient's hospitalization | mg/dL | Labs | 0 | | | | | labs |
| **creatininefluid_sd** | Creatinine Fluid (SD) | continuous | Standard deviation of the creatinine fluid measurements during this patient's hospitalization | mg/dL | Labs | 0 | | | | | labs |
| **creatininepoc_25** | Creatinine point-of-care (25th Percentile) | continuous | Creatinine point-of-care at the 25th percentile of this patient's measurements | mg/dL | Labs | 0 | | | | | labs |
| **creatininepoc_75** | Creatinine point-of-care (75th Percentile) | continuous | Creatinine point-of-care at the 75th percentile of this patient's measurements | mg/dL | Labs | 0 | | | | | labs |
| **creatininepoc_count** | Creatinine point-of-care (Count) | continuous | Count of creatinine point-of-care measurements for this patient | Number of measurements | Labs | 0 | | | | | labs |
| **creatininepoc_first** | Creatinine point-of-care (First) | continuous | First creatinine point-of-care measurement for this patient's hospitalization | mg/dL | Labs | 0 | | | | | labs |
| **creatininepoc_last** | Creatinine point-of-care (Last) | continuous | Last creatinine point-of-care measurement for this patient's hospitalization | mg/dL | Labs | 0 | | | | | labs |
| **creatininepoc_max** | Creatinine point-of-care (Max) | continuous | Maximum creatinine point-of-care measurement for this patient's hospitalization | mg/dL | Labs | 0 | | | | | labs |
| **creatininepoc_mean** | Creatinine point-of-care (Mean) | continuous | Average creatinine point-of-care measurement for this patient's hospitalization | mg/dL | Labs | 0 | | | | | labs |
| **creatininepoc_median** | Creatinine point-of-care (Median) | continuous | Median creatinine point-of-care measurement for this patient's hospitalization | mg/dL | Labs | 0 | | | | | labs |
| **creatininepoc_min** | Creatinine point-of-care (Min) | continuous | Minimum creatinine point-of-care measurement for this patient's hospitalization | mg/dL | Labs | 0 | | | | | labs |
| **creatininepoc_sd** | Creatinine point-of-care (SD) | continuous | Standard deviation of the creatinine point-of-care measurements during this patient's hospitalization | mg/dL | Labs | 0 | | | | | labs |
| **creatinineurine_25** | Creatinine Urine (25th Percentile) | continuous | Creatinine Urine at the 25th percentile of this patient's measurements | mg/dL | Labs | 0 | | | | | labs |
| **creatinineurine_75** | Creatinine Urine (75th Percentile) | continuous | Creatinine Urine at the 75th percentile of this patient's measurements | mg/dL | Labs | 0 | | | | | labs |
| **creatinineurine_count** | Creatinine Urine (Count) | continuous | Count of creatinine urine measurements for this patient | Number of measurements | Labs | 0 | | | | | labs |
| **creatinineurine_first** | Creatinine Urine (First) | continuous | First creatinine urine measurement for this patient's hospitalization | mg/dL | Labs | 0 | | | | | labs |
| **creatinineurine_last** | Creatinine Urine (Last) | continuous | Last creatinine urine measurement for this patient's hospitalization | mg/dL | Labs | 0 | | | | | labs |
| **creatinineurine_max** | Creatinine Urine (Max) | continuous | Maximum creatinine urine measurement for this patient's hospitalization | mg/dL | Labs | 0 | | | | | labs |
| **creatinineurine_mean** | Creatinine Urine (Mean) | continuous | Average creatinine urine measurement for this patient's hospitalization | mg/dL | Labs | 0 | | | | | labs |
| **creatinineurine_median** | Creatinine Urine (Median) | continuous | Median creatinine urine measurement for this patient's hospitalization | mg/dL | Labs | 0 | | | | | labs |
| **creatinineurine_min** | Creatinine Urine (Min) | continuous | Minimum creatinine urine measurement for this patient's hospitalization | mg/dL | Labs | 0 | | | | | labs |
| **creatinineurine_sd** | Creatinine Urine (SD) | continuous | Standard deviation of the creatinine urine measurements during this patient's hospitalization | mg/dL | Labs | 0 | | | | | labs |
| **crp_25** | C-Reactive Protein (25th Percentile) | continuous | C-Reactive Protein at the 25th percentile of this patient's measurements | mg/L | Labs | 0 | | | | | labs |
| **crp_75** | C-Reactive Protein (75th Percentile) | continuous | C-Reactive Protein at the 75th percentile of this patient's measurements | mg/L | Labs | 0 | | | | | labs |
| **crp_count** | C-Reactive Protein (Count) | continuous | Count of C-Reactive Protein measurements for this patient | Number of measurements | Labs | 0 | | | | | labs |
| **crp_first** | C-Reactive Protein (First) | continuous | First C-Reactive Protein measurement for this patient's hospitalization | mg/L | Labs | 0 | | | | | labs |
| **crp_last** | C-Reactive Protein (Last) | continuous | Last C-Reactive Protein measurement for this patient's hospitalization | mg/L | Labs | 0 | | | | | labs |
| **crp_max** | C-Reactive Protein (Max) | continuous | Maximum C-Reactive Protein measurement for this patient's hospitalization | mg/L | Labs | 0 | | | | | labs |
| **crp_mean** | C-Reactive Protein (Mean) | continuous | Average C-Reactive Protein measurement for this patient's hospitalization | mg/L | Labs | 0 | | | | | labs |
| **crp_median** | C-Reactive Protein (Median) | continuous | Median C-Reactive Protein measurement for this patient's hospitalization | mg/L | Labs | 0 | | | | | labs |
| **crp_min** | C-Reactive Protein (Min) | continuous | Minimum C-Reactive Protein measurement for this patient's hospitalization | mg/L | Labs | 0 | | | | | labs |
| **crp_sd** | C-Reactive Protein (SD) | continuous | Standard deviation of the C-Reactive Protein measurements during this patient's hospitalization | mg/L | Labs | 0 | | | | | labs |
| **cyclosporin_25** | Cyclosporin (25th Percentile) | continuous | Cyclosporin at the 25th percentile of this patient's measurements | ng/mL | Labs | 0 | | | | | labs |
| **cyclosporin_75** | Cyclosporin (75th Percentile) | continuous | Cyclosporin at the 75th percentile of this patient's measurements | ng/mL | Labs | 0 | | | | | labs |
| **cyclosporin_count** | Cyclosporin (Count) | continuous | Count of cyclosporin measurements for this patient | Number of measurements | Labs | 0 | | | | | labs |
| **cyclosporin_first** | Cyclosporin (First) | continuous | First cyclosporin measurement for this patient's hospitalization | ng/mL | Labs | 0 | | | | | labs |
| **cyclosporin_last** | Cyclosporin (Last) | continuous | Last cyclosporin measurement for this patient's hospitalization | ng/mL | Labs | 0 | | | | | labs |
| **cyclosporin_max** | Cyclosporin (Max) | continuous | Maximum cyclosporin measurement for this patient's hospitalization | ng/mL | Labs | 0 | | | | | labs |
| **cyclosporin_mean** | Cyclosporin (Mean) | continuous | Average cyclosporin measurement for this patient's hospitalization | ng/mL | Labs | 0 | | | | | labs |
| **cyclosporin_median** | Cyclosporin (Median) | continuous | Median cyclosporin measurement for this patient's hospitalization | ng/mL | Labs | 0 | | | | | labs |
| **cyclosporin_min** | Cyclosporin (Min) | continuous | Minimum cyclosporin measurement for this patient's hospitalization | ng/mL | Labs | 0 | | | | | labs |
| **cyclosporin_sd** | Cyclosporin (SD) | continuous | Standard deviation of the cyclosporin measurements during this patient's hospitalization | ng/mL | Labs | 0 | | | | | labs |
| **ddimer_25** | D-dimer (25th Percentile) | continuous | D-dimer at the 25th percentile of this patient's measurements | ng/mL | Labs | 0 | | | | | labs |
| **ddimer_75** | D-dimer (75th Percentile) | continuous | D-dimer at the 75th percentile of this patient's measurements | ng/mL | Labs | 0 | | | | | labs |
| **ddimer_count** | D-dimer (Count) | continuous | Count of D-dimer measurements for this patient | Number of measurements | Labs | 0 | | | | | labs |
| **ddimer_first** | D-dimer (First) | continuous | First D-dimer measurement for this patient's hospitalization | ng/mL | Labs | 0 | | | | | labs |
| **ddimer_last** | D-dimer (Last) | continuous | Last D-dimer measurement for this patient's hospitalization | ng/mL | Labs | 0 | | | | | labs |
| **ddimer_max** | D-dimer (Max) | continuous | Maximum D-dimer measurement for this patient's hospitalization | ng/mL | Labs | 0 | | | | | labs |
| **ddimer_mean** | D-dimer (Mean) | continuous | Average D-dimer measurement for this patient's hospitalization | ng/mL | Labs | 0 | | | | | labs |
| **ddimer_median** | D-dimer (Median) | continuous | Median D-dimer measurement for this patient's hospitalization | ng/mL | Labs | 0 | | | | | labs |
| **ddimer_min** | D-dimer (Min) | continuous | Minimum D-dimer measurement for this patient's hospitalization | ng/mL | Labs | 0 | | | | | labs |
| **ddimer_sd** | D-dimer (SD) | continuous | Standard deviation of the D-dimer measurements during this patient's hospitalization | ng/mL | Labs | 0 | | | | | labs |
| **diglevel_25** | Digoxin Level (25th Percentile) | continuous | Digoxin Level at the 25th percentile of this patient's measurements | ng/mL | Labs | 0 | | | | | labs |
| **diglevel_75** | Digoxin Level (75th Percentile) | continuous | Digoxin Level at the 75th percentile of this patient's measurements | ng/mL | Labs | 0 | | | | | labs |
| **diglevel_count** | Digoxin Level (Count) | continuous | Count of digoxin level measurements for this patient | Number of measurements | Labs | 0 | | | | | labs |

| | | | | | | | | | | | |
|---|---|---|---|---|---|---|---|---|---|---|---|
| **diglevel_first** | Digoxin Level (First) | continuous | First digoxin level measurement for this patient's hospitalization | ng/mL | Labs | 0 | | | | | labs |
| **diglevel_last** | Digoxin Level (Last) | continuous | Last digoxin level measurement for this patient's hospitalization | ng/mL | Labs | 0 | | | | | labs |
| **diglevel_max** | Digoxin Level (Max) | continuous | Maximum digoxin level measurement for this patient's hospitalization | ng/mL | Labs | 0 | | | | | labs |
| **diglevel_mean** | Digoxin Level (Mean) | continuous | Average digoxin level measurement for this patient's hospitalization | ng/mL | Labs | 0 | | | | | labs |
| **diglevel_median** | Digoxin Level (Median) | continuous | Median digoxin level measurement for this patient's hospitalization | ng/mL | Labs | 0 | | | | | labs |
| **diglevel_min** | Digoxin Level (Min) | continuous | Minimum digoxin level measurement for this patient's hospitalization | ng/mL | Labs | 0 | | | | | labs |
| **diglevel_sd** | Digoxin Level (SD) | continuous | Standard deviation of the digoxin level measurements during this patient's hospitalization | ng/mL | Labs | 0 | | | | | labs |
| **dsdna_25** | Double-Stranded DNA (25th Percentile) | continuous | Double-Stranded DNA at the 25th percentile of this patient's measurements | IU/mL | Labs | 0 | | | | | labs |
| **dsdna_75** | Double-Stranded DNA (75th Percentile) | continuous | Double-Stranded DNA at the 75th percentile of this patient's measurements | IU/mL | Labs | 0 | | | | | labs |
| **dsdna_count** | Double-Stranded DNA (Count) | continuous | Count of Double-Stranded DNA measurements for this patient | Number of measurements | Labs | 0 | | | | | labs |
| **dsdna_first** | Double-Stranded DNA (First) | continuous | First Double-Stranded DNA measurement for this patient's hospitalization | IU/mL | Labs | 0 | | | | | labs |
| **dsdna_last** | Double-Stranded DNA (Last) | continuous | Last Double-Stranded DNA measurement for this patient's hospitalization | IU/mL | Labs | 0 | | | | | labs |
| **dsdna_max** | Double-Stranded DNA (Max) | continuous | Maximum Double-Stranded DNA measurement for this patient's hospitalization | IU/mL | Labs | 0 | | | | | labs |
| **dsdna_mean** | Double-Stranded DNA (Mean) | continuous | Average Double-Stranded DNA measurement for this patient's hospitalization | IU/mL | Labs | 0 | | | | | labs |
| **dsdna_median** | Double-Stranded DNA (Median) | continuous | Median Double-Stranded DNA measurement for this patient's hospitalization | IU/mL | Labs | 0 | | | | | labs |
| **dsdna_min** | Double-Stranded DNA (Min) | continuous | Minimum Double-Stranded DNA measurement for this patient's hospitalization | IU/mL | Labs | 0 | | | | | labs |
| **dsdna_sd** | Double-Stranded DNA (SD) | continuous | Standard deviation of the Double-Stranded DNA measurements during this patient's hospitalization | IU/mL | Labs | 0 | | | | | labs |
| **ecgheartrate_25** | ECG Heart Rate (25th Percentile) | continuous | ECG Heart Rate at the 25th percentile of this patient's measurements | bpm | Labs | 0 | | | | | labs |
| **ecgheartrate_75** | ECG Heart Rate (75th Percentile) | continuous | ECG Heart Rate at the 75th percentile of this patient's measurements | bpm | Labs | 0 | | | | | labs |
| **ecgheartrate_count** | ECG Heart Rate (Count) | continuous | Count of ECG Heart Rate measurements for this patient | Number of measurements | Labs | 0 | | | | | labs |
| **ecgheartrate_first** | ECG Heart Rate (First) | continuous | First ECG Heart Rate measurement for this patient's hospitalization | bpm | Labs | 0 | | | | | labs |
| **ecgheartrate_last** | ECG Heart Rate (Last) | continuous | Last ECG Heart Rate measurement for this patient's hospitalization | bpm | Labs | 0 | | | | | labs |
| **ecgheartrate_max** | ECG Heart Rate (Max) | continuous | Maximum ECG Heart Rate measurement for this patient's hospitalization | bpm | Labs | 0 | | | | | labs |
| **ecgheartrate_mean** | ECG Heart Rate (Mean) | continuous | Average ECG Heart Rate measurement for this patient's hospitalization | bpm | Labs | 0 | | | | | labs |
| **ecgheartrate_median** | ECG Heart Rate (Median) | continuous | Median ECG Heart Rate measurement for this patient's hospitalization | bpm | Labs | 0 | | | | | labs |
| **ecgheartrate_min** | ECG Heart Rate (Min) | continuous | Minimum ECG Heart Rate measurement for this patient's hospitalization | bpm | Labs | 0 | | | | | labs |
| **ecgheartrate_sd** | ECG Heart Rate (SD) | continuous | Standard deviation of the ECG Heart Rate measurements during this patient's hospitalization | bpm | Labs | 0 | | | | | labs |
| **ecghr_25** | ECG HR (25th Percentile) | continuous | ECG HR at the 25th percentile of this patient's measurements | bpm | Labs | 0 | | | | | labs |
| **ecghr_75** | ECG HR (75th Percentile) | continuous | ECG HR at the 75th percentile of this patient's measurements | bpm | Labs | 0 | | | | | labs |
| **ecghr_count** | ECG HR (Count) | continuous | Count of ECG HR measurements for this patient | Number of measurements | Labs | 0 | | | | | labs |
| **ecghr_first** | ECG HR (First) | continuous | First ECG HR measurement for this patient's hospitalization | bpm | Labs | 0 | | | | | labs |
| **ecghr_last** | ECG HR (Last) | continuous | Last ECG HR measurement for this patient's hospitalization | bpm | Labs | 0 | | | | | labs |
| **ecghr_max** | ECG HR (Max) | continuous | Maximum ECG HR measurement for this patient's hospitalization | bpm | Labs | 0 | | | | | labs |
| **ecghr_mean** | ECG HR (Mean) | continuous | Average ECG HR measurement for this patient's hospitalization | bpm | Labs | 0 | | | | | labs |
| **ecghr_median** | ECG HR (Median) | continuous | Median ECG HR measurement for this patient's hospitalization | bpm | Labs | 0 | | | | | labs |
| **ecghr_min** | ECG HR (Min) | continuous | Minimum ECG HR measurement for this patient's hospitalization | bpm | Labs | 0 | | | | | labs |
| **ecghr_sd** | ECG HR (SD) | continuous | Standard deviation of the ECG HR measurements during this patient's hospitalization | bpm | Labs | 0 | | | | | labs |
| **ecgp_25** | ECG P (25th Percentile) | continuous | ECG P at the 25th percentile of this patient's measurements | ms | Labs | 0 | | | | | labs |
| **ecgp_75** | ECG P (75th Percentile) | continuous | ECG P at the 75th percentile of this patient's measurements | ms | Labs | 0 | | | | | labs |
| **ecgp_count** | ECG P (Count) | continuous | Count of ECG P measurements for this patient | Number of measurements | Labs | 0 | | | | | labs |
| **ecgp_first** | ECG P (First) | continuous | First ECG P measurement for this patient's hospitalization | ms | Labs | 0 | | | | | labs |
| **ecgp_last** | ECG P (Last) | continuous | Last ECG P measurement for this patient's hospitalization | ms | Labs | 0 | | | | | labs |
| **ecgp_max** | ECG P (Max) | continuous | Maximum ECG P measurement for this patient's hospitalization | ms | Labs | 0 | | | | | labs |
| **ecgp_mean** | ECG P (Mean) | continuous | Average ECG P measurement for this patient's hospitalization | ms | Labs | 0 | | | | | labs |
| **ecgp_median** | ECG P (Median) | continuous | Median ECG P measurement for this patient's hospitalization | ms | Labs | 0 | | | | | labs |
| **ecgp_min** | ECG P (Min) | continuous | Minimum ECG P measurement for this patient's hospitalization | ms | Labs | 0 | | | | | labs |
| **ecgp_sd** | ECG P (SD) | continuous | Standard deviation of the ECG P measurements during this patient's hospitalization | ms | Labs | 0 | | | | | labs |
| **ecgpr_25** | ECG Period (25th Percentile) | continuous | ECG Period at the 25th percentile of this patient's measurements | ms | Labs | 0 | | | | | labs |
| **ecgpr_75** | ECG Period (75th Percentile) | continuous | ECG Period at the 75th percentile of this patient's measurements | ms | Labs | 0 | | | | | labs |
| **ecgpr_count** | ECG Period (Count) | continuous | Count of ECG Period measurements for this patient | Number of measurements | Labs | 0 | | | | | labs |
| **ecgpr_first** | ECG Period (First) | continuous | First ECG Period measurement for this patient's hospitalization | ms | Labs | 0 | | | | | labs |
| **ecgpr_last** | ECG Period (Last) | continuous | Last ECG Period measurement for this patient's hospitalization | ms | Labs | 0 | | | | | labs |
| **ecgpr_max** | ECG Period (Max) | continuous | Maximum ECG Period measurement for this patient's hospitalization | ms | Labs | 0 | | | | | labs |
| **ecgpr_mean** | ECG Period (Mean) | continuous | Average ECG Period measurement for this patient's hospitalization | ms | Labs | 0 | | | | | labs |
| **ecgpr_median** | ECG Period (Median) | continuous | Median ECG Period measurement for this patient's hospitalization | ms | Labs | 0 | | | | | labs |
| **ecgpr_min** | ECG Period (Min) | continuous | Minimum ECG Period measurement for this patient's hospitalization | ms | Labs | 0 | | | | | labs |
| **ecgpr_sd** | ECG Period (SD) | continuous | Standard deviation of the ECG Period measurements during this patient's hospitalization | ms | Labs | 0 | | | | | labs |
| **ecgpwave_25** | ECG P Wave (25th Percentile) | continuous | ECG P Wave at the 25th percentile of this patient's measurements | ms | Labs | 0 | | | | | labs |
| **ecgpwave_75** | ECG P Wave (75th Percentile) | continuous | ECG P Wave at the 75th percentile of this patient's measurements | ms | Labs | 0 | | | | | labs |
| **ecgpwave_count** | ECG P Wave (Count) | continuous | Count of ECG P Wave measurements for this patient | Number of measurements | Labs | 0 | | | | | labs |
| **ecgpwave_first** | ECG P Wave (First) | continuous | First ECG P Wave measurement for this patient's hospitalization | ms | Labs | 0 | | | | | labs |
| **ecgpwave_last** | ECG P Wave (Last) | continuous | Last ECG P Wave measurement for this patient's hospitalization | ms | Labs | 0 | | | | | labs |
| **ecgpwave_max** | ECG P Wave (Max) | continuous | Maximum ECG P Wave measurement for this patient's hospitalization | ms | Labs | 0 | | | | | labs |
| **ecgpwave_mean** | ECG P Wave (Mean) | continuous | Average ECG P Wave measurement for this patient's hospitalization | ms | Labs | 0 | | | | | labs |
| **ecgpwave_median** | ECG P Wave (Median) | continuous | Median ECG P Wave measurement for this patient's hospitalization | ms | Labs | 0 | | | | | labs |
| **ecgpwave_min** | ECG P Wave (Min) | continuous | Minimum ECG P Wave measurement for this patient's hospitalization | ms | Labs | 0 | | | | | labs |
| **ecgpwave_sd** | ECG P Wave (SD) | continuous | Standard deviation of the ECG P Wave measurements during this patient's hospitalization | ms | Labs | 0 | | | | | labs |
| **ecgqrs_25** | ECG QRS (25th Percentile) | continuous | ECG QRS at the 25th percentile of this patient's measurements | ms | Labs | 0 | | | | | labs |
| **ecgqrs_75** | ECG QRS (75th Percentile) | continuous | ECG QRS at the 75th percentile of this patient's measurements | ms | Labs | 0 | | | | | labs |
| **ecgqrs_count** | ECG QRS (Count) | continuous | Count of ECG QRS measurements for this patient | Number of measurements | Labs | 0 | | | | | labs |
| **ecgqrs_first** | ECG QRS (First) | continuous | First ECG QRS measurement for this patient's hospitalization | ms | Labs | 0 | | | | | labs |
| **ecgqrs_last** | ECG QRS (Last) | continuous | Last ECG QRS measurement for this patient's hospitalization | ms | Labs | 0 | | | | | labs |
| **ecgqrs_max** | ECG QRS (Max) | continuous | Maximum ECG QRS measurement for this patient's hospitalization | ms | Labs | 0 | | | | | labs |
| **ecgqrs_mean** | ECG QRS (Mean) | continuous | Average ECG QRS measurement for this patient's hospitalization | ms | Labs | 0 | | | | | labs |
| **ecgqrs_median** | ECG QRS (Median) | continuous | Median ECG QRS measurement for this patient's hospitalization | ms | Labs | 0 | | | | | labs |
| **ecgqrs_min** | ECG QRS (Min) | continuous | Minimum ECG QRS measurement for this patient's hospitalization | ms | Labs | 0 | | | | | labs |
| **ecgqrs_sd** | ECG QRS (SD) | continuous | Standard deviation of the ECG QRS measurements during this patient's hospitalization | ms | Labs | 0 | | | | | labs |
| **ecgqrsaxis_25** | ECG QRS Axis (25th Percentile) | continuous | ECG QRS Axis at the 25th percentile of this patient's measurements | ms | Labs | 0 | | | | | labs |
| **ecgqrsaxis_75** | ECG QRS Axis (75th Percentile) | continuous | ECG QRS Axis at the 75th percentile of this patient's measurements | ms | Labs | 0 | | | | | labs |
| **ecgqrsaxis_count** | ECG QRS Axis (Count) | continuous | Count of ECG QRS Axis measurements for this patient | Number of measurements | Labs | 0 | | | | | labs |
| **ecgqrsaxis_first** | ECG QRS Axis (First) | continuous | First ECG QRS Axis measurement for this patient's hospitalization | ms | Labs | 0 | | | | | labs |
| **ecgqrsaxis_last** | ECG QRS Axis (Last) | continuous | Last ECG QRS Axis measurement for this patient's hospitalization | ms | Labs | 0 | | | | | labs |
| **ecgqrsaxis_max** | ECG QRS Axis (Max) | continuous | Maximum ECG QRS Axis measurement for this patient's hospitalization | ms | Labs | 0 | | | | | labs |
| **ecgqrsaxis_mean** | ECG QRS Axis (Mean) | continuous | Average ECG QRS Axis measurement for this patient's hospitalization | ms | Labs | 0 | | | | | labs |
| **ecgqrsaxis_median** | ECG QRS Axis (Median) | continuous | Median ECG QRS Axis measurement for this patient's hospitalization | ms | Labs | 0 | | | | | labs |
| **ecgqrsaxis_min** | ECG QRS Axis (Min) | continuous | Minimum ECG QRS Axis measurement for this patient's hospitalization | ms | Labs | 0 | | | | | labs |
| **ecgqrsaxis_sd** | ECG QRS Axis (SD) | continuous | Standard deviation of the ECG QRS Axis measurements during this patient's hospitalization | ms | Labs | 0 | | | | | labs |
| **ecgqrsint_25** | ECG QRS Interval (25th Percentile) | continuous | ECG QRS Interval at the 25th percentile of this patient's measurements | ms | Labs | 0 | | | | | labs |
| **ecgqrsint_75** | ECG QRS Interval (75th Percentile) | continuous | ECG QRS Interval at the 75th percentile of this patient's measurements | ms | Labs | 0 | | | | | labs |
| **ecgqrsint_count** | ECG QRS Interval (Count) | continuous | Count of ECG QRS Interval measurements for this patient | Number of measurements | Labs | 0 | | | | | labs |
| **ecgqrsint_first** | ECG QRS Interval (First) | continuous | First ECG QRS Interval measurement for this patient's hospitalization | ms | Labs | 0 | | | | | labs |
| **ecgqrsint_last** | ECG QRS Interval (Last) | continuous | Last ECG QRS Interval measurement for this patient's hospitalization | ms | Labs | 0 | | | | | labs |
| **ecgqrsint_max** | ECG QRS Interval (Max) | continuous | Maximum ECG QRS Interval measurement for this patient's hospitalization | ms | Labs | 0 | | | | | labs |
| **ecgqrsint_mean** | ECG QRS Interval (Mean) | continuous | Average ECG QRS Interval measurement for this patient's hospitalization | ms | Labs | 0 | | | | | labs |
| **ecgqrsint_median** | ECG QRS Interval (Median) | continuous | Median ECG QRS Interval measurement for this patient's hospitalization | ms | Labs | 0 | | | | | labs |
| **ecgqrsint_min** | ECG QRS Interval (Min) | continuous | Minimum ECG QRS Interval measurement for this patient's hospitalization | ms | Labs | 0 | | | | | labs |
| **ecgqrsint_sd** | ECG QRS Interval (SD) | continuous | Standard deviation of the ECG QRS Interval measurements during this patient's hospitalization | ms | Labs | 0 | | | | | labs |
| **ecgqt_25** | ECG QT (25th Percentile) | continuous | ECG QT at the 25th percentile of this patient's measurements | ms | Labs | 0 | | | | | labs |
| **ecgqt_75** | ECG QT (75th Percentile) | continuous | ECG QT at the 75th percentile of this patient's measurements | ms | Labs | 0 | | | | | labs |
| **ecgqt_count** | ECG QT (Count) | continuous | Count of ECG QT measurements for this patient | Number of measurements | Labs | 0 | | | | | labs |
| **ecgqt_first** | ECG QT (First) | continuous | First ECG QT measurement for this patient's hospitalization | ms | Labs | 0 | | | | | labs |
| **ecgqt_last** | ECG QT (Last) | continuous | Last ECG QT measurement for this patient's hospitalization | ms | Labs | 0 | | | | | labs |
| **ecgqt_max** | ECG QT (Max) | continuous | Maximum ECG QT measurement for this patient's hospitalization | ms | Labs | 0 | | | | | labs |
| **ecgqt_mean** | ECG QT (Mean) | continuous | Average ECG QT measurement for this patient's hospitalization | ms | Labs | 0 | | | | | labs |
| **ecgqt_median** | ECG QT (Median) | continuous | Median ECG QT measurement for this patient's hospitalization | ms | Labs | 0 | | | | | labs |
| **ecgqt_min** | ECG QT (Min) | continuous | Minimum ECG QT measurement for this patient's hospitalization | ms | Labs | 0 | | | | | labs |
| **ecgqt_sd** | ECG QT (SD) | continuous | Standard deviation of the ECG QT measurements during this patient's hospitalization | ms | Labs | 0 | | | | | labs |
| **ecgraxis_25** | ECG Right Axis (25th Percentile) | continuous | ECG Right Axis at the 25th percentile of this patient's measurements | deg. | Labs | 0 | | | | | labs |
| **ecgraxis_75** | ECG Right Axis (75th Percentile) | continuous | ECG Right Axis at the 75th percentile of this patient's measurements | deg. | Labs | 0 | | | | | labs |
| **ecgraxis_count** | ECG Right Axis (Count) | continuous | Count of ECG Right Axis measurements for this patient | Number of measurements | Labs | 0 | | | | | labs |
| **ecgraxis_first** | ECG Right Axis (First) | continuous | First ECG Right Axis measurement for this patient's hospitalization | deg. | Labs | 0 | | | | | labs |
| **ecgraxis_last** | ECG Right Axis (Last) | continuous | Last ECG Right Axis measurement for this patient's hospitalization | deg. | Labs | 0 | | | | | labs |
| **ecgraxis_max** | ECG Right Axis (Max) | continuous | Maximum ECG Right Axis measurement for this patient's hospitalization | deg. | Labs | 0 | | | | | labs |
| **ecgraxis_mean** | ECG Right Axis (Mean) | continuous | Average ECG Right Axis measurement for this patient's hospitalization | deg. | Labs | 0 | | | | | labs |
| **ecgraxis_median** | ECG Right Axis (Median) | continuous | Median ECG Right Axis measurement for this patient's hospitalization | deg. | Labs | 0 | | | | | labs |
| **ecgraxis_min** | ECG Right Axis (Min) | continuous | Minimum ECG Right Axis measurement for this patient's hospitalization | deg. | Labs | 0 | | | | | labs |
| **ecgraxis_sd** | ECG R Axis (SD) | continuous | Standard deviation of the ECG R Axis measurements during this patient's hospitalization | deg. | Labs | 0 | | | | | labs |
| **ecgtaxis_25** | ECG T Axis (25th Percentile) | continuous | ECG T Axis at the 25th percentile of this patient's measurements | deg. | Labs | 0 | | | | | labs |
| **ecgtaxis_75** | ECG T Axis (75th Percentile) | continuous | ECG T Axis at the 75th percentile of this patient's measurements | deg. | Labs | 0 | | | | | labs |
| **ecgtaxis_count** | ECG T Axis (Count) | continuous | Count of ECG T Axis measurements for this patient | Number of measurements | Labs | 0 | | | | | labs |
| **ecgtaxis_first** | ECG T Axis (First) | continuous | First ECG T Axis measurement for this patient's hospitalization | deg. | Labs | 0 | | | | | labs |
| **ecgtaxis_last** | ECG T Axis (Last) | continuous | Last ECG T Axis measurement for this patient's hospitalization | deg. | Labs | 0 | | | | | labs |
| **ecgtaxis_max** | ECG T Axis (Max) | continuous | Maximum ECG T Axis measurement for this patient's hospitalization | deg. | Labs | 0 | | | | | labs |
| **ecgtaxis_mean** | ECG T Axis (Mean) | continuous | Average ECG T Axis measurement for this patient's hospitalization | deg. | Labs | 0 | | | | | labs |

| | | | | | | | | | | | |
|---|---|---|---|---|---|---|---|---|---|---|---|
| **ecgtaxis_median** | ECG T Axis (Median) | continuous | Median ECG T Axis measurement for this patient's hospitalization | deg. | Labs | 0 | | | | | labs |
| **ecgtaxis_min** | ECG T Axis (Min) | continuous | Minimum ECG T Axis measurement for this patient's hospitalization | deg. | Labs | 0 | | | | | labs |
| **ecgtaxis_sd** | ECG T Axis (SD) | continuous | Standard deviation of the ECG T Axis measurements during this patient's hospitalization | deg. | Labs | 0 | | | | | labs |
| **egfr_25** | Estimated Glomerular Filtration Rate (25th Percentile) | continuous | Estimated Glomerular Filtration Rate at the 25th percentile of this patient's measurements | mL/min/m2 | Labs | 0 | | | | | labs |
| **egfr_75** | Estimated Glomerular Filtration Rate (75th Percentile) | continuous | Estimated Glomerular Filtration Rate at the 75th percentile of this patient's measurements | mL/min/m2 | Labs | 0 | | | | | labs |
| **egfr_count** | Estimated Glomerular Filtration Rate (Count) | continuous | Count of Estimated Glomerular Filtration Rate measurements for this patient | Number of measurements | Labs | 0 | | | | | labs |
| **egfr_first** | Estimated Glomerular Filtration Rate (First) | continuous | First Estimated Glomerular Filtration Rate measurement for this patient's hospitalization | mL/min/m2 | Labs | 0 | | | | | labs |
| **egfr_last** | Estimated Glomerular Filtration Rate (Last) | continuous | Last Estimated Glomerular Filtration Rate measurement for this patient's hospitalization | mL/min/m2 | Labs | 0 | | | | | labs |
| **egfr_max** | Estimated Glomerular Filtration Rate (Max) | continuous | Maximum Estimated Glomerular Filtration Rate measurement for this patient's hospitalization | mL/min/m2 | Labs | 0 | | | | | labs |
| **egfr_mean** | Estimated Glomerular Filtration Rate (Mean) | continuous | Average Estimated Glomerular Filtration Rate measurement for this patient's hospitalization | mL/min/m2 | Labs | 0 | | | | | labs |
| **egfr_median** | Estimated Glomerular Filtration Rate (Median) | continuous | Median Estimated Glomerular Filtration Rate measurement for this patient's hospitalization | mL/min/m2 | Labs | 0 | | | | | labs |
| **egfr_min** | Estimated Glomerular Filtration Rate (Min) | continuous | Minimum Estimated Glomerular Filtration Rate measurement for this patient's hospitalization | mL/min/m2 | Labs | 0 | | | | | labs |
| **egfr_sd** | Estimated Glomerular Filtration Rate (SD) | continuous | Standard deviation of the Estimated Glomerular Filtration Rate measurements during this patient's hospit | mL/min/m2 | Labs | 0 | | | | | labs |
| **egfraa_first** | Estimated Glomerular Filtration Rate AA (First) | categorical | Results of the first Estimated Glomerular Filtration Rate AA test during this patient's hospitalization | mL/min/m2 | Labs | 0 | | | | | labs |
| **egfraa_last** | Estimated Glomerular Filtration Rate AA (Last) | categorical | Results of the last Estimated Glomerular Filtration Rate AA test during this patient's hospitalization | mL/min/m2 | Labs | 0 | | | | | labs |
| **ejectionfraction_25** | Ejection Fraction (25th Percentile) | continuous | Ejection Fraction at the 25th percentile of this patient's measurements | % | Labs | 0 | | | | | labs |
| **ejectionfraction_75** | Ejection Fraction (75th Percentile) | continuous | Ejection Fraction at the 75th percentile of this patient's measurements | % | Labs | 0 | | | | | labs |
| **ejectionfraction_count** | Ejection Fraction (Count) | continuous | Count of Ejection Fraction measurements for this patient | Number of measurements | Labs | 0 | | | | | labs |
| **ejectionfraction_first** | Ejection Fraction (First) | continuous | First Ejection Fraction measurement for this patient's hospitalization | % | Labs | 0 | | | | | labs |
| **ejectionfraction_last** | Ejection Fraction (Last) | continuous | Last Ejection Fraction measurement for this patient's hospitalization | % | Labs | 0 | | | | | labs |
| **ejectionfraction_max** | Ejection Fraction (Max) | continuous | Maximum Ejection Fraction measurement for this patient's hospitalization | % | Labs | 0 | | | | | labs |
| **ejectionfraction_mean** | Ejection Fraction (Mean) | continuous | Average Ejection Fraction measurement for this patient's hospitalization | % | Labs | 0 | | | | | labs |
| **ejectionfraction_median** | Ejection Fraction (Median) | continuous | Median Ejection Fraction measurement for this patient's hospitalization | % | Labs | 0 | | | | | labs |
| **ejectionfraction_min** | Ejection Fraction (Min) | continuous | Minimum Ejection Fraction measurement for this patient's hospitalization | % | Labs | 0 | | | | | labs |
| **ejectionfraction_sd** | Ejection Fraction (SD) | continuous | Standard deviation of the Ejection Fraction measurements during this patient's hospitalization | % | Labs | 0 | | | | | labs |
| **eos_abs_25** | Absolute Eosinophil (25th Percentile) | continuous | Absolute Eosinophil at the 25th percentile of this patient's measurements | % | Labs | 0 | | | | | labs |
| **eos_abs_75** | Absolute Eosinophil (75th Percentile) | continuous | Absolute Eosinophil at the 75th percentile of this patient's measurements | % | Labs | 0 | | | | | labs |
| **eos_abs_count** | Absolute Eosinophil (Count) | continuous | Count of Absolute Eosinophil measurements for this patient | Number of measurements | Labs | 0 | | | | | labs |
| **eos_abs_first** | Absolute Eosinophil (First) | continuous | First Absolute Eosinophil measurement for this patient's hospitalization | % | Labs | 0 | | | | | labs |
| **eos_abs_last** | Absolute Eosinophil (Last) | continuous | Last Absolute Eosinophil measurement for this patient's hospitalization | % | Labs | 0 | | | | | labs |
| **eos_abs_max** | Absolute Eosinophil (Max) | continuous | Maximum Absolute Eosinophil measurement for this patient's hospitalization | % | Labs | 0 | | | | | labs |
| **eos_abs_mean** | Absolute Eosinophil (Mean) | continuous | Average Absolute Eosinophil measurement for this patient's hospitalization | % | Labs | 0 | | | | | labs |
| **eos_abs_median** | Absolute Eosinophil (Median) | continuous | Median Absolute Eosinophil measurement for this patient's hospitalization | % | Labs | 0 | | | | | labs |
| **eos_abs_min** | Absolute Eosinophil (Min) | continuous | Minimum Absolute Eosinophil measurement for this patient's hospitalization | % | Labs | 0 | | | | | labs |
| **eos_abs_sd** | Absolute Eosinophil (SD) | continuous | Standard deviation of the Absolute Eosinophil measurements during this patient's hospitalization | % | Labs | 0 | | | | | labs |
| **eos_percent_25** | Eosinophil Percent (25th Percentile) | continuous | Eosinophil Percent at the 25th percentile of this patient's measurements | % | Labs | 0 | | | | | labs |
| **eos_percent_75** | Eosinophil Percent (75th Percentile) | continuous | Eosinophil Percent at the 75th percentile of this patient's measurements | % | Labs | 0 | | | | | labs |
| **eos_percent_count** | Eosinophil Percent (Count) | continuous | Count of Eosinophil Percent measurements for this patient | Number of measurements | Labs | 0 | | | | | labs |
| **eos_percent_first** | Eosinophil Percent (First) | continuous | First Eosinophil Percent measurement for this patient's hospitalization | % | Labs | 0 | | | | | labs |
| **eos_percent_last** | Eosinophil Percent (Last) | continuous | Last Eosinophil Percent measurement for this patient's hospitalization | % | Labs | 0 | | | | | labs |
| **eos_percent_max** | Eosinophil Percent (Max) | continuous | Maximum Eosinophil Percent measurement for this patient's hospitalization | % | Labs | 0 | | | | | labs |
| **eos_percent_mean** | Eosinophil Percent (Mean) | continuous | Average Eosinophil Percent measurement for this patient's hospitalization | % | Labs | 0 | | | | | labs |
| **eos_percent_median** | Eosinophil Percent (Median) | continuous | Median Eosinophil Percent measurement for this patient's hospitalization | % | Labs | 0 | | | | | labs |
| **eos_percent_min** | Eosinophil Percent (Min) | continuous | Minimum Eosinophil Percent measurement for this patient's hospitalization | % | Labs | 0 | | | | | labs |
| **eos_percent_sd** | Eosinophil Percent (SD) | continuous | Standard deviation of the Eosinophil Percent measurements during this patient's hospitalization | % | Labs | 0 | | | | | labs |
| **ethanol_25** | Ethanol (25th Percentile) | continuous | Ethanol at the 25th percentile of this patient's measurements | mg/dL | Labs | 0 | | | | | labs |
| **ethanol_75** | Ethanol (75th Percentile) | continuous | Ethanol at the 75th percentile of this patient's measurements | mg/dL | Labs | 0 | | | | | labs |
| **ethanol_count** | Ethanol (Count) | continuous | Count of Ethanol measurements for this patient | Number of measurements | Labs | 0 | | | | | labs |
| **ethanol_first** | Ethanol (First) | continuous | First Ethanol measurement for this patient's hospitalization | mg/dL | Labs | 0 | | | | | labs |
| **ethanol_last** | Ethanol (Last) | continuous | Last Ethanol measurement for this patient's hospitalization | mg/dL | Labs | 0 | | | | | labs |
| **ethanol_max** | Ethanol (Max) | continuous | Maximum Ethanol measurement for this patient's hospitalization | mg/dL | Labs | 0 | | | | | labs |
| **ethanol_mean** | Ethanol (Mean) | continuous | Average Ethanol measurement for this patient's hospitalization | mg/dL | Labs | 0 | | | | | labs |
| **ethanol_median** | Ethanol (Median) | continuous | Median Ethanol measurement for this patient's hospitalization | mg/dL | Labs | 0 | | | | | labs |
| **ethanol_min** | Ethanol (Min) | continuous | Minimum Ethanol measurement for this patient's hospitalization | mg/dL | Labs | 0 | | | | | labs |
| **ethanol_sd** | Ethanol (SD) | continuous | Standard deviation of the Ethanol measurements during this patient's hospitalization | mg/dL | Labs | 0 | | | | | labs |
| **ferritin_25** | Ferritin (25th Percentile) | continuous | Ferritin at the 25th percentile of this patient's measurements | ng/mL | Labs | 0 | | | | | labs |
| **ferritin_75** | Ferritin (75th Percentile) | continuous | Ferritin at the 75th percentile of this patient's measurements | ng/mL | Labs | 0 | | | | | labs |
| **ferritin_count** | Ferritin (Count) | continuous | Count of Ferritin measurements for this patient | Number of measurements | Labs | 0 | | | | | labs |
| **ferritin_first** | Ferritin (First) | continuous | First Ferritin measurement for this patient's hospitalization | ng/mL | Labs | 0 | | | | | labs |
| **ferritin_last** | Ferritin (Last) | continuous | Last Ferritin measurement for this patient's hospitalization | ng/mL | Labs | 0 | | | | | labs |
| **ferritin_max** | Ferritin (Max) | continuous | Maximum Ferritin measurement for this patient's hospitalization | ng/mL | Labs | 0 | | | | | labs |
| **ferritin_mean** | Ferritin (Mean) | continuous | Average Ferritin measurement for this patient's hospitalization | ng/mL | Labs | 0 | | | | | labs |
| **ferritin_median** | Ferritin (Median) | continuous | Median Ferritin measurement for this patient's hospitalization | ng/mL | Labs | 0 | | | | | labs |
| **ferritin_min** | Ferritin (Min) | continuous | Minimum Ferritin measurement for this patient's hospitalization | ng/mL | Labs | 0 | | | | | labs |
| **ferritin_sd** | Ferritin (SD) | continuous | Standard deviation of the Ferritin measurements during this patient's hospitalization | ng/mL | Labs | 0 | | | | | labs |
| **fibrinogen_25** | Fibrinogen (25th Percentile) | continuous | Fibrinogen at the 25th percentile of this patient's measurements | mg/dL | Labs | 0 | | | | | labs |
| **fibrinogen_75** | Fibrinogen (75th Percentile) | continuous | Fibrinogen at the 75th percentile of this patient's measurements | mg/dL | Labs | 0 | | | | | labs |
| **fibrinogen_count** | Fibrinogen (Count) | continuous | Count of Fibrinogen measurements for this patient | Number of measurements | Labs | 0 | | | | | labs |
| **fibrinogen_first** | Fibrinogen (First) | continuous | First Fibrinogen measurement for this patient's hospitalization | mg/dL | Labs | 0 | | | | | labs |
| **fibrinogen_last** | Fibrinogen (Last) | continuous | Last Fibrinogen measurement for this patient's hospitalization | mg/dL | Labs | 0 | | | | | labs |
| **fibrinogen_max** | Fibrinogen (Max) | continuous | Maximum Fibrinogen measurement for this patient's hospitalization | mg/dL | Labs | 0 | | | | | labs |
| **fibrinogen_mean** | Fibrinogen (Mean) | continuous | Average Fibrinogen measurement for this patient's hospitalization | mg/dL | Labs | 0 | | | | | labs |
| **fibrinogen_median** | Fibrinogen (Median) | continuous | Median Fibrinogen measurement for this patient's hospitalization | mg/dL | Labs | 0 | | | | | labs |
| **fibrinogen_min** | Fibrinogen (Min) | continuous | Minimum Fibrinogen measurement for this patient's hospitalization | mg/dL | Labs | 0 | | | | | labs |
| **fibrinogen_sd** | Fibrinogen (SD) | continuous | Standard deviation of the Fibrinogen measurements during this patient's hospitalization | mg/dL | Labs | 0 | | | | | labs |
| **fio2_25** | Oxygen Flow Rate (25th Percentile) | continuous | Oxygen Flow Rate at the 25th percentile of this patient's measurements | mm Hg | Labs | 0 | | | | | labs |
| **fio2_75** | Oxygen Flow Rate (75th Percentile) | continuous | Oxygen Flow Rate at the 75th percentile of this patient's measurements | mm Hg | Labs | 0 | | | | | labs |
| **fio2_count** | Oxygen Flow Rate (Count) | continuous | Count of Oxygen Flow Rate measurements for this patient | Number of measurements | Labs | 0 | | | | | labs |
| **fio2_first** | Oxygen Flow Rate (First) | continuous | First Oxygen Flow Rate measurement for this patient's hospitalization | mm Hg | Labs | 0 | | | | | labs |
| **fio2_last** | Oxygen Flow Rate (Last) | continuous | Last Oxygen Flow Rate measurement for this patient's hospitalization | mm Hg | Labs | 0 | | | | | labs |
| **fio2_max** | Oxygen Flow Rate (Max) | continuous | Maximum Oxygen Flow Rate measurement for this patient's hospitalization | mm Hg | Labs | 0 | | | | | labs |
| **fio2_mean** | Oxygen Flow Rate (Mean) | continuous | Average Oxygen Flow Rate measurement for this patient's hospitalization | mm Hg | Labs | 0 | | | | | labs |
| **fio2_median** | Oxygen Flow Rate (Median) | continuous | Median Oxygen Flow Rate measurement for this patient's hospitalization | mm Hg | Labs | 0 | | | | | labs |
| **fio2_min** | Oxygen Flow Rate (Min) | continuous | Minimum Oxygen Flow Rate measurement for this patient's hospitalization | mm Hg | Labs | 0 | | | | | labs |
| **fio2_sd** | Oxygen Flow Rate (SD) | continuous | Standard deviation of the Oxygen Flow Rate measurements during this patient's hospitalization | mm Hg | Labs | 0 | | | | | labs |
| **first_aki_time** | | | | | Labs | 0 | | | | | |
| **flua_first** | Influenza A (First) | categorical | Results of the first influenza A test during this patient's hospitalization | | Labs | 0 | | | | | |
| **flua_last** | Influenza A (Last) | categorical | Results of the last influenza A test during this patient's hospitalization | | Labs | 0 | | | | | |
| **fluaab_first** | | | | | Labs | 0 | | | | | |
| **fluaab_last** | | | | | Labs | 0 | | | | | |
| **flub_first** | Influenza B (First) | categorical | Results of the first influenza B test during this patient's hospitalization | | Labs | 0 | | | | | |
| **flub_last** | Influenza B (First) | categorical | Results of the last influenza B test during this patient's hospitalization | | Labs | 0 | | | | | |
| **flubab_first** | | | | | Labs | 0 | | | | | |
| **flubab_last** | | | | | Labs | 0 | | | | | |
| **flubantibody_first** | Influenza B antibody test (First) | categorical | Results of the first influenza B antibody test during this patient's hospitalization | | Labs | 0 | | | | | labs |
| **flubantibody_last** | Influenza B antibody test (Last) | categorical | Results of the last influenza B antibody test during this patient's hospitalization | | Labs | 0 | | | | | labs |
| **folate_25** | Folate (25th Percentile) | continuous | Folate at the 25th percentile of this patient's measurements | ng/mL | Labs | 0 | | | | | labs |
| **folate_75** | Folate (75th Percentile) | continuous | Folate at the 75th percentile of this patient's measurements | ng/mL | Labs | 0 | | | | | labs |
| **folate_count** | Folate (Count) | continuous | Count of folate measurements for this patient | Number of measurements | Labs | 0 | | | | | labs |
| **folate_first** | Folate (First) | continuous | First folate measurement for this patient's hospitalization | ng/mL | Labs | 0 | | | | | labs |
| **folate_last** | Folate (Last) | continuous | Last folate measurement for this patient's hospitalization | ng/mL | Labs | 0 | | | | | labs |
| **folate_max** | Folate (Max) | continuous | Maximum folate measurement for this patient's hospitalization | ng/mL | Labs | 0 | | | | | labs |
| **folate_mean** | Folate (Mean) | continuous | Average folate measurement for this patient's hospitalization | ng/mL | Labs | 0 | | | | | labs |
| **folate_median** | Folate (Median) | continuous | Median folate measurement for this patient's hospitalization | ng/mL | Labs | 0 | | | | | labs |
| **folate_min** | Folate (Min) | continuous | Minimum folate measurement for this patient's hospitalization | ng/mL | Labs | 0 | | | | | labs |
| **folate_sd** | Folate (SD) | continuous | Standard deviation of the folate measurements during this patient's hospitalization | ng/mL | Labs | 0 | | | | | labs |
| **gammaglob_25** | Gamma Globulin (25th Percentile) | continuous | Gamma Globulin at the 25th percentile of this patient's measurements | g/dL | Labs | 0 | | | | | labs |
| **gammaglob_75** | Gamma Globulin (75th Percentile) | continuous | Gamma Globulin at the 75th percentile of this patient's measurements | g/dL | Labs | 0 | | | | | labs |
| **gammaglob_count** | Gamma Globulin (Count) | continuous | Count of Gamma Globulin measurements for this patient | Number of measurements | Labs | 0 | | | | | labs |
| **gammaglob_first** | Gamma Globulin (First) | continuous | First Gamma Globulin measurement for this patient's hospitalization | g/dL | Labs | 0 | | | | | labs |
| **gammaglob_last** | Gamma Globulin (Last) | continuous | Last Gamma Globulin measurement for this patient's hospitalization | g/dL | Labs | 0 | | | | | labs |
| **gammaglob_max** | Gamma Globulin (Max) | continuous | Maximum Gamma Globulin measurement for this patient's hospitalization | g/dL | Labs | 0 | | | | | labs |
| **gammaglob_mean** | Gamma Globulin (Mean) | continuous | Average Gamma Globulin measurement for this patient's hospitalization | g/dL | Labs | 0 | | | | | labs |
| **gammaglob_median** | Gamma Globulin (Median) | continuous | Median Gamma Globulin measurement for this patient's hospitalization | g/dL | Labs | 0 | | | | | labs |
| **gammaglob_min** | Gamma Globulin (Min) | continuous | Minimum Gamma Globulin measurement for this patient's hospitalization | g/dL | Labs | 0 | | | | | labs |
| **gammaglob_sd** | Gamma Globulin (SD) | continuous | Standard deviation of the Gamma Globulin measurements during this patient's hospitalization | g/dL | Labs | 0 | | | | | labs |
| **gbm_25** | Glioblastoma Multiforme (25th Percentile) | continuous | Glioblastoma Multiforme at the 25th percentile of this patient's measurements | EU/mL | Labs | 0 | | | | | labs |
| **gbm_75** | Glioblastoma Multiforme (75th Percentile) | continuous | Glioblastoma Multiforme at the 75th percentile of this patient's measurements | EU/mL | Labs | 0 | | | | | labs |
| **gbm_count** | Glioblastoma Multiforme (Count) | continuous | Count of Glioblastoma Multiforme measurements for this patient | Number of measurements | Labs | 0 | | | | | labs |
| **gbm_first** | Glioblastoma Multiforme (First) | continuous | First Glioblastoma Multiforme measurement for this patient's hospitalization | EU/mL | Labs | 0 | | | | | labs |
| **gbm_last** | Glioblastoma Multiforme (Last) | continuous | Last Glioblastoma Multiforme measurement for this patient's hospitalization | EU/mL | Labs | 0 | | | | | labs |
| **gbm_max** | Glioblastoma Multiforme (Max) | continuous | Maximum Glioblastoma Multiforme measurement for this patient's hospitalization | EU/mL | Labs | 0 | | | | | labs |
| **gbm_mean** | Glioblastoma Multiforme (Mean) | continuous | Average Glioblastoma Multiforme measurement for this patient's hospitalization | EU/mL | Labs | 0 | | | | | labs |
| **gbm_median** | Glioblastoma Multiforme (Median) | continuous | Median Glioblastoma Multiforme measurement for this patient's hospitalization | EU/mL | Labs | 0 | | | | | labs |

| | | | | | | | | | | | |
|---|---|---|---|---|---|---|---|---|---|---|---|
| **gbm_min** | Glioblastoma Multiforme (Min) | continuous | Minimum Glioblastoma Multiforme measurement for this patient's hospitalization | EU/mL | Labs | 0 | | | | | labs |
| **gbm_sd** | Glioblastoma Multiforme (SD) | continuous | Standard deviation of the Glioblastoma Multiforme measurements during this patient's hospitalization | EU/mL | Labs | 0 | | | | | labs |
| **gentamicin_lab_25** | Gentamicin (25th Percentile) | continuous | Gentamicin at the 25th percentile of this patient's measurements | μg/mL | Labs | 0 | | | | | labs |
| **gentamicin_lab_75** | Gentamicin (75th Percentile) | continuous | Gentamicin at the 75th percentile of this patient's measurements | μg/mL | Labs | 0 | | | | | labs |
| **gentamicin_lab_count** | Gentamicin (Count) | continuous | Count of gentamicin measurements for this patient | Number of measurements | Labs | 0 | | | | | labs |
| **gentamicin_lab_first** | Gentamicin (First) | continuous | First gentamicin measurement for this patient's hospitalization | μg/mL | Labs | 0 | | | | | labs |
| **gentamicin_lab_last** | Gentamicin (Last) | continuous | Last gentamicin measurement for this patient's hospitalization | μg/mL | Labs | 0 | | | | | labs |
| **gentamicin_lab_max** | Gentamicin (Max) | continuous | Maximum gentamicin measurement for this patient's hospitalization | μg/mL | Labs | 0 | | | | | labs |
| **gentamicin_lab_mean** | Gentamicin (Mean) | continuous | Average gentamicin measurement for this patient's hospitalization | μg/mL | Labs | 0 | | | | | labs |
| **gentamicin_lab_median** | Gentamicin (Median) | continuous | Median gentamicin measurement for this patient's hospitalization | μg/mL | Labs | 0 | | | | | labs |
| **gentamicin_lab_min** | Gentamicin (Min) | continuous | Minimum gentamicin measurement for this patient's hospitalization | μg/mL | Labs | 0 | | | | | labs |
| **gentamicin_lab_sd** | Gentamicin (SD) | continuous | Standard deviation of the gentamicin measurements during this patient's hospitalization | μg/mL | Labs | 0 | | | | | labs |
| **ggt_25** | Gamma-glutamyl transferase (25th Percentile) | continuous | Gamma-glutamyl transferase at the 25th percentile of this patient's measurements | U/L | Labs | 0 | | | | | labs |
| **ggt_75** | Gamma-glutamyl transferase (75th Percentile) | continuous | Gamma-glutamyl transferase at the 75th percentile of this patient's measurements | U/L | Labs | 0 | | | | | labs |
| **ggt_count** | Gamma-glutamyl transferase (Count) | continuous | Count of gamma-glutamyl transferase measurements for this patient | Number of measurements | Labs | 0 | | | | | labs |
| **ggt_first** | Gamma-glutamyl transferase (First) | continuous | First gamma-glutamyl transferase measurement for this patient's hospitalization | U/L | Labs | 0 | | | | | labs |
| **ggt_last** | Gamma-glutamyl transferase (Last) | continuous | Last gamma-glutamyl transferase measurement for this patient's hospitalization | U/L | Labs | 0 | | | | | labs |
| **ggt_max** | Gamma-glutamyl transferase (Max) | continuous | Maximum gamma-glutamyl transferase measurement for this patient's hospitalization | U/L | Labs | 0 | | | | | labs |
| **ggt_mean** | Gamma-glutamyl transferase (Mean) | continuous | Average gamma-glutamyl transferase measurement for this patient's hospitalization | U/L | Labs | 0 | | | | | labs |
| **ggt_median** | Gamma-glutamyl transferase (Median) | continuous | Median gamma-glutamyl transferase measurement for this patient's hospitalization | U/L | Labs | 0 | | | | | labs |
| **ggt_min** | Gamma-glutamyl transferase (Min) | continuous | Minimum gamma-glutamyl transferase measurement for this patient's hospitalization | U/L | Labs | 0 | | | | | labs |
| **ggt_sd** | Gamma-glutamyl transferase (SD) | continuous | Standard deviation of the gamma-glutamyl transferase measurements during this patient's hospitalizatio | U/L | Labs | 0 | | | | | labs |
| **globulin_25** | Globulin (25th Percentile) | continuous | Globulin at the 25th percentile of this patient's measurements | g/dL | Labs | 0 | | | | | labs |
| **globulin_75** | Globulin (75th Percentile) | continuous | Globulin at the 75th percentile of this patient's measurements | g/dL | Labs | 0 | | | | | labs |
| **globulin_count** | Globulin (Count) | continuous | Count of globulin measurements for this patient | Number of measurements | Labs | 0 | | | | | labs |
| **globulin_first** | Globulin (First) | continuous | First globulin measurement for this patient's hospitalization | g/dL | Labs | 0 | | | | | labs |
| **globulin_last** | Globulin (Last) | continuous | Last globulin measurement for this patient's hospitalization | g/dL | Labs | 0 | | | | | labs |
| **globulin_max** | Globulin (Max) | continuous | Maximum globulin measurement for this patient's hospitalization | g/dL | Labs | 0 | | | | | labs |
| **globulin_mean** | Globulin (Mean) | continuous | Average globulin measurement for this patient's hospitalization | g/dL | Labs | 0 | | | | | labs |
| **globulin_median** | Globulin (Median) | continuous | Median globulin measurement for this patient's hospitalization | g/dL | Labs | 0 | | | | | labs |
| **globulin_min** | Globulin (Min) | continuous | Minimum globulin measurement for this patient's hospitalization | g/dL | Labs | 0 | | | | | labs |
| **globulin_sd** | Globulin (SD) | continuous | Standard deviation of the globulin measurements during this patient's hospitalization | g/dL | Labs | 0 | | | | | labs |
| **glucose_25** | Glucose (25th Percentile) | continuous | Glucose at the 25th percentile of this patient's measurements | mg/dL | Labs | 0 | | | | | labs |
| **glucose_75** | Glucose (75th Percentile) | continuous | Glucose at the 75th percentile of this patient's measurements | mg/dL | Labs | 0 | | | | | labs |
| **glucose_count** | Glucose (Count) | continuous | Count of glucose measurements for this patient | Number of measurements | Labs | 0 | | | | | labs |
| **glucose_first** | Glucose (First) | continuous | First glucose measurement for this patient's hospitalization | mg/dL | Labs | 0 | | | | | labs |
| **glucose_last** | Glucose (Last) | continuous | Last glucose measurement for this patient's hospitalization | mg/dL | Labs | 0 | | | | | labs |
| **glucose_max** | Glucose (Max) | continuous | Maximum glucose measurement for this patient's hospitalization | mg/dL | Labs | 0 | | | | | labs |
| **glucose_mean** | Glucose (Mean) | continuous | Average glucose measurement for this patient's hospitalization | mg/dL | Labs | 0 | | | | | labs |
| **glucose_median** | Glucose (Median) | continuous | Median glucose measurement for this patient's hospitalization | mg/dL | Labs | 0 | | | | | labs |
| **glucose_min** | Glucose (Min) | continuous | Minimum glucose measurement for this patient's hospitalization | mg/dL | Labs | 0 | | | | | labs |
| **glucose_sd** | Glucose (SD) | continuous | Standard deviation of the glucose measurements during this patient's hospitalization | mg/dL | Labs | 0 | | | | | labs |
| **gramstain_first** | Gram Stain (First) | categorical | Results of the first Gram Stain test during this patient's hospitalization | | Labs | 0 | | | | | |
| **gramstain_last** | Gram Stain (Last) | categorical | Results of the last Gram Stain test during this patient's hospitalization | | Labs | 0 | | | | | |
| **granulocytes_25** | Granulocytes (25th Percentile) | continuous | Granulocytes at the 25th percentile of this patient's measurements | mm3 | Labs | 0 | | | | | labs |
| **granulocytes_75** | Granulocytes (75th Percentile) | continuous | Granulocytes at the 75th percentile of this patient's measurements | mm3 | Labs | 0 | | | | | labs |
| **granulocytes_count** | Granulocytes (Count) | continuous | Count of granulocytes measurements for this patient | Number of measurements | Labs | 0 | | | | | labs |
| **granulocytes_first** | Granulocytes (First) | continuous | First granulocytes measurement for this patient's hospitalization | mm3 | Labs | 0 | | | | | labs |
| **granulocytes_last** | Granulocytes (Last) | continuous | Last granulocytes measurement for this patient's hospitalization | mm3 | Labs | 0 | | | | | labs |
| **granulocytes_max** | Granulocytes (Max) | continuous | Maximum granulocytes measurement for this patient's hospitalization | mm3 | Labs | 0 | | | | | labs |
| **granulocytes_mean** | Granulocytes (Mean) | continuous | Average granulocytes measurement for this patient's hospitalization | mm3 | Labs | 0 | | | | | labs |
| **granulocytes_median** | Granulocytes (Median) | continuous | Median granulocytes measurement for this patient's hospitalization | mm3 | Labs | 0 | | | | | labs |
| **granulocytes_min** | Granulocytes (Min) | continuous | Minimum granulocytes measurement for this patient's hospitalization | mm3 | Labs | 0 | | | | | labs |
| **granulocytes_sd** | Granulocytes (SD) | continuous | Standard deviation of the granulocytes measurements during this patient's hospitalization | mm3 | Labs | 0 | | | | | labs |
| **haptoglobin_25** | Haptoglobin (25th Percentile) | continuous | Haptoglobin at the 25th percentile of this patient's measurements | g/L | Labs | 0 | | | | | labs |
| **haptoglobin_75** | Haptoglobin (75th Percentile) | continuous | haptoglobin at the 75th percentile of this patient's measurements | g/L | Labs | 0 | | | | | labs |
| **haptoglobin_count** | Haptoglobin (Count) | continuous | Count of haptoglobin measurements for this patient | Number of measurements | Labs | 0 | | | | | labs |
| **haptoglobin_first** | Haptoglobin (First) | continuous | First haptoglobin measurement for this patient's hospitalization | g/L | Labs | 0 | | | | | labs |
| **haptoglobin_last** | Haptoglobin (Last) | continuous | Last haptoglobin measurement for this patient's hospitalization | g/L | Labs | 0 | | | | | labs |
| **haptoglobin_max** | Haptoglobin (Max) | continuous | Maximum haptoglobin measurement for this patient's hospitalization | g/L | Labs | 0 | | | | | labs |
| **haptoglobin_mean** | Haptoglobin (Mean) | continuous | Average haptoglobin measurement for this patient's hospitalization | g/L | Labs | 0 | | | | | labs |
| **haptoglobin_median** | Haptoglobin (Median) | continuous | Median haptoglobin measurement for this patient's hospitalization | g/L | Labs | 0 | | | | | labs |
| **haptoglobin_min** | Haptoglobin (Min) | continuous | Minimum haptoglobin measurement for this patient's hospitalization | g/L | Labs | 0 | | | | | labs |
| **haptoglobin_sd** | Haptoglobin (SD) | continuous | Standard deviation of the haptoglobin measurements during this patient's hospitalization | g/L | Labs | 0 | | | | | labs |
| **hba1c_25** | Hemoglobin A1c (25th Percentile) | continuous | Hemoglobin A1c at the 25th percentile of this patient's measurements | g/dL | Labs | 0 | | | | | labs |
| **hba1c_75** | Hemoglobin A1c (75th Percentile) | continuous | Hemoglobin A1c at the 75th percentile of this patient's measurements | g/dL | Labs | 0 | | | | | labs |
| **hba1c_count** | Hemoglobin A1c (Count) | continuous | Count of hemoglobin A1c measurements for this patient | Number of measurements | Labs | 0 | | | | | labs |
| **hba1c_first** | Hemoglobin A1c (First) | continuous | First hemoglobin A1c measurement for this patient's hospitalization | g/dL | Labs | 0 | | | | | labs |
| **hba1c_last** | Hemoglobin A1c (Last) | continuous | Last hemoglobin A1c measurement for this patient's hospitalization | g/dL | Labs | 0 | | | | | labs |
| **hba1c_max** | Hemoglobin A1c (Max) | continuous | Maximum hemoglobin A1c measurement for this patient's hospitalization | g/dL | Labs | 0 | | | | | labs |
| **hba1c_mean** | Hemoglobin A1c (Mean) | continuous | Average hemoglobin A1c measurement for this patient's hospitalization | g/dL | Labs | 0 | | | | | labs |
| **hba1c_median** | Hemoglobin A1c (Median) | continuous | Median hemoglobin A1c measurement for this patient's hospitalization | g/dL | Labs | 0 | | | | | labs |
| **hba1c_min** | Hemoglobin A1c (Min) | continuous | Minimum hemoglobin A1c measurement for this patient's hospitalization | g/dL | Labs | 0 | | | | | labs |
| **hba1c_sd** | Hemoglobin A1c (SD) | continuous | Standard deviation of the hemoglobin A1c measurements during this patient's hospitalization | g/dL | Labs | 0 | | | | | labs |
| **hbeantibody_first** | Hepatitis B e-Antibody (First) | categorical | Results of the first Hepatitis B e-Antibody test during this patient's hospitalization | mIU | Labs | 0 | | | | | labs |
| **hbeantibody_last** | Hepatitis B e-Antibody (Last) | categorical | Results of the last Hepatitis B e-Antibody test during this patient's hospitalization | mIU | Labs | 0 | | | | | labs |
| **hbeantigen_first** | Hepatitis B e-Antigen (First) | categorical | Results of the first Hepatitis B e-Antigen test during this patient's hospitalization | IU/mL | Labs | 0 | | | | | labs |
| **hbeantigen_last** | Hepatitis B e-Antigen (Last) | categorical | Results of the last Hepatitis B e-Antigen test during this patient's hospitalization | IU/mL | Labs | 0 | | | | | labs |
| **hbsab_25** | Hepatitis B Surface Antibody (25th Percentile) | continuous | Hepatitis B Surface Antibody at the 25th percentile of this patient's measurements | mIU | Labs | 0 | | | | | labs |
| **hbsab_75** | Hepatitis B Surface Antibody (75th Percentile) | continuous | Hepatitis B Surface Antibody at the 75th percentile of this patient's measurements | mIU | Labs | 0 | | | | | labs |
| **hbsab_count** | Hepatitis B Surface Antibody (Count) | continuous | Count of Hepatitis B Surface Antibody measurements for this patient | Number of measurements | Labs | 0 | | | | | labs |
| **hbsab_first** | Hepatitis B Surface Antibody (First) | continuous | First Hepatitis B Surface Antibody measurement for this patient's hospitalization | mIU | Labs | 0 | | | | | labs |
| **hbsab_last** | Hepatitis B Surface Antibody (Last) | continuous | Last Hepatitis B Surface Antibody measurement for this patient's hospitalization | mIU | Labs | 0 | | | | | labs |
| **hbsab_max** | Hepatitis B Surface Antibody (Max) | continuous | Maximum Hepatitis B Surface Antibody measurement for this patient's hospitalization | mIU | Labs | 0 | | | | | labs |
| **hbsab_mean** | Hepatitis B Surface Antibody (Mean) | continuous | Average Hepatitis B Surface Antibody measurement for this patient's hospitalization | mIU | Labs | 0 | | | | | labs |
| **hbsab_median** | Hepatitis B Surface Antibody (Median) | continuous | Median Hepatitis B Surface Antibody measurement for this patient's hospitalization | mIU | Labs | 0 | | | | | labs |
| **hbsab_min** | Hepatitis B Surface Antibody (Min) | continuous | Minimum Hepatitis B Surface Antibody measurement for this patient's hospitalization | mIU | Labs | 0 | | | | | labs |
| **hbsab_sd** | Hepatitis B Surface Antibody (SD) | continuous | Standard deviation of the Hepatitis B Surface Antibody measurements during this patient's hospitalizatio | mIU | Labs | 0 | | | | | labs |
| **hcg_25** | Human Chorionic Gonadotropin (25th Percentile) | continuous | Human Chorionic Gonadotropin at the 25th percentile of this patient's measurements | mIU/mL | Labs | 0 | | | | | labs |
| **hcg_75** | Human Chorionic Gonadotropin (75th Percentile) | continuous | Human Chorionic Gonadotropin at the 75th percentile of this patient's measurements | mIU/mL | Labs | 0 | | | | | labs |
| **hcg_count** | Human Chorionic Gonadotropin (Count) | continuous | Count of Human Chorionic Gonadotropin measurements for this patient | Number of measurements | Labs | 0 | | | | | labs |
| **hcg_first** | Human Chorionic Gonadotropin (First) | continuous | First Human Chorionic Gonadotropin measurement for this patient's hospitalization | mIU/mL | Labs | 0 | | | | | labs |
| **hcg_last** | Human Chorionic Gonadotropin (Last) | continuous | Last Human Chorionic Gonadotropin measurement for this patient's hospitalization | mIU/mL | Labs | 0 | | | | | labs |
| **hcg_max** | Human Chorionic Gonadotropin (Max) | continuous | Maximum Human Chorionic Gonadotropin measurement for this patient's hospitalization | mIU/mL | Labs | 0 | | | | | labs |
| **hcg_mean** | Human Chorionic Gonadotropin (Mean) | continuous | Average Human Chorionic Gonadotropin measurement for this patient's hospitalization | mIU/mL | Labs | 0 | | | | | labs |
| **hcg_median** | Human Chorionic Gonadotropin (Median) | continuous | Median Human Chorionic Gonadotropin measurement for this patient's hospitalization | mIU/mL | Labs | 0 | | | | | labs |
| **hcg_min** | Human Chorionic Gonadotropin (Min) | continuous | Minimum Human Chorionic Gonadotropin measurement for this patient's hospitalization | mIU/mL | Labs | 0 | | | | | labs |
| **hcg_sd** | Human Chorionic Gonadotropin (SD) | continuous | Standard deviation of the Human Chorionic Gonadotropin measurements during this patient's hospitaliza | mIU/mL | Labs | 0 | | | | | labs |
| **hct_25** | Hematocrit (25th Percentile) | continuous | Hematocrit at the 25th percentile of this patient's measurements | % | Labs | 0 | | | | | labs |
| **hct_75** | Hematocrit (75th Percentile) | continuous | Hematocrit at the 75th percentile of this patient's measurements | % | Labs | 0 | | | | | labs |
| **hct_count** | Hematocrit (Count) | continuous | Count of hematocrit measurements for this patient | Number of measurements | Labs | 0 | | | | | labs |
| **hct_first** | Hematocrit (First) | continuous | First hematocrit measurement for this patient's hospitalization | % | Labs | 0 | | | | | labs |
| **hct_last** | Hematocrit (Last) | continuous | Last hematocrit measurement for this patient's hospitalization | % | Labs | 0 | | | | | labs |
| **hct_max** | Hematocrit (Max) | continuous | Maximum hematocrit measurement for this patient's hospitalization | % | Labs | 0 | | | | | labs |
| **hct_mean** | Hematocrit (Mean) | continuous | Average hematocrit measurement for this patient's hospitalization | % | Labs | 0 | | | | | labs |
| **hct_median** | Hematocrit (Median) | continuous | Median hematocrit measurement for this patient's hospitalization | % | Labs | 0 | | | | | labs |
| **hct_min** | Hematocrit (Min) | continuous | Minimum hematocrit measurement for this patient's hospitalization | % | Labs | 0 | | | | | labs |
| **hct_sd** | Hematocrit (SD) | continuous | Standard deviation of the hematocrit measurements during this patient's hospitalization | % | Labs | 0 | | | | | labs |
| **hcvab_first** | Hepatitis C Virus Antibody (First) | categorical | Results of the first Hepatitis C Virus Antibody test during this patient's hospitalization | IU/mL | Labs | 0 | | | | | labs |
| **hcvab_last** | Hepatitis C Virus Antibody (Last) | categorical | Results of the last Hepatitis C Virus Antibody test during this patient's hospitalization | IU/mL | Labs | 0 | | | | | labs |
| **hcvgenotype_first** | Hepatitis C Virus Genotype (First) | categorical | Results of the first Hepatitis C Virus Genotype test during this patient's hospitalization | IU/mL | Labs | 0 | | | | | labs |
| **hcvgenotype_last** | Hepatitis C Virus Genotype (Last) | categorical | Results of the last Hepatitis C Virus Genotype test during this patient's hospitalization | IU/mL | Labs | 0 | | | | | labs |
| **hcvlog_first** | Hepatitis C Virus Log (First) | categorical | Results of the first Hepatitis C Virus Log during this patient's hospitalization | | Labs | 0 | | | | | |
| **hcvlog_last** | Hepatitis C Virus Log (Last) | categorical | Results of the last Hepatitis C Virus Log during this patient's hospitalization | | Labs | 0 | | | | | |
| **hcvquant_first** | Hepatitis C Virus Quantitative (First) | categorical | Results of the first Hepatitis C Virus Quantitative test during this patient's hospitalization | IU/L | Labs | 0 | | | | | labs |
| **hcvquant_last** | Hepatitis C Virus Quantitative (Last) | categorical | Results of the last Hepatitis C Virus Quantitative test during this patient's hospitalization | IU/L | Labs | 0 | | | | | labs |
| **hebpsab_first** | Hepatitis B Surface Antibody (First) | categorical | First Hepatitis B Surface Antibody measurement for this patient's hospitalization | | Labs | 0 | | | | | labs |
| **hebpsab_last** | Hepatitis B Surface Antibody (Last) | categorical | Last Hepatitis B Surface Antibody measurement for this patient's hospitalization | | Labs | 0 | | | | | labs |
| **hemoglobin_25** | Hemoglobin (25th Percentile) | continuous | Hemoglobin at the 25th percentile of this patient's measurements | g/dL | Labs | 0 | | | | | labs |
| **hemoglobin_75** | Hemoglobin (75th Percentile) | continuous | Hemoglobin at the 75th percentile of this patient's measurements | g/dL | Labs | 0 | | | | | labs |
| **hemoglobin_count** | Hemoglobin (Count) | continuous | Count of hemoglobin measurements for this patient | Number of measurements | Labs | 0 | | | | | labs |
| **hemoglobin_first** | Hemoglobin (First) | continuous | First hemoglobin measurement for this patient's hospitalization | g/dL | Labs | 0 | | | | | labs |
| **hemoglobin_last** | Hemoglobin (Last) | continuous | Last hemoglobin measurement for this patient's hospitalization | g/dL | Labs | 0 | | | | | labs |
| **hemoglobin_max** | Hemoglobin (Max) | continuous | Maximum hemoglobin measurement for this patient's hospitalization | g/dL | Labs | 0 | | | | | labs |

| | | | | | | | | | | | |
|---|---|---|---|---|---|---|---|---|---|---|---|
| **hemoglobin_mean** | Hemoglobin (Mean) | continuous | Average hemoglobin measurement for this patient's hospitalization | g/dL | Labs | 0 | | | | | labs |
| **hemoglobin_median** | Hemoglobin (Median) | continuous | Median hemoglobin measurement for this patient's hospitalization | g/dL | Labs | 0 | | | | | labs |
| **hemoglobin_min** | Hemoglobin (Min) | continuous | Minimum hemoglobin measurement for this patient's hospitalization | g/dL | Labs | 0 | | | | | labs |
| **hemoglobin_sd** | Hemoglobin (SD) | continuous | Standard deviation of the hemoglobin measurements during this patient's hospitalization | g/dL | Labs | 0 | | | | | labs |
| **hepbcab_first** | Hepatitis B CAB (First) | categorical | Results of the first Hepatitis B CAB test during this patient's hospitalization | | Labs | 0 | | | | | labs |
| **hepbcab_last** | Hepatitis B CAB (Last) | categorical | Results of the last Hepatitis B CAB test during this patient's hospitalization | | Labs | 0 | | | | | labs |
| **hepbcigm_first** | Hepatitis B Cytoplasmic Immunoglobulin (First) | categorical | Results of the first Hepatitis B Cytoplasmic Immunoglobulin test during this patient's hospitalization | | Labs | 0 | | | | | labs |
| **hepbcigm_last** | Hepatitis B Cytoplasmic Immunoglobulin (Last) | categorical | Results of the last Hepatitis B Cytoplasmic Immunoglobulin test during this patient's hospitalization | | Labs | 0 | | | | | labs |
| **hepbcore_first** | Hepatitis B Core (First) | categorical | Results of the first Hepatitis B Core test during this patient's hospitalization | | Labs | 0 | | | | | labs |
| **hepbcore_last** | Hepatitis B Core (Last) | categorical | Results of the last Hepatitis B Core test during this patient's hospitalization | | Labs | 0 | | | | | labs |
| **hepbdna_first** | Hepatitis B DNA Test (First) | categorical | Results of the first Hepatitis B DNA test during this patient's hospitalization | | Labs | 0 | | | | | |
| **hepbdna_last** | Hepatitis B DNA Test (Last) | categorical | Results of the last Hepatitis B DNA test during this patient's hospitalization | | Labs | 0 | | | | | |
| **hepbquantcat_first** | | | | | Labs | 0 | | | | | labs |
| **hepbquantcat_last** | | | | | Labs | 0 | | | | | labs |
| **hepbsab_first** | Hepatitis B Surface Antibody (First) | categorical | First Hepatitis B Surface Antibody measurement for this patient's hospitalization | | Labs | 0 | | | | | labs |
| **hepbsab_last** | Hepatitis B Surface Antibody (Last) | categorical | Last Hepatitis B Surface Antibody measurement for this patient's hospitalization | | Labs | 0 | | | | | labs |
| **hepbsag_first** | Hepatitis B Surface Antigen (First) | categorical | First Hepatitis B Surface Antigen measurement for this patient's hospitalization | | Labs | 0 | | | | | labs |
| **hepbsag_last** | Hepatitis B Surface Antigen (Last) | categorical | Last Hepatitis B Surface Antigen measurement for this patient's hospitalization | | Labs | 0 | | | | | labs |
| **historical_aki** | Historical Acquired Kidney Injury | categorical | Acquired Kidney Injury during admission | | Labs | 0 | | | | | aki_flag |
| **historical_aki_stage** | Historical Acquired Kidney Injury Stage | categorical | Stage of Historical Acquired Kidney Injury diagnosis for this patient | | Labs | 0 | | | | | |
| **historical_baseline_crea** | Historical Baseline Creatinine | continuous | Historical baseline creatinine levels | Mg/dL | Labs | 0 | | | | | aki_flag |
| **hittlevel_first** | | categorical | | | Labs | 0 | | | | | labs |
| **hittlevel_last** | | categorical | | | Labs | 0 | | | | | labs |
| **hiv1gen_first** | HIV 1 Genotype (First) | categorical | Results of the first HIV 1 Genotype test during this patient's hospitalization | | Labs | 0 | | | | | labs |
| **hiv1gen_last** | HIV 1 Genotype (Last) | categorical | Results of the last HIV 1 Genotype test during this patient's hospitalization | | Labs | 0 | | | | | labs |
| **hiv1rna_first** | HIV 1 RNA (First) | categorical | Results of the first HIV 1 RNA test during this patient's hospitalization | | Labs | 0 | | | | | labs |
| **hiv1rna_last** | HIV 1 RNA (Last) | categorical | Results of the last HIV 1 RNA test during this patient's hospitalization | | Labs | 0 | | | | | labs |
| **hiv2ant_first** | HIV 2 Antibody (First) | categorical | Results of the first HIV 2 Antibody test during this patient's hospitalization | | Labs | 0 | | | | | labs |
| **hiv2ant_last** | HIV 2 Antibody (Last) | categorical | Results of the last HIV 2 Antibody test during this patient's hospitalization | | Labs | 0 | | | | | labs |
| **hivab_25** | HIV Antibody (25th Percentile) | continuous | HIV Antibody at the 25th percentile of this patient's measurements | +/- | Labs | 0 | | | | | labs |
| **hivab_75** | HIV Antibody (75th Percentile) | continuous | HIV Antibody at the 75th percentile of this patient's measurements | +/- | Labs | 0 | | | | | labs |
| **hivab_count** | HIV Antibody (Count) | continuous | Count of HIV Antibody measurements for this patient | Number of measurements | Labs | 0 | | | | | labs |
| **hivab_first** | HIV Antibody (First) | continuous | First HIV Antibody measurement for this patient's hospitalization | +/- | Labs | 0 | | | | | labs |
| **hivab_last** | HIV Antibody (Last) | continuous | Last HIV Antibody measurement for this patient's hospitalization | +/- | Labs | 0 | | | | | labs |
| **hivab_max** | HIV Antibody (Max) | continuous | Maximum HIV Antibody measurement for this patient's hospitalization | +/- | Labs | 0 | | | | | labs |
| **hivab_mean** | HIV Antibody (Mean) | continuous | Average HIV Antibody measurement for this patient's hospitalization | +/- | Labs | 0 | | | | | labs |
| **hivab_median** | HIV Antibody (Median) | continuous | Median HIV Antibody measurement for this patient's hospitalization | +/- | Labs | 0 | | | | | labs |
| **hivab_min** | HIV Antibody (Min) | continuous | Minimum HIV Antibody measurement for this patient's hospitalization | +/- | Labs | 0 | | | | | labs |
| **hivab_sd** | HIV Antibody (SD) | continuous | Standard deviation of the HIV Antibody measurements during this patient's hospitalization | +/- | Labs | 0 | | | | | labs |
| **hivlog_first** | HIV Log (First) | categorical | First HIV Log measurement for this patient's hospitalization | | Labs | 0 | | | | | |
| **hivlog_last** | HIV Log (Last) | categorical | Last HIV Log measurement for this patient's hospitalization | | Labs | 0 | | | | | |
| **hivqnt_25** | HIV Quantitative (25th Percentile) | continuous | HIV Quantitative at the 25th percentile of this patient's measurements | | Labs | 0 | | | | | labs |
| **hivqnt_75** | HIV Quantitative (75th Percentile) | continuous | HIV Quantitative at the 75th percentile of this patient's measurements | | Labs | 0 | | | | | labs |
| **hivqnt_count** | HIV Quantitative (Count) | continuous | Count of HIV Quantitative measurements for this patient | Number of measurements | Labs | 0 | | | | | labs |
| **hivqnt_first** | HIV Quantitative (First) | continuous | First HIV Quantitative measurement for this patient's hospitalization | | Labs | 0 | | | | | labs |
| **hivqnt_last** | HIV Quantitative (Last) | continuous | Last HIV Quantitative measurement for this patient's hospitalization | | Labs | 0 | | | | | labs |
| **hivqnt_max** | HIV Quantitative (Max) | continuous | Maximum HIV Quantitative measurement for this patient's hospitalization | | Labs | 0 | | | | | labs |
| **hivqnt_mean** | HIV Quantitative (Mean) | continuous | Average HIV Quantitative measurement for this patient's hospitalization | | Labs | 0 | | | | | labs |
| **hivqnt_median** | HIV Quantitative (Median) | continuous | Median HIV Quantitative measurement for this patient's hospitalization | | Labs | 0 | | | | | labs |
| **hivqnt_min** | HIV Quantitative (Min) | continuous | Minimum HIV Quantitative measurement for this patient's hospitalization | | Labs | 0 | | | | | labs |
| **hivqnt_sd** | HIV Quantitative (SD) | continuous | Standard deviation of the HIV Quantitative measurements during this patient's hospitalization | | Labs | 0 | | | | | labs |
| **hmpv_first** | Human Metapneumovirus(First) | categorical | Results of the first Human Metapneumovirus test during this patient's hospitalization | | Labs | 0 | | | | | labs |
| **hmpv_last** | Human Metapneumovirus (Last) | categorical | Results of the last Human Metapneumovirus test during this patient's hospitalization | | Labs | 0 | | | | | labs |
| **hscrp_25** | High Sensitivity C-Reactive Protein (25th Percentile) | continuous | High Sensitivity C-Reactive Protein at the 25th percentile of this patient's measurements | mg/L | Labs | 0 | | | | | labs |
| **hscrp_75** | High Sensitivity C-Reactive Protein (75th Percentile) | continuous | High Sensitivity C-Reactive Protein at the 75th percentile of this patient's measurements | mg/L | Labs | 0 | | | | | labs |
| **hscrp_count** | High Sensitivity C-Reactive Protein (Count) | continuous | Count of High Sensitivity C-Reactive Protein measurements for this patient | Number of measurements | Labs | 0 | | | | | labs |
| **hscrp_first** | High Sensitivity C-Reactive Protein (First) | continuous | First High Sensitivity C-Reactive Protein measurement for this patient's hospitalization | mg/L | Labs | 0 | | | | | labs |
| **hscrp_last** | High Sensitivity C-Reactive Protein (Last) | continuous | Last High Sensitivity C-Reactive Protein measurement for this patient's hospitalization | mg/L | Labs | 0 | | | | | labs |
| **hscrp_max** | High Sensitivity C-Reactive Protein (Max) | continuous | Maximum High Sensitivity C-Reactive Protein measurement for this patient's hospitalization | mg/L | Labs | 0 | | | | | labs |
| **hscrp_mean** | High Sensitivity C-Reactive Protein (Mean) | continuous | Average High Sensitivity C-Reactive Protein measurement for this patient's hospitalization | mg/L | Labs | 0 | | | | | labs |
| **hscrp_median** | High Sensitivity C-Reactive Protein (Median) | continuous | Median High Sensitivity C-Reactive Protein measurement for this patient's hospitalization | mg/L | Labs | 0 | | | | | labs |
| **hscrp_min** | High Sensitivity C-Reactive Protein (Min) | continuous | Minimum High Sensitivity C-Reactive Protein measurement for this patient's hospitalization | mg/L | Labs | 0 | | | | | labs |
| **hscrp_sd** | High Sensitivity C-Reactive Protein (SD) | continuous | Standard deviation of the High Sensitivity C-Reactive Protein measurements during this patient's hospita | mg/L | Labs | 0 | | | | | labs |
| **icalcium_25** | Ionized Calcium (25th Percentile) | continuous | Ionized Calcium at the 25th percentile of this patient's measurements | mg/dL | Labs | 0 | | | | | labs |
| **icalcium_75** | Ionized Calcium (75th Percentile) | continuous | Ionized Calcium at the 75th percentile of this patient's measurements | mg/dL | Labs | 0 | | | | | labs |
| **icalcium_count** | Ionized Calcium (Count) | continuous | Count of Ionized Calcium measurements for this patient | Number of measurements | Labs | 0 | | | | | labs |
| **icalcium_first** | Ionized Calcium (First) | continuous | First Ionized Calcium measurement for this patient's hospitalization | mg/dL | Labs | 0 | | | | | labs |
| **icalcium_last** | Ionized Calcium (Last) | continuous | Last Ionized Calcium measurement for this patient's hospitalization | mg/dL | Labs | 0 | | | | | labs |
| **icalcium_max** | Ionized Calcium (Max) | continuous | Maximum Ionized Calcium measurement for this patient's hospitalization | mg/dL | Labs | 0 | | | | | labs |
| **icalcium_mean** | Ionized Calcium (Mean) | continuous | Average Ionized Calcium measurement for this patient's hospitalization | mg/dL | Labs | 0 | | | | | labs |
| **icalcium_median** | Ionized Calcium (Median) | continuous | Median Ionized Calcium measurement for this patient's hospitalization | mg/dL | Labs | 0 | | | | | labs |
| **icalcium_min** | Ionized Calcium (Min) | continuous | Minimum Ionized Calcium measurement for this patient's hospitalization | mg/dL | Labs | 0 | | | | | labs |
| **icalcium_sd** | Ionized Calcium (SD) | continuous | Standard deviation of the Ionized Calcium measurements during this patient's hospitalization | mg/dL | Labs | 0 | | | | | labs |
| **iga_25** | Immunoglobulin A Nephropathy (25th Percentile) | continuous | Immunoglobulin A Nephropathy at the 25th percentile of this patient's measurements | g/L | Labs | 0 | | | | | labs |
| **iga_75** | Immunoglobulin A Nephropathy (75th Percentile) | continuous | Immunoglobulin A Nephropathy at the 75th percentile of this patient's measurements | g/L | Labs | 0 | | | | | labs |
| **iga_count** | Immunoglobulin A Nephropathy (Count) | continuous | Count of Immunoglobulin A Nephropathy measurements for this patient | Number of measurements | Labs | 0 | | | | | labs |
| **iga_first** | Immunoglobulin A Nephropathy (First) | continuous | First Immunoglobulin A Nephropathy measurement for this patient's hospitalization | g/L | Labs | 0 | | | | | labs |
| **iga_last** | Immunoglobulin A Nephropathy (Last) | continuous | Last Immunoglobulin A Nephropathy measurement for this patient's hospitalization | g/L | Labs | 0 | | | | | labs |
| **iga_max** | Immunoglobulin A Nephropathy (Max) | continuous | Maximum Immunoglobulin A Nephropathy measurement for this patient's hospitalization | g/L | Labs | 0 | | | | | labs |
| **iga_mean** | Immunoglobulin A Nephropathy (Mean) | continuous | Average Immunoglobulin A Nephropathy measurement for this patient's hospitalization | g/L | Labs | 0 | | | | | labs |
| **iga_median** | Immunoglobulin A Nephropathy (Median) | continuous | Median Immunoglobulin A Nephropathy measurement for this patient's hospitalization | g/L | Labs | 0 | | | | | labs |
| **iga_min** | Immunoglobulin A Nephropathy (Min) | continuous | Minimum Immunoglobulin A Nephropathy measurement for this patient's hospitalization | g/L | Labs | 0 | | | | | labs |
| **iga_sd** | Immunoglobulin A Nephropathy (SD) | continuous | Standard deviation of the Immunoglobulin A Nephropathy measurements during this patient's hospitaliza | g/L | Labs | 0 | | | | | labs |
| **ige_25** | Immunoglobulin E (25th Percentile) | continuous | Immunoglobulin E at the 25th percentile of this patient's measurements | IU/mL | Labs | 0 | | | | | labs |
| **ige_75** | Immunoglobulin E (75th Percentile) | continuous | Immunoglobulin E at the 75th percentile of this patient's measurements | IU/mL | Labs | 0 | | | | | labs |
| **ige_count** | Immunoglobulin E (Count) | continuous | Count of Immunoglobulin E measurements for this patient | Number of measurements | Labs | 0 | | | | | labs |
| **ige_first** | Immunoglobulin E (First) | continuous | First Immunoglobulin E measurement for this patient's hospitalization | IU/mL | Labs | 0 | | | | | labs |
| **ige_last** | Immunoglobulin E (Last) | continuous | Last Immunoglobulin E measurement for this patient's hospitalization | IU/mL | Labs | 0 | | | | | labs |
| **ige_max** | Immunoglobulin E (Max) | continuous | Maximum Immunoglobulin E measurement for this patient's hospitalization | IU/mL | Labs | 0 | | | | | labs |
| **ige_mean** | Immunoglobulin E (Mean) | continuous | Average Immunoglobulin E measurement for this patient's hospitalization | IU/mL | Labs | 0 | | | | | labs |
| **ige_median** | Immunoglobulin E (Median) | continuous | Median Immunoglobulin E measurement for this patient's hospitalization | IU/mL | Labs | 0 | | | | | labs |
| **ige_min** | Immunoglobulin E (Min) | continuous | Minimum Immunoglobulin E measurement for this patient's hospitalization | IU/mL | Labs | 0 | | | | | labs |
| **ige_sd** | Immunoglobulin E (SD) | continuous | Standard deviation of the Immunoglobulin E measurements during this patient's hospitalization | IU/mL | Labs | 0 | | | | | labs |
| **igg_25** | Immunoglobulin G (25th Percentile) | continuous | Immunoglobulin G at the 25th percentile of this patient's measurements | g/L | Labs | 0 | | | | | labs |
| **igg_75** | Immunoglobulin G (75th Percentile) | continuous | Immunoglobulin G at the 75th percentile of this patient's measurements | g/L | Labs | 0 | | | | | labs |
| **igg_count** | Immunoglobulin G (Count) | continuous | Count of Immunoglobulin G measurements for this patient | Number of measurements | Labs | 0 | | | | | labs |
| **igg_first** | Immunoglobulin G (First) | continuous | First Immunoglobulin G measurement for this patient's hospitalization | g/L | Labs | 0 | | | | | labs |
| **igg_last** | Immunoglobulin G (Last) | continuous | Last Immunoglobulin G measurement for this patient's hospitalization | g/L | Labs | 0 | | | | | labs |
| **igg_max** | Immunoglobulin G (Max) | continuous | Maximum Immunoglobulin G measurement for this patient's hospitalization | g/L | Labs | 0 | | | | | labs |
| **igg_mean** | Immunoglobulin G (Mean) | continuous | Average Immunoglobulin G measurement for this patient's hospitalization | g/L | Labs | 0 | | | | | labs |
| **igg_median** | Immunoglobulin G (Median) | continuous | Median Immunoglobulin G measurement for this patient's hospitalization | g/L | Labs | 0 | | | | | labs |
| **igg_min** | Immunoglobulin G (Min) | continuous | Minimum Immunoglobulin G measurement for this patient's hospitalization | g/L | Labs | 0 | | | | | labs |
| **igg_sd** | Immunoglobulin G (SD) | continuous | Standard deviation of the Immunoglobulin G measurements during this patient's hospitalization | g/L | Labs | 0 | | | | | labs |
| **igm_25** | Immunoglobulin M (25th Percentile) | continuous | Immunoglobulin M at the 25th percentile of this patient's measurements | mg/100mL | Labs | 0 | | | | | labs |
| **igm_75** | Immunoglobulin M (75th Percentile) | continuous | Immunoglobulin M at the 75th percentile of this patient's measurements | mg/100mL | Labs | 0 | | | | | labs |
| **igm_count** | Immunoglobulin M (Count) | continuous | Count of Immunoglobulin M measurements for this patient | Number of measurements | Labs | 0 | | | | | labs |
| **igm_first** | Immunoglobulin M (First) | continuous | First Immunoglobulin M measurement for this patient's hospitalization | mg/100mL | Labs | 0 | | | | | labs |
| **igm_last** | Immunoglobulin M (Last) | continuous | Last Immunoglobulin M measurement for this patient's hospitalization | mg/100mL | Labs | 0 | | | | | labs |
| **igm_max** | Immunoglobulin M (Max) | continuous | Maximum Immunoglobulin M measurement for this patient's hospitalization | mg/100mL | Labs | 0 | | | | | labs |
| **igm_mean** | Immunoglobulin M (Mean) | continuous | Average Immunoglobulin M measurement for this patient's hospitalization | mg/100mL | Labs | 0 | | | | | labs |
| **igm_median** | Immunoglobulin M (Median) | continuous | Median Immunoglobulin M measurement for this patient's hospitalization | mg/100mL | Labs | 0 | | | | | labs |
| **igm_min** | Immunoglobulin M (Min) | continuous | Minimum Immunoglobulin M measurement for this patient's hospitalization | mg/100mL | Labs | 0 | | | | | labs |
| **igm_sd** | Immunoglobulin M (SD) | continuous | Standard deviation of the Immunoglobulin M measurements during this patient's hospitalization | mg/100mL | Labs | 0 | | | | | labs |
| **il10_25** | Interleukin 10 (25th Percentile) | continuous | Interleukin 10 at the 25th percentile of this patient's measurements | pg/mL | Labs | 0 | | | | | labs |
| **il10_75** | Interleukin 10 (75th Percentile) | continuous | Interleukin 10 at the 75th percentile of this patient's measurements | pg/mL | Labs | 0 | | | | | labs |
| **il10_count** | Interleukin 10 (Count) | continuous | Count of Interleukin 10 measurements for this patient | Number of measurements | Labs | 0 | | | | | labs |
| **il10_first** | Interleukin 10 (First) | continuous | First Interleukin 10 measurement for this patient's hospitalization | pg/mL | Labs | 0 | | | | | labs |
| **il10_last** | Interleukin 10 (Last) | continuous | Last Interleukin 10 measurement for this patient's hospitalization | pg/mL | Labs | 0 | | | | | labs |
| **il10_max** | Interleukin 10 (Max) | continuous | Maximum Interleukin 10 measurement for this patient's hospitalization | pg/mL | Labs | 0 | | | | | labs |
| **il10_mean** | Interleukin 10 (Mean) | continuous | Average Interleukin 10 measurement for this patient's hospitalization | pg/mL | Labs | 0 | | | | | labs |
| **il10_median** | Interleukin 10 (Median) | continuous | Median Interleukin 10 measurement for this patient's hospitalization | pg/mL | Labs | 0 | | | | | labs |
| **il10_min** | Interleukin 10 (Min) | continuous | Minimum Interleukin 10 measurement for this patient's hospitalization | pg/mL | Labs | 0 | | | | | labs |
| **il10_sd** | Interleukin 10 (SD) | continuous | Standard deviation of the Interleukin 10 measurements during this patient's hospitalization | pg/mL | Labs | 0 | | | | | labs |
| **il12_25** | Interleukin 12 (25th Percentile) | continuous | Interleukin 12 at the 25th percentile of this patient's measurements | pg/mL | Labs | 0 | | | | | labs |

| | | | | | | | | | | | |
|---|---|---|---|---|---|---|---|---|---|---|---|
| il12_75 | Interleukin 12 (75th Percentile) | continuous | Interleukin 12 at the 75th percentile of this patient's measurements | pg/mL | Labs | 0 | | | | | labs |
| il12_count | Interleukin 12 (Count) | continuous | Count of Interleukin 12 measurements for this patient | Number of measurements | Labs | 0 | | | | | labs |
| il12_first | Interleukin 12 (First) | continuous | First Interleukin 12 measurement for this patient's hospitalization | pg/mL | Labs | 0 | | | | | labs |
| il12_last | Interleukin 12 (Last) | continuous | Last Interleukin 12 measurement for this patient's hospitalization | pg/mL | Labs | 0 | | | | | labs |
| il12_max | Interleukin 12 (Max) | continuous | Maximum Interleukin 12 measurement for this patient's hospitalization | pg/mL | Labs | 0 | | | | | labs |
| il12_mean | Interleukin 12 (Mean) | continuous | Average Interleukin 12 measurement for this patient's hospitalization | pg/mL | Labs | 0 | | | | | labs |
| il12_median | Interleukin 12 (Median) | continuous | Median Interleukin 12 measurement for this patient's hospitalization | pg/mL | Labs | 0 | | | | | labs |
| il12_min | Interleukin 12 (Min) | continuous | Minimum Interleukin 12 measurement for this patient's hospitalization | pg/mL | Labs | 0 | | | | | labs |
| il12_sd | Interleukin 12 (SD) | continuous | Standard deviation of the Interleukin 12 measurements during this patient's hospitalization | pg/mL | Labs | 0 | | | | | labs |
| il13_25 | Interleukin 13 (25th Percentile) | continuous | Interleukin 13 at the 25th percentile of this patient's measurements | pg/mL | Labs | 0 | | | | | labs |
| il13_75 | Interleukin 13 (75th Percentile) | continuous | Interleukin 13 at the 75th percentile of this patient's measurements | pg/mL | Labs | 0 | | | | | labs |
| il13_count | Interleukin 13 (Count) | continuous | Count of Interleukin 13 measurements for this patient | Number of measurements | Labs | 0 | | | | | labs |
| il13_first | Interleukin 13 (First) | continuous | First Interleukin 13 measurement for this patient's hospitalization | pg/mL | Labs | 0 | | | | | labs |
| il13_last | Interleukin 13 (Last) | continuous | Last Interleukin 13 measurement for this patient's hospitalization | pg/mL | Labs | 0 | | | | | labs |
| il13_max | Interleukin 13 (Max) | continuous | Maximum Interleukin 13 measurement for this patient's hospitalization | pg/mL | Labs | 0 | | | | | labs |
| il13_mean | Interleukin 13 (Mean) | continuous | Average Interleukin 13 measurement for this patient's hospitalization | pg/mL | Labs | 0 | | | | | labs |
| il13_median | Interleukin 13 (Median) | continuous | Median Interleukin 13 measurement for this patient's hospitalization | pg/mL | Labs | 0 | | | | | labs |
| il13_min | Interleukin 13 (Min) | continuous | Minimum Interleukin 13 measurement for this patient's hospitalization | pg/mL | Labs | 0 | | | | | labs |
| il13_sd | Interleukin 13 (SD) | continuous | Standard deviation of the Interleukin 13 measurements during this patient's hospitalization | pg/mL | Labs | 0 | | | | | labs |
| il17_25 | Interleukin 17 (25th Percentile) | continuous | Interleukin 17 at the 25th percentile of this patient's measurements | pg/mL | Labs | 0 | | | | | labs |
| il17_75 | Interleukin 17 (75th Percentile) | continuous | Interleukin 17 at the 75th percentile of this patient's measurements | pg/mL | Labs | 0 | | | | | labs |
| il17_count | Interleukin 17 (Count) | continuous | Count of Interleukin 17 measurements for this patient | Number of measurements | Labs | 0 | | | | | labs |
| il17_first | Interleukin 17 (First) | continuous | First Interleukin 17 measurement for this patient's hospitalization | pg/mL | Labs | 0 | | | | | labs |
| il17_last | Interleukin 17 (Last) | continuous | Last Interleukin 17 measurement for this patient's hospitalization | pg/mL | Labs | 0 | | | | | labs |
| il17_max | Interleukin 17 (Max) | continuous | Maximum Interleukin 17 measurement for this patient's hospitalization | pg/mL | Labs | 0 | | | | | labs |
| il17_mean | Interleukin 17 (Mean) | continuous | Average Interleukin 17 measurement for this patient's hospitalization | pg/mL | Labs | 0 | | | | | labs |
| il17_median | Interleukin 17 (Median) | continuous | Median Interleukin 17 measurement for this patient's hospitalization | pg/mL | Labs | 0 | | | | | labs |
| il17_min | Interleukin 17 (Min) | continuous | Minimum Interleukin 17 measurement for this patient's hospitalization | pg/mL | Labs | 0 | | | | | labs |
| il17_sd | Interleukin 17 (SD) | continuous | Standard deviation of the Interleukin 17 measurements during this patient's hospitalization | pg/mL | Labs | 0 | | | | | labs |
| il1beta_25 | Interleukin 1 Beta (25th Percentile) | continuous | Interleukin 1 Beta at the 25th percentile of this patient's measurements | kDa | Labs | 0 | | | | | labs |
| il1beta_75 | Interleukin 1 Beta (75th Percentile) | continuous | Interleukin 1 Beta at the 75th percentile of this patient's measurements | kDa | Labs | 0 | | | | | labs |
| il1beta_count | Interleukin 1 Beta (Count) | continuous | Count of Interleukin 1 Beta measurements for this patient | Number of measurements | Labs | 0 | | | | | labs |
| il1beta_first | Interleukin 1 Beta (First) | continuous | First Interleukin 1 Beta measurement for this patient's hospitalization | kDa | Labs | 0 | | | | | labs |
| il1beta_last | Interleukin 1 Beta (Last) | continuous | Last Interleukin 1 Beta measurement for this patient's hospitalization | kDa | Labs | 0 | | | | | labs |
| il1beta_max | Interleukin 1 Beta (Max) | continuous | Maximum Interleukin 1 Beta measurement for this patient's hospitalization | kDa | Labs | 0 | | | | | labs |
| il1beta_mean | Interleukin 1 Beta (Mean) | continuous | Average Interleukin 1 Beta measurement for this patient's hospitalization | kDa | Labs | 0 | | | | | labs |
| il1beta_median | Interleukin 1 Beta (Median) | continuous | Median Interleukin 1 Beta measurement for this patient's hospitalization | kDa | Labs | 0 | | | | | labs |
| il1beta_min | Interleukin 1 Beta (Min) | continuous | Minimum Interleukin 1 Beta measurement for this patient's hospitalization | kDa | Labs | 0 | | | | | labs |
| il1beta_sd | Interleukin 1 Beta (SD) | continuous | Standard deviation of the Interleukin 1 Beta measurements during this patient's hospitalization | kDa | Labs | 0 | | | | | labs |
| il2_25 | Interleukin 2 (25th Percentile) | continuous | Interleukin 2 at the 25th percentile of this patient's measurements | pg/mL | Labs | 0 | | | | | labs |
| il2_75 | Interleukin 2 (75th Percentile) | continuous | Interleukin 2 at the 75th percentile of this patient's measurements | pg/mL | Labs | 0 | | | | | labs |
| il2_count | Interleukin 2 (Count) | continuous | Count of Interleukin 2 measurements for this patient | Number of measurements | Labs | 0 | | | | | labs |
| il2_first | Interleukin 2 (First) | continuous | First Interleukin 2 measurement for this patient's hospitalization | pg/mL | Labs | 0 | | | | | labs |
| il2_last | Interleukin 2 (Last) | continuous | Last Interleukin 2 measurement for this patient's hospitalization | pg/mL | Labs | 0 | | | | | labs |
| il2_max | Interleukin 2 (Max) | continuous | Maximum Interleukin 2 measurement for this patient's hospitalization | pg/mL | Labs | 0 | | | | | labs |
| il2_mean | Interleukin 2 (Mean) | continuous | Average Interleukin 2 measurement for this patient's hospitalization | pg/mL | Labs | 0 | | | | | labs |
| il2_median | Interleukin 2 (Median) | continuous | Median Interleukin 2 measurement for this patient's hospitalization | pg/mL | Labs | 0 | | | | | labs |
| il2_min | Interleukin 2 (Min) | continuous | Minimum Interleukin 2 measurement for this patient's hospitalization | pg/mL | Labs | 0 | | | | | labs |
| il2_sd | Interleukin 2 (SD) | continuous | Standard deviation of the Interleukin 2 measurements during this patient's hospitalization | pg/mL | Labs | 0 | | | | | labs |
| il2r_25 | Interleukin 2 Soluble Receptor (25th Percentile) | continuous | Interleukin 2 Soluble Receptor at the 25th percentile of this patient's measurements | U/mL | Labs | 0 | | | | | labs |
| il2r_75 | Interleukin 2 Soluble Receptor (75th Percentile) | continuous | Interleukin 2 Soluble Receptor at the 75th percentile of this patient's measurements | U/mL | Labs | 0 | | | | | labs |
| il2r_count | Interleukin 2 Soluble Receptor (Count) | continuous | Count of Interleukin 2 Soluble Receptor measurements for this patient | Number of measurements | Labs | 0 | | | | | labs |
| il2r_first | Interleukin 2 Soluble Receptor (First) | continuous | First Interleukin 2 Soluble Receptor measurement for this patient's hospitalization | U/mL | Labs | 0 | | | | | labs |
| il2r_last | Interleukin 2 Soluble Receptor (Last) | continuous | Last Interleukin 2 Soluble Receptor measurement for this patient's hospitalization | U/mL | Labs | 0 | | | | | labs |
| il2r_max | Interleukin 2 Soluble Receptor (Max) | continuous | Maximum Interleukin 2 Soluble Receptor measurement for this patient's hospitalization | U/mL | Labs | 0 | | | | | labs |
| il2r_mean | Interleukin 2 Soluble Receptor (Mean) | continuous | Average Interleukin 2 Soluble Receptor measurement for this patient's hospitalization | U/mL | Labs | 0 | | | | | labs |
| il2r_median | Interleukin 2 Soluble Receptor (Median) | continuous | Median Interleukin 2 Soluble Receptor measurement for this patient's hospitalization | U/mL | Labs | 0 | | | | | labs |
| il2r_min | Interleukin 2 Soluble Receptor (Min) | continuous | Minimum Interleukin 2 Soluble Receptor measurement for this patient's hospitalization | U/mL | Labs | 0 | | | | | labs |
| il2r_sd | Interleukin 2 Soluble Receptor (SD) | continuous | Standard deviation of the Interleukin 2 Soluble Receptor measurements during this patient's hospitalizati | U/mL | Labs | 0 | | | | | labs |
| il4_25 | Interleukin 4 (25th Percentile) | continuous | Interleukin 4 at the 25th percentile of this patient's measurements | pg/mL | Labs | 0 | | | | | labs |
| il4_75 | Interleukin 4 (75th Percentile) | continuous | Interleukin 4 at the 75th percentile of this patient's measurements | pg/mL | Labs | 0 | | | | | labs |
| il4_count | Interleukin 4 (Count) | continuous | Count of Interleukin 4 measurements for this patient | Number of measurements | Labs | 0 | | | | | labs |
| il4_first | Interleukin 4 (First) | continuous | First Interleukin 4 measurement for this patient's hospitalization | pg/mL | Labs | 0 | | | | | labs |
| il4_last | Interleukin 4 (Last) | continuous | Last Interleukin 4 measurement for this patient's hospitalization | pg/mL | Labs | 0 | | | | | labs |
| il4_max | Interleukin 4 (Max) | continuous | Maximum Interleukin 4 measurement for this patient's hospitalization | pg/mL | Labs | 0 | | | | | labs |
| il4_mean | Interleukin 4 (Mean) | continuous | Average Interleukin 4 measurement for this patient's hospitalization | pg/mL | Labs | 0 | | | | | labs |
| il4_median | Interleukin 4 (Median) | continuous | Median Interleukin 4 measurement for this patient's hospitalization | pg/mL | Labs | 0 | | | | | labs |
| il4_min | Interleukin 4 (Min) | continuous | Minimum Interleukin 4 measurement for this patient's hospitalization | pg/mL | Labs | 0 | | | | | labs |
| il4_sd | Interleukin 4 (SD) | continuous | Standard deviation of the Interleukin 4 measurements during this patient's hospitalization | pg/mL | Labs | 0 | | | | | labs |
| il5_25 | Interleukin 5 (25th Percentile) | continuous | Interleukin 5 at the 25th percentile of this patient's measurements | pg/mL | Labs | 0 | | | | | labs |
| il5_75 | Interleukin 5 (75th Percentile) | continuous | Interleukin 5 at the 75th percentile of this patient's measurements | pg/mL | Labs | 0 | | | | | labs |
| il5_count | Interleukin 5 (Count) | continuous | Count of Interleukin 5 measurements for this patient | Number of measurements | Labs | 0 | | | | | labs |
| il5_first | Interleukin 5 (First) | continuous | First Interleukin 5 measurement for this patient's hospitalization | pg/mL | Labs | 0 | | | | | labs |
| il5_last | Interleukin 5 (Last) | continuous | Last Interleukin 5 measurement for this patient's hospitalization | pg/mL | Labs | 0 | | | | | labs |
| il5_max | Interleukin 5 (Max) | continuous | Maximum Interleukin 5 measurement for this patient's hospitalization | pg/mL | Labs | 0 | | | | | labs |
| il5_mean | Interleukin 5 (Mean) | continuous | Average Interleukin 5 measurement for this patient's hospitalization | pg/mL | Labs | 0 | | | | | labs |
| il5_median | Interleukin 5 (Median) | continuous | Median Interleukin 5 measurement for this patient's hospitalization | pg/mL | Labs | 0 | | | | | labs |
| il5_min | Interleukin 5 (Min) | continuous | Minimum Interleukin 5 measurement for this patient's hospitalization | pg/mL | Labs | 0 | | | | | labs |
| il5_sd | Interleukin 5 (SD) | continuous | Standard deviation of the Interleukin 5 measurements during this patient's hospitalization | pg/mL | Labs | 0 | | | | | labs |
| il6_25 | Interleukin 6 (25th Percentile) | continuous | Interleukin 6 at the 25th percentile of this patient's measurements | pg/mL | Labs | 0 | | | | | labs |
| il6_75 | Interleukin 6 (75th Percentile) | continuous | Interleukin 6 at the 75th percentile of this patient's measurements | pg/mL | Labs | 0 | | | | | labs |
| il6_count | Interleukin 6 (Count) | continuous | Count of Interleukin 6 measurements for this patient | Number of measurements | Labs | 0 | | | | | labs |
| il6_first | Interleukin 6 (First) | continuous | First Interleukin 6 measurement for this patient's hospitalization | pg/mL | Labs | 0 | | | | | labs |
| il6_last | Interleukin 6 (Last) | continuous | Last Interleukin 6 measurement for this patient's hospitalization | pg/mL | Labs | 0 | | | | | labs |
| il6_max | Interleukin 6 (Max) | continuous | Maximum Interleukin 6 measurement for this patient's hospitalization | pg/mL | Labs | 0 | | | | | labs |
| il6_mean | Interleukin 6 (Mean) | continuous | Average Interleukin 6 measurement for this patient's hospitalization | pg/mL | Labs | 0 | | | | | labs |
| il6_median | Interleukin 6 (Median) | continuous | Median Interleukin 6 measurement for this patient's hospitalization | pg/mL | Labs | 0 | | | | | labs |
| il6_min | Interleukin 6 (Min) | continuous | Minimum Interleukin 6 measurement for this patient's hospitalization | pg/mL | Labs | 0 | | | | | labs |
| il6_sd | Interleukin 6 (SD) | continuous | Standard deviation of the Interleukin 6 measurements during this patient's hospitalization | pg/mL | Labs | 0 | | | | | labs |
| il8_25 | Interleukin 8 (25th Percentile) | continuous | Interleukin 8 at the 25th percentile of this patient's measurements | pg/mL | Labs | 0 | | | | | labs |
| il8_75 | Interleukin 8 (75th Percentile) | continuous | Interleukin 8 at the 75th percentile of this patient's measurements | pg/mL | Labs | 0 | | | | | labs |
| il8_count | Interleukin 8 (Count) | continuous | Count of Interleukin 8 measurements for this patient | Number of measurements | Labs | 0 | | | | | labs |
| il8_first | Interleukin 8 (First) | continuous | First Interleukin 8 measurement for this patient's hospitalization | pg/mL | Labs | 0 | | | | | labs |
| il8_last | Interleukin 8 (Last) | continuous | Last Interleukin 8 measurement for this patient's hospitalization | pg/mL | Labs | 0 | | | | | labs |
| il8_max | Interleukin 8 (Max) | continuous | Maximum Interleukin 8 measurement for this patient's hospitalization | pg/mL | Labs | 0 | | | | | labs |
| il8_mean | Interleukin 8 (Mean) | continuous | Average Interleukin 8 measurement for this patient's hospitalization | pg/mL | Labs | 0 | | | | | labs |
| il8_median | Interleukin 8 (Median) | continuous | Median Interleukin 8 measurement for this patient's hospitalization | pg/mL | Labs | 0 | | | | | labs |
| il8_min | Interleukin 8 (Min) | continuous | Minimum Interleukin 8 measurement for this patient's hospitalization | pg/mL | Labs | 0 | | | | | labs |
| il8_sd | Interleukin 8 (SD) | continuous | Standard deviation of the Interleukin 8 measurements during this patient's hospitalization | pg/mL | Labs | 0 | | | | | labs |
| ilgamma_25 | Interferon Gamma (25th Percentile) | continuous | Interferon Gamma at the 25th percentile of this patient's measurements | pg/mL | Labs | 0 | | | | | labs |
| ilgamma_75 | Interferon Gamma (75th Percentile) | continuous | Interferon Gamma at the 75th percentile of this patient's measurements | pg/mL | Labs | 0 | | | | | labs |
| ilgamma_count | Interferon Gamma (Count) | continuous | Count of Interferon Gamma measurements for this patient | Number of measurements | Labs | 0 | | | | | labs |
| ilgamma_first | Interferon Gamma (First) | continuous | First Interferon Gamma measurement for this patient's hospitalization | pg/mL | Labs | 0 | | | | | labs |
| ilgamma_last | Interferon Gamma (Last) | continuous | Last Interferon Gamma measurement for this patient's hospitalization | pg/mL | Labs | 0 | | | | | labs |
| ilgamma_max | Interferon Gamma (Max) | continuous | Maximum Interferon Gamma measurement for this patient's hospitalization | pg/mL | Labs | 0 | | | | | labs |
| ilgamma_mean | Interferon Gamma (Mean) | continuous | Average Interferon Gamma measurement for this patient's hospitalization | pg/mL | Labs | 0 | | | | | labs |
| ilgamma_median | Interferon Gamma (Median) | continuous | Median Interferon Gamma measurement for this patient's hospitalization | pg/mL | Labs | 0 | | | | | labs |
| ilgamma_min | Interferon Gamma (Min) | continuous | Minimum Interferon Gamma measurement for this patient's hospitalization | pg/mL | Labs | 0 | | | | | labs |
| ilgamma_sd | Interferon Gamma (SD) | continuous | Standard deviation of the Interferon Gamma measurements during this patient's hospitalization | pg/mL | Labs | 0 | | | | | labs |
| immaturegranabs_25 | Immature Granulocytes Absolute (25th Percentile) | continuous | Immature Granulocytes Absolute at the 25th percentile of this patient's measurements | gm/dL | Labs | 0 | | | | | labs |
| immaturegranabs_75 | Immature Granulocytes Absolute (75th Percentile) | continuous | Immature Granulocytes Absolute at the 75th percentile of this patient's measurements | gm/dL | Labs | 0 | | | | | labs |
| immaturegranabs_coun | Immature Granulocytes Absolute (Count) | continuous | Count of Immature Granulocytes Absolute measurements for this patient | Number of measurements | Labs | 0 | | | | | labs |
| immaturegranabs_first | Immature Granulocytes Absolute (First) | continuous | First Immature Granulocytes Absolute measurement for this patient's hospitalization | gm/dL | Labs | 0 | | | | | labs |
| immaturegranabs_last | Immature Granulocytes Absolute (Last) | continuous | Last Immature Granulocytes Absolute measurement for this patient's hospitalization | gm/dL | Labs | 0 | | | | | labs |
| immaturegranabs_max | Immature Granulocytes Absolute (Max) | continuous | Maximum Immature Granulocytes Absolute measurement for this patient's hospitalization | gm/dL | Labs | 0 | | | | | labs |
| immaturegranabs_mear | Immature Granulocytes Absolute (Mean) | continuous | Average Immature Granulocytes Absolute measurement for this patient's hospitalization | gm/dL | Labs | 0 | | | | | labs |
| immaturegranabs_medi | Immature Granulocytes Absolute (Median) | continuous | Median Immature Granulocytes Absolute measurement for this patient's hospitalization | gm/dL | Labs | 0 | | | | | labs |
| immaturegranabs_min | Immature Granulocytes Absolute (Min) | continuous | Minimum Immature Granulocytes Absolute measurement for this patient's hospitalization | gm/dL | Labs | 0 | | | | | labs |
| immaturegranabs_sd | Immature Granulocytes Absolute (SD) | continuous | Standard deviation of the Immature Granulocytes Absolute measurements during this patient's hospitaliz | gm/dL | Labs | 0 | | | | | labs |
| immaturegranpercent_2 | Immature Granulocytes Percentage (25th Percentile) | continuous | Immature Granulocytes Percentage at the 25th percentile of this patient's measurements | % | Labs | 0 | | | | | labs |
| immaturegranpercent_7 | Immature Granulocytes Percentage (75th Percentile) | continuous | Immature Granulocytes Percentage at the 75th percentile of this patient's measurements | % | Labs | 0 | | | | | labs |
| immaturegranpercent_c | Immature Granulocytes Percentage (Count) | continuous | Count of Immature Granulocytes Percentage measurements for this patient | Number of measurements | Labs | 0 | | | | | labs |
| immaturegranpercent_fi | Immature Granulocytes Percentage (First) | continuous | First Immature Granulocytes Percentage measurement for this patient's hospitalization | % | Labs | 0 | | | | | labs |
| immaturegranpercent_la | Immature Granulocytes Percentage (Last) | continuous | Last Immature Granulocytes Percentage measurement for this patient's hospitalization | % | Labs | 0 | | | | | labs |

| | | | | | | | | | | | |
|---|---|---|---|---|---|---|---|---|---|---|---|
| immaturegranpercent_n | Immature Granulocytes Percentage (Max) | continuous | Maximum Immature Granulocytes Percentage measurement for this patient's hospitalization | % | Labs | 0 | | | | | labs |
| immaturegranpercent_n | Immature Granulocytes Percentage (Mean) | continuous | Average Immature Granulocytes Percentage measurement for this patient's hospitalization | % | Labs | 0 | | | | | labs |
| immaturegranpercent_n | Immature Granulocytes Percentage (Median) | continuous | Median Immature Granulocytes Percentage measurement for this patient's hospitalization | % | Labs | 0 | | | | | labs |
| immaturegranpercent_n | Immature Granulocytes Percentage (Min) | continuous | Minimum Immature Granulocytes Percentage measurement for this patient's hospitalization | % | Labs | 0 | | | | | labs |
| immaturegranpercent_s | Immature Granulocytes Percentage (SD) | continuous | Standard deviation of the Immature Granulocytes Percentage measurements during this patient's hospita | % | Labs | 0 | | | | | labs |
| imputed_baseline | Imputed Baseline | continuous | Imputed Baseline recorded for this patient | | Labs | 0 | | | | | aki_flag |
| imputed_baseline_creat | Imputed Baseline Creatinine | continuous | Imputed Baseline Creatinine recorded for this patient | | Labs | 0 | | | | | aki_flag |
| imputed_egfr | Imputed Estimated Glomerular Filtration Rate | continuous | Imputed Estimated Glomerular Filtrationn Rate recorded for this patient | | Labs | 0 | | | | | aki_flag |
| inr_25 | International Normalized Ratio (25th Percentile) | continuous | International Normalized Ratio at the 25th percentile of this patient's measurements | score | Labs | 0 | | | | | labs |
| inr_75 | International Normalized Ratio (75th Percentile) | continuous | International Normalized Ratio at the 75th percentile of this patient's measurements | score | Labs | 0 | | | | | labs |
| inr_count | International Normalized Ratio (Count) | continuous | Count of International Normalized Ratio measurements for this patient | Number of measurements | Labs | 0 | | | | | labs |
| inr_first | International Normalized Ratio (First) | continuous | First International Normalized Ratio measurement for this patient's hospitalization | score | Labs | 0 | | | | | labs |
| inr_last | International Normalized Ratio (Last) | continuous | Last International Normalized Ratio measurement for this patient's hospitalization | score | Labs | 0 | | | | | labs |
| inr_max | International Normalized Ratio (Max) | continuous | Maximum International Normalized Ratio measurement for this patient's hospitalization | score | Labs | 0 | | | | | labs |
| inr_mean | International Normalized Ratio (Mean) | continuous | Average International Normalized Ratio measurement for this patient's hospitalization | score | Labs | 0 | | | | | labs |
| inr_median | International Normalized Ratio (Median) | continuous | Median International Normalized Ratio measurement for this patient's hospitalization | score | Labs | 0 | | | | | labs |
| inr_min | International Normalized Ratio (Min) | continuous | Minimum International Normalized Ratio measurement for this patient's hospitalization | score | Labs | 0 | | | | | labs |
| inr_sd | International Normalized Ratio (SD) | continuous | Standard deviation of the International Normalized Ratio measurements during this patient's hospitalizati | score | Labs | 0 | | | | | labs |
| iron_25 | Iron (25th Percentile) | continuous | Iron at the 25th percentile of this patient's measurements | mcg/dL | Labs | 0 | | | | | labs |
| iron_75 | Iron (75th Percentile) | continuous | Iron at the 75th percentile of this patient's measurements | mcg/dL | Labs | 0 | | | | | labs |
| iron_count | Iron (Count) | continuous | Count of Iron measurements for this patient | Number of measurements | Labs | 0 | | | | | labs |
| iron_first | Iron (First) | continuous | First Iron measurement for this patient's hospitalization | mcg/dL | Labs | 0 | | | | | labs |
| iron_last | Iron (Last) | continuous | Last Iron measurement for this patient's hospitalization | mcg/dL | Labs | 0 | | | | | labs |
| iron_max | Iron (Max) | continuous | Maximum Iron measurement for this patient's hospitalization | mcg/dL | Labs | 0 | | | | | labs |
| iron_mean | Iron (Mean) | continuous | Average Iron measurement for this patient's hospitalization | mcg/dL | Labs | 0 | | | | | labs |
| iron_median | Iron (Median) | continuous | Median Iron measurement for this patient's hospitalization | mcg/dL | Labs | 0 | | | | | labs |
| iron_min | Iron (Min) | continuous | Minimum Iron measurement for this patient's hospitalization | mcg/dL | Labs | 0 | | | | | labs |
| iron_sd | Iron (SD) | continuous | Standard deviation of the Iron measurements during this patient's hospitalization | mcg/dL | Labs | 0 | | | | | labs |
| ironsat_25 | Iron Saturation (25th Percentile) | continuous | Iron Saturation at the 25th percentile of this patient's measurements | % | Labs | 0 | | | | | labs |
| ironsat_75 | Iron Saturation (75th Percentile) | continuous | Iron Saturation at the 75th percentile of this patient's measurements | % | Labs | 0 | | | | | labs |
| ironsat_count | Iron Saturation (Count) | continuous | Count of Iron Saturation measurements for this patient | Number of measurements | Labs | 0 | | | | | labs |
| ironsat_first | Iron Saturation (First) | continuous | First Iron Saturation measurement for this patient's hospitalization | % | Labs | 0 | | | | | labs |
| ironsat_last | Iron Saturation (Last) | continuous | Last Iron Saturation measurement for this patient's hospitalization | % | Labs | 0 | | | | | labs |
| ironsat_max | Iron Saturation (Max) | continuous | Maximum Iron Saturation measurement for this patient's hospitalization | % | Labs | 0 | | | | | labs |
| ironsat_mean | Iron Saturation (Mean) | continuous | Average Iron Saturation measurement for this patient's hospitalization | % | Labs | 0 | | | | | labs |
| ironsat_median | Iron Saturation (Median) | continuous | Median Iron Saturation measurement for this patient's hospitalization | % | Labs | 0 | | | | | labs |
| ironsat_min | Iron Saturation (Min) | continuous | Minimum Iron Saturation measurement for this patient's hospitalization | % | Labs | 0 | | | | | labs |
| ironsat_sd | Iron Saturation (SD) | continuous | Standard deviation of the Iron Saturation measurements during this patient's hospitalization | z | Labs | 0 | | | | | labs |
| kappafree_25 | Kappa Free (25th Percentile) | continuous | Kappa Free at the 25th percentile of this patient's measurements | mg/L | Labs | 0 | | | | | labs |
| kappafree_75 | Kappa Free (75th Percentile) | continuous | Kappa Free at the 75th percentile of this patient's measurements | mg/L | Labs | 0 | | | | | labs |
| kappafree_count | Kappa Free (Count) | continuous | Count of Kappa Free measurements for this patient | Number of measurements | Labs | 0 | | | | | labs |
| kappafree_first | Kappa Free (First) | continuous | First Kappa Free measurement for this patient's hospitalization | mg/L | Labs | 0 | | | | | labs |
| kappafree_last | Kappa Free (Last) | continuous | Last Kappa Free measurement for this patient's hospitalization | mg/L | Labs | 0 | | | | | labs |
| kappafree_max | Kappa Free (Max) | continuous | Maximum Kappa Free measurement for this patient's hospitalization | mg/L | Labs | 0 | | | | | labs |
| kappafree_mean | Kappa Free (Mean) | continuous | Average Kappa Free measurement for this patient's hospitalization | mg/L | Labs | 0 | | | | | labs |
| kappafree_median | Kappa Free (Median) | continuous | Median Kappa Free measurement for this patient's hospitalization | mg/L | Labs | 0 | | | | | labs |
| kappafree_min | Kappa Free (Min) | continuous | Minimum Kappa Free measurement for this patient's hospitalization | mg/L | Labs | 0 | | | | | labs |
| kappafree_sd | Kappa Free (SD) | continuous | Standard deviation of the Kappa Free measurements during this patient's hospitalization | mg/L | Labs | 0 | | | | | labs |
| klratio_25 | Potassium Iodide Ratio (25th Percentile) | continuous | Potassium Iodide Ratio at the 25th percentile of this patient's measurements | | Labs | 0 | | | | | labs |
| klratio_75 | Potassium Iodide Ratio (75th Percentile) | continuous | Potassium Iodide Ratio at the 75th percentile of this patient's measurements | | Labs | 0 | | | | | labs |
| klratio_count | Potassium Iodide Ratio (Count) | continuous | Count of Potassium Iodide Ratio measurements for this patient | Number of measurements | Labs | 0 | | | | | labs |
| klratio_first | Potassium Iodide Ratio (First) | continuous | First Potassium Iodide Ratio measurement for this patient's hospitalization | | Labs | 0 | | | | | labs |
| klratio_last | Potassium Iodide Ratio (Last) | continuous | Last Potassium Iodide Ratio measurement for this patient's hospitalization | | Labs | 0 | | | | | labs |
| klratio_max | Potassium Iodide Ratio (Max) | continuous | Maximum Potassium Iodide Ratio measurement for this patient's hospitalization | | Labs | 0 | | | | | labs |
| klratio_mean | Potassium Iodide Ratio (Mean) | continuous | Average Potassium Iodide Ratio measurement for this patient's hospitalization | | Labs | 0 | | | | | labs |
| klratio_median | Potassium Iodide Ratio (Median) | continuous | Median Potassium Iodide Ratio measurement for this patient's hospitalization | | Labs | 0 | | | | | labs |
| klratio_min | Potasssium Iodide Ratio (Minimum) | continuous | Minimum Potassium Iodide Ratio measurement for this patient's hospitalization | | Labs | 0 | | | | | |
| klratio_sd | Potassium Iodide Ratio (SD) | continuous | Standard deviation of the Potassium Iodide Ratio measurements during this patient's hospitalization | | Labs | 0 | | | | | labs |
| klratioserum_first | Potassium Iodide Ratio Serum (First) | categorical | First Potassium Iodide Ratio Serum measurement for this patient's hospitalization | | Labs | 0 | | | | | |
| klratioserum_last | Potassium Iodide Ratio Serum (Last) | categorical | Last Potassium Iodide Ratio Serum measurement for this patient's hospitalization | | Labs | 0 | | | | | |
| kur_25 | | | | | Labs | 0 | | | | | labs |
| kur_75 | | | | | Labs | 0 | | | | | labs |
| kur_count | | | | | Labs | 0 | | | | | labs |
| kur_first | | | | | Labs | 0 | | | | | labs |
| kur_last | | | | | Labs | 0 | | | | | labs |
| kur_max | | | | | Labs | 0 | | | | | labs |
| kur_mean | | | | | Labs | 0 | | | | | labs |
| kur_median | | | | | Labs | 0 | | | | | labs |
| kur_min | | | | | Labs | 0 | | | | | labs |
| kur_sd | | | | | Labs | 0 | | | | | labs |
| lactate_25 | Lactate (25th Percentile) | continuous | Lactate at the 25th percentile of this patient's measurements | mmol/L | Labs | 0 | | | | | labs |
| lactate_75 | Lactate (75th Percentile) | continuous | Lactate at the 75th percentile of this patient's measurements | mmol/L | Labs | 0 | | | | | labs |
| lactate_count | Lactate (Count) | continuous | Count of lactate measurements for this patient | Number of measurements | Labs | 0 | | | | | labs |
| lactate_first | Lactate (First) | continuous | First lactate measurement for this patient's hospitalization | mmol/L | Labs | 0 | | | | | labs |
| lactate_last | Lactate (Last) | continuous | Last lactate measurement for this patient's hospitalization | mmol/L | Labs | 0 | | | | | labs |
| lactate_max | Lactate (Max) | continuous | Maximum lactate measurement for this patient's hospitalization | mmol/L | Labs | 0 | | | | | labs |
| lactate_mean | Lactate (Mean) | continuous | Average lactate measurement for this patient's hospitalization | mmol/L | Labs | 0 | | | | | labs |
| lactate_median | Lactate (Median) | continuous | Median lactate measurement for this patient's hospitalization | mmol/L | Labs | 0 | | | | | labs |
| lactate_min | Lactate (Min) | continuous | Minimum lactate measurement for this patient's hospitalization | mmol/L | Labs | 0 | | | | | labs |
| lactate_sd | Lactate (SD) | continuous | Standard deviation of the lactate measurements during this patient's hospitalization | mmol/L | Labs | 0 | | | | | labs |
| lactatedehyd_25 | Lactate Dehydrogenase (25th Percentile) | continuous | Lactate Dehydrogenase at the 25th percentile of this patient's measurements | U/L | Labs | 0 | | | | | labs |
| lactatedehyd_75 | Lactate Dehydrogenase (75th Percentile) | continuous | Lactate Dehydrogenase at the 75th percentile of this patient's measurements | U/L | Labs | 0 | | | | | labs |
| lactatedehyd_count | Lactate Dehydrogenase (Count) | continuous | Count of Lactate Dehydrogenase measurements for this patient | Number of measurements | Labs | 0 | | | | | labs |
| lactatedehyd_first | Lactate Dehydrogenase (First) | continuous | First Lactate Dehydrogenase measurement for this patient's hospitalization | U/L | Labs | 0 | | | | | labs |
| lactatedehyd_last | Lactate Dehydrogenase (Last) | continuous | Last Lactate Dehydrogenase measurement for this patient's hospitalization | U/L | Labs | 0 | | | | | labs |
| lactatedehyd_max | Lactate Dehydrogenase (Max) | continuous | Maximum Lactate Dehydrogenase measurement for this patient's hospitalization | U/L | Labs | 0 | | | | | labs |
| lactatedehyd_mean | Lactate Dehydrogenase (Mean) | continuous | Average Lactate Dehydrogenase measurement for this patient's hospitalization | U/L | Labs | 0 | | | | | labs |
| lactatedehyd_median | Lactate Dehydrogenase (Median) | continuous | Median Lactate Dehydrogenase measurement for this patient's hospitalization | U/L | Labs | 0 | | | | | labs |
| lactatedehyd_min | Lactate Dehydrogenase (Min) | continuous | Minimum Lactate Dehydrogenase measurement for this patient's hospitalization | U/L | Labs | 0 | | | | | labs |
| lactatedehyd_sd | Lactate Dehydrogenase (SD) | continuous | Standard deviation of the Lactate Dehydrogenase measurements during this patient's hospitalization | U/L | Labs | 0 | | | | | labs |
| lambdafree_25 | Lambda Free Light Chains (25th Percentile) | continuous | Lambda Free Light Chains at the 25th percentile of this patient's measurements | mg/L | Labs | 0 | | | | | labs |
| lambdafree_75 | Lambda Free Light Chains (75th Percentile) | continuous | Lambda Free Light Chains at the 75th percentile of this patient's measurements | mg/L | Labs | 0 | | | | | labs |
| lambdafree_count | Lambda Free Light Chains (Count) | continuous | Count of Lambda Free Light Chains measurements for this patient | Number of measurements | Labs | 0 | | | | | labs |
| lambdafree_first | Lambda Free Light Chains (First) | continuous | First Lambda Free Light Chains measurement for this patient's hospitalization | mg/L | Labs | 0 | | | | | labs |
| lambdafree_last | Lambda Free Light Chains (Last) | continuous | Last Lambda Free Light Chains measurement for this patient's hospitalization | mg/L | Labs | 0 | | | | | labs |
| lambdafree_max | Lambda Free Light Chains (Max) | continuous | Maximum Lambda Free Light Chains measurement for this patient's hospitalization | mg/L | Labs | 0 | | | | | labs |
| lambdafree_mean | Lambda Free Light Chains (Mean) | continuous | Average Lambda Free Light Chains measurement for this patient's hospitalization | mg/L | Labs | 0 | | | | | labs |
| lambdafree_median | Lambda Free Light Chains (Median) | continuous | Median Lambda Free Light Chains measurement for this patient's hospitalization | mg/L | Labs | 0 | | | | | labs |
| lambdafree_min | Lambda Free Light Chains (Min) | continuous | Minimum Lambda Free Light Chains measurement for this patient's hospitalization | mg/L | Labs | 0 | | | | | labs |
| lambdafree_sd | Lambda Free Light Chains (SD) | continuous | Standard deviation of the Lambda Free Light Chains measurements during this patient's hospitalization | mg/L | Labs | 0 | | | | | labs |
| leadblood_25 | Lead Blood (25th Percentile) | continuous | Lead Blood at the 25th percentile of this patient's measurements | μg/dL | Labs | 0 | | | | | labs |
| leadblood_75 | Lead Blood (75th Percentile) | continuous | Lead Blood at the 75th percentile of this patient's measurements | μg/dL | Labs | 0 | | | | | labs |
| leadblood_count | Lead Blood (Count) | continuous | Count of Lead Blood measurements for this patient | Number of measurements | Labs | 0 | | | | | labs |
| leadblood_first | Lead Blood (First) | continuous | First Lead Blood measurement for this patient's hospitalization | μg/dL | Labs | 0 | | | | | labs |
| leadblood_last | Lead Blood (Last) | continuous | Last Lead Blood measurement for this patient's hospitalization | μg/dL | Labs | 0 | | | | | labs |
| leadblood_max | Lead Blood (Max) | continuous | Maximum Lead Blood measurement for this patient's hospitalization | μg/dL | Labs | 0 | | | | | labs |
| leadblood_mean | Lead Blood (Mean) | continuous | Average Lead Blood measurement for this patient's hospitalization | μg/dL | Labs | 0 | | | | | labs |
| leadblood_median | Lead Blood (Median) | continuous | Median Lead Blood measurement for this patient's hospitalization | μg/dL | Labs | 0 | | | | | labs |
| leadblood_min | Lead Blood (Min) | continuous | Minimum Lead Blood measurement for this patient's hospitalization | μg/dL | Labs | 0 | | | | | labs |
| leadblood_sd | Lead Blood (SD) | continuous | Standard deviation of the Lead Blood measurements during this patient's hospitalization | μg/dL | Labs | 0 | | | | | labs |
| legionella_first | Legionella (First) | categorical | Results of the first Legionella test during this patient's hospitalization | | Labs | 0 | | | | | labs |
| legionella_last | Legionella (Last) | categorical | Results of the last Legionella test during this patient's hospitalization | | Labs | 0 | | | | | labs |
| lipase_25 | Lipase (25th Percentile) | continuous | Lipase at the 25th percentile of this patient's measurements | U/L | Labs | 0 | | | | | labs |
| lipase_75 | Lipase (75th Percentile) | continuous | Lipase at the 75th percentile of this patient's measurements | U/L | Labs | 0 | | | | | labs |
| lipase_count | Lipase (Count) | continuous | Count of Lipase measurements for this patient | Number of measurements | Labs | 0 | | | | | labs |
| lipase_first | Lipase (First) | continuous | First Lipase measurement for this patient's hospitalization | U/L | Labs | 0 | | | | | labs |
| lipase_last | Lipase (Last) | continuous | Last Lipase measurement for this patient's hospitalization | U/L | Labs | 0 | | | | | labs |
| lipase_max | Lipase (Max) | continuous | Maximum Lipase measurement for this patient's hospitalization | U/L | Labs | 0 | | | | | labs |
| lipase_mean | Lipase (Mean) | continuous | Average Lipase measurement for this patient's hospitalization | U/L | Labs | 0 | | | | | labs |
| lipase_median | Lipase (Median) | continuous | Median Lipase measurement for this patient's hospitalization | U/L | Labs | 0 | | | | | labs |
| lipase_min | Lipase (Min) | continuous | Minimum Lipase measurement for this patient's hospitalization | U/L | Labs | 0 | | | | | labs |
| lipase_sd | Lipase (SD) | continuous | Standard deviation of the Lipase measurements during this patient's hospitalization | U/L | Labs | 0 | | | | | labs |
| lithiumlevel_25 | Lithium Level (25th Percentile) | continuous | Lithium Level at the 25th percentile of this patient's measurements | mmol/L | Labs | 0 | | | | | labs |
| lithiumlevel_75 | Lithium Level (75th Percentile) | continuous | Lithium Level at the 75th percentile of this patient's measurements | mmol/L | Labs | 0 | | | | | labs |

| | | | | | | | | | | | |
|---|---|---|---|---|---|---|---|---|---|---|---|
| **lithiumlevel_count** | Lithium Level (Count) | continuous | Count of Lithium Level measurements for this patient | Number of measurements | Labs | 0 | | | | | labs |
| **lithiumlevel_first** | Lithium Level (First) | continuous | First Lithium Level measurement for this patient's hospitalization | mmol/L | Labs | 0 | | | | | labs |
| **lithiumlevel_last** | Lithium Level (Last) | continuous | Last Lithium Level measurement for this patient's hospitalization | mmol/L | Labs | 0 | | | | | labs |
| **lithiumlevel_max** | Lithium Level (Max) | continuous | Maximum Lithium Level measurement for this patient's hospitalization | mmol/L | Labs | 0 | | | | | labs |
| **lithiumlevel_mean** | Lithium Level (Mean) | continuous | Average Lithium Level measurement for this patient's hospitalization | mmol/L | Labs | 0 | | | | | labs |
| **lithiumlevel_median** | Lithium Level (Median) | continuous | Median Lithium Level measurement for this patient's hospitalization | mmol/L | Labs | 0 | | | | | labs |
| **lithiumlevel_min** | Lithium Level (Min) | continuous | Minimum Lithium Level measurement for this patient's hospitalization | mmol/L | Labs | 0 | | | | | labs |
| **lithiumlevel_sd** | Lithium Level (SD) | continuous | Standard deviation of the Lithium Level measurements during this patient's hospitalization | mmol/L | Labs | 0 | | | | | labs |
| **lymeab_first** | Lyme Antibody (First) | categorical | Results of the first Lyme Antibody test during this patient's hospitalization | | Labs | 0 | | | | | labs |
| **lymeab_last** | Lyme Antibody (Last) | categorical | Results of the last Lyme Antibody test during this patient's hospitalization | | Labs | 0 | | | | | labs |
| **lymewestern_first** | Lyme Western Blot (First) | categorical | Results of the first Lyme Western Blot during this patient's hospitalization | | Labs | 0 | | | | | labs |
| **lymewestern_last** | Lyme Western Blot (Last) | categorical | Results of the last Lyme Western Blot during this patient's hospitalization | | Labs | 0 | | | | | labs |
| **lymphocyte_abs_25** | Lymphocyte Absolute (25th Percentile) | continuous | Lymphocyte Absolute at the 25th percentile of this patient's measurements | µL | Labs | 0 | | | | | labs |
| **lymphocyte_abs_75** | Lymphocyte Absolute (75th Percentile) | continuous | Lymphocyte Absolute at the 75th percentile of this patient's measurements | µL | Labs | 0 | | | | | labs |
| **lymphocyte_abs_count** | Lymphocyte Absolute (Count) | continuous | Count of Lymphocyte Absolute measurements for this patient | Number of measurements | Labs | 0 | | | | | labs |
| **lymphocyte_abs_first** | Lymphocyte Absolute (First) | continuous | First Lymphocyte Absolute measurement for this patient's hospitalization | µL | Labs | 0 | | | | | labs |
| **lymphocyte_abs_last** | Lymphocyte Absolute (Last) | continuous | Last Lymphocyte Absolute measurement for this patient's hospitalization | µL | Labs | 0 | | | | | labs |
| **lymphocyte_abs_max** | Lymphocyte Absolute (Max) | continuous | Maximum Lymphocyte Absolute measurement for this patient's hospitalization | µL | Labs | 0 | | | | | labs |
| **lymphocyte_abs_mean** | Lymphocyte Absolute (Mean) | continuous | Average Lymphocyte Absolute measurement for this patient's hospitalization | µL | Labs | 0 | | | | | labs |
| **lymphocyte_abs_mediar** | Lymphocyte Absolute (Median) | continuous | Median Lymphocyte Absolute measurement for this patient's hospitalization | µL | Labs | 0 | | | | | labs |
| **lymphocyte_abs_min** | Lymphocyte Absolute (Min) | continuous | Minimum Lymphocyte Absolute measurement for this patient's hospitalization | µL | Labs | 0 | | | | | labs |
| **lymphocyte_abs_sd** | Lymphocyte Absolute (SD) | continuous | Standard deviation of the Lymphocyte Absolute measurements during this patient's hospitalization | µL | Labs | 0 | | | | | labs |
| **lymphocyte_percent_25** | Lymphocyte Percentage (25th Percentile) | continuous | Lymphocyte Percentage at the 25th percentile of this patient's measurements | % | Labs | 0 | | | | | labs |
| **lymphocyte_percent_75** | Lymphocyte Percentage (75th Percentile) | continuous | Lymphocyte Percentage at the 75th percentile of this patient's measurements | % | Labs | 0 | | | | | labs |
| **lymphocyte_percent_co** | Lymphocyte Percentage (Count) | continuous | Count of Lymphocyte Percentage measurements for this patient | Number of measurements | Labs | 0 | | | | | labs |
| **lymphocyte_percent_firs** | Lymphocyte Percentage (First) | continuous | First Lymphocyte Percentage measurement for this patient's hospitalization | % | Labs | 0 | | | | | labs |
| **lymphocyte_percent_las** | Lymphocyte Percentage (Last) | continuous | Last Lymphocyte Percentage measurement for this patient's hospitalization | % | Labs | 0 | | | | | labs |
| **lymphocyte_percent_ma** | Lymphocyte Percentage (Max) | continuous | Maximum Lymphocyte Percentage measurement for this patient's hospitalization | % | Labs | 0 | | | | | labs |
| **lymphocyte_percent_me** | Lymphocyte Percentage (Mean) | continuous | Average Lymphocyte Percentage measurement for this patient's hospitalization | % | Labs | 0 | | | | | labs |
| **lymphocyte_percent_me** | Lymphocyte Percentage (Median) | continuous | Median Lymphocyte Percentage measurement for this patient's hospitalization | % | Labs | 0 | | | | | labs |
| **lymphocyte_percent_mi** | Lymphocyte Percentage (Min) | continuous | Minimum Lymphocyte Percentage measurement for this patient's hospitalization | % | Labs | 0 | | | | | labs |
| **lymphocyte_percent_sd** | Lymphocyte Percentage (SD) | continuous | Standard deviation of the Lymphocyte Percentage measurements during this patient's hospitalization | % | Labs | 0 | | | | | labs |
| **magnesium_25** | Magnesium (25th Percentile) | continuous | Magnesium at the 25th percentile of this patient's measurements | mg/dL | Labs | 0 | | | | | labs |
| **magnesium_75** | Magnesium (75th Percentile) | continuous | Magnesium at the 75th percentile of this patient's measurements | mg/dL | Labs | 0 | | | | | labs |
| **magnesium_count** | Magnesium (Count) | continuous | Count of Magnesium measurements for this patient | Number of measurements | Labs | 0 | | | | | labs |
| **magnesium_first** | Magnesium (First) | continuous | First Magnesium measurement for this patient's hospitalization | mg/dL | Labs | 0 | | | | | labs |
| **magnesium_last** | Magnesium (Last) | continuous | Last Magnesium measurement for this patient's hospitalization | mg/dL | Labs | 0 | | | | | labs |
| **magnesium_max** | Magnesium (Max) | continuous | Maximum Magnesium measurement for this patient's hospitalization | mg/dL | Labs | 0 | | | | | labs |
| **magnesium_mean** | Magnesium (Mean) | continuous | Average Magnesium measurement for this patient's hospitalization | mg/dL | Labs | 0 | | | | | labs |
| **magnesium_median** | Magnesium (Median) | continuous | Median Magnesium measurement for this patient's hospitalization | mg/dL | Labs | 0 | | | | | labs |
| **magnesium_min** | Magnesium (Min) | continuous | Minimum Magnesium measurement for this patient's hospitalization | mg/dL | Labs | 0 | | | | | labs |
| **magnesium_sd** | Magnesium (SD) | continuous | Standard deviation of the Magnesium measurements during this patient's hospitalization | mg/dL | Labs | 0 | | | | | labs |
| **mcg_25** | | continuous | | | Labs | 0 | | | | | labs |
| **mcg_75** | | continuous | | | Labs | 0 | | | | | labs |
| **mcg_count** | | continuous | | | Labs | 0 | | | | | labs |
| **mcg_first** | | continuous | | | Labs | 0 | | | | | labs |
| **mcg_last** | | continuous | | | Labs | 0 | | | | | labs |
| **mcg_max** | | continuous | | | Labs | 0 | | | | | labs |
| **mcg_mean** | | continuous | | | Labs | 0 | | | | | labs |
| **mcg_median** | | continuous | | | Labs | 0 | | | | | labs |
| **mcg_min** | | continuous | | | Labs | 0 | | | | | labs |
| **mcg_sd** | | continuous | | | Labs | 0 | | | | | labs |
| **mch_25** | Mean Corpuscular Hemoglobin (25th Percentile) | continuous | Mean Corpuscular Hemoglobin at the 25th percentile of this patient's measurements | pg/cell | Labs | 0 | | | | | labs |
| **mch_75** | Mean Corpuscular Hemoglobin (75th Percentile) | continuous | Mean Corpuscular Hemoglobin at the 75th percentile of this patient's measurements | pg/cell | Labs | 0 | | | | | labs |
| **mch_count** | Mean Corpuscular Hemoglobin (Count) | continuous | Count of Mean Corpuscular Hemoglobin measurements for this patient | Number of measurements | Labs | 0 | | | | | labs |
| **mch_first** | Mean Corpuscular Hemoglobin (First) | continuous | First Mean Corpuscular Hemoglobin measurement for this patient's hospitalization | pg/cell | Labs | 0 | | | | | labs |
| **mch_last** | Mean Corpuscular Hemoglobin (Last) | continuous | Last Mean Corpuscular Hemoglobin measurement for this patient's hospitalization | pg/cell | Labs | 0 | | | | | labs |
| **mch_max** | Mean Corpuscular Hemoglobin (Max) | continuous | Maximum Mean Corpuscular Hemoglobin measurement for this patient's hospitalization | pg/cell | Labs | 0 | | | | | labs |
| **mch_mean** | Mean Corpuscular Hemoglobin (Mean) | continuous | Average Mean Corpuscular Hemoglobin measurement for this patient's hospitalization | pg/cell | Labs | 0 | | | | | labs |
| **mch_median** | Mean Corpuscular Hemoglobin (Median) | continuous | Median Mean Corpuscular Hemoglobin measurement for this patient's hospitalization | pg/cell | Labs | 0 | | | | | labs |
| **mch_min** | Mean Corpuscular Hemoglobin (Min) | continuous | Minimum Mean Corpuscular Hemoglobin measurement for this patient's hospitalization | pg/cell | Labs | 0 | | | | | labs |
| **mch_sd** | Mean Corpuscular Hemoglobin (SD) | continuous | Standard deviation of the Mean Corpuscular Hemoglobin measurements during this patient's hospitaliza | pg/cell | Labs | 0 | | | | | labs |
| **mchc_25** | Mean Corpuscular Hemoglobin Concentration (25th Perce | continuous | Mean Corpuscular Hemoglobin Concentration at the 25th percentile of this patient's measurements | g/dL | Labs | 0 | | | | | labs |
| **mchc_75** | Mean Corpuscular Hemoglobin Concentration (75th Perce | continuous | Mean Corpuscular Hemoglobin Concentration at the 75th percentile of this patient's measurements | g/dL | Labs | 0 | | | | | labs |
| **mchc_count** | Mean Corpuscular Hemoglobin Concentration (Count) | continuous | Count of Mean Corpuscular Hemoglobin Concentration measurements for this patient | Number of measurements | Labs | 0 | | | | | labs |
| **mchc_first** | Mean Corpuscular Hemoglobin Concentration (First) | continuous | First Mean Corpuscular Hemoglobin Concentration measurement for this patient's hospitalization | g/dL | Labs | 0 | | | | | labs |
| **mchc_last** | Mean Corpuscular Hemoglobin Concentration (Last) | continuous | Last Mean Corpuscular Hemoglobin Concentration measurement for this patient's hospitalization | g/dL | Labs | 0 | | | | | labs |
| **mchc_max** | Mean Corpuscular Hemoglobin Concentration (Max) | continuous | Maximum Mean Corpuscular Hemoglobin Concentration measurement for this patient's hospitalization | g/dL | Labs | 0 | | | | | labs |
| **mchc_mean** | Mean Corpuscular Hemoglobin Concentration (Mean) | continuous | Average Mean Corpuscular Hemoglobin Concentration measurement for this patient's hospitalization | g/dL | Labs | 0 | | | | | labs |
| **mchc_median** | Mean Corpuscular Hemoglobin Concentration (Median) | continuous | Median Mean Corpuscular Hemoglobin Concentration measurement for this patient's hospitalization | g/dL | Labs | 0 | | | | | labs |
| **mchc_min** | Mean Corpuscular Hemoglobin Concentration (Min) | continuous | Minimum Mean Corpuscular Hemoglobin Concentration measurement for this patient's hospitalization | g/dL | Labs | 0 | | | | | labs |
| **mchc_sd** | Mean Corpuscular Hemoglobin Concentration (SD) | continuous | Standard deviation of the Mean Corpuscular Hemoglobin Concentration measurements during this patie | g/dL | Labs | 0 | | | | | labs |
| **mcv_25** | Mean Corposcular Volume (25th Percentile) | continuous | Mean Corposcular Volume at the 25th percentile of this patient's measurements | fL. | Labs | 0 | | | | | labs |
| **mcv_75** | Mean Corposcular Volume (75th Percentile) | continuous | Mean Corposcular Volume at the 75th percentile of this patient's measurements | fL. | Labs | 0 | | | | | labs |
| **mcv_count** | Mean Corposcular Volume (Count) | continuous | Count of Mean Corposcular Volume measurements for this patient | Number of measurements | Labs | 0 | | | | | labs |
| **mcv_first** | Mean Corposcular Volume (First) | continuous | First Mean Corposcular Volume measurement for this patient's hospitalization | fL. | Labs | 0 | | | | | labs |
| **mcv_last** | Mean Corposcular Volume (Last) | continuous | Last Mean Corposcular Volume measurement for this patient's hospitalization | fL. | Labs | 0 | | | | | labs |
| **mcv_max** | Mean Corposcular Volume (Max) | continuous | Maximum Mean Corposcular Volume measurement for this patient's hospitalization | fL. | Labs | 0 | | | | | labs |
| **mcv_mean** | Mean Corposcular Volume (Mean) | continuous | Average Mean Corposcular Volume measurement for this patient's hospitalization | fL. | Labs | 0 | | | | | labs |
| **mcv_median** | Mean Corposcular Volume (Median) | continuous | Median Mean Corposcular Volume measurement for this patient's hospitalization | fL. | Labs | 0 | | | | | labs |
| **mcv_min** | Mean Corposcular Volume (Min) | continuous | Minimum Mean Corposcular Volume measurement for this patient's hospitalization | fL. | Labs | 0 | | | | | labs |
| **mcv_sd** | Mean Corposcular Volume (SD) | continuous | Standard deviation of the Mean Corposcular Volume measurements during this patient's hospitalization | fL. | Labs | 0 | | | | | labs |
| **methanolblood_first** | Methanol Blood (First) | categorical | Results of the first Methanol Blood test during this patient's hospitalization | | Labs | 0 | | | | | labs |
| **methanolblood_last** | Methanol BLood (Last) | categorical | Results of the last Methanol Blood test during this patient's hospitalization | | Labs | 0 | | | | | labs |
| **methgb_25** | | continuous | | | Labs | 0 | | | | | labs |
| **methgb_75** | | continuous | | | Labs | 0 | | | | | labs |
| **methgb_count** | | continuous | | | Labs | 0 | | | | | labs |
| **methgb_first** | | continuous | | | Labs | 0 | | | | | labs |
| **methgb_last** | | continuous | | | Labs | 0 | | | | | labs |
| **methgb_max** | | continuous | | | Labs | 0 | | | | | labs |
| **methgb_mean** | | continuous | | | Labs | 0 | | | | | labs |
| **methgb_median** | | continuous | | | Labs | 0 | | | | | labs |
| **methgb_min** | | continuous | | | Labs | 0 | | | | | labs |
| **methgb_sd** | | continuous | | | Labs | 0 | | | | | labs |
| **mma_25** | Methylmalonic Acidemia (25th Percentile) | continuous | Methylmalonic Acidemia at the 25th percentile of this patient's measurements | ?mol/mmol | Labs | 0 | | | | | labs |
| **mma_75** | Methylmalonic Acidemia (75th Percentile) | continuous | Methylmalonic Acidemia at the 75th percentile of this patient's measurements | ?mol/mmol | Labs | 0 | | | | | labs |
| **mma_count** | Methylmalonic Acidemia (Count) | continuous | Count of Methylmalonic Acidemia measurements for this patient | Number of measurements | Labs | 0 | | | | | labs |
| **mma_first** | Methylmalonic Acidemia (First) | continuous | First Methylmalonic Acidemia measurement for this patient's hospitalization | ?mol/mmol | Labs | 0 | | | | | labs |
| **mma_last** | Methylmalonic Acidemia (Last) | continuous | Last Methylmalonic Acidemia measurement for this patient's hospitalization | ?mol/mmol | Labs | 0 | | | | | labs |
| **mma_max** | Methylmalonic Acidemia (Max) | continuous | Maximum Methylmalonic Acidemia measurement for this patient's hospitalization | ?mol/mmol | Labs | 0 | | | | | labs |
| **mma_mean** | Methylmalonic Acidemia (Mean) | continuous | Average Methylmalonic Acidemia measurement for this patient's hospitalization | ?mol/mmol | Labs | 0 | | | | | labs |
| **mma_median** | Methylmalonic Acidemia (Median) | continuous | Median Methylmalonic Acidemia measurement for this patient's hospitalization | ?mol/mmol | Labs | 0 | | | | | labs |
| **mma_min** | Methylmalonic Acidemia (Min) | continuous | Minimum Methylmalonic Acidemia measurement for this patient's hospitalization | ?mol/mmol | Labs | 0 | | | | | labs |
| **mma_sd** | Methylmalonic Acidemia (SD) | continuous | Standard deviation of the Methylmalonic Acidemia measurements during this patient's hospitalization | ?mol/mmol | Labs | 0 | | | | | labs |
| **monocyte_abs_25** | Monocyte Absolute (25th Percentile) | continuous | Monocyte Absolute at the 25th percentile of this patient's measurements | % | Labs | 0 | | | | | labs |
| **monocyte_abs_75** | Monocyte Absolute (75th Percentile) | continuous | Monocyte Absolute at the 75th percentile of this patient's measurements | % | Labs | 0 | | | | | labs |
| **monocyte_abs_count** | Monocyte Absolute (Count) | continuous | Count of Monocyte Absolute measurements for this patient | Number of measurements | Labs | 0 | | | | | labs |
| **monocyte_abs_first** | Monocyte Absolute (First) | continuous | First Monocyte Absolute measurement for this patient's hospitalization | % | Labs | 0 | | | | | labs |
| **monocyte_abs_last** | Monocyte Absolute (Last) | continuous | Last Monocyte Absolute measurement for this patient's hospitalization | % | Labs | 0 | | | | | labs |
| **monocyte_abs_max** | Monocyte Absolute (Max) | continuous | Maximum Monocyte Absolute measurement for this patient's hospitalization | % | Labs | 0 | | | | | labs |
| **monocyte_abs_mean** | Monocyte Absolute (Mean) | continuous | Average Monocyte Absolute measurement for this patient's hospitalization | % | Labs | 0 | | | | | labs |
| **monocyte_abs_median** | Monocyte Absolute (Median) | continuous | Median Monocyte Absolute measurement for this patient's hospitalization | % | Labs | 0 | | | | | labs |
| **monocyte_abs_min** | Monocyte Absolute (Min) | continuous | Minimum Monocyte Absolute measurement for this patient's hospitalization | % | Labs | 0 | | | | | labs |
| **monocyte_abs_sd** | Monocyte Absolute (SD) | continuous | Standard deviation of the Monocyte Absolute measurements during this patient's hospitalization | % | Labs | 0 | | | | | labs |
| **monocyte_percent_25** | Monocyte Percentage (25th Percentile) | continuous | Monocyte Percentage at the 25th percentile of this patient's measurements | % | Labs | 0 | | | | | labs |
| **monocyte_percent_75** | Monocyte Percentage (75th Percentile) | continuous | Monocyte Percentage at the 75th percentile of this patient's measurements | % | Labs | 0 | | | | | labs |
| **monocyte_percent_cour** | Monocyte Percentage (Count) | continuous | Count of Monocyte Percentage measurements for this patient | Number of measurements | Labs | 0 | | | | | labs |
| **monocyte_percent_first** | Monocyte Percentage (First) | continuous | First Monocyte Percentage measurement for this patient's hospitalization | % | Labs | 0 | | | | | labs |
| **monocyte_percent_last** | Monocyte Percentage (Last) | continuous | Last Monocyte Percentage measurement for this patient's hospitalization | % | Labs | 0 | | | | | labs |
| **monocyte_percent_max** | Monocyte Percentage (Max) | continuous | Maximum Monocyte Percentage measurement for this patient's hospitalization | % | Labs | 0 | | | | | labs |
| **monocyte_percent_mea** | Monocyte Percentage (Mean) | continuous | Average Monocyte Percentage measurement for this patient's hospitalization | % | Labs | 0 | | | | | labs |
| **monocyte_percent_med** | Monocyte Percentage (Median) | continuous | Median Monocyte Percentage measurement for this patient's hospitalization | % | Labs | 0 | | | | | labs |
| **monocyte_percent_min** | Monocyte Percentage (Min) | continuous | Minimum Monocyte Percentage measurement for this patient's hospitalization | % | Labs | 0 | | | | | labs |
| **monocyte_percent_sd** | Monocyte Percentage (SD) | continuous | Standard deviation of the Monocyte Percentage measurements during this patient's hospitalization | % | Labs | 0 | | | | | labs |

| | | | | | | | | | | | |
|---|---|---|---|---|---|---|---|---|---|---|---|
| **mpv_25** | Mean Platelet Volume (25th Percentile) | continuous | Mean Platelet Volume at the 25th percentile of this patient's measurements | fL. | Labs | 0 | | | | | labs |
| **mpv_75** | Mean Platelet Volume (75th Percentile) | continuous | Mean Platelet Volume at the 75th percentile of this patient's measurements | fL. | Labs | 0 | | | | | labs |
| **mpv_count** | Mean Platelet Volume (Count) | continuous | Count of Mean Platelet Volume measurements for this patient | Number of measurements | Labs | 0 | | | | | labs |
| **mpv_first** | Mean Platelet Volume (First) | continuous | First Mean Platelet Volume measurement for this patient's hospitalization | fL. | Labs | 0 | | | | | labs |
| **mpv_last** | Mean Platelet Volume (Last) | continuous | Last Mean Platelet Volume measurement for this patient's hospitalization | fL. | Labs | 0 | | | | | labs |
| **mpv_max** | Mean Platelet Volume (Max) | continuous | Maximum Mean Platelet Volume measurement for this patient's hospitalization | fL. | Labs | 0 | | | | | labs |
| **mpv_mean** | Mean Platelet Volume (Mean) | continuous | Average Mean Platelet Volume measurement for this patient's hospitalization | fL. | Labs | 0 | | | | | labs |
| **mpv_median** | Mean Platelet Volume (Median) | continuous | Median Mean Platelet Volume measurement for this patient's hospitalization | fL. | Labs | 0 | | | | | labs |
| **mpv_min** | Mean Platelet Volume (Min) | continuous | Minimum Mean Platelet Volume measurement for this patient's hospitalization | fL. | Labs | 0 | | | | | labs |
| **mpv_sd** | Mean Platelet Volume (SD) | continuous | Standard deviation of the Mean Platelet Volume measurements during this patient's hospitalization | fL. | Labs | 0 | | | | | labs |
| **myoglobin_25** | Myoglobin (25th Percentile) | continuous | Myoglobin at the 25th percentile of this patient's measurements | ng/mL | Labs | 0 | | | | | labs |
| **myoglobin_75** | Myoglobin (75th Percentile) | continuous | Myoglobin at the 75th percentile of this patient's measurements | ng/mL | Labs | 0 | | | | | labs |
| **myoglobin_count** | Myoglobin (Count) | continuous | Count of Myoglobin measurements for this patient | Number of measurements | Labs | 0 | | | | | labs |
| **myoglobin_first** | Myoglobin (First) | continuous | First Myoglobin measurement for this patient's hospitalization | ng/mL | Labs | 0 | | | | | labs |
| **myoglobin_last** | Myoglobin (Last) | continuous | Last Myoglobin measurement for this patient's hospitalization | ng/mL | Labs | 0 | | | | | labs |
| **myoglobin_max** | Myoglobin (Max) | continuous | Maximum Myoglobin measurement for this patient's hospitalization | ng/mL | Labs | 0 | | | | | labs |
| **myoglobin_mean** | Myoglobin (Mean) | continuous | Average Myoglobin measurement for this patient's hospitalization | ng/mL | Labs | 0 | | | | | labs |
| **myoglobin_median** | Myoglobin (Median) | continuous | Median Myoglobin measurement for this patient's hospitalization | ng/mL | Labs | 0 | | | | | labs |
| **myoglobin_min** | Myoglobin (Min) | continuous | Minimum Myoglobin measurement for this patient's hospitalization | ng/mL | Labs | 0 | | | | | labs |
| **myoglobin_sd** | Myoglobin (SD) | continuous | Standard deviation of the Myoglobin measurements during this patient's hospitalization | ng/mL | Labs | 0 | | | | | labs |
| **myourine_first** | Myoglobin Urine (First) | categorical | Results of the first myoglobin in urine test during this patient's hospitalization | | Labs | 0 | | | | | labs |
| **myourine_last** | Myoglobin Urine (Last) | categorical | Results of the last myoglobin in urine test during this patient's hospitalization | | Labs | 0 | | | | | labs |
| **neutrophil_abs_25** | Absolute Neutrophil (25th Percentile) | continuous | Absolute neutrophil count at the 25th percentile of this patient's measurements | | Labs | 0 | | | | | labs |
| **neutrophil_abs_75** | Absolute Neutrophil (75th Percentile) | continuous | Absolute neutrophil count at the 75th percentile of this patient's measurements | | Labs | 0 | | | | | labs |
| **neutrophil_abs_count** | Absolute Neutrophil (Count) | continuous | Count of absolute neutrophil measurements for this patient | Number of measurements | Labs | 0 | | | | | labs |
| **neutrophil_abs_first** | Absolute Neutrophil (First) | continuous | First absolute neutrophil measurement for this patient's hospitalization | | Labs | 0 | | | | | labs |
| **neutrophil_abs_last** | Absolute Neutrophil (Last) | continuous | Last absolute neutrophil measurement for this patient's hospitalization | | Labs | 0 | | | | | labs |
| **neutrophil_abs_max** | Absolute Neutrophil (Max) | continuous | Maximum absolute neutrophil count measurement for this patient's hospitalization | | Labs | 0 | | | | | labs |
| **neutrophil_abs_mean** | Absolute Neutrophil (Mean) | continuous | Average absolute neutrophil count measurement for this patient's hospitalization | | Labs | 0 | | | | | labs |
| **neutrophil_abs_median** | Absolute Neutrophil (Median) | continuous | Median absolute neutrophil count measurement for this patient's hospitalization | | Labs | 0 | | | | | labs |
| **neutrophil_abs_min** | Absolute Neutrophil (Min) | continuous | Minimum absolute neutrophil count measurement for this patient's hospitalization | | Labs | 0 | | | | | labs |
| **neutrophil_abs_sd** | Absolute Neutrophil (SD) | continuous | Standard deviation of the absolute neutrophil count measurements during this patient's hospitalization | | Labs | 0 | | | | | labs |
| **neutrophil_percent_25** | Neutrophil Percentage (25th Percentile) | continuous | Neutrophil percentage at the 25th percentile of this patient's measurements | % | Labs | 0 | | | | | labs |
| **neutrophil_percent_75** | Neutrophil Percentage (75th Percentile) | continuous | Neutrophil percentage at the 75th percentile of this patient's measurements | % | Labs | 0 | | | | | labs |
| **neutrophil_percent_cou** | Neutrophil Percentage (Count) | continuous | Count of neutrophil percentage measurements for this patient | Number of measurements | Labs | 0 | | | | | labs |
| **neutrophil_percent_first** | Neutrophil Percentage (First) | continuous | First neutrophil percentage measurement for this patient's hospitalization | % | Labs | 0 | | | | | labs |
| **neutrophil_percent_last** | Neutrophil Percentage (Last) | continuous | Last neutrophil percentage measurement for this patient's hospitalization | % | Labs | 0 | | | | | labs |
| **neutrophil_percent_max** | Neutrophil Percentage (Max) | continuous | Maximum neutrophil percentage measurement for this patient's hospitalization | % | Labs | 0 | | | | | labs |
| **neutrophil_percent_mea** | Neutrophil Percentage (Mean) | continuous | Average neutrophil percentage measurement for this patient's hospitalization | % | Labs | 0 | | | | | labs |
| **neutrophil_percent_med** | Neutrophil Percentage (Median) | continuous | Median neutrophil percentage measurement for this patient's hospitalization | % | Labs | 0 | | | | | labs |
| **neutrophil_percent_min** | Neutrophil Percentage (Min) | continuous | Minimum neutrophil percentage measurement for this patient's hospitalization | % | Labs | 0 | | | | | labs |
| **neutrophil_percent_sd** | Neutrophil Percentage (SD) | continuous | Standard deviation of the neutrophil percentage measurements during this patient's hospitalization | % | Labs | 0 | | | | | labs |
| **norovirus_first** | Norovirus (First) | categorical | Results of the first norovirus test during this patient's hospitalization | | Labs | 0 | | | | | labs |
| **norovirus_last** | Norovirus (Last) | categorical | Results of the last norovirus test during this patient's hospitalization | | Labs | 0 | | | | | labs |
| **nrbc_25** | Nucleated Red Blood Cells (25th Percentile) | continuous | Nucleated red blood cells at the 25th percentile of this patient's measurements | NRBCs | Labs | 0 | | | | | labs |
| **nrbc_75** | Nucleated Red Blood Cells (75th Percentile) | continuous | Nucleated red blood cells at the 75th percentile of this patient's measurements | NRBCs | Labs | 0 | | | | | labs |
| **nrbc_count** | Nucleated Red Blood Cells (Count) | continuous | Count of nucleated red blood cell measurements for this patient | Number of measurements | Labs | 0 | | | | | labs |
| **nrbc_first** | Nucleated Red Blood Cells (First) | continuous | First nucleated red blood cell measurement for this patient's hospitalization | NRBCs | Labs | 0 | | | | | labs |
| **nrbc_last** | Nucleated Red Blood Cells (Last) | continuous | Last nucleated red blood cell measurement for this patient's hospitalization | NRBCs | Labs | 0 | | | | | labs |
| **nrbc_max** | Nucleated Red Blood Cells (Max) | continuous | Maximum nucleated red blood cell measurement for this patient's hospitalization | NRBCs | Labs | 0 | | | | | labs |
| **nrbc_mean** | Nucleated Red Blood Cells (Mean) | continuous | Average nucleated red blood cell measurement for this patient's hospitalization | NRBCs | Labs | 0 | | | | | labs |
| **nrbc_median** | Nucleated Red Blood Cells (Median) | continuous | Median nucleated red blood cell measurement for this patient's hospitalization | NRBCs | Labs | 0 | | | | | labs |
| **nrbc_min** | Nucleated Red Blood Cells (Min) | continuous | Minimum nucleated red blood cell measurement for this patient's hospitalization | NRBCs | Labs | 0 | | | | | labs |
| **nrbc_sd** | Nucleated Red Blood Cells (SD) | continuous | Standard deviation of the nucleated red blood cell measurements during this patient's hospitalization | NRBCs | Labs | 0 | | | | | labs |
| **nrbcdiff_25** | Nucleated Red Blood Cells Difference (25th Percentile) | continuous | Nucleated Red Blood Cells Difference at the 25th percentile of this patient's measurements | NRBCs | Labs | 0 | | | | | labs |
| **nrbcdiff_75** | Nucleated Red Blood Cells Difference (75th Percentile) | continuous | Nucleated Red Blood Cells Difference at the 75th percentile of this patient's measurements | NRBCs | Labs | 0 | | | | | labs |
| **nrbcdiff_count** | Nucleated Red Blood Cells Difference (Count) | continuous | Count of Nucleated Red Blood Cells Difference measurements for this patient | Number of measurements | Labs | 0 | | | | | labs |
| **nrbcdiff_first** | Nucleated Red Blood Cells Difference (First) | continuous | First Nucleated Red Blood Cells Difference measurement for this patient's hospitalization | NRBCs | Labs | 0 | | | | | labs |
| **nrbcdiff_last** | Nucleated Red Blood Cells Difference (Last) | continuous | Last Nucleated Red Blood Cells Difference measurement for this patient's hospitalization | NRBCs | Labs | 0 | | | | | labs |
| **nrbcdiff_max** | Nucleated Red Blood Cells Difference (Max) | continuous | Maximum Nucleated Red Blood Cells Difference measurement for this patient's hospitalization | NRBCs | Labs | 0 | | | | | labs |
| **nrbcdiff_mean** | Nucleated Red Blood Cells Difference (Mean) | continuous | Average Nucleated Red Blood Cells Difference measurement for this patient's hospitalization | NRBCs | Labs | 0 | | | | | labs |
| **nrbcdiff_median** | Nucleated Red Blood Cells Difference (Median) | continuous | Median Nucleated Red Blood Cells Difference measurement for this patient's hospitalization | NRBCs | Labs | 0 | | | | | labs |
| **nrbcdiff_min** | Nucleated Red Blood Cells Difference (Min) | continuous | Minimum Nucleated Red Blood Cells Difference measurement for this patient's hospitalization | NRBCs | Labs | 0 | | | | | labs |
| **nrbcdiff_sd** | Nucleated Red Blood Cells Difference (SD) | continuous | Standard deviation of the Nucleated Red Blood Cells Difference measurements during this patient's hosp | NRBCs | Labs | 0 | | | | | labs |
| **nucleatedrbcpercent_25** | Nucleated Red Blood Cells Percentage (25th Percentile) | continuous | Nucleated RBCs percentage at the 25th percentile of this patient's measurements | % | Labs | 0 | | | | | labs |
| **nucleatedrbcpercent_75** | Nucleated Red Blood Cells Percentage (75th Percentile) | continuous | Nucleated RBCs percentage at the 75th percentile of this patient's measurements | % | Labs | 0 | | | | | labs |
| **nucleatedrbcpercent_co** | Nucleated Red Blood Cells Percentage (Count) | continuous | Count of nucleated RBCs percentage measurements for this patient | Number of measurements | Labs | 0 | | | | | labs |
| **nucleatedrbcpercent_fir** | Nucleated Red Blood Cells Percentage (First) | continuous | First nucleated RBCs percentage measurement for this patient's hospitalization | % | Labs | 0 | | | | | labs |
| **nucleatedrbcpercent_la** | Nucleated Red Blood Cells Percentage (Last) | continuous | Last nucleated RBCs percentage measurement for this patient's hospitalization | % | Labs | 0 | | | | | labs |
| **nucleatedrbcpercent_m** | Nucleated Red Blood Cells Percentage (Max) | continuous | Maximum nucleated RBCs percentage measurement for this patient's hospitalization | % | Labs | 0 | | | | | labs |
| **nucleatedrbcpercent_m** | Nucleated Red Blood Cells Percentage (Mean) | continuous | Average nucleated RBCs percentage measurement for this patient's hospitalization | % | Labs | 0 | | | | | labs |
| **nucleatedrbcpercent_m** | Nucleated Red Blood Cells Percentage (Median) | continuous | Median nucleated RBCs percentage measurement for this patient's hospitalization | % | Labs | 0 | | | | | labs |
| **nucleatedrbcpercent_m** | Nucleated Red Blood Cells Percentage (Min) | continuous | Minimum nucleated RBCs percentage measurement for this patient's hospitalization | % | Labs | 0 | | | | | labs |
| **nucleatedrbcpercent_sd** | Nucleated Red Blood Cells Percentage (SD) | continuous | Standard deviation of the nucleated RBCs percentage measurements during this patient's hospitalizatior | % | Labs | 0 | | | | | labs |
| **o2sat_25** | Oxygen Saturation (25th Percentile) | continuous | Oxygen saturation level at the 25th percentile of this patient's measurements | mmHg | Labs | 0 | | | | | labs |
| **o2sat_75** | Oxygen Saturation (75th Percentile) | continuous | Oxygen saturation level at the 75th percentile of this patient's measurements | mmHg | Labs | 0 | | | | | labs |
| **o2sat_count** | Oxygen Saturation (Count) | continuous | Count of oxygen saturation measurements for this patient | Number of measurements | Labs | 0 | | | | | labs |
| **o2sat_first** | Oxygen Saturation (First) | continuous | First oxygen saturation measurement for this patient's hospitalization | mmHg | Labs | 0 | | | | | labs |
| **o2sat_last** | Oxygen Saturation (Last) | continuous | Last oxygen saturation measurement for this patient's hospitalization | mmHg | Labs | 0 | | | | | labs |
| **o2sat_max** | Oxygen Saturation (Max) | continuous | Maximum oxygen saturation level measurement for this patient's hospitalization | mmHg | Labs | 0 | | | | | labs |
| **o2sat_mean** | Oxygen Saturation (Mean) | continuous | Average oxygen saturation level measurement for this patient's hospitalization | mmHg | Labs | 0 | | | | | labs |
| **o2sat_median** | Oxygen Saturation (Median) | continuous | Median oxygen saturation level measurement for this patient's hospitalization | mmHg | Labs | 0 | | | | | labs |
| **o2sat_min** | Oxygen Saturation (Min) | continuous | Minimum oxygen saturation level measurement for this patient's hospitalization | mmHg | Labs | 0 | | | | | labs |
| **o2sat_sd** | Oxygen Saturation (SD) | continuous | Standard deviation of the oxygen saturation level measurements during this patient's hospitalization | mmHg | Labs | 0 | | | | | labs |
| **o2satart_25** | Arterial Oxygen Saturation (25th Percentile) | continuous | Arterial oxygen saturation level at the 25th percentile of this patient's measurements | mmHg | Labs | 0 | | | | | labs |
| **o2satart_75** | Arterial Oxygen Saturation (75th Percentile) | continuous | Arterial oxygen saturation level at the 75th percentile of this patient's measurements | mmHg | Labs | 0 | | | | | labs |
| **o2satart_count** | Arterial Oxygen Saturation (Count) | continuous | Count of arterial oxygen saturation measurements for this patient | Number of measurements | Labs | 0 | | | | | labs |
| **o2satart_first** | Arterial Oxygen Saturation (First) | continuous | First arterial oxygen saturation measurement for this patient's hospitalization | mmHg | Labs | 0 | | | | | labs |
| **o2satart_last** | Arterial Oxygen Saturation (Last) | continuous | Last arterial oxygen saturation measurement for this patient's hospitalization | mmHg | Labs | 0 | | | | | labs |
| **o2satart_max** | Arterial Oxygen Saturation (Max) | continuous | Maximum arterial oxygen saturation level measurement for this patient's hospitalization | mmHg | Labs | 0 | | | | | labs |
| **o2satart_mean** | Arterial Oxygen Saturation (Mean) | continuous | Average arterial oxygen saturation level measurement for this patient's hospitalization | mmHg | Labs | 0 | | | | | labs |
| **o2satart_median** | Arterial Oxygen Saturation (Median) | continuous | Median arterial oxygen saturation level measurement for this patient's hospitalization | mmHg | Labs | 0 | | | | | labs |
| **o2satart_min** | Arterial Oxygen Saturation (Min) | continuous | Minimum arterial oxygen saturation level measurement for this patient's hospitalization | mmHg | Labs | 0 | | | | | labs |
| **o2satart_sd** | Arterial Oxygen Saturation (SD) | continuous | Standard deviation of the arterial oxygen saturation level measurements during this patient's hospitalizati | mmHg | Labs | 0 | | | | | labs |
| **o2satmixedvenous_first** | Mixed Venous Oxygen Saturation (First) | categorical | Results of the first opiate urine test during this patient's hospitalization | | Labs | 0 | | | | | |
| **o2satmixedvenous_last** | Mixed Venous Oxygen Saturation (Last) | categorical | Results of the last opiate urine test during this patient's hospitalization | | Labs | 0 | | | | | |
| **o2satvenous_first** | Venous Oxygen Saturation (First) | categorical | Results of the first opiate urine test during this patient's hospitalization | | Labs | 0 | | | | | |
| **o2satvenous_last** | Venous Oxygen Saturation (Last) | categorical | Results of the last opiate urine test during this patient's hospitalization | | Labs | 0 | | | | | |
| **opiateurine_first** | Opiate Urine (First) | categorical | Results of the first opiate urine test during this patient's hospitalization | | Labs | 0 | | | | | labs |
| **opiateurine_last** | Opiate Urine (Last) | categorical | Results of the last opiate urine test during this patient's hospitalization | | Labs | 0 | | | | | labs |
| **osmolality_25** | Osmolality (25th Percentile) | continuous | Osmolality level at the 25th percentile of this patient's measurements | mOsm/kg | Labs | 0 | | | | | labs |
| **osmolality_75** | Osmolality (75th Percentile) | continuous | Osmolality level at the 75th percentile of this patient's measurements | mOsm/kg | Labs | 0 | | | | | labs |
| **osmolality_count** | Osmolality (Count) | continuous | Count of osmolality level measurements for this patient | Number of measurements | Labs | 0 | | | | | labs |
| **osmolality_first** | Osmolality (First) | continuous | First osmolality level measurement for this patient's hospitalization | mOsm/kg | Labs | 0 | | | | | labs |
| **osmolality_last** | Osmolality (Last) | continuous | Last osmolality level measurement for this patient's hospitalization | mOsm/kg | Labs | 0 | | | | | labs |
| **osmolality_max** | Osmolality (Max) | continuous | Maximum osmolality level measurement for this patient's hospitalization | mOsm/kg | Labs | 0 | | | | | labs |
| **osmolality_mean** | Osmolality (Mean) | continuous | Average osmolality level measurement for this patient's hospitalization | mOsm/kg | Labs | 0 | | | | | labs |
| **osmolality_median** | Osmolality (Median) | continuous | Median osmolality level measurement for this patient's hospitalization | mOsm/kg | Labs | 0 | | | | | labs |
| **osmolality_min** | Osmolality (Min) | continuous | Minimum osmolality level measurement for this patient's hospitalization | mOsm/kg | Labs | 0 | | | | | labs |
| **osmolality_sd** | Osmolality (SD) | continuous | Standard deviation of the osmolality level measurements during this patient's hospitalization | mOsm/kg | Labs | 0 | | | | | labs |
| **osmolurine_25** | Urine Osmolality (25th Percentile) | continuous | Urine osmolality level at the 25th percentile of this patient's measurements | mOsm/kg | Labs | 0 | | | | | labs |
| **osmolurine_75** | Urine Osmolality (75th Percentile) | continuous | Urine osmolality level at the 75th percentile of this patient's measurements | mOsm/kg | Labs | 0 | | | | | labs |
| **osmolurine_count** | Urine Osmolality (Count) | continuous | Count of urine osmolality level measurements for this patient | Number of measurements | Labs | 0 | | | | | labs |
| **osmolurine_first** | Urine Osmolality (First) | continuous | First urine osmolality level measurement for this patient's hospitalization | mOsm/kg | Labs | 0 | | | | | labs |
| **osmolurine_last** | Urine Osmolality (Last) | continuous | Last urine osmolality level measurement for this patient's hospitalization | mOsm/kg | Labs | 0 | | | | | labs |
| **osmolurine_max** | Urine Osmolality (Max) | continuous | Maximum urine osmolality level measurement for this patient's hospitalization | mOsm/kg | Labs | 0 | | | | | labs |
| **osmolurine_mean** | Urine Osmolality (Mean) | continuous | Average urine osmolality level measurement for this patient's hospitalization | mOsm/kg | Labs | 0 | | | | | labs |
| **osmolurine_median** | Urine Osmolality (Median) | continuous | Median urine osmolality level measurement for this patient's hospitalization | mOsm/kg | Labs | 0 | | | | | labs |
| **osmolurine_min** | Urine Osmolality (Min) | continuous | Minimum urine osmolality level measurement for this patient's hospitalization | mOsm/kg | Labs | 0 | | | | | labs |
| **osmolurine_sd** | Urine Osmolality (SD) | continuous | Standard deviation of the urine osmolality level measurements during this patient's hospitalization | mOsm/kg | Labs | 0 | | | | | labs |
| **othercorona_first** | Other Corona Test (First) | categorical | Results of the first other corona test during this patient's hospitalization | | Labs | 0 | | | | | |
| **othercorona_last** | Other Corona Test (Last) | categorical | Results of the last other corona test during this patient's hospitalization | | Labs | 0 | | | | | |
| **parainfluenza_first** | Parainfluenza (First) | categorical | Results of the first parainfluenza test during this patient's hospitalization | | Labs | 0 | | | | | labs |
| **parainfluenza_last** | Parainfluenza (Last) | categorical | Results of the last parainfluenza test during this patient's hospitalization | | Labs | 0 | | | | | labs |

| | | | | | | | | | | | |
|---|---|---|---|---|---|---|---|---|---|---|---|
| **pco2_25** | Partial Pressure of Carbon Dioxide (25th Percentile) | continuous | Partial pressure of carbon dioxide at the 25th percentile of this patient's measurements | mmHg | Labs | 0 | | | | | labs |
| **pco2_75** | Partial Pressure of Carbon Dioxide (75th Percentile) | continuous | Partial pressure of carbon dioxide at the 75th percentile of this patient's measurements | mmHg | Labs | 0 | | | | | labs |
| **pco2_count** | Partial Pressure of Carbon Dioxide (Count) | continuous | Count of partial pressure (CO2) measurements for this patient | Number of measurements | Labs | 0 | | | | | labs |
| **pco2_first** | Partial Pressure of Carbon Dioxide (First) | continuous | First partial pressure (CO2) measurement for this patient's hospitalization | mmHg | Labs | 0 | | | | | labs |
| **pco2_last** | Partial Pressure of Carbon Dioxide (Last) | continuous | Last partial pressure (CO2) measurement for this patient's hospitalization | mmHg | Labs | 0 | | | | | labs |
| **pco2_max** | Partial Pressure of Carbon Dioxide (Max) | continuous | Maximum partial pressure (CO2) measurement for this patient's hospitalization | mmHg | Labs | 0 | | | | | labs |
| **pco2_mean** | Partial Pressure of Carbon Dioxide (Mean) | continuous | Average partial pressure (CO2) measurement for this patient's hospitalization | mmHg | Labs | 0 | | | | | labs |
| **pco2_median** | Partial Pressure of Carbon Dioxide (Median) | continuous | Median partial pressure (CO2) measurement for this patient's hospitalization | mmHg | Labs | 0 | | | | | labs |
| **pco2_min** | Partial Pressure of Carbon Dioxide (Min) | continuous | Minimum partial pressure (CO2) measurement for this patient's hospitalization | mmHg | Labs | 0 | | | | | labs |
| **pco2_sd** | Partial Pressure of Carbon Dioxide (SD) | continuous | Standard deviation of the partial pressure of carbon dioxide measurements during this patient's hospitali | mmHg | Labs | 0 | | | | | labs |
| **pcratio_25** | Protein/Creatinine Ratio (25th Percentile) | continuous | Protein/Creatinine ratio at the 25th percentile of this patient's measurements | mg/mg | Labs | 0 | | | | | labs |
| **pcratio_75** | Protein/Creatinine Ratio (75th Percentile) | continuous | Protein/Creatinine ratio at the 75th percentile of this patient's measurements | mg/mg | Labs | 0 | | | | | labs |
| **pcratio_count** | Protein/Creatinine Ratio (Count) | continuous | Count of protein/creatinine ratio measurements for this patient | Number of measurements | Labs | 0 | | | | | labs |
| **pcratio_first** | Protein/Creatinine Ratio (First) | continuous | First protein/creatinine ratio measurement for this patient's hospitalization | mg/mg | Labs | 0 | | | | | labs |
| **pcratio_last** | Protein/Creatinine Ratio (Last) | continuous | Last protein/creatinine ratio measurement for this patient's hospitalization | mg/mg | Labs | 0 | | | | | labs |
| **pcratio_max** | Protein/Creatinine Ratio (Max) | continuous | Maximum protein/creatinine ratio measurement for this patient's hospitalization | mg/mg | Labs | 0 | | | | | labs |
| **pcratio_mean** | Protein/Creatinine Ratio (Mean) | continuous | Average protein/creatinine ratio measurement for this patient's hospitalization | mg/mg | Labs | 0 | | | | | labs |
| **pcratio_median** | Protein/Creatinine Ratio (Median) | continuous | Median protein/creatinine ratio measurement for this patient's hospitalization | mg/mg | Labs | 0 | | | | | labs |
| **pcratio_min** | Protein/Creatinine Ratio (Min) | continuous | Minimum protein/creatinine ratio measurement for this patient's hospitalization | mg/mg | Labs | 0 | | | | | labs |
| **pcratio_sd** | Protein/Creatinine Ratio (SD) | continuous | Standard deviation of the protein/creatinine ratio measurements during this patient's hospitalization | mg/mg | Labs | 0 | | | | | labs |
| **peep_25** | Positive End-Expiratory Pressure (25th Percentile) | continuous | Positive end-expiratory pressure at the 25th percentile of this patient's measurements | cm H2O | Labs | 0 | | | | | labs |
| **peep_75** | Positive End-Expiratory Pressure (75th Percentile) | continuous | Positive end-expiratory pressure at the 75th percentile of this patient's measurements | cm/H2O | Labs | 0 | | | | | labs |
| **peep_count** | Positive End-Expiratory Pressure (Count) | continuous | Count of positive end-expiratory pressure measurements for this patient | Number of measurements | Labs | 0 | | | | | labs |
| **peep_first** | Positive End-Expiratory Pressure (First) | continuous | First positive end-expiratory pressure measurement for this patient's hospitalization | cm H2O | Labs | 0 | | | | | labs |
| **peep_last** | Positive End-Expiratory Pressure (Last) | continuous | Last positive end-expiratory pressure measurement for this patient's hospitalization | cm H2O | Labs | 0 | | | | | labs |
| **peep_max** | Positive End-Expiratory Pressure (Max) | continuous | Maximum positive end-expiratory pressure measurement for this patient's hospitalization | cm H2O | Labs | 0 | | | | | labs |
| **peep_mean** | Positive End-Expiratory Pressure (Mean) | continuous | Average positive end-expiratory pressure measurement for this patient's hospitalization | cm H2O | Labs | 0 | | | | | labs |
| **peep_median** | Positive End-Expiratory Pressure (Median) | continuous | Median positive end-expiratory pressure measurement for this patient's hospitalization | cm H2O | Labs | 0 | | | | | labs |
| **peep_min** | Positive End-Expiratory Pressure (Min) | continuous | Minimum positive end-expiratory pressure measurement for this patient's hospitalization | cm H2O | Labs | 0 | | | | | labs |
| **peep_sd** | Positive End-Expiratory Pressure (SD) | continuous | Standard deviation of the positive end-expiratory pressure measurements during this patient's hospitaliz | cm H2O | Labs | 0 | | | | | labs |
| **ph_25** | pH (25th Percentile) | continuous | pH level at the 25th percentile of this patient's measurements | | Labs | 0 | | | | | labs |
| **ph_75** | pH (75th Percentile) | continuous | pH level at the 75th percentile of this patient's measurements | | Labs | 0 | | | | | labs |
| **ph_count** | pH (Count) | continuous | Count of pH measurements for this patient | Number of measurements | Labs | 0 | | | | | labs |
| **ph_first** | pH (First) | continuous | First pH measurement for this patient's hospitalization | | Labs | 0 | | | | | labs |
| **ph_last** | pH (Last) | continuous | Last pH measurement for this patient's hospitalization | | Labs | 0 | | | | | labs |
| **ph_max** | pH (Max) | continuous | Maximum pH measurement for this patient's hospitalization | | Labs | 0 | | | | | labs |
| **ph_mean** | pH (Mean) | continuous | Average pH measurement for this patient's hospitalization | | Labs | 0 | | | | | labs |
| **ph_median** | pH (Median) | continuous | Median pH measurement for this patient's hospitalization | | Labs | 0 | | | | | labs |
| **ph_min** | pH (Min) | continuous | Minimum pH measurement for this patient's hospitalization | | Labs | 0 | | | | | labs |
| **ph_sd** | pH (SD) | continuous | Standard deviation of the pH measurements during this patient's hospitalization | | Labs | 0 | | | | | labs |
| **pharterial_25** | Arterial Blood Gas (25th Percentile) | continuous | Arterial blood gas level at the 25th percentile of this patient's measurements | | Labs | 0 | | | | | labs |
| **pharterial_75** | Arterial Blood Gas (75th Percentile) | continuous | Arterial blood gas level at the 75th percentile of this patient's measurements | | Labs | 0 | | | | | labs |
| **pharterial_count** | Arterial Blood Gas (Count) | continuous | Count of arterial blood gas measurements for this patient | Number of measurements | Labs | 0 | | | | | labs |
| **pharterial_first** | Arterial Blood Gas (First) | continuous | First arterial blood gas level measurement for this patient's hospitalization | | Labs | 0 | | | | | labs |
| **pharterial_last** | Arterial Blood Gas (Last) | continuous | Last arterial blood gas level measurement for this patient's hospitalization | | Labs | 0 | | | | | labs |
| **pharterial_max** | Arterial Blood Gas (Max) | continuous | Maximum arterial blood gas level measurement for this patient's hospitalization | | Labs | 0 | | | | | labs |
| **pharterial_mean** | Arterial Blood Gas (Mean) | continuous | Average arterial blood gas level measurement for this patient's hospitalization | | Labs | 0 | | | | | labs |
| **pharterial_median** | Arterial Blood Gas (Median) | continuous | Median arterial blood gas level measurement for this patient's hospitalization | | Labs | 0 | | | | | labs |
| **pharterial_min** | Arterial Blood Gas (Min) | continuous | Minimum arterial blood gas level measurement for this patient's hospitalization | | Labs | 0 | | | | | labs |
| **pharterial_sd** | Arterial Blood Gas (SD) | continuous | Standard deviation of the arterial blood gas level measurements during this patient's hospitalization | | Labs | 0 | | | | | labs |
| **phmv_25** | Physalis Mottle Virus (25th Percentile) | continuous | Physalis Mottle Virus at the 25th percentile of this patient's measurements | | Labs | 0 | | | | | labs |
| **phmv_75** | Physalis Mottle Virus (75th Percentile) | continuous | Physalis Mottle Virus at the 75th percentile of this patient's measurements | | Labs | 0 | | | | | labs |
| **phmv_count** | Physalis Mottle Virus (Count) | continuous | Count of Physalis Mottle Virus measurements for this patient | | Labs | 0 | | | | | labs |
| **phmv_first** | Physalis Mottle Virus (First) | continuous | First Physalis Mottle Virus measurement for this patient's hospitalization | | Labs | 0 | | | | | labs |
| **phmv_last** | Physalis Mottle Virus (Last) | continuous | Last Physalis Mottle Virus measurement for this patient's hospitalization | | Labs | 0 | | | | | labs |
| **phmv_max** | Physalis Mottle Virus (Max) | continuous | Maximum Physalis Mottle Virus measurement for this patient's hospitalization | | Labs | 0 | | | | | labs |
| **phmv_mean** | Physalis Mottle Virus (Mean) | continuous | Average Physalis Mottle Virus measurement for this patient's hospitalization | | Labs | 0 | | | | | labs |
| **phmv_median** | Physalis Mottle Virus (Median) | continuous | Median Physalis Mottle Virus measurement for this patient's hospitalization | | Labs | 0 | | | | | labs |
| **phmv_min** | Physalis Mottle Virus (Min) | continuous | Minimum Physalis Mottle Virus measurement for this patient's hospitalization | | Labs | 0 | | | | | labs |
| **phmv_sd** | Physalis Mottle Virus (SD) | continuous | Standard deviation of the Physalis Mottle Virus measurements during this patient's hospitalization | | Labs | 0 | | | | | labs |
| **phosphorous_25** | Phosphorous (25th Percentile) | continuous | Phosphorous level at the 25th percentile of this patient's measurements | mg/dl | Labs | 0 | | | | | labs |
| **phosphorous_75** | Phosphorous (75th Percentile) | continuous | Phosphorous level at the 75th percentile of this patient's measurements | mg/dl | Labs | 0 | | | | | labs |
| **phosphorous_count** | Phosphorous (Count) | continuous | Count of phosphorous level measurements for this patient | Number of measurements | Labs | 0 | | | | | labs |
| **phosphorous_first** | Phosphorous (First) | continuous | First phosphorous level measurement for this patient's hospitalization | mg/dl | Labs | 0 | | | | | labs |
| **phosphorous_last** | Phosphorous (Last) | continuous | Last phosphorous level measurement for this patient's hospitalization | mg/dl | Labs | 0 | | | | | labs |
| **phosphorous_max** | Phosphorous (Max) | continuous | Maximum phosphorous level measurement for this patient's hospitalization | mg/dl | Labs | 0 | | | | | labs |
| **phosphorous_mean** | Phosphorous (Mean) | continuous | Average phosphorous level measurement for this patient's hospitalization | mg/dl | Labs | 0 | | | | | labs |
| **phosphorous_median** | Phosphorous (Median) | continuous | Median phosphorous level measurement for this patient's hospitalization | mg/dl | Labs | 0 | | | | | labs |
| **phosphorous_min** | Phosphorous (Min) | continuous | Minimum phosphorous level measurement for this patient's hospitalization | mg/dl | Labs | 0 | | | | | labs |
| **phosphorous_sd** | Phosphorous (SD) | continuous | Standard deviation of the phosphorous level measurements during this patient's hospitalization | mg/dl | Labs | 0 | | | | | labs |
| **phvenous_25** | Venous pH (25th Percentile) | continuous | Venous pH level at the 25th percentile of this patient's measurements | | Labs | 0 | | | | | labs |
| **phvenous_75** | Venous pH (75th Percentile) | continuous | Venous pH level at the 75th percentile of this patient's measurements | | Labs | 0 | | | | | labs |
| **phvenous_count** | Venous pH (Count) | continuous | Count of venous pH measurements for this patient | Number of measurements | Labs | 0 | | | | | labs |
| **phvenous_first** | Venous pH (First) | continuous | First venous pH measurement for this patient's hospitalization | | Labs | 0 | | | | | labs |
| **phvenous_last** | Venous pH (Last) | continuous | Last venous pH measurement for this patient's hospitalization | | Labs | 0 | | | | | labs |
| **phvenous_max** | Venous pH (Max) | continuous | Maximum venous pH measurement for this patient's hospitalization | | Labs | 0 | | | | | labs |
| **phvenous_mean** | Venous pH (Mean) | continuous | Average venous pH measurement for this patient's hospitalization | | Labs | 0 | | | | | labs |
| **phvenous_median** | Venous pH (Median) | continuous | Median venous pH measurement for this patient's hospitalization | | Labs | 0 | | | | | labs |
| **phvenous_min** | Venous pH (Min) | continuous | Minimum venous pH measurement for this patient's hospitalization | | Labs | 0 | | | | | labs |
| **phvenous_sd** | Venous pH (SD) | continuous | Standard deviation of the venous pH measurements during this patient's hospitalization | | Labs | 0 | | | | | labs |
| **plateletcount_25** | Platelet Count (25th Percentile) | continuous | Platelet count at the 25th percentile of this patient's measurements | Platelets/Liter of blood | Labs | 0 | | | | | labs |
| **plateletcount_75** | Platelet Count (75th Percentile) | continuous | Platelet count at the 75th percentile of this patient's measurements | Platelets/Liter of blood | Labs | 0 | | | | | labs |
| **plateletcount_count** | Platelet Count (Count) | continuous | Count of platelet measurements for this patient | Number of measurements | Labs | 0 | | | | | labs |
| **plateletcount_first** | Platelet Count (First) | continuous | First platelet count measurement for this patient's hospitalization | Platelets/Liter of blood | Labs | 0 | | | | | labs |
| **plateletcount_last** | Platelet Count (Last) | continuous | Last platelet count measurement for this patient's hospitalization | Platelets/Liter of blood | Labs | 0 | | | | | labs |
| **plateletcount_max** | Platelet Count (Max) | continuous | Maximum platelet count measurement for this patient's hospitalization | Platelets/Liter of blood | Labs | 0 | | | | | labs |
| **plateletcount_mean** | Platelet Count (Mean) | continuous | Average platelet count measurement for this patient's hospitalization | Platelets/Liter of blood | Labs | 0 | | | | | labs |
| **plateletcount_median** | Platelet Count (Median) | continuous | Median platelet count measurement for this patient's hospitalization | Platelets/Liter of blood | Labs | 0 | | | | | labs |
| **plateletcount_min** | Platelet Count (Min) | continuous | Minimum platelet count measurement for this patient's hospitalization | Platelets/Liter of blood | Labs | 0 | | | | | labs |
| **plateletcount_sd** | Platelet Count (SD) | continuous | Standard deviation of the platelet count measurements during this patient's hospitalization | Platelets/Liter of blood | Labs | 0 | | | | | labs |
| **po2_first** | Partial Pressure of Oxygen (First) | categorical | First Partial Pressure of Oxygen measurement for this patient's hospitalization | | Labs | 0 | | | | | |
| **po2_last** | Partial Pressure of Oxygen (Last) | categorical | Last Partial Pressure of Oxygen measurement for this patient's hospitalization | | Labs | 0 | | | | | |
| **po2art_first** | Arterial Partial Pressure of Oxygen (First) | categorical | First Arterial Partial Pressure of Oxygen count measurement for this patient's hospitalization | | Labs | 0 | | | | | |
| **po2art_last** | Arterial Partial Pressure of Oxygen (Last) | categorical | Last Arterial Partial Pressure of Oxygen count measurement for this patient's hospitalization | | Labs | 0 | | | | | |
| **po2mv_25** | | continuous | | | Labs | 0 | | | | | labs |
| **po2mv_75** | | continuous | | | Labs | 0 | | | | | labs |
| **po2mv_count** | | continuous | | | Labs | 0 | | | | | labs |
| **po2mv_first** | | continuous | | | Labs | 0 | | | | | labs |
| **po2mv_last** | | continuous | | | Labs | 0 | | | | | labs |
| **po2mv_max** | | continuous | | | Labs | 0 | | | | | labs |
| **po2mv_mean** | | continuous | | | Labs | 0 | | | | | labs |
| **po2mv_median** | | continuous | | | Labs | 0 | | | | | labs |
| **po2mv_min** | | continuous | | | Labs | 0 | | | | | labs |
| **po2mv_sd** | | continuous | | | Labs | 0 | | | | | labs |
| **po2ven_first** | Venous Partial Pressure of Oxygen (First) | categorical | First Venous Partial Pressure of Oxygen count measurement for this patient's hospitalization | | Labs | 0 | | | | | |
| **po2ven_last** | Venous Partial Pressure of Oxygen (Last) | categorical | Last Venous Partial Pressure of Oxygen count measurement for this patient's hospitalization | | Labs | 0 | | | | | |
| **potassium_25** | Potassium (25th Percentile) | continuous | Potassium level at the 25th percentile of this patient's measurements | mmol/L | Labs | 0 | | | | | labs |
| **potassium_75** | Potassium (75th Percentile) | continuous | Potassium level at the 75th percentile of this patient's measurements | mmol/L | Labs | 0 | | | | | labs |
| **potassium_count** | Potassium (Count) | continuous | Count of potassium level measurements for this patient | Number of measurements | Labs | 0 | | | | | labs |
| **potassium_first** | Potassium (First) | continuous | First potassium level measurement for this patient's hospitalization | mmol/L | Labs | 0 | | | | | labs |
| **potassium_last** | Potassium (Last) | continuous | Last potassium level measurement for this patient's hospitalization | mmol/L | Labs | 0 | | | | | labs |
| **potassium_max** | Potassium (Max) | continuous | Maximum potassium level measurement for this patient's hospitalization | mmol/L | Labs | 0 | | | | | labs |
| **potassium_mean** | Potassium (Mean) | continuous | Average potassium level measurement for this patient's hospitalization | mmol/L | Labs | 0 | | | | | labs |
| **potassium_median** | Potassium (Median) | continuous | Median potassium level measurement for this patient's hospitalization | mmol/L | Labs | 0 | | | | | labs |
| **potassium_min** | Potassium (Min) | continuous | Minimum potassium level measurement for this patient's hospitalization | mmol/L | Labs | 0 | | | | | labs |
| **potassium_sd** | Potassium (SD) | continuous | Standard deviation of the potassium level measurements during this patient's hospitalization | mmol/L | Labs | 0 | | | | | labs |
| **prealbumin_25** | Prealbumin (25th Percentile) | continuous | Prealbumin level at the 25th percentile of this patient's measurements | mg/dL | Labs | 0 | | | | | labs |
| **prealbumin_75** | Prealbumin (75th Percentile) | continuous | Prealbumin level at the 75th percentile of this patient's measurements | mg/dL | Labs | 0 | | | | | labs |
| **prealbumin_count** | Prealbumin (Count) | continuous | Count of prealbumin level measurements for this patient | Number of measurements | Labs | 0 | | | | | labs |
| **prealbumin_first** | Prealbumin (First) | continuous | First prealbumin level measurement for this patient's hospitalization | mg/dL | Labs | 0 | | | | | labs |
| **prealbumin_last** | Prealbumin (Last) | continuous | Last prealbumin level measurement for this patient's hospitalization | mg/dL | Labs | 0 | | | | | labs |
| **prealbumin_max** | Prealbumin (Max) | continuous | Maximum prealbumin level measurement for this patient's hospitalization | mg/dL | Labs | 0 | | | | | labs |
| **prealbumin_mean** | Prealbumin (Mean) | continuous | Average prealbumin level measurement for this patient's hospitalization | mg/dL | Labs | 0 | | | | | labs |
| **prealbumin_median** | Prealbumin (Median) | continuous | Median prealbumin level measurement for this patient's hospitalization | mg/dL | Labs | 0 | | | | | labs |

| | | | | | | | | | | | |
|---|---|---|---|---|---|---|---|---|---|---|---|
| **prealbumin_min** | Prealbumin (Min) | continuous | Minimum prealbumin level measurement for this patient's hospitalization | mg/dL | Labs | 0 | | | | | labs |
| **prealbumin_sd** | Prealbumin (SD) | continuous | Standard deviation of the prealbumin level measurements during this patient's hospitalization | mg/dL | Labs | 0 | | | | | labs |
| **pregnancy_first** | Pregnancy (First) | categorical | Results of the first pregnancy test during this patient's hospitalization | | Labs | 0 | | | | | labs |
| **pregnancy_last** | Pregnancy (Last) | categorical | Results of the last pregnancy test during this patient's hospitalization | | Labs | 0 | | | | | labs |
| **procalcitonin_25** | Procalcitonin (25th Percentile) | continuous | Procalcitonin level at the 25th percentile of this patient's measurements | ng/mL | Labs | 0 | | | | | labs |
| **procalcitonin_75** | Procalcitonin (75th Percentile) | continuous | Procalcitonin level at the 75th percentile of this patient's measurements | ng/mL | Labs | 0 | | | | | labs |
| **procalcitonin_count** | Procalcitonin (Count) | continuous | Count of procalcitonin level measurements for this patient | Number of measurements | Labs | 0 | | | | | labs |
| **procalcitonin_first** | Procalcitonin (First) | continuous | First procalcitonin level measurement for this patient's hospitalization | ng/mL | Labs | 0 | | | | | labs |
| **procalcitonin_last** | Procalcitonin (Last) | continuous | Last procalcitonin level measurement for this patient's hospitalization | ng/mL | Labs | 0 | | | | | labs |
| **procalcitonin_max** | Procalcitonin (Max) | continuous | Maximum procalcitonin level measurement for this patient's hospitalization | ng/mL | Labs | 0 | | | | | labs |
| **procalcitonin_mean** | Procalcitonin (Mean) | continuous | Average procalcitonin level measurement for this patient's hospitalization | ng/mL | Labs | 0 | | | | | labs |
| **procalcitonin_median** | Procalcitonin (Median) | continuous | Median procalcitonin level measurement for this patient's hospitalization | ng/mL | Labs | 0 | | | | | labs |
| **procalcitonin_min** | Procalcitonin (Min) | continuous | Minimum procalcitonin level measurement for this patient's hospitalization | ng/mL | Labs | 0 | | | | | labs |
| **procalcitonin_sd** | Procalcitonin (SD) | continuous | Standard deviation of the procalcitonin level measurements during this patient's hospitalization | ng/mL | Labs | 0 | | | | | labs |
| **protein_total_25** | Total Protein (25th Percentile) | continuous | Total protein level at the 25th percentile of this patient's measurements | g/dL | Labs | 0 | | | | | labs |
| **protein_total_75** | Total Protein (75th Percentile) | continuous | Total protein level at the 75th percentile of this patient's measurements | g/dL | Labs | 0 | | | | | labs |
| **protein_total_count** | Total Protein (Count) | continuous | Count of total protein level measurements for this patient | Number of measurements | Labs | 0 | | | | | labs |
| **protein_total_first** | Total Protein (First) | continuous | First total protein level measurement for this patient's hospitalization | g/dL | Labs | 0 | | | | | labs |
| **protein_total_last** | Total Protein (Last) | continuous | Last total protein level measurement for this patient's hospitalization | g/dL | Labs | 0 | | | | | labs |
| **protein_total_max** | Total Protein (Max) | continuous | Maximum total protein level measurement for this patient's hospitalization | g/dL | Labs | 0 | | | | | labs |
| **protein_total_mean** | Total Protein (Mean) | continuous | Average total protein level measurement for this patient's hospitalization | g/dL | Labs | 0 | | | | | labs |
| **protein_total_median** | Total Protein (Median) | continuous | Median total protein level measurement for this patient's hospitalization | g/dL | Labs | 0 | | | | | labs |
| **protein_total_min** | Total Protein (Min) | continuous | Minimum total protein level measurement for this patient's hospitalization | g/dL | Labs | 0 | | | | | labs |
| **protein_total_sd** | Total Protein (SD) | continuous | Standard deviation of the total protein level measurements during this patient's hospitalization | g/dL | Labs | 0 | | | | | labs |
| **proteinserum_25** | Protein Serum (25th Percentile) | continuous | Protein serum level at the 25th percentile of this patient's measurements | g/dL | Labs | 0 | | | | | labs |
| **proteinserum_75** | Protein Serum (75th Percentile) | continuous | Protein serum level at the 75th percentile of this patient's measurements | g/dL | Labs | 0 | | | | | labs |
| **proteinserum_count** | Protein Serum (Count) | continuous | Count of protein serum measurements for this patient | Number of measurements | Labs | 0 | | | | | labs |
| **proteinserum_first** | Protein Serum (First) | continuous | First protein serum level measurement for this patient's hospitalization | g/dL | Labs | 0 | | | | | labs |
| **proteinserum_last** | Protein Serum (Last) | continuous | Last protein serum level measurement for this patient's hospitalization | g/dL | Labs | 0 | | | | | labs |
| **proteinserum_max** | Protein Serum (Max) | continuous | Maximum protein serum level measurement for this patient's hospitalization | g/dL | Labs | 0 | | | | | labs |
| **proteinserum_mean** | Protein Serum (Mean) | continuous | Average protein serum level measurement for this patient's hospitalization | g/dL | Labs | 0 | | | | | labs |
| **proteinserum_median** | Protein Serum (Median) | continuous | Median protein serum level measurement for this patient's hospitalization | g/dL | Labs | 0 | | | | | labs |
| **proteinserum_min** | Protein Serum (Min) | continuous | Minimum protein serum level measurement for this patient's hospitalization | g/dL | Labs | 0 | | | | | labs |
| **proteinserum_sd** | Protein Serum (SD) | continuous | Standard deviation of the protein serum measurements during this patient's hospitalization | g/dL | Labs | 0 | | | | | labs |
| **psa_25** | Prostate-Specific Antigen (25th Percentile) | continuous | Prostate-specific antigen level at the 25th percentile of this patient's measurements | ng/mL | Labs | 0 | | | | | labs |
| **psa_75** | Prostate-Specific Antigen (75th Percentile) | continuous | Prostate-specific antigen level at the 75th percentile of this patient's measurements | ng/mL | Labs | 0 | | | | | labs |
| **psa_count** | Prostate-Specific Antigen (Count) | continuous | Count of prostate-specific antigen level measurements for this patient | Number of measurements | Labs | 0 | | | | | labs |
| **psa_first** | Prostate-Specific Antigen (First) | continuous | First prostate-specific antigen level measurement for this patient's hospitalization | ng/mL | Labs | 0 | | | | | labs |
| **psa_last** | Prostate-Specific Antigen (Last) | continuous | Last prostate-specific antigen level measurement for this patient's hospitalization | ng/mL | Labs | 0 | | | | | labs |
| **psa_max** | Prostate-Specific Antigen (Max) | continuous | Maximum prostate-specific antigen level measurement for this patient's hospitalization | ng/mL | Labs | 0 | | | | | labs |
| **psa_mean** | Prostate-Specific Antigen (Mean) | continuous | Average prostate-specific antigen level measurement for this patient's hospitalization | ng/mL | Labs | 0 | | | | | labs |
| **psa_median** | Prostate-Specific Antigen (Median) | continuous | Median prostate-specific antigen level measurement for this patient's hospitalization | ng/mL | Labs | 0 | | | | | labs |
| **psa_min** | Prostate-Specific Antigen (Min) | continuous | Minimum prostate-specific antigen level measurement for this patient's hospitalization | ng/mL | Labs | 0 | | | | | labs |
| **psa_sd** | Prostate-Specific Antigen (SD) | continuous | Standard deviation of the y measurements during this patient's hospitalization | ng/mL | Labs | 0 | | | | | labs |
| **pt_25** | Prothrombin Time (25th Percentile) | continuous | Prothrombin time at the 25th percentile of this patient's measurements | Seconds | Labs | 0 | | | | | labs |
| **pt_75** | Prothrombin Time (75th Percentile) | continuous | Prothrombin time at the 75th percentile of this patient's measurements | Seconds | Labs | 0 | | | | | labs |
| **pt_count** | Prothrombin Time (Count) | continuous | Count of prothrombin time measurements for this patient | Number of measurements | Labs | 0 | | | | | labs |
| **pt_first** | Prothrombin Time (First) | continuous | First prothrombin time measurement for this patient's hospitalization | Seconds | Labs | 0 | | | | | labs |
| **pt_last** | Prothrombin Time (Last) | continuous | Last prothrombin time measurement for this patient's hospitalization | Seconds | Labs | 0 | | | | | labs |
| **pt_max** | Prothrombin Time (Max) | continuous | Maximum prothrombin time measurement for this patient's hospitalization | Seconds | Labs | 0 | | | | | labs |
| **pt_mean** | Prothrombin Time (Mean) | continuous | Average prothrombin time measurement for this patient's hospitalization | Seconds | Labs | 0 | | | | | labs |
| **pt_median** | Prothrombin Time (Median) | continuous | Median prothrombin time measurement for this patient's hospitalization | Seconds | Labs | 0 | | | | | labs |
| **pt_min** | Prothrombin Time (Min) | continuous | Minimum prothrombin time measurement for this patient's hospitalization | Seconds | Labs | 0 | | | | | labs |
| **pt_sd** | Prothrombin Time (SD) | continuous | Standard deviation of the prothrombin time measurements during this patient's hospitalization | Seconds | Labs | 0 | | | | | labs |
| **pthintact_25** | Parathyroid Hormone (25th Percentile) | continuous | Parathyroid hormone level at the 25th percentile of this patient's measurements | pg/mL | Labs | 0 | | | | | labs |
| **pthintact_75** | Parathyroid Hormone (75th Percentile) | continuous | Parathyroid hormone level at the 75th percentile of this patient's measurements | pg/mL | Labs | 0 | | | | | labs |
| **pthintact_count** | Parathyroid Hormone (Count) | continuous | Count of parathyroid hormone level measurements for this patient | Number of measurements | Labs | 0 | | | | | labs |
| **pthintact_first** | Parathyroid Hormone (First) | continuous | First parathyroid hormone level measurement for this patient's hospitalization | pg/mL | Labs | 0 | | | | | labs |
| **pthintact_last** | Parathyroid Hormone (Last) | continuous | Last parathyroid hormone level measurement for this patient's hospitalization | pg/mL | Labs | 0 | | | | | labs |
| **pthintact_max** | Parathyroid Hormone (Max) | continuous | Maximum parathyroid hormone level measurement for this patient's hospitalization | pg/mL | Labs | 0 | | | | | labs |
| **pthintact_mean** | Parathyroid Hormone (Mean) | continuous | Average parathyroid hormone level measurement for this patient's hospitalization | pg/mL | Labs | 0 | | | | | labs |
| **pthintact_median** | Parathyroid Hormone (Median) | continuous | Median parathyroid hormone level measurement for this patient's hospitalization | pg/mL | Labs | 0 | | | | | labs |
| **pthintact_min** | Parathyroid Hormone (Min) | continuous | Minimum parathyroid hormone level measurement for this patient's hospitalization | pg/mL | Labs | 0 | | | | | labs |
| **pthintact_sd** | Parathyroid Hormone (SD) | continuous | Standard deviation of the parathyroid hormone level measurements during this patient's hospitalization | pg/mL | Labs | 0 | | | | | labs |
| **ptt_25** | Partial Thromboplastin Time (25th Percentile) | continuous | Partial thromboplastin time at the 25th percentile of this patient's measurements | Seconds | Labs | 0 | | | | | labs |
| **ptt_75** | Partial Thromboplastin Time (75th Percentile) | continuous | Partial thromboplastin time at the 75th percentile of this patient's measurements | Seconds | Labs | 0 | | | | | labs |
| **ptt_count** | Partial Thromboplastin Time (Count) | continuous | Count of partial thromboplastin time measurements for this patient | Number of measurements | Labs | 0 | | | | | labs |
| **ptt_first** | Partial Thromboplastin Time (First) | continuous | First partial thromboplastin time measurement for this patient's hospitalization | Seconds | Labs | 0 | | | | | labs |
| **ptt_last** | Partial Thromboplastin Time (Last) | continuous | Last partial thromboplastin time measurement for this patient's hospitalization | Seconds | Labs | 0 | | | | | labs |
| **ptt_max** | Partial Thromboplastin Time (Max) | continuous | Maximum partial thromboplastin time measurement for this patient's hospitalization | Seconds | Labs | 0 | | | | | labs |
| **ptt_mean** | Partial Thromboplastin Time (Mean) | continuous | Average partial thromboplastin time measurement for this patient's hospitalization | Seconds | Labs | 0 | | | | | labs |
| **ptt_median** | Partial Thromboplastin Time (Median) | continuous | Median partial thromboplastin time measurement for this patient's hospitalization | Seconds | Labs | 0 | | | | | labs |
| **ptt_min** | Partial Thromboplastin Time (Min) | continuous | Minimum partial thromboplastin time measurement for this patient's hospitalization | Seconds | Labs | 0 | | | | | labs |
| **ptt_sd** | Partial Thromboplastin Time (SD) | continuous | Standard deviation of the partial thromboplastin time measurements during this patient's hospitalization | Seconds | Labs | 0 | | | | | labs |
| **rapamycin_25** | Sirolimus (25th Percentile) | continuous | Sirolimus level at the 25th percentile of this patient's measurements | ng/mL | Labs | 0 | | | | | labs |
| **rapamycin_75** | Sirolimus (75th Percentile) | continuous | Sirolimus level at the 75th percentile of this patient's measurements | ng/mL | Labs | 0 | | | | | labs |
| **rapamycin_count** | Sirolimus (Count) | continuous | Count of sirolimus level measurements for this patient | Number of measurements | Labs | 0 | | | | | labs |
| **rapamycin_first** | Sirolimus (First) | continuous | First sirolimus level measurement for this patient's hospitalization | ng/mL | Labs | 0 | | | | | labs |
| **rapamycin_last** | Sirolimus (Last) | continuous | Last sirolimus level measurement for this patient's hospitalization | ng/mL | Labs | 0 | | | | | labs |
| **rapamycin_max** | Sirolimus (Max) | continuous | Maximum sirolimus level measurement for this patient's hospitalization | ng/mL | Labs | 0 | | | | | labs |
| **rapamycin_mean** | Sirolimus (Mean) | continuous | Average sirolimus level measurement for this patient's hospitalization | ng/mL | Labs | 0 | | | | | labs |
| **rapamycin_median** | Sirolimus (Median) | continuous | Median sirolimus level measurement for this patient's hospitalization | ng/mL | Labs | 0 | | | | | labs |
| **rapamycin_min** | Sirolimus (Min) | continuous | Minimum sirolimus level measurement for this patient's hospitalization | ng/mL | Labs | 0 | | | | | labs |
| **rapamycin_sd** | Sirolimus (SD) | continuous | Standard deviation of the sirolimus level measurements during this patient's hospitalization | ng/mL | Labs | 0 | | | | | labs |
| **rbcc_25** | Red Blood Cell Count (25th Percentile) | continuous | Red blood cell count at the 25th percentile of this patient's measurements | million cells/mcL | Labs | 0 | | | | | labs |
| **rbcc_75** | Red Blood Cell Count (75th Percentile) | continuous | Red blood cell count at the 75th percentile of this patient's measurements | million cells/mcL | Labs | 0 | | | | | labs |
| **rbcc_count** | Red Blood Cell Count (Count) | continuous | Count of red blood cell measurements for this patient | Number of measurements | Labs | 0 | | | | | labs |
| **rbcc_first** | Red Blood Cell Count (First) | continuous | First red blood cell count measurement for this patient's hospitalization | million cells/mcL | Labs | 0 | | | | | labs |
| **rbcc_last** | Red Blood Cell Count (Last) | continuous | Last red blood cell count measurement for this patient's hospitalization | million cells/mcL | Labs | 0 | | | | | labs |
| **rbcc_max** | Red Blood Cell Count (Max) | continuous | Maximum red blood cell count measurement for this patient's hospitalization | million cells/mcL | Labs | 0 | | | | | labs |
| **rbcc_mean** | Red Blood Cell Count (Mean) | continuous | Average red blood cell count measurement for this patient's hospitalization | million cells/mcL | Labs | 0 | | | | | labs |
| **rbcc_median** | Red Blood Cell Count (Median) | continuous | Median red blood cell count measurement for this patient's hospitalization | million cells/mcL | Labs | 0 | | | | | labs |
| **rbcc_min** | Red Blood Cell Count (Min) | continuous | Minimum red blood cell count measurement for this patient's hospitalization | million cells/mcL | Labs | 0 | | | | | labs |
| **rbcc_sd** | Red Blood Cell Count (SD) | continuous | Standard deviation of the red blood cell count measurements during this patient's hospitalization | million cells/mcL | Labs | 0 | | | | | labs |
| **rdw_25** | Red Cell Distribution Width (25th Percentile) | continuous | Red cell distribution width at the 25th percentile of this patient's measurements | % | Labs | 0 | | | | | labs |
| **rdw_75** | Red Cell Distribution Width (75th Percentile) | continuous | Red cell distribution width at the 75th percentile of this patient's measurements | % | Labs | 0 | | | | | labs |
| **rdw_count** | Red Cell Distribution Width (Count) | continuous | Count of red cell distribution width measurements for this patient | Number of measurements | Labs | 0 | | | | | labs |
| **rdw_first** | Red Cell Distribution Width (First) | continuous | First red cell distribution width measurement for this patient's hospitalization | % | Labs | 0 | | | | | labs |
| **rdw_last** | Red Cell Distribution Width (Last) | continuous | Last red cell distribution width measurement for this patient's hospitalization | % | Labs | 0 | | | | | labs |
| **rdw_max** | Red Cell Distribution Width (Max) | continuous | Maximum red cell distribution width measurement for this patient's hospitalization | % | Labs | 0 | | | | | labs |
| **rdw_mean** | Red Cell Distribution Width (Mean) | continuous | Average red cell distribution width measurement for this patient's hospitalization | % | Labs | 0 | | | | | labs |
| **rdw_median** | Red Cell Distribution Width (Median) | continuous | Median red cell distribution width measurement for this patient's hospitalization | % | Labs | 0 | | | | | labs |
| **rdw_min** | Red Cell Distribution Width (Min) | continuous | Minimum red cell distribution width measurement for this patient's hospitalization | % | Labs | 0 | | | | | labs |
| **rdw_sd** | Red Cell Distribution Width (SD) | continuous | Standard deviation of the red cell distribution width measurements during this patient's hospitalization | % | Labs | 0 | | | | | labs |
| **reticabs_25** | Absolute Reticulocyte (25th Percentile) | continuous | Absolute reticulocyte count at the 25th percentile of this patient's measurements | | Labs | 0 | | | | | labs |
| **reticabs_75** | Absolute Reticulocyte (75th Percentile) | continuous | Absolute reticulocyte count at the 75th percentile of this patient's measurements | | Labs | 0 | | | | | labs |
| **reticabs_count** | Absolute Reticulocyte (Count) | continuous | Count of absolute reticulocyte measurements for this patient | Number of measurements | Labs | 0 | | | | | labs |
| **reticabs_first** | Absolute Reticulocyte (First) | continuous | First absolute reticulocyte measurement for this patient's hospitalization | | Labs | 0 | | | | | labs |
| **reticabs_last** | Absolute Reticulocyte (Last) | continuous | Last absolute reticulocyte measurement for this patient's hospitalization | | Labs | 0 | | | | | labs |
| **reticabs_max** | Absolute Reticulocyte (Max) | continuous | Maximum absolute reticulocyte measurement for this patient's hospitalization | | Labs | 0 | | | | | labs |
| **reticabs_mean** | Absolute Reticulocyte (Mean) | continuous | Average absolute reticulocyte measurement for this patient's hospitalization | | Labs | 0 | | | | | labs |
| **reticabs_median** | Absolute Reticulocyte (Median) | continuous | Median absolute reticulocyte measurement for this patient's hospitalization | | Labs | 0 | | | | | labs |
| **reticabs_min** | Absolute Reticulocyte (Min) | continuous | Minimum absolute reticulocyte measurement for this patient's hospitalization | | Labs | 0 | | | | | labs |
| **reticabs_sd** | Absolute Reticulocyte (SD) | continuous | Standard deviation of the absolute reticulocyte measurements during this patient's hospitalization | | Labs | 0 | | | | | labs |
| **reticpct_25** | Reticulocyte Percentage (25th Percentile) | continuous | Reticulocyte percentage at the 25th percentile of this patient's measurements | % | Labs | 0 | | | | | labs |
| **reticpct_75** | Reticulocyte Percentage (75th Percentile) | continuous | Reticulocyte percentage at the 75th percentile of this patient's measurements | % | Labs | 0 | | | | | labs |
| **reticpct_count** | Reticulocyte Percentage (Count) | continuous | Count of reticulocyte percentage measurements for this patient | Number of measurements | Labs | 0 | | | | | labs |
| **reticpct_first** | Reticulocyte Percentage (First) | continuous | First reticulocyte percentage measurement for this patient's hospitalization | % | Labs | 0 | | | | | labs |
| **reticpct_last** | Reticulocyte Percentage (Last) | continuous | Last reticulocyte percentage measurement for this patient's hospitalization | % | Labs | 0 | | | | | labs |
| **reticpct_max** | Reticulocyte Percentage (Max) | continuous | Maximum reticulocyte percentage measurement for this patient's hospitalization | % | Labs | 0 | | | | | labs |
| **reticpct_mean** | Reticulocyte Percentage (Mean) | continuous | Average reticulocyte percentage measurement for this patient's hospitalization | % | Labs | 0 | | | | | labs |
| **reticpct_median** | Reticulocyte Percentage (Median) | continuous | Median reticulocyte percentage measurement for this patient's hospitalization | % | Labs | 0 | | | | | labs |
| **reticpct_min** | Reticulocyte Percentage (Min) | continuous | Minimum reticulocyte percentage measurement for this patient's hospitalization | % | Labs | 0 | | | | | labs |
| **reticpct_sd** | Reticulocyte Percentage (SD) | continuous | Standard deviation of the reticulocyte percentage measurements during this patient's hospitalization | % | Labs | 0 | | | | | labs |

| | | | | | | | | | | | |
|---|---|---|---|---|---|---|---|---|---|---|---|
| **rf_25** | Rheumatoid Factor (25th Percentile) | continuous | Rheumatoid factor at the 25th percentile of this patient's measurements | IU/ml | Labs | 0 | | | | | labs |
| **rf_75** | Rheumatoid Factor (75th Percentile) | continuous | Rheumatoid factor at the 75th percentile of this patient's measurements | IU/ml | Labs | 0 | | | | | labs |
| **rf_count** | Rheumatoid Factor (Count) | continuous | Count of rheumatoid factor measurements for this patient | Number of measurements | Labs | 0 | | | | | labs |
| **rf_first** | Rheumatoid Factor (First) | continuous | First rheumatoid factor measurement for this patient's hospitalization | IU/ml | Labs | 0 | | | | | labs |
| **rf_last** | Rheumatoid Factor (Last) | continuous | Last rheumatoid factor measurement for this patient's hospitalization | IU/ml | Labs | 0 | | | | | labs |
| **rf_max** | Rheumatoid Factor (Max) | continuous | Maximum rheumatoid factor measurement for this patient's hospitalization | IU/ml | Labs | 0 | | | | | labs |
| **rf_mean** | Rheumatoid Factor (Mean) | continuous | Average rheumatoid factor measurement for this patient's hospitalization | IU/ml | Labs | 0 | | | | | labs |
| **rf_median** | Rheumatoid Factor (Median) | continuous | Median rheumatoid factor measurement for this patient's hospitalization | IU/ml | Labs | 0 | | | | | labs |
| **rf_min** | Rheumatoid Factor (Min) | continuous | Minimum rheumatoid factor measurement for this patient's hospitalization | IU/ml | Labs | 0 | | | | | labs |
| **rf_sd** | Rheumatoid Factor (SD) | continuous | Standard deviation of the rheumatoid factor measurements during this patient's hospitalization | IU/ml | Labs | 0 | | | | | labs |
| **rhinovirus_first** | Rhinovirus (First) | categorical | Results of the first rhinovirus test during this patient's hospitalization | | Labs | 0 | | | | | labs |
| **rhinovirus_last** | Rhinovirus (Last) | categorical | Results of the last rhinovirus test during this patient's hospitalization | | Labs | 0 | | | | | labs |
| **rsv_first** | Respiratory Syncytial Virus (First) | categorical | Results of the first respiratory syncytial virus test during this patient's hospitalization | | Labs | 0 | | | | | labs |
| **rsv_last** | Respiratory Syncytial Virus (Last) | categorical | Results of the last respiratory syncytial virus test during this patient's hospitalization | | Labs | 0 | | | | | labs |
| **running_aki_stage** | Running AKI Stage | categorical | Stage of Running AKI diagnosis for this patient | | Labs | 0 | | | | | |
| **salicylate_25** | Salicylate (25th Percentile) | continuous | Salicylate level at the 25th percentile of this patient's measurements | mg/dL | Labs | 0 | | | | | labs |
| **salicylate_75** | Salicylate (75th Percentile) | continuous | Salicylate level at the 75th percentile of this patient's measurements | mg/dL | Labs | 0 | | | | | labs |
| **salicylate_count** | Salicylate (Count) | continuous | Count of salicylate level measurements for this patient | Number of measurements | Labs | 0 | | | | | labs |
| **salicylate_first** | Salicylate (First) | continuous | First salicylate level measurement for this patient's hospitalization | mg/dL | Labs | 0 | | | | | labs |
| **salicylate_last** | Salicylate (Last) | continuous | Last salicylate level measurement for this patient's hospitalization | mg/dL | Labs | 0 | | | | | labs |
| **salicylate_max** | Salicylate (Max) | continuous | Maximum salicylate level measurement for this patient's hospitalization | mg/dL | Labs | 0 | | | | | labs |
| **salicylate_mean** | Salicylate (Mean) | continuous | Average salicylate level measurement for this patient's hospitalization | mg/dL | Labs | 0 | | | | | labs |
| **salicylate_median** | Salicylate (Median) | continuous | Median salicylate level measurement for this patient's hospitalization | mg/dL | Labs | 0 | | | | | labs |
| **salicylate_min** | Salicylate (Min) | continuous | Minimum salicylate level measurement for this patient's hospitalization | mg/dL | Labs | 0 | | | | | labs |
| **salicylate_sd** | Salicylate (SD) | continuous | Standard deviation of the salicylate level measurements during this patient's hospitalization | mg/dL | Labs | 0 | | | | | labs |
| **schistocytes_first** | Schistocyte (First) | categorical | Results of the first schistocyte blood test during this patient's hospitalization | | Labs | 0 | | | | | labs |
| **schistocytes_last** | Schistocyte (Last) | categorical | Results of the last schistocyte blood test during this patient's hospitalization | | Labs | 0 | | | | | labs |
| **sedrate_25** | Sedimentation Rate (25th Percentile) | continuous | Sedimentation rate at the 25th percentile of this patient's measurements | mm/hr | Labs | 0 | | | | | labs |
| **sedrate_75** | Sedimentation Rate (75th Percentile) | continuous | Sedimentation rate at the 75th percentile of this patient's measurements | mm/hr | Labs | 0 | | | | | labs |
| **sedrate_count** | Sedimentation Rate (Count) | continuous | Count of sedimentation rate measurements for this patient | Number of measurements | Labs | 0 | | | | | labs |
| **sedrate_first** | Sedimentation Rate (First) | continuous | First sedimentation rate measurement for this patient's hospitalization | mm/hr | Labs | 0 | | | | | labs |
| **sedrate_last** | Sedimentation Rate (Last) | continuous | Last sedimentation rate measurement for this patient's hospitalization | mm/hr | Labs | 0 | | | | | labs |
| **sedrate_max** | Sedimentation Rate (Max) | continuous | Maximum sedimentation rate measurement for this patient's hospitalization | mm/hr | Labs | 0 | | | | | labs |
| **sedrate_mean** | Sedimentation Rate (Mean) | continuous | Average sedimentation rate measurement for this patient's hospitalization | mm/hr | Labs | 0 | | | | | labs |
| **sedrate_median** | Sedimentation Rate (Median) | continuous | Median sedimentation rate measurement for this patient's hospitalization | mm/hr | Labs | 0 | | | | | labs |
| **sedrate_min** | Sedimentation Rate (Min) | continuous | Minimum sedimentation rate measurement for this patient's hospitalization | mm/hr | Labs | 0 | | | | | labs |
| **sedrate_sd** | Sedimentation Rate (SD) | continuous | Standard deviation of the sedimentation rate measurements during this patient's hospitalization | mm/hr | Labs | 0 | | | | | labs |
| **sodium_25** | Sodium (25th Percentile) | continuous | Sodium level at the 25th percentile of this patient's measurements | mEq/L | Labs | 0 | | | | | labs |
| **sodium_75** | Sodium (75th Percentile) | continuous | Sodium level at the 75th percentile of this patient's measurements | mEq/L | Labs | 0 | | | | | labs |
| **sodium_count** | Sodium (Count) | continuous | Count of sodium level measurements for this patient | Number of measurements | Labs | 0 | | | | | labs |
| **sodium_first** | Sodium (First) | continuous | First sodium level measurement for this patient's hospitalization | mEq/L | Labs | 0 | | | | | labs |
| **sodium_last** | Sodium (Last) | continuous | Last sodium level measurement for this patient's hospitalization | mEq/L | Labs | 0 | | | | | labs |
| **sodium_max** | Sodium (Max) | continuous | Maximum sodium level measurement for this patient's hospitalization | mEq/L | Labs | 0 | | | | | labs |
| **sodium_mean** | Sodium (Mean) | continuous | Average sodium level measurement for this patient's hospitalization | mEq/L | Labs | 0 | | | | | labs |
| **sodium_median** | Sodium (Median) | continuous | Median sodium level measurement for this patient's hospitalization | mEq/L | Labs | 0 | | | | | labs |
| **sodium_min** | Sodium (Min) | continuous | Minimum sodium level measurement for this patient's hospitalization | mEq/L | Labs | 0 | | | | | labs |
| **sodium_sd** | Sodium (SD) | continuous | Standard deviation of the sodium level measurements during this patient's hospitalization | mEq/L | Labs | 0 | | | | | labs |
| **spep_first** | Serum Protein Electrophoresis (First) | categorical | Results of the first serum protein electrophoresis test during this patient's hospitalization | | Labs | 0 | | | | | labs |
| **spep_last** | Serum Protein Electrophoresis (Last) | categorical | Results of the last serum protein electrophoresis test during this patient's hospitalization | | Labs | 0 | | | | | labs |
| **streppneumo_first** | Streptococcus Pneumoniae (First) | categorical | Results of the first streptococcus pneumoniae test during this patient's hospitalization | | Labs | 0 | | | | | labs |
| **streppneumo_last** | Streptococcus Pneumoniae (Last) | categorical | Results of the last streptococcus pneumoniae test during this patient's hospitalization | | Labs | 0 | | | | | labs |
| **t3_25** | Triiodothyronine (25th Percentile) | continuous | Triiodothyronine level at the 25th percentile of this patient's measurements | ng/dL | Labs | 0 | | | | | labs |
| **t3_75** | Triiodothyronine (75th Percentile) | continuous | Triiodothyronine level at the 75th percentile of this patient's measurements | ng/dL | Labs | 0 | | | | | labs |
| **t3_count** | Triiodothyronine (Count) | continuous | Count of triiodothyronine level measurements for this patient | Number of measurements | Labs | 0 | | | | | labs |
| **t3_first** | Triiodothyronine (First) | continuous | First triiodothyronine level measurement for this patient's hospitalization | ng/dL | Labs | 0 | | | | | labs |
| **t3_last** | Triiodothyronine (Last) | continuous | Last triiodothyronine level measurement for this patient's hospitalization | ng/dL | Labs | 0 | | | | | labs |
| **t3_max** | Triiodothyronine (Max) | continuous | Maximum triiodothyronine level measurement for this patient's hospitalization | ng/dL | Labs | 0 | | | | | labs |
| **t3_mean** | Triiodothyronine (Mean) | continuous | Average triiodothyronine level measurement for this patient's hospitalization | ng/dL | Labs | 0 | | | | | labs |
| **t3_median** | Triiodothyronine (Median) | continuous | Median triiodothyronine level measurement for this patient's hospitalization | ng/dL | Labs | 0 | | | | | labs |
| **t3_min** | Triiodothyronine (Min) | continuous | Minimum triiodothyronine level measurement for this patient's hospitalization | ng/dL | Labs | 0 | | | | | labs |
| **t3_sd** | Triiodothyronine (SD) | continuous | Standard deviation of the triiodothyronine level measurements during this patient's hospitalization | ng/dL | Labs | 0 | | | | | labs |
| **t4free_25** | Free Thyroxine (25th Percentile) | continuous | Free thyroxine level at the 25th percentile of this patient's measurements | ng/dl | Labs | 0 | | | | | labs |
| **t4free_75** | Free Thyroxine (75th Percentile) | continuous | Free thyroxine level at the 75th percentile of this patient's measurements | ng/dl | Labs | 0 | | | | | labs |
| **t4free_count** | Free Thyroxine (Count) | continuous | Count of free thyroxine level measurements for this patient | Number of measurements | Labs | 0 | | | | | labs |
| **t4free_first** | Free Thyroxine (First) | continuous | First free thyroxine level measurement for this patient's hospitalization | ng/dl | Labs | 0 | | | | | labs |
| **t4free_last** | Free Thyroxine (Last) | continuous | Last free thyroxine level measurement for this patient's hospitalization | ng/dl | Labs | 0 | | | | | labs |
| **t4free_max** | Free Thyroxine (Max) | continuous | Maximum free thyroxine level measurement for this patient's hospitalization | ng/dl | Labs | 0 | | | | | labs |
| **t4free_mean** | Free Thyroxine (Mean) | continuous | Average free thyroxine level measurement for this patient's hospitalization | ng/dl | Labs | 0 | | | | | labs |
| **t4free_median** | Free Thyroxine (Median) | continuous | Median free thyroxine level measurement for this patient's hospitalization | ng/dl | Labs | 0 | | | | | labs |
| **t4free_min** | Free Thyroxine (Min) | continuous | Minimum free thyroxine level measurement for this patient's hospitalization | ng/dl | Labs | 0 | | | | | labs |
| **t4free_sd** | Free Thyroxine (SD) | continuous | Standard deviation of the free thyroxine level measurements during this patient's hospitalization | ng/dl | Labs | 0 | | | | | labs |
| **t4total_25** | Total Thyroxine (25th Percentile) | continuous | Total thyroxine level at the 25th percentile of this patient's measurements | ?g/dL | Labs | 0 | | | | | labs |
| **t4total_75** | Total Thyroxine (75th Percentile) | continuous | Total thyroxine level at the 75th percentile of this patient's measurements | ?g/dL | Labs | 0 | | | | | labs |
| **t4total_count** | Total Thyroxine (Count) | continuous | Count of total thyroxine level measurements for this patient | Number of measurements | Labs | 0 | | | | | labs |
| **t4total_first** | Total Thyroxine (First) | continuous | First total thyroxine level measurement for this patient's hospitalization | ?g/dL | Labs | 0 | | | | | labs |
| **t4total_last** | Total Thyroxine (Last) | continuous | Last total thyroxine level measurement for this patient's hospitalization | ?g/dL | Labs | 0 | | | | | labs |
| **t4total_max** | Total Thyroxine (Max) | continuous | Maximum total thyroxine level measurement for this patient's hospitalization | ?g/dL | Labs | 0 | | | | | labs |
| **t4total_mean** | Total Thyroxine (Mean) | continuous | Average total thyroxine level measurement for this patient's hospitalization | ?g/dL | Labs | 0 | | | | | labs |
| **t4total_median** | Total Thyroxine (Median) | continuous | Median total thyroxine level measurement for this patient's hospitalization | ?g/dL | Labs | 0 | | | | | labs |
| **t4total_min** | Total Thyroxine (Min) | continuous | Minimum total thyroxine level measurement for this patient's hospitalization | ?g/dL | Labs | 0 | | | | | labs |
| **t4total_sd** | Total Thyroxine (SD) | continuous | Standard deviation of the total thyroxine level measurements during this patient's hospitalization | ?g/dL | Labs | 0 | | | | | labs |
| **tacrolevel_25** | Tacrolimus Level (25th Percentile) | continuous | Tacrolimus level at the 25th percentile of this patient's measurements | ng/mL | Labs | 0 | | | | | labs |
| **tacrolevel_75** | Tacrolimus Level (75th Percentile) | continuous | Tacrolimus level at the 75th percentile of this patient's measurements | ng/mL | Labs | 0 | | | | | labs |
| **tacrolevel_count** | Tacrolimus Level (Count) | continuous | Count of tacrolimus level measurements for this patient | Number of measurements | Labs | 0 | | | | | labs |
| **tacrolevel_first** | Tacrolimus Level (First) | continuous | First tacrolimus level measurement for this patient's hospitalization | ng/mL | Labs | 0 | | | | | labs |
| **tacrolevel_last** | Tacrolimus Level (Last) | continuous | Last tacrolimus level measurement for this patient's hospitalization | ng/mL | Labs | 0 | | | | | labs |
| **tacrolevel_max** | Tacrolimus Level (Max) | continuous | Maximum tacrolimus level measurement for this patient's hospitalization | ng/mL | Labs | 0 | | | | | labs |
| **tacrolevel_mean** | Tacrolimus Level (Mean) | continuous | Average tacrolimus level measurement for this patient's hospitalization | ng/mL | Labs | 0 | | | | | labs |
| **tacrolevel_median** | Tacrolimus Level (Median) | continuous | Median tacrolimus level measurement for this patient's hospitalization | ng/mL | Labs | 0 | | | | | labs |
| **tacrolevel_min** | Tacrolimus Level (Min) | continuous | Minimum tacrolimus level measurement for this patient's hospitalization | ng/mL | Labs | 0 | | | | | labs |
| **tacrolevel_sd** | Tacrolimus Level (SD) | continuous | Standard deviation of the tacrolimus level measurements during this patient's hospitalization | ng/mL | Labs | 0 | | | | | labs |
| **temp_25** | Temperature (25th Percentile) | continuous | Temperature level at the 25th percentile of this patient's measurements | °C | Labs | 0 | | | | | labs |
| **temp_75** | Temperature (75th Percentile) | continuous | Temperature level at the 75th percentile of this patient's measurements | °C | Labs | 0 | | | | | labs |
| **temp_count** | Temperature (Count) | continuous | Count of temperature measurements for this patient | Number of measurements | Labs | 0 | | | | | labs |
| **temp_first** | Temperature (First) | continuous | First temperature measurement for this patient's hospitalization | °C | Labs | 0 | | | | | labs |
| **temp_last** | Temperature (Last) | continuous | Last temperature measurement for this patient's hospitalization | °C | Labs | 0 | | | | | labs |
| **temp_max** | Temperature (Max) | continuous | Maximum temperature measurement for this patient's hospitalization | °C | Labs | 0 | | | | | labs |
| **temp_mean** | Temperature (Mean) | continuous | Average temperature measurement for this patient's hospitalization | °C | Labs | 0 | | | | | labs |
| **temp_median** | Temperature (Median) | continuous | Median temperature measurement for this patient's hospitalization | °C | Labs | 0 | | | | | labs |
| **temp_min** | Temperature (Min) | continuous | Minimum temperature measurement for this patient's hospitalization | °C | Labs | 0 | | | | | labs |
| **temp_sd** | Temperature (SD) | continuous | Standard deviation of the temperature measurements during this patient's hospitalization | °C | Labs | 0 | | | | | labs |
| **thrombintime_25** | Thrombin Time (25th Percentile) | continuous | Thrombin time at the 25th percentile of this patient's measurements | Seconds | Labs | 0 | | | | | labs |
| **thrombintime_75** | Thrombin Time (75th Percentile) | continuous | Thrombin time at the 75th percentile of this patient's measurements | Seconds | Labs | 0 | | | | | labs |
| **thrombintime_count** | Thrombin Time (Count) | continuous | Count of thrombin time measurements for this patient | Number of measurements | Labs | 0 | | | | | labs |
| **thrombintime_first** | Thrombin Time (First) | continuous | First thrombin time measurement for this patient's hospitalization | Seconds | Labs | 0 | | | | | labs |
| **thrombintime_last** | Thrombin Time (Last) | continuous | Last thrombin time measurement for this patient's hospitalization | Seconds | Labs | 0 | | | | | labs |
| **thrombintime_max** | Thrombin Time (Max) | continuous | Maximum thrombin time measurement for this patient's hospitalization | Seconds | Labs | 0 | | | | | labs |
| **thrombintime_mean** | Thrombin Time (Mean) | continuous | Average thrombin time measurement for this patient's hospitalization | Seconds | Labs | 0 | | | | | labs |
| **thrombintime_median** | Thrombin Time (Median) | continuous | Median thrombin time measurement for this patient's hospitalization | Seconds | Labs | 0 | | | | | labs |
| **thrombintime_min** | Thrombin Time (Min) | continuous | Minimum thrombin time measurement for this patient's hospitalization | Seconds | Labs | 0 | | | | | labs |
| **thrombintime_sd** | Thrombin Time (SD) | continuous | Standard deviation of the thrombin time measurements during this patient's hospitalization | Seconds | Labs | 0 | | | | | labs |
| **tibc_25** | Total Iron Binding Capacity (25th Percentile) | continuous | Total iron binding capacity at the 25th percentile of this patient's measurements | mcg/dL | Labs | 0 | | | | | labs |
| **tibc_75** | Total Iron Binding Capacity (75th Percentile) | continuous | Total iron binding capacity at the 75th percentile of this patient's measurements | mcg/dL | Labs | 0 | | | | | labs |
| **tibc_count** | Total Iron Binding Capacity (Count) | continuous | Count of total iron binding capacity measurements for this patient | Number of measurements | Labs | 0 | | | | | labs |
| **tibc_first** | Total Iron Binding Capacity (First) | continuous | First total iron binding capacity measurement for this patient's hospitalization | mcg/dL | Labs | 0 | | | | | labs |
| **tibc_last** | Total Iron Binding Capacity (Last) | continuous | Last total iron binding capacity measurement for this patient's hospitalization | mcg/dL | Labs | 0 | | | | | labs |
| **tibc_max** | Total Iron Binding Capacity (Max) | continuous | Maximum total iron binding capacity measurement for this patient's hospitalization | mcg/dL | Labs | 0 | | | | | labs |
| **tibc_mean** | Total Iron Binding Capacity (Mean) | continuous | Average total iron binding capacity measurement for this patient's hospitalization | mcg/dL | Labs | 0 | | | | | labs |
| **tibc_median** | Total Iron Binding Capacity (Median) | continuous | Median total iron binding capacity measurement for this patient's hospitalization | mcg/dL | Labs | 0 | | | | | labs |
| **tibc_min** | Total Iron Binding Capacity (Min) | continuous | Minimum total iron binding capacity measurement for this patient's hospitalization | mcg/dL | Labs | 0 | | | | | labs |
| **tibc_sd** | Total Iron Binding Capacity (SD) | continuous | Standard deviation of the total iron binding capacity measurements during this patient's hospitalization | mcg/dL | Labs | 0 | | | | | labs |
| **tidalvolume_25** | Tidal Volume (25th Percentile) | continuous | Tidal volume at the 25th percentile of this patient's measurements | mL/inspiration | Labs | 0 | | | | | labs |
| **tidalvolume_75** | Tidal Volume (75th Percentile) | continuous | Tidal volume at the 75th percentile of this patient's measurements | mL/inspiration | Labs | 0 | | | | | labs |
| **tidalvolume_count** | Tidal Volume (Count) | continuous | Count of tidal volume measurements for this patient | Number of measurements | Labs | 0 | | | | | labs |

| | | | | | | | | | | | |
|---|---|---|---|---|---|---|---|---|---|---|---|
| **tidalvolume_first** | Tidal Volume (First) | continuous | First tidal volume measurement for this patient's hospitalization | mL/inspiration | Labs | 0 | | | | | labs |
| **tidalvolume_last** | Tidal Volume (Last) | continuous | Last tidal volume measurement for this patient's hospitalization | mL/inspiration | Labs | 0 | | | | | labs |
| **tidalvolume_max** | Tidal Volume (Max) | continuous | Maximum tidal volume measurement for this patient's hospitalization | mL/inspiration | Labs | 0 | | | | | labs |
| **tidalvolume_mean** | Tidal Volume (Mean) | continuous | Average tidal volume measurement for this patient's hospitalization | mL/inspiration | Labs | 0 | | | | | labs |
| **tidalvolume_median** | Tidal Volume (Median) | continuous | Median tidal volume measurement for this patient's hospitalization | mL/inspiration | Labs | 0 | | | | | labs |
| **tidalvolume_min** | Tidal Volume (Min) | continuous | Minimum tidal volume measurement for this patient's hospitalization | mL/inspiration | Labs | 0 | | | | | labs |
| **tidalvolume_sd** | Tidal Volume (SD) | continuous | Standard deviation of the tidal volume measurements during this patient's hospitalization | mL/inspiration | Labs | 0 | | | | | labs |
| **time_to_first_pos_covid** | Time To First Positive COVID test | continuous | Time since the patient's first positive COVID-19 test | days | Labs | 0 | | | | | labs |
| **timedurinecreatinine_25** | Creatinine Timed Urine (25th Percentile) | continuous | Creatinine timed urine value at the 25th percentile of this patient's measurements | | Labs | 0 | | | | | labs |
| **timedurinecreatinine_75** | Creatinine Timed Urine (75th Percentile) | continuous | Creatinine timed urine value at the 75th percentile of this patient's measurements | | Labs | 0 | | | | | labs |
| **timedurinecreatinine_co** | Creatinine Timed Urine (Count) | continuous | Count of creatinine timed urine measurements for this patient | Number of measurements | Labs | 0 | | | | | labs |
| **timedurinecreatinine_fir** | Creatinine Timed Urine (First) | continuous | First creatinine timed urine measurement for this patient's hospitalization | | Labs | 0 | | | | | labs |
| **timedurinecreatinine_las** | Creatinine Timed Urine (Last) | continuous | Last creatinine timed urine measurement for this patient's hospitalization | | Labs | 0 | | | | | labs |
| **timedurinecreatinine_ma** | Creatinine Timed Urine (Max) | continuous | Maximum creatinine timed urine measurement for this patient's hospitalization | | Labs | 0 | | | | | labs |
| **timedurinecreatinine_me** | Creatinine Timed Urine (Mean) | continuous | Average creatinine timed urine measurement for this patient's hospitalization | | Labs | 0 | | | | | labs |
| **timedurinecreatinine_me** | Creatinine Timed Urine (Median) | continuous | Median creatinine timed urine measurement for this patient's hospitalization | | Labs | 0 | | | | | labs |
| **timedurinecreatinine_mi** | Creatinine Timed Urine (Min) | continuous | Minimum creatinine timed urine measurement for this patient's hospitalization | | Labs | 0 | | | | | labs |
| **timedurinecreatinine_sd** | Creatinine Timed Urine (SD) | continuous | Standard deviation of the y measurements during this patient's hospitalization | | Labs | 0 | | | | | labs |
| **timedurineduration_25** | Timed Urine Duration (25th Percentile) | continuous | Timed Urine Duration at the 25th percentile of this patient's measurements | sec. | Labs | 0 | | | | | labs |
| **timedurineduration_75** | Timed Urine Duration (75th Percentile) | continuous | Timed Urine Duration at the 75th percentile of this patient's measurements | sec. | Labs | 0 | | | | | labs |
| **timedurineduration_cou** | Timed Urine Duration (Count) | continuous | Count of Timed Urine Duration measurements for this patient | Number of measurements | Labs | 0 | | | | | labs |
| **timedurineduration_first** | Timed Urine Duration (First) | continuous | First Timed Urine Duration measurement for this patient's hospitalization | sec. | Labs | 0 | | | | | labs |
| **timedurineduration_last** | Timed Urine Duration (Last) | continuous | Last Timed Urine Duration measurement for this patient's hospitalization | sec. | Labs | 0 | | | | | labs |
| **timedurineduration_max** | Timed Urine Duration (Max) | continuous | Maximum Timed Urine Duration measurement for this patient's hospitalization | sec. | Labs | 0 | | | | | labs |
| **timedurineduration_mea** | Timed Urine Duration (Mean) | continuous | Average Timed Urine Duration measurement for this patient's hospitalization | sec. | Labs | 0 | | | | | labs |
| **timedurineduration_med** | Timed Urine Duration (Median) | continuous | Median Timed Urine Duration measurement for this patient's hospitalization | sec. | Labs | 0 | | | | | labs |
| **timedurineduration_min** | Timed Urine Duration (Min) | continuous | Minimum Timed Urine Duration measurement for this patient's hospitalization | sec. | Labs | 0 | | | | | labs |
| **timedurineduration_sd** | Timed Urine Duration (SD) | continuous | Standard deviation of the Timed Urine Duration measurements during this patient's hospitalization | sec. | Labs | 0 | | | | | labs |
| **tnfalpha_25** | Tumour Necrosis Factor Alpha (25th Percentile) | continuous | TNF alpha level at the 25th percentile of this patient's measurements | pg/ml | Labs | 0 | | | | | labs |
| **tnfalpha_75** | Tumour Necrosis Factor Alpha (75th Percentile) | continuous | TNF alpha level at the 75th percentile of this patient's measurements | pg/ml | Labs | 0 | | | | | labs |
| **tnfalpha_count** | Tumour Necrosis Factor Alpha (Count) | continuous | Count of TNF alpha level measurements for this patient | Number of measurements | Labs | 0 | | | | | labs |
| **tnfalpha_first** | Tumour Necrosis Factor Alpha (First) | continuous | First TNF alpha level measurement for this patient's hospitalization | pg/ml | Labs | 0 | | | | | labs |
| **tnfalpha_last** | Tumour Necrosis Factor Alpha (Last) | continuous | Last TNF alpha level measurement for this patient's hospitalization | pg/ml | Labs | 0 | | | | | labs |
| **tnfalpha_max** | Tumour Necrosis Factor Alpha (Max) | continuous | Maximum TNF alpha level measurement for this patient's hospitalization | pg/ml | Labs | 0 | | | | | labs |
| **tnfalpha_mean** | Tumour Necrosis Factor Alpha (Mean) | continuous | Average TNF alpha level measurement for this patient's hospitalization | pg/ml | Labs | 0 | | | | | labs |
| **tnfalpha_median** | Tumour Necrosis Factor Alpha (Median) | continuous | Median TNF alpha level measurement for this patient's hospitalization | pg/ml | Labs | 0 | | | | | labs |
| **tnfalpha_min** | Tumour Necrosis Factor Alpha (Min) | continuous | Minimum TNF alpha level measurement for this patient's hospitalization | pg/ml | Labs | 0 | | | | | labs |
| **tnfalpha_sd** | Tumour Necrosis Factor Alpha (SD) | continuous | Standard deviation of the TNF alpha level measurements during this patient's hospitalization | pg/ml | Labs | 0 | | | | | labs |
| **tobramycin_lev_first** | Tobramycin Level (First) | categorical | First Tobramycin level measurement for this patient's hospitalization | | Labs | 0 | | | | | |
| **tobramycin_lev_last** | Tobramycin Level (Last) | categorical | Last Tobramycin level measurement for this patient's hospitalization | | Labs | 0 | | | | | |
| **totalurine_25** | Total Urine (25th Percentile) | continuous | Total urine level at the 25th percentile of this patient's measurements | milliliters/day | Labs | 0 | | | | | labs |
| **totalurine_75** | Total Urine (75th Percentile) | continuous | Total urine level at the 75th percentile of this patient's measurements | milliliters/day | Labs | 0 | | | | | labs |
| **totalurine_count** | Total Urine (Count) | continuous | Count of total urine measurements for this patient | Number of measurements | Labs | 0 | | | | | labs |
| **totalurine_first** | Total Urine (First) | continuous | First total urine measurement for this patient's hospitalization | milliliters/day | Labs | 0 | | | | | labs |
| **totalurine_last** | Total Urine (Last) | continuous | Last total urine measurement for this patient's hospitalization | milliliters/day | Labs | 0 | | | | | labs |
| **totalurine_max** | Total Urine (Max) | continuous | Maximum total urine measurement for this patient's hospitalization | milliliters/day | Labs | 0 | | | | | labs |
| **totalurine_mean** | Total Urine (Mean) | continuous | Average total urine measurement for this patient's hospitalization | milliliters/day | Labs | 0 | | | | | labs |
| **totalurine_median** | Total Urine (Median) | continuous | Median total urine measurement for this patient's hospitalization | milliliters/day | Labs | 0 | | | | | labs |
| **totalurine_min** | Total Urine (Min) | continuous | Minimum total urine measurement for this patient's hospitalization | milliliters/day | Labs | 0 | | | | | labs |
| **totalurine_sd** | Total Urine (SD) | continuous | Standard deviation of the total urine measurements during this patient's hospitalization | milliliters/day | Labs | 0 | | | | | labs |
| **triglycerides_25** | Triglycerides (25th Percentile) | continuous | Triglyceride level at the 25th percentile of this patient's measurements | mg/dL | Labs | 0 | | | | | labs |
| **triglycerides_75** | Triglycerides (75th Percentile) | continuous | Triglyceride level at the 75th percentile of this patient's measurements | mg/dL | Labs | 0 | | | | | labs |
| **triglycerides_count** | Triglycerides (Count) | continuous | Count of triglyceride level measurements for this patient | Number of measurements | Labs | 0 | | | | | labs |
| **triglycerides_first** | Triglycerides (First) | continuous | First triglyceride level measurement for this patient's hospitalization | mg/dL | Labs | 0 | | | | | labs |
| **triglycerides_last** | Triglycerides (Last) | continuous | Last triglyceride level measurement for this patient's hospitalization | mg/dL | Labs | 0 | | | | | labs |
| **triglycerides_max** | Triglycerides (Max) | continuous | Maximum triglyceride level measurement for this patient's hospitalization | mg/dL | Labs | 0 | | | | | labs |
| **triglycerides_mean** | Triglycerides (Mean) | continuous | Average triglyceride level measurement for this patient's hospitalization | mg/dL | Labs | 0 | | | | | labs |
| **triglycerides_median** | Triglycerides (Median) | continuous | Median triglyceride level measurement for this patient's hospitalization | mg/dL | Labs | 0 | | | | | labs |
| **triglycerides_min** | Triglycerides (Min) | continuous | Minimum triglyceride level measurement for this patient's hospitalization | mg/dL | Labs | 0 | | | | | labs |
| **triglycerides_sd** | Triglycerides (SD) | continuous | Standard deviation of the triglyceride level measurements during this patient's hospitalization | mg/dL | Labs | 0 | | | | | labs |
| **troponin_i_first** | | | | | Labs | 0 | | | | | |
| **troponin_i_last** | | | | | Labs | 0 | | | | | |
| **troponin_t_first** | | | | | Labs | 0 | | | | | |
| **troponin_t_last** | | | | | Labs | 0 | | | | | |
| **tsh_25** | Thyroid Stimulating Hormone (25th Percentile) | continuous | TSH level at the 25th percentile of this patient's measurements | mU/L | Labs | 0 | | | | | labs |
| **tsh_75** | Thyroid Stimulating Hormone (75th Percentile) | continuous | TSH level at the 75th percentile of this patient's measurements | mU/L | Labs | 0 | | | | | labs |
| **tsh_count** | Thyroid Stimulating Hormone (Count) | continuous | Count of TSH level measurements for this patient | Number of measurements | Labs | 0 | | | | | labs |
| **tsh_first** | Thyroid Stimulating Hormone (First) | continuous | First TSH level measurement for this patient's hospitalization | mU/L | Labs | 0 | | | | | labs |
| **tsh_last** | Thyroid Stimulating Hormone (Last) | continuous | Last TSH level measurement for this patient's hospitalization | mU/L | Labs | 0 | | | | | labs |
| **tsh_max** | Thyroid Stimulating Hormone (Max) | continuous | Maximum TSH level measurement for this patient's hospitalization | mU/L | Labs | 0 | | | | | labs |
| **tsh_mean** | Thyroid Stimulating Hormone (Mean) | continuous | Average TSH level measurement for this patient's hospitalization | mU/L | Labs | 0 | | | | | labs |
| **tsh_median** | Thyroid Stimulating Hormone (Median) | continuous | Median TSH level measurement for this patient's hospitalization | mU/L | Labs | 0 | | | | | labs |
| **tsh_min** | Thyroid Stimulating Hormone (Min) | continuous | Minimum TSH level measurement for this patient's hospitalization | mU/L | Labs | 0 | | | | | labs |
| **tsh_sd** | Thyroid Stimulating Hormone (SD) | continuous | Standard deviation of the TSH level measurements during this patient's hospitalization | mU/L | Labs | 0 | | | | | labs |
| **uaappearance_first** | Urinalysis Appearance (First) | categorical | Results of the first urinalysis appearance test during this patient's hospitalization | | Labs | 0 | | | | | labs |
| **uaappearance_last** | Urinalysis Appearance (Last) | categorical | Results of the last urinalysis appearance test during this patient's hospitalization | | Labs | 0 | | | | | labs |
| **uabacteria_first** | Urinalysis Bacteria (First) | categorical | Results of the first urinalysis bacteria test during this patient's hospitalization | | Labs | 0 | | | | | labs |
| **uabacteria_last** | Urinalysis Bacteria (Last) | categorical | Results of the last urinalysis bacteria test during this patient's hospitalization | | Labs | 0 | | | | | labs |
| **uabili_first** | Urinalysis Bilirubin (First) | categorical | Results of the first urinalysis bilirubin test during this patient's hospitalization | | Labs | 0 | | | | | labs |
| **uabili_last** | Urinalysis Bilirubin (Last) | categorical | Results of the last urinalysis bilirubin test during this patient's hospitalization | | Labs | 0 | | | | | labs |
| **uablood_first** | Urinalysis Blood (First) | categorical | Results of the first urinalysis blood test during this patient's hospitalization | | Labs | 0 | | | | | labs |
| **uablood_last** | Urinalysis Blood (Last) | categorical | Results of the last urinalysis blood test during this patient's hospitalization | | Labs | 0 | | | | | labs |
| **uacaoxalate_first** | Calcium Oxalate in Urine (First) | categorical | First Calcium Oxalate in Urine measurement for this patient's hospitalization | | Labs | 0 | | | | | |
| **uacaoxalate_last** | Calcium Oxalate in Urine (Last) | categorical | Last Calcium Oxalate in Urine measurement for this patient's hospitalization | | Labs | 0 | | | | | |
| **uacapyrophosphate_firs** | Urine Calcium Pyrophosphate (First) | categorical | Results of the first Urine calcium pyrophosphate test during this patient's hospitalization | | Labs | 0 | | | | | labs |
| **uacapyrophosphate_las** | Urine Calcium Pyrophosphate (Last) | categorical | Results of the last Urine calcium pyrophosphate test during this patient's hospitalization | | Labs | 0 | | | | | labs |
| **uaclarity_first** | Urinalysis Clarity (First) | categorical | Results of the first urinalysis clarity test during this patient's hospitalization | | Labs | 0 | | | | | labs |
| **uaclarity_last** | Urinalysis Clarity (Last) | categorical | Results of the last urinalysis clarity test during this patient's hospitalization | | Labs | 0 | | | | | labs |
| **uacolor_first** | Urinalysis color (First) | categorical | Results of the first urinalysis color test during this patient's hospitalization | | Labs | 0 | | | | | labs |
| **uacolor_last** | Urinalysis color (Last) | categorical | Results of the last urinalysis color test during this patient's hospitalization | | Labs | 0 | | | | | labs |
| **uaglucose_first** | Urinalysis Glucose (First) | categorical | Results of the first urinalysis glucose test during this patient's hospitalization | | Labs | 0 | | | | | labs |
| **uaglucose_last** | Urinalysis Glucose (Last) | categorical | Results of the last urinalysis glucose test during this patient's hospitalization | | Labs | 0 | | | | | labs |
| **uagrancasts_first** | Urinalysis Granular Casts (First) | categorical | Results of the first urinalysis granular casts test during this patient's hospitalization | | Labs | 0 | | | | | labs |
| **uagrancasts_last** | Urinalysis Granular Casts (Last) | categorical | Results of the last urinalysis granular casts test during this patient's hospitalization | | Labs | 0 | | | | | labs |
| **uahycasts_first** | Urinalysis Hyaline Casts (First) | categorical | Results of the first urinalysis hyaline casts test during this patient's hospitalization | | Labs | 0 | | | | | labs |
| **uahycasts_last** | Urinalysis Hyaline Casts (Last) | categorical | Results of the last urinalysis hyaline casts test during this patient's hospitalization | | Labs | 0 | | | | | labs |
| **uaketones_first** | Urinalysis Ketones (First) | categorical | Results of the first urinalysis ketones test during this patient's hospitalization | | Labs | 0 | | | | | labs |
| **uaketones_last** | Urinalysis Ketones (Last) | categorical | Results of the last urinalysis ketones test during this patient's hospitalization | | Labs | 0 | | | | | labs |
| **ualeukest_first** | Urinalysis Leukoesterase (First) | categorical | Results of the first urinalysis leukoesterase test during this patient's hospitalization | | Labs | 0 | | | | | labs |
| **ualeukest_last** | Urinalysis Leukoesterase (Last) | categorical | Results of the last urinalysis leukoesterase test during this patient's hospitalization | | Labs | 0 | | | | | labs |
| **uanitrite_first** | Urinalysis Nitrite (First) | categorical | Results of the first urinalysis nitrite test during this patient's hospitalization | | Labs | 0 | | | | | labs |
| **uanitrite_last** | Urinalysis Nitrite (Last) | categorical | Results of the last urinalysis nitrite test during this patient's hospitalization | | Labs | 0 | | | | | labs |
| **uaph_25** | Urinalysis pH (25th Percentile) | continuous | Urinalysis pH level at the 25th percentile of this patient's measurements | | Labs | 0 | | | | | labs |
| **uaph_75** | Urinalysis pH (75th Percentile) | continuous | Urinalysis pH level at the 75th percentile of this patient's measurements | | Labs | 0 | | | | | labs |
| **uaph_count** | Urinalysis pH (Count) | continuous | Count of urinalysis pH level measurements for this patient | Number of measurements | Labs | 0 | | | | | labs |
| **uaph_first** | Urinalysis pH (First) | continuous | First urinalysis pH level measurement for this patient's hospitalization | | Labs | 0 | | | | | labs |
| **uaph_last** | Urinalysis pH (Last) | continuous | Last urinalysis pH level measurement for this patient's hospitalization | | Labs | 0 | | | | | labs |
| **uaph_max** | Urinalysis pH (Max) | continuous | Maximum urinalysis pH level measurement for this patient's hospitalization | | Labs | 0 | | | | | labs |
| **uaph_mean** | Urinalysis pH (Mean) | continuous | Average urinalysis pH level measurement for this patient's hospitalization | | Labs | 0 | | | | | labs |
| **uaph_median** | Urinalysis pH (Median) | continuous | Median urinalysis pH level measurement for this patient's hospitalization | | Labs | 0 | | | | | labs |
| **uaph_min** | Urinalysis pH (Min) | continuous | Minimum urinalysis pH level measurement for this patient's hospitalization | | Labs | 0 | | | | | labs |
| **uaph_sd** | Urinalysis pH (SD) | continuous | Standard deviation of the urinalysis pH level measurements during this patient's hospitalization | | Labs | 0 | | | | | labs |
| **uaprotein_first** | Urinalysis Protein (First) | categorical | Results of the first urinalysis protein test during this patient's hospitalization | | Labs | 0 | | | | | labs |
| **uaprotein_last** | Urinalysis Protein (Last) | categorical | Results of the last urinalysis protein test during this patient's hospitalization | | Labs | 0 | | | | | labs |
| **uarbcs_first** | Urinalysis Red Blood Cells (First) | categorical | Results of the first urinalysis red blood cells test during this patient's hospitalization | | Labs | 0 | | | | | labs |
| **uarbcs_last** | Urinalysis Red Blood Cells (Last) | categorical | Results of the last urinalysis red blood cells test during this patient's hospitalization | | Labs | 0 | | | | | labs |
| **uarte_first** | Urinalysis Renal Tubular Epithelial Cells (First) | categorical | Results of the first urinalysis renal tubular epithelial cells test during this patient's hospitalization | | Labs | 0 | | | | | labs |
| **uarte_last** | Urinalysis Renal Tubular Epithelial Cells (Last) | categorical | Results of the last urinalysis renal tubular epithelial cells test during this patient's hospitalization | | Labs | 0 | | | | | labs |
| **uaspecgrav_25** | Urinalysis Specific Gravity (25th Percentile) | continuous | Urinalysis specific gravity level at the 25th percentile of this patient's measurements | | Labs | 0 | | | | | labs |
| **uaspecgrav_75** | Urinalysis Specific Gravity (75th Percentile) | continuous | Urinalysis specific gravity level at the 75th percentile of this patient's measurements | | Labs | 0 | | | | | labs |
| **uaspecgrav_count** | Urinalysis Specific Gravity (Count) | continuous | Count of urinalysis specific gravity level measurements for this patient | Number of measurements | Labs | 0 | | | | | labs |
| **uaspecgrav_first** | Urinalysis Specific Gravity (First) | continuous | First urinalysis specific gravity level measurement for this patient's hospitalization | | Labs | 0 | | | | | labs |
| **uaspecgrav_last** | Urinalysis Specific Gravity (Last) | continuous | Last urinalysis specific gravity level measurement for this patient's hospitalization | | Labs | 0 | | | | | labs |
| **uaspecgrav_max** | Urinalysis Specific Gravity (Max) | continuous | Maximum urinalysis specific gravity level measurement for this patient's hospitalization | | Labs | 0 | | | | | labs |

| | | | | | | | | | | | |
|---|---|---|---|---|---|---|---|---|---|---|---|
| uaspecgrav_mean | Urinalysis Specific Gravity (Mean) | continuous | Average urinalysis specific gravity level measurement for this patient's hospitalization | | Labs | 0 | | | | | labs |
| uaspecgrav_median | Urinalysis Specific Gravity (Median) | continuous | Median urinalysis specific gravity level measurement for this patient's hospitalization | | Labs | 0 | | | | | labs |
| uaspecgrav_min | Urinalysis Specific Gravity (Min) | continuous | Minimum urinalysis specific gravity level measurement for this patient's hospitalization | | Labs | 0 | | | | | labs |
| uaspecgrav_sd | Urinalysis Specific Gravity (SD) | continuous | Standard deviation of the urinalysis specific gravity level measurements during this patient's hospitalization | | Labs | 0 | | | | | labs |
| uauricacid_first | Urinalysis Uric Acid (First) | categorical | Results of the first urinalysis uric acid test during this patient's hospitalization | | Labs | 0 | | | | | labs |
| uauricacid_last | Urinalysis Uric Acid (Last) | categorical | Results of the last urinalysis uric acid test during this patient's hospitalization | | Labs | 0 | | | | | labs |
| uawbccast_first | Urinalysis White Blood Cell Casts (First) | categorical | Results of the first urinalysis white blood cell casts test during this patient's hospitalization | | Labs | 0 | | | | | labs |
| uawbccast_last | Urinalysis White Blood Cell Casts (Last) | categorical | Results of the last urinalysis white blood cell casts test during this patient's hospitalization | | Labs | 0 | | | | | labs |
| uawbcclumps_first | Urinalysis White Blood Cell Clumps (First) | categorical | Results of the first urinalysis white blood cell clumps test during this patient's hospitalization | | Labs | 0 | | | | | labs |
| uawbcclumps_last | Urinalysis White Blood Cell Clumps (Last) | categorical | Results of the last urinalysis white blood cell clumps test during this patient's hospitalization | | Labs | 0 | | | | | labs |
| uawbcs_first | Urinalysis White Blood Cells (First) | categorical | Results of the first urinalysis white blood cells test during this patient's hospitalization | | Labs | 0 | | | | | labs |
| uawbcs_last | Urinalysis White Blood Cells (Last) | categorical | Results of the last urinalysis white blood cells test during this patient's hospitalization | | Labs | 0 | | | | | labs |
| ucreat24h_25 | Urine Creatinine 24 hour (25th Percentile) | continuous | Urine creatinine level (24 hour) at the 25th percentile of this patient's measurements | mg/24 hours | Labs | 0 | | | | | labs |
| ucreat24h_75 | Urine Creatinine 24 hour (75th Percentile) | continuous | Urine creatinine level (24 hour) at the 75th percentile of this patient's measurements | mg/24 hours | Labs | 0 | | | | | labs |
| ucreat24h_count | Urine Creatinine 24 hour (Count) | continuous | Count of urine creatinine level (24 hour) measurements for this patient | Number of measurements | Labs | 0 | | | | | labs |
| ucreat24h_first | Urine Creatinine 24 hour (First) | continuous | First urine creatinine level (24 hour) measurement for this patient's hospitalization | mg/24 hours | Labs | 0 | | | | | labs |
| ucreat24h_last | Urine Creatinine 24 hour (Last) | continuous | Last urine creatinine level (24 hour) measurement for this patient's hospitalization | mg/24 hours | Labs | 0 | | | | | labs |
| ucreat24h_max | Urine Creatinine 24 hour (Max) | continuous | Maximum urine creatinine level (24 hour) measurement for this patient's hospitalization | mg/24 hours | Labs | 0 | | | | | labs |
| ucreat24h_mean | Urine Creatinine 24 hour (Mean) | continuous | Average urine creatinine level (24 hour) measurement for this patient's hospitalization | mg/24 hours | Labs | 0 | | | | | labs |
| ucreat24h_median | Urine Creatinine 24 hour (Median) | continuous | Median urine creatinine level (24 hour) measurement for this patient's hospitalization | mg/24 hours | Labs | 0 | | | | | labs |
| ucreat24h_min | Urine Creatinine 24 hour (Min) | continuous | Minimum urine creatinine level (24 hour) measurement for this patient's hospitalization | mg/24 hours | Labs | 0 | | | | | labs |
| ucreat24h_sd | Urine Creatinine 24 hour (SD) | continuous | Standard deviation of the urine creatinine level (24 hour) measurements during this patient's hospitalizati | mg/24 hours | Labs | 0 | | | | | labs |
| ucreatrandom_25 | Random Urine Creatinine (25th Percentile) | continuous | Random urine creatinine level at the 25th percentile of this patient's measurements | | Labs | 0 | | | | | labs |
| ucreatrandom_75 | Random Urine Creatinine (75th Percentile) | continuous | Random urine creatinine level at the 75th percentile of this patient's measurements | | Labs | 0 | | | | | labs |
| ucreatrandom_count | Random Urine Creatinine (Count) | continuous | Count of random urine creatinine level measurements for this patient | Number of measurements | Labs | 0 | | | | | labs |
| ucreatrandom_first | Random Urine Creatinine (First) | continuous | First random urine creatinine level measurement for this patient's hospitalization | | Labs | 0 | | | | | labs |
| ucreatrandom_last | Random Urine Creatinine (Last) | continuous | Last random urine creatinine level measurement for this patient's hospitalization | | Labs | 0 | | | | | labs |
| ucreatrandom_max | Random Urine Creatinine (Max) | continuous | Maximum random urine creatinine level measurement for this patient's hospitalization | | Labs | 0 | | | | | labs |
| ucreatrandom_mean | Random Urine Creatinine (Mean) | continuous | Average random urine creatinine level measurement for this patient's hospitalization | | Labs | 0 | | | | | labs |
| ucreatrandom_median | Random Urine Creatinine (Median) | continuous | Median random urine creatinine level measurement for this patient's hospitalization | | Labs | 0 | | | | | labs |
| ucreatrandom_min | Random Urine Creatinine (Min) | continuous | Minimum random urine creatinine level measurement for this patient's hospitalization | | Labs | 0 | | | | | labs |
| ucreatrandom_sd | Random Urine Creatinine (SD) | continuous | Standard deviation of the random urine creatinine level measurements during this patient's hospitalization | | Labs | 0 | | | | | labs |
| ukappa_25 | Urine Kappa (25th Percentile) | continuous | Kappa light chain (urine) at the 25th percentile of this patient's measurements | mg/L | Labs | 0 | | | | | labs |
| ukappa_75 | Urine Kappa (75th Percentile) | continuous | Kappa light chain (urine) at the 75th percentile of this patient's measurements | mg/L | Labs | 0 | | | | | labs |
| ukappa_count | Urine Kappa (Count) | continuous | Count of kappa light chain (urine) measurements for this patient | Number of measurements | Labs | 0 | | | | | labs |
| ukappa_first | Urine Kappa (First) | continuous | First kappa light chain (urine) measurement for this patient's hospitalization | mg/L | Labs | 0 | | | | | labs |
| ukappa_last | Urine Kappa (Last) | continuous | Last kappa light chain (urine) measurement for this patient's hospitalization | mg/L | Labs | 0 | | | | | labs |
| ukappa_max | Urine Kappa (Max) | continuous | Maximum kappa light chain (urine) measurement for this patient's hospitalization | mg/L | Labs | 0 | | | | | labs |
| ukappa_mean | Urine Kappa (Mean) | continuous | Average kappa light chain (urine) measurement for this patient's hospitalization | mg/L | Labs | 0 | | | | | labs |
| ukappa_median | Urine Kappa (Median) | continuous | Median kappa light chain (urine) measurement for this patient's hospitalization | mg/L | Labs | 0 | | | | | labs |
| ukappa_min | Urine Kappa (Min) | continuous | Minimum kappa light chain (urine) measurement for this patient's hospitalization | mg/L | Labs | 0 | | | | | labs |
| ukappa_sd | Urine Kappa (SD) | continuous | Standard deviation of the kappa light chain (urine) measurements during this patient's hospitalization | mg/L | Labs | 0 | | | | | labs |
| ulambda_25 | g (25th Percentile) | continuous | y at the 25th percentile of this patient's measurements | z | Labs | 0 | | | | | |
| ulambda_75 | g (75th Percentile) | continuous | y at the 75th percentile of this patient's measurements | z | Labs | 0 | | | | | |
| ulambda_count | g (Count) | continuous | Count of y measurements for this patient | Number of measurements | Labs | 0 | | | | | |
| ulambda_first | g (First) | continuous | First y measurement for this patient's hospitalization | z | Labs | 0 | | | | | |
| ulambda_last | g (Last) | continuous | Last y measurement for this patient's hospitalization | z | Labs | 0 | | | | | |
| ulambda_max | g (Max) | continuous | Maximum y measurement for this patient's hospitalization | z | Labs | 0 | | | | | |
| ulambda_mean | g (Mean) | continuous | Average y measurement for this patient's hospitalization | z | Labs | 0 | | | | | |
| ulambda_median | g (Median) | continuous | Median y measurement for this patient's hospitalization | z | Labs | 0 | | | | | |
| ulambda_min | g (Min) | continuous | Minimum y measurement for this patient's hospitalization | z | Labs | 0 | | | | | |
| ulambda_sd | g (SD) | continuous | Standard deviation of the y measurements during this patient's hospitalization | z | Labs | 0 | | | | | |
| upep_first | Urine Protein Electrophoresis (First) | categorical | Results of the first urine protein electrophoresis test during this patient's hospitalization | | Labs | 0 | | | | | labs |
| upep_last | Urine Protein Electrophoresis (Last) | categorical | Results of the last urine protein electrophoresis test during this patient's hospitalization | | Labs | 0 | | | | | labs |
| uprotein_25 | Urine Protein (25th Percentile) | continuous | Urine protein level at the 25th percentile of this patient's measurements | mg/dL | Labs | 0 | | | | | labs |
| uprotein_75 | Urine Protein (75th Percentile) | continuous | Urine protein level at the 75th percentile of this patient's measurements | mg/dL | Labs | 0 | | | | | labs |
| uprotein_count | Urine Protein (Count) | continuous | Count of urine protein level measurements for this patient | Number of measurements | Labs | 0 | | | | | labs |
| uprotein_first | Urine Protein (First) | continuous | First urine protein level measurement for this patient's hospitalization | mg/dL | Labs | 0 | | | | | labs |
| uprotein_last | Urine Protein (Last) | continuous | Last urine protein level measurement for this patient's hospitalization | mg/dL | Labs | 0 | | | | | labs |
| uprotein_max | Urine Protein (Max) | continuous | Maximum urine protein level measurement for this patient's hospitalization | mg/dL | Labs | 0 | | | | | labs |
| uprotein_mean | Urine Protein (Mean) | continuous | Average urine protein level measurement for this patient's hospitalization | mg/dL | Labs | 0 | | | | | labs |
| uprotein_median | Urine Protein (Median) | continuous | Median urine protein level measurement for this patient's hospitalization | mg/dL | Labs | 0 | | | | | labs |
| uprotein_min | Urine Protein (Min) | continuous | Minimum urine protein level measurement for this patient's hospitalization | mg/dL | Labs | 0 | | | | | labs |
| uprotein_sd | Urine Protein (SD) | continuous | Standard deviation of the urine protein level measurements during this patient's hospitalization | mg/dL | Labs | 0 | | | | | labs |
| uricacid_25 | Uric Acid (25th Percentile) | continuous | Uric acid level at the 25th percentile of this patient's measurements | mg/dL | Labs | 0 | | | | | labs |
| uricacid_75 | Uric Acid (75th Percentile) | continuous | Uric acid level at the 75th percentile of this patient's measurements | mg/dL | Labs | 0 | | | | | labs |
| uricacid_count | Uric Acid (Count) | continuous | Count of uric acid level measurements for this patient | Number of measurements | Labs | 0 | | | | | labs |
| uricacid_first | Uric Acid (First) | continuous | First uric acid level measurement for this patient's hospitalization | mg/dL | Labs | 0 | | | | | labs |
| uricacid_last | Uric Acid (Last) | continuous | Last uric acid level measurement for this patient's hospitalization | mg/dL | Labs | 0 | | | | | labs |
| uricacid_max | Uric Acid (Max) | continuous | Maximum uric acid level measurement for this patient's hospitalization | mg/dL | Labs | 0 | | | | | labs |
| uricacid_mean | Uric Acid (Mean) | continuous | Average uric acid level measurement for this patient's hospitalization | mg/dL | Labs | 0 | | | | | labs |
| uricacid_median | Uric Acid (Median) | continuous | Median uric acid level measurement for this patient's hospitalization | mg/dL | Labs | 0 | | | | | labs |
| uricacid_min | Uric Acid (Min) | continuous | Minimum uric acid level measurement for this patient's hospitalization | mg/dL | Labs | 0 | | | | | labs |
| uricacid_sd | Uric Acid (SD) | continuous | Standard deviation of the uric acid level measurements during this patient's hospitalization | mg/dL | Labs | 0 | | | | | labs |
| urinecalrandom_first | Random Urine Calcium (First) | categorical | Results of the first random urine calcium test during this patient's hospitalization | | Labs | 0 | | | | | labs |
| urinecalrandom_last | Random Urine Calcium (Last) | categorical | Results of the last random urine calcium test during this patient's hospitalization | | Labs | 0 | | | | | labs |
| urinecx_first | Urine Culture (First) | categorical | Results of the first urine culture test during this patient's hospitalization | | Labs | 0 | | | | | labs |
| urinecx_last | Urine Culture (Last) | categorical | Results of the last urine culture test during this patient's hospitalization | | Labs | 0 | | | | | labs |
| usodium_25 | Urine Sodium (25th Percentile) | continuous | Urine sodium level at the 25th percentile of this patient's measurements | mEq/day | Labs | 0 | | | | | labs |
| usodium_75 | Urine Sodium (75th Percentile) | continuous | Urine sodium level at the 75th percentile of this patient's measurements | mEq/day | Labs | 0 | | | | | labs |
| usodium_count | Urine Sodium (Count) | continuous | Count of urine sodium level measurements for this patient | Number of measurements | Labs | 0 | | | | | labs |
| usodium_first | Urine Sodium (First) | continuous | First urine sodium level measurement for this patient's hospitalization | mEq/day | Labs | 0 | | | | | labs |
| usodium_last | Urine Sodium (Last) | continuous | Last urine sodium level measurement for this patient's hospitalization | mEq/day | Labs | 0 | | | | | labs |
| usodium_max | Urine Sodium (Max) | continuous | Maximum urine sodium level measurement for this patient's hospitalization | mEq/day | Labs | 0 | | | | | labs |
| usodium_mean | Urine Sodium (Mean) | continuous | Average urine sodium level measurement for this patient's hospitalization | mEq/day | Labs | 0 | | | | | labs |
| usodium_median | Urine Sodium (Median) | continuous | Median urine sodium level measurement for this patient's hospitalization | mEq/day | Labs | 0 | | | | | labs |
| usodium_min | Urine Sodium (Min) | continuous | Minimum urine sodium level measurement for this patient's hospitalization | mEq/day | Labs | 0 | | | | | labs |
| usodium_sd | Urine Sodium (SD) | continuous | Standard deviation of the urine sodium level measurements during this patient's hospitalization | mEq/day | Labs | 0 | | | | | labs |
| uunrandom_25 | Random Urine Urea Nitrogen (25th Percentile) | continuous | Random UUN level at the 25th percentile of this patient's measurements | mmol/day | Labs | 0 | | | | | labs |
| uunrandom_75 | Random Urine Urea Nitrogen (75th Percentile) | continuous | Random UUN level at the 75th percentile of this patient's measurements | mmol/day | Labs | 0 | | | | | labs |
| uunrandom_count | Random Urine Urea Nitrogen (Count) | continuous | Count of random UUN level measurements for this patient | Number of measurements | Labs | 0 | | | | | labs |
| uunrandom_first | Random Urine Urea Nitrogen (First) | continuous | First random UUN level measurement for this patient's hospitalization | mmol/day | Labs | 0 | | | | | labs |
| uunrandom_last | Random Urine Urea Nitrogen (Last) | continuous | Last random UUN level measurement for this patient's hospitalization | mmol/day | Labs | 0 | | | | | labs |
| uunrandom_max | Random Urine Urea Nitrogen (Max) | continuous | Maximum random UUN level measurement for this patient's hospitalization | mmol/day | Labs | 0 | | | | | labs |
| uunrandom_mean | Random Urine Urea Nitrogen (Mean) | continuous | Average random UUN level measurement for this patient's hospitalization | mmol/day | Labs | 0 | | | | | labs |
| uunrandom_median | Random Urine Urea Nitrogen (Median) | continuous | Median random UUN level measurement for this patient's hospitalization | mmol/day | Labs | 0 | | | | | labs |
| uunrandom_min | Random Urine Urea Nitrogen (Min) | continuous | Minimum random UUN level measurement for this patient's hospitalization | mmol/day | Labs | 0 | | | | | labs |
| uunrandom_sd | Random Urine Urea Nitrogen (SD) | continuous | Standard deviation of the random UUN level measurements during this patient's hospitalization | mmol/day | Labs | 0 | | | | | labs |
| valproiclevel_25 | Valproic Acid Level (25th Percentile) | continuous | Valproic acid level at the 25th percentile of this patient's measurements | mcg/mL | Labs | 0 | | | | | labs |
| valproiclevel_75 | Valproic Acid Level (75th Percentile) | continuous | Valproic acid level at the 75th percentile of this patient's measurements | mcg/mL | Labs | 0 | | | | | labs |
| valproiclevel_count | Valproic Acid Level (Count) | continuous | Count of valproic acid level measurements for this patient | Number of measurements | Labs | 0 | | | | | labs |
| valproiclevel_first | Valproic Acid Level (First) | continuous | First valproic acid level measurement for this patient's hospitalization | mcg/mL | Labs | 0 | | | | | labs |
| valproiclevel_last | Valproic Acid Level (Last) | continuous | Last valproic acid level measurement for this patient's hospitalization | mcg/mL | Labs | 0 | | | | | labs |
| valproiclevel_max | Valproic Acid Level (Max) | continuous | Maximum valproic acid level measurement for this patient's hospitalization | mcg/mL | Labs | 0 | | | | | labs |
| valproiclevel_mean | Valproic Acid Level (Mean) | continuous | Average valproic acid level measurement for this patient's hospitalization | mcg/mL | Labs | 0 | | | | | labs |
| valproiclevel_median | Valproic Acid Level (Median) | continuous | Median valproic acid level measurement for this patient's hospitalization | mcg/mL | Labs | 0 | | | | | labs |
| valproiclevel_min | Valproic Acid Level (Min) | continuous | Minimum valproic acid level measurement for this patient's hospitalization | mcg/mL | Labs | 0 | | | | | labs |
| valproiclevel_sd | Valproic Acid Level (SD) | continuous | Standard deviation of the valproic acid level measurements during this patient's hospitalization | mcg/mL | Labs | 0 | | | | | labs |
| vancomycinlevel_25 | Vancomycin Level (25th Percentile) | continuous | Vancomycin level at the 25th percentile of this patient's measurements | mcg/mL | Labs | 0 | | | | | labs |
| vancomycinlevel_75 | Vancomycin Level (75th Percentile) | continuous | Vancomycin level at the 75th percentile of this patient's measurements | mcg/mL | Labs | 0 | | | | | labs |
| vancomycinlevel_count | Vancomycin Level (Count) | continuous | Count of vancomycin level measurements for this patient | Number of measurements | Labs | 0 | | | | | labs |
| vancomycinlevel_first | Vancomycin Level (First) | continuous | First vancomycin level measurement for this patient's hospitalization | mcg/mL | Labs | 0 | | | | | labs |
| vancomycinlevel_last | Vancomycin Level (Last) | continuous | Last vancomycin level measurement for this patient's hospitalization | mcg/mL | Labs | 0 | | | | | labs |
| vancomycinlevel_max | Vancomycin Level (Max) | continuous | Maximum vancomycin level measurement for this patient's hospitalization | mcg/mL | Labs | 0 | | | | | labs |
| vancomycinlevel_mean | Vancomycin Level (Mean) | continuous | Average vancomycin level measurement for this patient's hospitalization | mcg/mL | Labs | 0 | | | | | labs |
| vancomycinlevel_media | Vancomycin Level (Median) | continuous | Median vancomycin level measurement for this patient's hospitalization | mcg/mL | Labs | 0 | | | | | labs |
| vancomycinlevel_min | Vancomycin Level (Min) | continuous | Minimum vancomycin level measurement for this patient's hospitalization | mcg/mL | Labs | 0 | | | | | labs |
| vancomycinlevel_sd | Vancomycin Level (SD) | continuous | Standard deviation of the vancomycin level measurements during this patient's hospitalization | mcg/mL | Labs | 0 | | | | | labs |
| vancorandom_25 | Random Vancomycin (25th Percentile) | continuous | Random vancomycin level at the 25th percentile of this patient's measurements | mcg/mL | Labs | 0 | | | | | labs |
| vancorandom_75 | Random Vancomycin (75th Percentile) | continuous | Random vancomycin level at the 75th percentile of this patient's measurements | mcg/mL | Labs | 0 | | | | | labs |
| vancorandom_count | Random Vancomycin (Count) | continuous | Count of random vancomycin level measurements for this patient | Number of measurements | Labs | 0 | | | | | labs |
| vancorandom_first | Random Vancomycin (First) | continuous | First random vancomycin level measurement for this patient's hospitalization | mcg/mL | Labs | 0 | | | | | labs |
| vancorandom_last | Random Vancomycin (Last) | continuous | Last random vancomycin level measurement for this patient's hospitalization | mcg/mL | Labs | 0 | | | | | labs |
| vancorandom_max | Random Vancomycin (Max) | continuous | Maximum random vancomycin level measurement for this patient's hospitalization | mcg/mL | Labs | 0 | | | | | labs |

| | | | | | | | | | | | |
|---|---|---|---|---|---|---|---|---|---|---|---|
| **vancorandom_mean** | Random Vancomycin (Mean) | continuous | Average random vancomycin level measurement for this patient's hospitalization | mcg/mL | Labs | 0 | | | | | labs |
| **vancorandom_median** | Random Vancomycin (Median) | continuous | Median random vancomycin level measurement for this patient's hospitalization | mcg/mL | Labs | 0 | | | | | labs |
| **vancorandom_min** | Random Vancomycin (Min) | continuous | Minimum random vancomycin level measurement for this patient's hospitalization | mcg/mL | Labs | 0 | | | | | labs |
| **vancorandom_sd** | Random Vancomycin (SD) | continuous | Standard deviation of the random vancomycin level measurements during this patient's hospitalization | mcg/mL | Labs | 0 | | | | | labs |
| **vancotrough_25** | Vancomycin Trough (25th Percentile) | continuous | vancomycin trough level at the 25th percentile of this patient's measurements | mcg/mL | Labs | 0 | | | | | labs |
| **vancotrough_75** | Vancomycin Trough (75th Percentile) | continuous | vancomycin trough level at the 75th percentile of this patient's measurements | mcg/mL | Labs | 0 | | | | | labs |
| **vancotrough_count** | Vancomycin Trough (Count) | continuous | Count of vancomycin trough level measurements for this patient | Number of measurements | Labs | 0 | | | | | labs |
| **vancotrough_first** | Vancomycin Trough (First) | continuous | First vancomycin trough level measurement for this patient's hospitalization | mcg/mL | Labs | 0 | | | | | labs |
| **vancotrough_last** | Vancomycin Trough (Last) | continuous | Last vancomycin trough level measurement for this patient's hospitalization | mcg/mL | Labs | 0 | | | | | labs |
| **vancotrough_max** | Vancomycin Trough (Max) | continuous | Maximum vancomycin trough level measurement for this patient's hospitalization | mcg/mL | Labs | 0 | | | | | labs |
| **vancotrough_mean** | Vancomycin Trough (Mean) | continuous | Average vancomycin trough level measurement for this patient's hospitalization | mcg/mL | Labs | 0 | | | | | labs |
| **vancotrough_median** | Vancomycin Trough (Median) | continuous | Median vancomycin trough level measurement for this patient's hospitalization | mcg/mL | Labs | 0 | | | | | labs |
| **vancotrough_min** | Vancomycin Trough (Min) | continuous | Minimum vancomycin trough level measurement for this patient's hospitalization | mcg/mL | Labs | 0 | | | | | labs |
| **vancotrough_sd** | Vancomycin Trough (SD) | continuous | Standard deviation of the vancomycin trough level measurements during this patient's hospitalization | mcg/mL | Labs | 0 | | | | | labs |
| **venousbicarb_25** | Venous Bicarbonate (25th Percentile) | continuous | Venous bicarbonate level at the 25th percentile of this patient's measurements | mmol/L | Labs | 0 | | | | | labs |
| **venousbicarb_75** | Venous Bicarbonate (75th Percentile) | continuous | Venous bicarbonate level at the 75th percentile of this patient's measurements | mmol/L | Labs | 0 | | | | | labs |
| **venousbicarb_count** | Venous Bicarbonate (Count) | continuous | Count of venous bicarbonate level measurements for this patient | Number of measurements | Labs | 0 | | | | | labs |
| **venousbicarb_first** | Venous Bicarbonate (First) | continuous | First venous bicarbonate level measurement for this patient's hospitalization | mmol/L | Labs | 0 | | | | | labs |
| **venousbicarb_last** | Venous Bicarbonate (Last) | continuous | Last venous bicarbonate level measurement for this patient's hospitalization | mmol/L | Labs | 0 | | | | | labs |
| **venousbicarb_max** | Venous Bicarbonate (Max) | continuous | Maximum venous bicarbonate level measurement for this patient's hospitalization | mmol/L | Labs | 0 | | | | | labs |
| **venousbicarb_mean** | Venous Bicarbonate (Mean) | continuous | Average venous bicarbonate level measurement for this patient's hospitalization | mmol/L | Labs | 0 | | | | | labs |
| **venousbicarb_median** | Venous Bicarbonate (Median) | continuous | Median venous bicarbonate level measurement for this patient's hospitalization | mmol/L | Labs | 0 | | | | | labs |
| **venousbicarb_min** | Venous Bicarbonate (Min) | continuous | Minimum venous bicarbonate level measurement for this patient's hospitalization | mmol/L | Labs | 0 | | | | | labs |
| **venousbicarb_sd** | Venous Bicarbonate (SD) | continuous | Standard deviation of the venous bicarbonate level measurements during this patient's hospitalization | mmol/L | Labs | 0 | | | | | labs |
| **vitamind125_25** | 1,25-Dihydroxyvitamin D (25th Percentile) | continuous | 1,25-Dihydroxyvitamin D level at the 25th percentile of this patient's measurements | ng/mL | Labs | 0 | | | | | labs |
| **vitamind125_75** | 1,25-Dihydroxyvitamin D (75th Percentile) | continuous | 1,25-Dihydroxyvitamin D level at the 75th percentile of this patient's measurements | ng/mL | Labs | 0 | | | | | labs |
| **vitamind125_count** | 1,25-Dihydroxyvitamin D (Count) | continuous | Count of 1,25-dihydroxyvitamin D level measurements for this patient | Number of measurements | Labs | 0 | | | | | labs |
| **vitamind125_first** | 1,25-Dihydroxyvitamin D (First) | continuous | First 1,25-dihydroxyvitamin D level measurement for this patient's hospitalization | ng/mL | Labs | 0 | | | | | labs |
| **vitamind125_last** | 1,25-Dihydroxyvitamin D (Last) | continuous | Last 1,25-dihydroxyvitamin D level measurement for this patient's hospitalization | ng/mL | Labs | 0 | | | | | labs |
| **vitamind125_max** | 1,25-Dihydroxyvitamin D (Max) | continuous | Maximum 1,25-dihydroxyvitamin D level measurement for this patient's hospitalization | ng/mL | Labs | 0 | | | | | labs |
| **vitamind125_mean** | 1,25-Dihydroxyvitamin D (Mean) | continuous | Average 1,25-dihydroxyvitamin D level measurement for this patient's hospitalization | ng/mL | Labs | 0 | | | | | labs |
| **vitamind125_median** | 1,25-Dihydroxyvitamin D (Median) | continuous | Median 1,25-dihydroxyvitamin D level measurement for this patient's hospitalization | ng/mL | Labs | 0 | | | | | labs |
| **vitamind125_min** | 1,25-Dihydroxyvitamin D (Min) | continuous | Minimum 1,25-dihydroxyvitamin D level measurement for this patient's hospitalization | ng/mL | Labs | 0 | | | | | labs |
| **vitamind125_sd** | 1,25-Dihydroxyvitamin D (SD) | continuous | Standard deviation of the 1,25-dihydroxyvitamin D level measurements during this patient's hospitalizatic | ng/mL | Labs | 0 | | | | | labs |
| **vitamind25_25** | 25-Hydroxyvitamin D (25th Percentile) | continuous | 25-Hydroxyvitamin D level at the 25th percentile of this patient's measurements | ng/mL | Labs | 0 | | | | | labs |
| **vitamind25_75** | 25-Hydroxyvitamin D (75th Percentile) | continuous | 25-Hydroxyvitamin D level at the 75th percentile of this patient's measurements | ng/mL | Labs | 0 | | | | | labs |
| **vitamind25_count** | 25-Hydroxyvitamin D (Count) | continuous | Count of 25-hydroxyvitamin D level measurements for this patient | Number of measurements | Labs | 0 | | | | | labs |
| **vitamind25_first** | 25-Hydroxyvitamin D (First) | continuous | First 25-hydroxyvitamin D level measurement for this patient's hospitalization | ng/mL | Labs | 0 | | | | | labs |
| **vitamind25_last** | 25-Hydroxyvitamin D (Last) | continuous | Last 25-hydroxyvitamin D level measurement for this patient's hospitalization | ng/mL | Labs | 0 | | | | | labs |
| **vitamind25_max** | 25-Hydroxyvitamin D (Max) | continuous | Maximum 25-hydroxyvitamin D level measurement for this patient's hospitalization | ng/mL | Labs | 0 | | | | | labs |
| **vitamind25_mean** | 25-Hydroxyvitamin D (Mean) | continuous | Average 25-hydroxyvitamin D level measurement for this patient's hospitalization | ng/mL | Labs | 0 | | | | | labs |
| **vitamind25_median** | 25-Hydroxyvitamin D (Median) | continuous | Median 25-hydroxyvitamin D level measurement for this patient's hospitalization | ng/mL | Labs | 0 | | | | | labs |
| **vitamind25_min** | 25-Hydroxyvitamin D (Min) | continuous | Minimum 25-hydroxyvitamin D level measurement for this patient's hospitalization | ng/mL | Labs | 0 | | | | | labs |
| **vitamind25_sd** | 25-Hydroxyvitamin D (SD) | continuous | Standard deviation of the 25-hydroxyvitamin D level measurements during this patient's hospitalization | ng/mL | Labs | 0 | | | | | labs |
| **wbcc_25** | White Blood Cell Count (25th Percentile) | continuous | White blood cell count at the 25th percentile of this patient's measurements | WBCs/L | Labs | 0 | | | | | labs |
| **wbcc_75** | White Blood Cell Count (75th Percentile) | continuous | White blood cell count at the 75th percentile of this patient's measurements | WBCs/L | Labs | 0 | | | | | labs |
| **wbcc_count** | White Blood Cell Count (Count) | continuous | Count of white blood cell measurements for this patient | Number of measurements | Labs | 0 | | | | | labs |
| **wbcc_first** | White Blood Cell Count (First) | continuous | First white blood cell measurement for this patient's hospitalization | WBCs/L | Labs | 0 | | | | | labs |
| **wbcc_last** | White Blood Cell Count (Last) | continuous | Last white blood cell measurement for this patient's hospitalization | WBCs/L | Labs | 0 | | | | | labs |
| **wbcc_max** | White Blood Cell Count (Max) | continuous | Maximum white blood cell count measurement for this patient's hospitalization | WBCs/L | Labs | 0 | | | | | labs |
| **wbcc_mean** | White Blood Cell Count (Mean) | continuous | Average white blood cell count measurement for this patient's hospitalization | WBCs/L | Labs | 0 | | | | | labs |
| **wbcc_median** | White Blood Cell Count (Median) | continuous | Median white blood cell count measurement for this patient's hospitalization | WBCs/L | Labs | 0 | | | | | labs |
| **wbcc_min** | White Blood Cell Count (Min) | continuous | Minimum white blood cell count measurement for this patient's hospitalization | WBCs/L | Labs | 0 | | | | | labs |
| **wbcc_sd** | White Blood Cell Count (SD) | continuous | Standard deviation of the white blood cell count measurements during this patient's hospitalization | WBCs/L | Labs | 0 | | | | | labs |
| **abacavir** | Abacavir | categorical | Patient received abacavir during admission | | Medications | 0 | | | | | med |
| **acearbrenin** | Ace Arbrenin | categorical | Patient received Ace Arbrenin during admission | | Medications | 0 | | | | | med |
| **aceinhibitor** | Ace Inhibitor | categorical | Patient received Ace Inhibitor during admission | | Medications | 0 | | | | | med |
| **acetaminophenmed** | Acetaminophen Medication | categorical | Patient received Acetaminophen Medication during admission | | Medications | 0 | | | | | med |
| **acetazolamide** | Acetazolamide | categorical | Patient received Acetazolamide during admission | | Medications | 0 | | | | | med |
| **acetylcysteine** | Acetylcysteine | categorical | Patient received Acetylcysteine during admission | | Medications | 0 | | | | | med |
| **acyclovir** | Acyclovir | categorical | Patient received Acyclovir during admission | | Medications | 0 | | | | | med |
| **adalimumab** | Adalimumab | categorical | Patient received Adalimumab during admission | | Medications | 0 | | | | | |
| **adenosine** | Adenosine | categorical | Patient received Adenosine during admission | | Medications | 0 | | | | | med |
| **albuminmed** | Albuminmed | categorical | Patient received Albuminmed during admission | | Medications | 0 | | | | | med |
| **albuterol** | Albuterol | categorical | Patient received Albuterol during admission | | Medications | 0 | | | | | med |
| **allopurinol** | Allopurinol | categorical | Patient received Allopurinol during admission | | Medications | 0 | | | | | med |
| **alprazolam** | Alprazolam | categorical | Patient received Alprazolam during admission | | Medications | 0 | | | | | med |
| **alteplase** | Alteplase | categorical | Patient received Alteplase during admission | | Medications | 0 | | | | | med |
| **amantadine** | Amantadine | categorical | Patient received Amantadine during admission | | Medications | 0 | | | | | med |
| **amikacin** | Amikacin | categorical | Patient received Amikacin during admission | | Medications | 0 | | | | | med |
| **amikacin_ag** | Amikacin Antigen | categorical | Patient received Amikacin Antigen during the | | Medications | 0 | | | | | |
| **amiloride** | Amiloride | categorical | Patient received Amiloride during admission | | Medications | 0 | | | | | med |
| **aminocaproic** | Aminocaproic | categorical | Patient received Aminocaproic during admission | | Medications | 0 | | | | | med |
| **aminoglycoside** | Aminoglycoside | categorical | Patient received Aminoglycoside during admission | | Medications | 0 | | | | | med |
| **amiodarone** | Amiodarone | categorical | Patient received Amiodarone during admission | | Medications | 0 | | | | | med |
| **amitriptyline** | Amitriptyline | categorical | Patient received Amitriptyline during admission | | Medications | 0 | | | | | med |
| **amlodipine** | Amlodipine | categorical | Patient received Amlodipine during admission | | Medications | 0 | | | | | med |
| **amoxicillin** | Amoxicillin | categorical | Patient received Amoxicillin during admission | | Medications | 0 | | | | | med |
| **amphotericin** | Amphotericin | categorical | Patient received Amphotericin during admission | | Medications | 0 | | | | | med |
| **amphotericin_both** | Amphotericin (Both) | categorical | Patient received Amphotericin (Both) during admission | | Medications | 0 | | | | | med |
| **amphotericin_lipid** | Amphotericin B Lipid | categorical | Patient received Amphotericin B Lipid during admission | | Medications | 0 | | | | | med |
| **ampicillin** | Ampicillin | categorical | Patient received Ampicillin during admission | | Medications | 0 | | | | | med |
| **anidulafungin** | Anidulafungin | categorical | Patient received Anidulafungin during admission | | Medications | 0 | | | | | med |
| **antibiotic** | Antibiotic | categorical | Patient received an antibiotic during admission | | Medications | 0 | | | | | med |
| **antidepressants** | Antidepressants | categorical | Patient received Antidepressants during admission | | Medications | 0 | | | | | med |
| **antifungal** | Antifungal | categorical | Patient received Antifungal during admission | | Medications | 0 | | | | | med |
| **antifungal_all** | Antifungal (All) | categorical | Patient received Antifungal (All) during admission | | Medications | 0 | | | | | med |
| **arb** | Angiotensin II Receptor Blockers | categorical | Patient received Angiotensin II Receptor Blockers during admission | | Medications | 0 | | | | | med |
| **aripiprazole** | Aripiprazole | categorical | Patient received Aripiprazole during admission | | Medications | 0 | | | | | med |
| **aspirin** | Aspirin | categorical | Patient received Aspirin during admission | | Medications | 0 | | | | | med |
| **atazanavir** | Atazanavir | categorical | Patient received Atazanavir during admission | | Medications | 0 | | | | | med |
| **atenolol** | Atenolol | categorical | Patient received Atenolol during admission | | Medications | 0 | | | | | med |
| **atorvastatin** | Atorvastatin | categorical | Patient received Atorvastatin during admission | | Medications | 0 | | | | | med |
| **atovaquone** | Atovaquone | categorical | Patient received Atovaquone during admission | | Medications | 0 | | | | | med |
| **atropine** | Atropine | categorical | Patient received Atropine during admission | | Medications | 0 | | | | | med |
| **augmentin** | Augmentin | categorical | Patient received Augmentin during admission | | Medications | 0 | | | | | |
| **azacitidine** | Azacitidine | categorical | Patient received Azacitidine during admission | | Medications | 0 | | | | | med |
| **azathioprine** | Azathioprine | categorical | Patient received Azathioprine during admission | | Medications | 0 | | | | | med |
| **azithromycin** | Azithromycin | categorical | Patient received Azithromycin during admission | | Medications | 0 | | | | | med |
| **aztreonam** | Aztreonam | categorical | Patient received Aztreonam during admission | | Medications | 0 | | | | | med |
| **baclofen** | Baclofen | categorical | Patient received Baclofen during admission | | Medications | 0 | | | | | med |
| **bactrim** | Bactrim | categorical | Patient received Bactrim during admission | | Medications | 0 | | | | | med |
| **bendamustine** | Bendamustine | categorical | Patient received Bendamustine during admission | | Medications | 0 | | | | | med |
| **benztropine** | Benztropine | categorical | Patient received Benztropine during admission | | Medications | 0 | | | | | med |
| **betablocker** | Beta Blocker | categorical | Patient received Beta Blocker during admission | | Medications | 0 | | | | | med |
| **betalactam** | Beta Lactam | categorical | Patient received Beta Lactam during admission | | Medications | 0 | | | | | med |
| **bevacizumab** | Bevacizumab | categorical | Patient received Bevacizumab during admission | | Medications | 0 | | | | | med |
| **bicarbdrip** | Bicarbdrip | categorical | Patient received Bicarbdrip during admission | | Medications | 0 | | | | | med |
| **bicitra** | Bicitra | categorical | Patient received Bicitra during admission | | Medications | 0 | | | | | med |
| **bisacodyl** | Bisacodyl | categorical | Patient received Bisacodyl during admission | | Medications | 0 | | | | | med |
| **bleomycin** | Bleomycin | categorical | Patient received Bleomycin during admission | | Medications | 0 | | | | | med |
| **bortezomib** | Bortezomib | categorical | Patient received Bortezomib during admission | | Medications | 0 | | | | | med |
| **budesonide** | Budesonide | categorical | Patient received Budesonide during admission | | Medications | 0 | | | | | med |
| **bumetanide** | Bumetanide | categorical | Patient received Bumetanide during admission | | Medications | 0 | | | | | med |
| **buprenorphine** | Buprenorphine | categorical | Patient received Buprenorphine during admission | | Medications | 0 | | | | | med |
| **bupropion** | Bupropion | categorical | Patient received Bupropion during admission | | Medications | 0 | | | | | med |
| **buspirone** | Buspirone | categorical | Patient received Buspirone during admission | | Medications | 0 | | | | | med |
| **calcitriol** | Calcitriol | categorical | Patient received Calcitriol during admission | | Medications | 0 | | | | | med |
| **calciummed** | Calciummed | categorical | Patient received Calciummed during admission | | Medications | 0 | | | | | med |
| **captopril** | Captopril | categorical | Patient received Captopril during admission | | Medications | 0 | | | | | med |
| **carbamazepine** | Carbamazepine | categorical | Patient received Carbamazepine during admission | | Medications | 0 | | | | | med |
| **carboplatin** | Carboplatin | categorical | Patient received Carboplatin during admission | | Medications | 0 | | | | | med |
| **carvedilol** | Carvedilol | categorical | Patient received Carvedilol during admission | | Medications | 0 | | | | | med |
| **ccb** | Calcium Channel Blocker | categorical | Patient received Calcium Channel Blocker during admission | | Medications | 0 | | | | | med |

| | | | | | | | | | | | |
|---|---|---|---|---|---|---|---|---|---|---|---|
| **cefadroxil** | Cefadroxil | categorical | Patient received Cefadroxil during admission | | Medications | 0 | | | | | med |
| **cefazolin** | Cefazolin | categorical | Patient received Cefazolin during admission | | Medications | 0 | | | | | med |
| **cefdinir** | Cefdinir | categorical | Patient received Cefdinir during admission | | Medications | 0 | | | | | med |
| **cefepime** | Cefepime | categorical | Patient received Cefepime during admission | | Medications | 0 | | | | | med |
| **cefoxitin** | Cefoxitin | categorical | Patient received Cefoxitin during admission | | Medications | 0 | | | | | med |
| **cefpodoxime** | Cefpodoxime | categorical | Patient received Cefpodoxime during admission | | Medications | 0 | | | | | med |
| **ceftaroline** | Ceftaroline | categorical | Patient received Ceftaroline during admission | | Medications | 0 | | | | | med |
| **ceftazidime** | Ceftazidime | categorical | Patient received Ceftazidime during admission | | Medications | 0 | | | | | med |
| **ceftolozane** | Ceftolozane | categorical | Patient received Ceftolozane during admission | | Medications | 0 | | | | | med |
| **ceftriaxone** | Ceftriaxone | categorical | Patient received Ceftriaxone during admission | | Medications | 0 | | | | | med |
| **cefuroxime** | Cefuroxime | categorical | Patient received Cefuroxime during admission | | Medications | 0 | | | | | med |
| **celecoxib** | Celecoxib | categorical | Patient received Celecoxib during admission | | Medications | 0 | | | | | med |
| **cephalexin** | Cephalexin | categorical | Patient received Cephalexin during admission | | Medications | 0 | | | | | med |
| **chemo** | Chemotherapy | categorical | Patient received Chemotherapy during admission | | Medications | 0 | | | | | med |
| **chlorambucil** | Chlorambucil | categorical | Patient received Chlorambucil during admission | | Medications | 0 | | | | | med |
| **chloramphenicol** | Chloramphenicol | categorical | Patient received Chloramphenicol during admission | | Medications | 0 | | | | | med |
| **chlordiazepoxide** | Chlordiazepoxide | categorical | Patient received Chlordiazepoxide during admission | | Medications | 0 | | | | | med |
| **chlorpromazine** | Chlorpromazine | categorical | Patient received Chlorpromazine during admission | | Medications | 0 | | | | | med |
| **chlorthalidone** | Chlorthalidone | categorical | Patient received Chlorthalidone during admission | | Medications | 0 | | | | | med |
| **cinacalcet** | Cinacalcet | categorical | Patient received Cinacalcet during admission | | Medications | 0 | | | | | med |
| **ciprofloxacin** | Ciprofloxacin | categorical | Patient received Ciprofloxacin during admission | | Medications | 0 | | | | | med |
| **cisatracurium** | Cisatracurium | categorical | Patient received Cisatracurium during admission | | Medications | 0 | | | | | med |
| **cisplatin** | Cisplatin | categorical | Patient received Cisplatin during admission | | Medications | 0 | | | | | med |
| **citalopram** | Citalopram | categorical | Patient received Citalopram during admission | | Medications | 0 | | | | | med |
| **citrate** | Citrate | categorical | Patient received Citrate during admission | | Medications | 0 | | | | | med |
| **clarithromycin** | Clarithromycin | categorical | Patient received Clarithromycin during admission | | Medications | 0 | | | | | med |
| **clindamycin** | Clindamycin | categorical | Patient received Clindamycin during admission | | Medications | 0 | | | | | med |
| **clonazepam** | Clonazepam | categorical | Patient received Clonazepam during admission | | Medications | 0 | | | | | med |
| **clonidine** | Clonidine | categorical | Patient received Clonidine during admission | | Medications | 0 | | | | | med |
| **clopidogrel** | Clopidogrel | categorical | Patient received Clopidogrel during admission | | Medications | 0 | | | | | med |
| **clotrimazole** | Clotrimazole | categorical | Patient received Clotrimazole during admission | | Medications | 0 | | | | | med |
| **clozapine** | Clozapine | categorical | Patient received Clozapine during admission | | Medications | 0 | | | | | med |
| **codeine** | Codeine | categorical | Patient received Codeine during admission | | Medications | 0 | | | | | med |
| **colchicine** | Colchicine | categorical | Patient received Colchicine during admission | | Medications | 0 | | | | | med |
| **crystalloid** | Crystalloid | categorical | Patient received Crystalloid during admission | | Medications | 0 | | | | | med |
| **cyclobenzaprine** | Cyclobenzaprine | categorical | Patient received Cyclobenzaprine during admission | | Medications | 0 | | | | | med |
| **cyclophosphamide** | Cyclophosphamide | categorical | Patient received Cyclophosphamide during admission | | Medications | 0 | | | | | med |
| **cyclosporine** | Cyclosporine | categorical | Patient received Cyclosporine during admission | | Medications | 0 | | | | | med |
| **cytarabine** | Cytarabine | categorical | Patient received Cytarabine during admission | | Medications | 0 | | | | | med |
| **d50drip** | D50 Drip | categorical | Patient received D50 Drip during admission | | Medications | 0 | | | | | med |
| **d5halfdrip** | D5 Half Drip | categorical | Patient received D5 Half Drip during admission | | Medications | 0 | | | | | med |
| **d5lrdrip** | D5IR Drip | categorical | Patient received D5IR Drip during admission | | Medications | 0 | | | | | med |
| **d5nsdrip** | D5NS Drip | categorical | Patient received D5NS Drip during admission | | Medications | 0 | | | | | med |
| **d5wdrip** | D5W Drip | categorical | Patient received D5W Drip during admission | | Medications | 0 | | | | | med |
| **dapsone** | Dapsone | categorical | Patient received Dapsone during admission | | Medications | 0 | | | | | med |
| **daptomycin** | Daptomycin | categorical | Patient received Daptomycin during admission | | Medications | 0 | | | | | med |
| **darbepoetin** | Darbepoetin | categorical | Patient received Darbepoetin during admission | | Medications | 0 | | | | | med |
| **darunavir** | Darunavir | categorical | Patient received Darunavir during admission | | Medications | 0 | | | | | med |
| **desipramine** | Desipramine | categorical | Patient received Desipramine during admission | | Medications | 0 | | | | | med |
| **desmopressin** | Desmopressin | categorical | Patient received Desmopressin during admission | | Medications | 0 | | | | | med |
| **desvenlafaxine** | Desvenlafaxine | categorical | Patient received Desvenlafaxine during admission | | Medications | 0 | | | | | med |
| **dexamethasone** | Dexamethasone | categorical | Patient received Dexamethasone during admission | | Medications | 0 | | | | | med |
| **dexlansoprazole** | Dexlansoprazole | categorical | Patient received Dexlansoprazole during admission | | Medications | 0 | | | | | med |
| **dexmedetomidine** | Dexmedetomidine | categorical | Patient received Dexmedetomidine during admission | | Medications | 0 | | | | | med |
| **dialysisdrug** | Dialysis Drug | categorical | Patient received Dialysis Drug during admission | | Medications | 0 | | | | | med |
| **diatrizoate_meglumine** | Diatrizoate Meglumine | categorical | Patient received Diatrizoate Meglumine during admission | | Medications | 0 | | | | | med |
| **diazepam** | Diazepam | categorical | Patient received Diazepam during admission | | Medications | 0 | | | | | med |
| **diazoxide** | Diazoxide | categorical | Patient received Diazoxide during admission | | Medications | 0 | | | | | med |
| **dicloxacillin** | Dicloxacillin | categorical | Patient received Dicloxacillin during admission | | Medications | 0 | | | | | med |
| **digibind** | Digibind | categorical | Patient received Digibind during admission | | Medications | 0 | | | | | med |
| **digoxin** | Digoxin | categorical | Patient received Digoxin during admission | | Medications | 0 | | | | | med |
| **diltiazem** | Diltiazem | categorical | Patient received Diltiazem during admission | | Medications | 0 | | | | | med |
| **diphenhydramine** | Diphenhydramine | categorical | Patient received Diphenhydramine during admission | | Medications | 0 | | | | | med |
| **diuretic** | Diuretic | categorical | Patient received Diuretic during admission | | Medications | 0 | | | | | med |
| **dobutamine** | Dobutamine | categorical | Patient received Dobutamine during admission | | Medications | 0 | | | | | med |
| **docetaxel** | Docetaxel | categorical | Patient received Docetaxel during admission | | Medications | 0 | | | | | med |
| **docusate** | Docusate | categorical | Patient received Docusate during admission | | Medications | 0 | | | | | med |
| **dofetilide** | Dofetilide | categorical | Patient received Dofetilide during admission | | Medications | 0 | | | | | med |
| **dolutegravir** | Dolutegravir | categorical | Patient received Dolutegravir during admission | | Medications | 0 | | | | | med |
| **donepezil** | Donepezil | categorical | Patient received Donepezil during admission | | Medications | 0 | | | | | med |
| **dopamine** | Dopamine | categorical | Patient received Dopamine during admission | | Medications | 0 | | | | | med |
| **doxazosin** | Doxazosin | categorical | Patient received Doxazosin during admission | | Medications | 0 | | | | | med |
| **doxorubicin** | Doxorubicin | categorical | Patient received Doxorubicin during admission | | Medications | 0 | | | | | med |
| **doxycycline** | Doxycycline | categorical | Patient received Doxycycline during admission | | Medications | 0 | | | | | med |
| **dronabinol** | Dronabinol | categorical | Patient received Dronabinol during admission | | Medications | 0 | | | | | med |
| **duloxetine** | Duloxetine | categorical | Patient received Duloxetine during admission | | Medications | 0 | | | | | med |
| **empagliflozin** | Empagliflozin | categorical | Patient received Empagliflozin during admission | | Medications | 0 | | | | | med |
| **emtricitabine** | Emtricitabine | categorical | Patient received Emtricitabine during admission | | Medications | 0 | | | | | med |
| **enalapril** | Enalapril | categorical | Patient received Enalapril during admission | | Medications | 0 | | | | | med |
| **enalaprilat** | Enalaprilat | categorical | Patient received Enalaprilat during admission | | Medications | 0 | | | | | med |
| **enoxaparin** | Enoxaparin | categorical | Patient received Enoxaparin during admission | | Medications | 0 | | | | | med |
| **entecavir** | Entecavir | categorical | Patient received Entecavir during admission | | Medications | 0 | | | | | med |
| **entresto** | Entresto | categorical | Patient received Entestro during admission | | Medications | 0 | | | | | med |
| **ephedrine** | Ephedrine | categorical | Patient received Ephedrine during admission | | Medications | 0 | | | | | med |
| **epinephrine** | Epinephrine | categorical | Patient received Epinephrine during admission | | Medications | 0 | | | | | med |
| **eplerenone** | Eplerenone | categorical | Patient received Eplerenone during admission | | Medications | 0 | | | | | med |
| **epoetin** | Epoetin | categorical | Patient received Epoetin during admission | | Medications | 0 | | | | | |
| **ertapenem** | Ertapenem | categorical | Patient received Ertapenem during admission | | Medications | 0 | | | | | med |
| **erythromycin** | Erythromycin | categorical | Patient received Erythromycin during admission | | Medications | 0 | | | | | med |
| **escitalopram** | Escitalopram | categorical | Patient received Escitalopram during admission | | Medications | 0 | | | | | med |
| **esmolol** | Esmolol | categorical | Patient received Esmolol during admission | | Medications | 0 | | | | | med |
| **esomeprazole** | Esomeprazole | categorical | Patient received Esomeprazole during admission | | Medications | 0 | | | | | med |
| **estradiol** | Estradiol | categorical | Patient received Estradiol during admission | | Medications | 0 | | | | | med |
| **etanercept** | Etanercept | categorical | Patient received Etanercept during admission | | Medications | 0 | | | | | med |
| **ethacrynicacid** | Ethacrynicacid | categorical | Patient received Ethacrynicacid during admission | | Medications | 0 | | | | | med |
| **ethambutol** | Ethambutol | categorical | Patient received Ethambutol during admission | | Medications | 0 | | | | | med |
| **etoposide** | Etoposide | categorical | Patient received Etoposide during admission | | Medications | 0 | | | | | med |
| **everolimus** | Everolimus | categorical | Patient received Everolimus during admission | | Medications | 0 | | | | | med |
| **ezetimibe** | Ezetimibe | categorical | Patient received Ezetimibe during admission | | Medications | 0 | | | | | med |
| **famciclovir** | Famciclovir | categorical | Patient received Famciclovir during admission | | Medications | 0 | | | | | med |
| **famotidine** | Famotidine | categorical | Patient received Famotidine during admission | | Medications | 0 | | | | | med |
| **febuxostat** | Febuxostat | categorical | Patient received Febuxostat during admission | | Medications | 0 | | | | | med |
| **fenofibrate** | Fenofibrate | categorical | Patient received Fenofibrate during admission | | Medications | 0 | | | | | med |
| **fentanyl** | Fentanyl | categorical | Patient received Fentanyl during admission | | Medications | 0 | | | | | med |
| **finasteride** | Finasteride | categorical | Patient received Finasteride during admission | | Medications | 0 | | | | | med |
| **flecainide** | Flecainide | categorical | Patient received Flecainide during admission | | Medications | 0 | | | | | med |
| **fluconazole** | Fluconazole | categorical | Patient received Fluconazole during admission | | Medications | 0 | | | | | med |
| **fludrocortisone** | Fludrocortisone | categorical | Patient received Fludrocortisone during admission | | Medications | 0 | | | | | med |
| **fluoroquinolone** | Fluoroquinolone | categorical | Patient received Fluoroquinolone during admission | | Medications | 0 | | | | | med |
| **fluorouracil** | Fluorouracil | categorical | Patient received Fluorouracil during admission | | Medications | 0 | | | | | med |
| **fluoxetine** | Fluoxetine | categorical | Patient received Fluoxetine during admission | | Medications | 0 | | | | | med |
| **fluphenazine** | Fluphenazine | categorical | Patient received Fluphenazine during admission | | Medications | 0 | | | | | med |
| **folicacid** | Folicacid | categorical | Patient received Folicacid during admission | | Medications | 0 | | | | | med |
| **fondaparinux** | Fondaparinux | categorical | Patient received Fondaparinux during admission | | Medications | 0 | | | | | med |
| **foscarnet** | Foscarnet | categorical | Patient received Foscarnet during admission | | Medications | 0 | | | | | med |
| **fosfomycin** | Fosfomycin | categorical | Patient received Fosfomycin during admission | | Medications | 0 | | | | | med |
| **fosphenytoin** | Fosphenytoin | categorical | Patient received Fosphenytoin during admission | | Medications | 0 | | | | | med |
| **furosemide** | Furosemide | categorical | Patient received Furosemide during admission | | Medications | 0 | | | | | med |
| **gabapentin** | Gabapentin | categorical | Patient received Gabapentin during admission | | Medications | 0 | | | | | med |
| **ganciclovir** | Ganciclovir | categorical | Patient received Ganciclovir during admission | | Medications | 0 | | | | | med |
| **gemcitabine** | Gemcitabine | categorical | Patient received Gemcitabine during admission | | Medications | 0 | | | | | med |
| **gentamicin** | Gentamicin | categorical | Patient received Gentamicin during admission | | Medications | 0 | | | | | med |
| **gentamicin_ag** | Gentamicin Antigen | categorical | Patient received Gentamicin Antigen during admission | | Medications | 0 | | | | | |
| **glimepiride** | Glimepiride | categorical | Patient received Glimepiride during admission | | Medications | 0 | | | | | med |

| | | | | | | | | | | | |
|---|---|---|---|---|---|---|---|---|---|---|---|
| **glipizide** | Glipizide | categorical | Patient received Glipizide during admission | | Medications | 0 | | | | | med |
| **globulinmed** | Globulin Medication | categorical | Patient received Globulin Medication during admission | | Medications | 0 | | | | | med |
| **glucagon** | Glucagon | categorical | Patient received Glucagon during admission | | Medications | 0 | | | | | med |
| **guaifenesin** | Guaifenesin | categorical | Patient received Guaifenesin during admission | | Medications | 0 | | | | | med |
| **haart** | Highly Active Antiretroviral Therapy | categorical | Patient received Highly Active Antiretroviral Therapy during admission | | Medications | 0 | | | | | med |
| **halfsalinedrip** | Half Saline Drip | categorical | Patient received Half Saline Drip during admission | | Medications | 0 | | | | | med |
| **haloperidol** | Haloperidol | categorical | Patient received Haloperidol during admission | | Medications | 0 | | | | | med |
| **hctz** | Hydrochlorothiazide | categorical | Patient received Hydrochlorothiazide during admission | | Medications | 0 | | | | | med |
| **heparin** | Heparin | categorical | Patient received Heparin during admission | | Medications | 0 | | | | | med |
| **htsaline** | Hypertonic Saline | categorical | Patient received Hypertonic Saline during admission | | Medications | 0 | | | | | med |
| **hydralazine** | Hydralazine | categorical | Patient received Hydralazine during admission | | Medications | 0 | | | | | med |
| **hydrocodone** | Hydrocodone | categorical | Patient received Hydrocodone during admission | | Medications | 0 | | | | | med |
| **hydrocortisone** | Hydrocortisone | categorical | Patient received Hydrocortisone during admission | | Medications | 0 | | | | | med |
| **hydromorphone** | Hydromorphone | categorical | Patient received Hydromorphone during admission | | Medications | 0 | | | | | med |
| **hydroxychloroquine** | Hydroxychloroquine | categorical | Patient received Hydroxychloroquine during admission | | Medications | 0 | | | | | med |
| **hydroxyurea** | Hydroxyurea | categorical | Patient received Hydroxyurea during admission | | Medications | 0 | | | | | med |
| **hydroxyzine** | Hydroxyzine | categorical | Patient received Hydroxyzine during admission | | Medications | 0 | | | | | med |
| **ibuprofen** | Ibuprofen | categorical | Patient received Ibuprofen during admission | | Medications | 0 | | | | | med |
| **ifosfamide** | Ifosfamide | categorical | Patient received Ifosfamide during admission | | Medications | 0 | | | | | med |
| **imdur** | Imdur | categorical | Patient received Imdur during admission | | Medications | 0 | | | | | med |
| **immune** | Immunosuppressant Drugs | categorical | Patient received Immunosuppressant Drugs during admission | | Medications | 0 | | | | | med |
| **immunomodulators** | Immunomodulators | categorical | Patient received Immunomodulators during admission | | Medications | 0 | | | | | med |
| **indomethacin** | Indomethacin | categorical | Patient received Indomethacin during admission | | Medications | 0 | | | | | med |
| **infliximab** | Infliximab | categorical | Patient received Infliximab during admission | | Medications | 0 | | | | | med |
| **insulin** | Insulin | categorical | Patient received Insulin during admission | | Medications | 0 | | | | | med |
| **insulin_aspart** | Insulin Aspart | categorical | Patient received Insulin Aspart during admission | | Medications | 0 | | | | | |
| **insulin_aspart_sc_scale** | Subcutaneous Insulin Aspart | categorical | Patient received Subcutaneous Insulin Aspart during admission |  | Medications | 0 |  |  |  |  |  |
| **insulin_conc** | Concentrated Insulin | categorical | Patient received Concentrated Insulin during admission | | Medications | 0 | | | | | |
| **insulin_csii** | Insulin CSII | categorical | Patient received Insulin CSII during admission | | Medications | 0 | | | | | |
| **insulin_degludec** | Insulin Degludec | categorical | Patient received Insulin Degludec during admission | | Medications | 0 | | | | | |
| **insulin_degludec_lira** | Insulin Degludec Liraglutide | categorical | Patient received Insulin Degludec Liraglutide during admission | | Medications | 0 | | | | | |
| **insulin_degludec_lira_pe** | Insulin Degludec Liraglutide Pen | categorical | Patient received Insulin Degludec Liraglutide Pen during admission | | Medications | 0 | | | | | |
| **insulin_degludec_u100_** | Insulin Degludec u100 Pen | categorical | Patient received Insulin Degludec u100 Pen during admission | | Medications | 0 | | | | | |
| **insulin_detemir** | Insulin Detemir | categorical | Patient recevied Insulin Detemir during admission | | Medications | 0 | | | | | |
| **insulin_detemir_pen** | Insulin Detemir Pen | categorical | Patient received Insulin Detemir Pen during admission | | Medications | 0 | | | | | |
| **insulin_detemir_sc** | Insulin Detemir Pen Subcutaneous | categorical | Patient received Insulin Detemir Pen Subcutaneous during admission | | Medications | 0 | | | | | |
| **insulin_glargine** | Insuline Glargine | categorical | Patient received Insulin Glargine during admission | | Medications | 0 | | | | | |
| **insulin_glargine_sc** | Insuline Glargine Subcutaneous | categorical | Patient received Insuline Glargine Subcutaneous during admission | | Medications | 0 | | | | | |
| **insulin_glargine_u100_p** | Insuline Glargine u100 Pen | categorical | Patient receved Insuline Glargine u100 Pen during admission | | Medications | 0 | | | | | |
| **insulin_glargine_u300_p** | Insuline Glargine u300 Pen | categorical | Patient received Insuline Glargine u300 Pen during admission | | Medications | 0 | | | | | |
| **insulin_gtt** | Insulin Glucose Tolerance Test | categorical | Patient received Insulin Glucose Tolerance Test during admission | | Medications | 0 | | | | | |
| **insulin_lispro** | Insulin Lispro | categorical | Patient receieved Insulin Lispro during admission | | Medications | 0 | | | | | |
| **insulin_lispro_csii** | Insulin Lispro CSII | categorical | Patient receieved Insulin Lispro CSII during admission | | Medications | 0 | | | | | |
| **insulin_lispro_pump** | Insulin Lispro Pump | categorical | Patient received Insulin Lispro Pump during admission | | Medications | 0 | | | | | |
| **insulin_lispro_scale** | Insulin Lispro Scale | categorical | Patient received Insulin Lispro Scale during admission | | Medications | 0 | | | | | |
| **insulin_lispro_u100_pen** | Insulin Lispro u100 Pen | categorical | Patient received Insulin Lispro u100 Pen during admission | | Medications | 0 | | | | | |
| **insulin_lispro_u100_sc** | Insulin Lispro u100 Subcutaneous | categorical | Patient received Insulin Lispro u100 Subcutaneous during admission | | Medications | 0 | | | | | |
| **insulin_long** | Insulin Long | categorical | Patient received Insulin Long during admission | | Medications | 0 | | | | | |
| **insulin_medium** | Insulin Medium | categorical | Patient received Insulin Medium during admission | | Medications | 0 | | | | | |
| **insulin_mix** | Insulin Mix | categorical | Patient received Insulin Mix during admission | | Medications | 0 | | | | | |
| **insulin_nph** | Insulin NPH | categorical | Patient received Insulin NPH during admission | | Medications | 0 | | | | | |
| **insulin_nph_pen** | Insulin NPH Pen | categorical | Patient received Insulin NPH Pen during admission | | Medications | 0 | | | | | |
| **insulin_nph_reg_7030** | Insulin NPH Reg 7030 | categorical | Patient received Insulin NPH Reg 7030 during admission | | Medications | 0 | | | | | |
| **insulin_nph_reg_mix** | Insulin NPH Reg Mix | categorical | Patient received Insulin NPH Reg Mix during admission | | Medications | 0 | | | | | |
| **insulin_nph_sc** | Insulin NPH Subcutaneous | categorical | Patient received Insulin NPH Subcutaneous during admission | | Medications | 0 | | | | | |
| **insulin_pump_bolus** | Insulin Pump Bolus | categorical | Patient received Insulin Pump Bolus during admission | | Medications | 0 | | | | | |
| **insulin_regular** | Insulin Regular | categorical | Patient received Insulin Regular during admission | | Medications | 0 | | | | | |
| **insulin_regular_bolus** | Insulin Regular Bolus | categorical | Patient received Insulin Regular Bolus during admission | | Medications | 0 | | | | | |
| **insulin_regular_conc** | Insulin Regular Concentrated | categorical | Patient received Insulin Regular Concentrated during admission | | Medications | 0 | | | | | |
| **insulin_regular_gtt** | Insulin Regular Glucose Tolerance Test | categorical | Patient received Insulin Regular Glucose Tolerance Test during admission | | Medications | 0 | | | | | |
| **insulin_regular_sc** | Insulin Regular Subcutaneous | categorical | Patient received Insulin Regular Subcutaneous during admission | | Medications | 0 | | | | | |
| **insulin_regular_scale** | Insulin Regular Scale | categorical | Patient received Insulin Regular Scale during admission | | Medications | 0 | | | | | |
| **insulin_regular_u500_pe** | Insulin Regular u500 Pen | categorical | Patient received Insulin Regular u500 Pen during admission | | Medications | 0 | | | | | |
| **insulin_short** | Insulin Short | categorical | Patient received Insulin Short during admission | | Medications | 0 | | | | | |
| **insulin_short_csii** | Insulin Short CSII | categorical | Patient received Insulin Short CSII during admission | | Medications | 0 | | | | | |
| **iodixanol** | Iodixanol | categorical | Patient received Iodixanol during admission | | Medications | 0 | | | | | med |
| **iohexol** | Iohexol | categorical | Patient received Iohexol during admission | | Medications | 0 | | | | | med |
| **iopamidol** | Iopamidol | categorical | Patient received Iopamidol during admission | | Medications | 0 | | | | | med |
| **iothalamate** | Iothalamate | categorical | Patient received Iothalamate during admission | | Medications | 0 | | | | | med |
| **ioversol** | Ioversol | categorical | Patient received Ioversol during admission | | Medications | 0 | | | | | med |
| **ipratropium** | Ipratropium | categorical | Patient received Ipratropium during admission | | Medications | 0 | | | | | med |
| **irbesartan** | Irbesartan | categorical | Patient received Irbesartan during admission | | Medications | 0 | | | | | |
| **irinotecan** | Irinotecan | categorical | Patient received Irinotecan during admission | | Medications | 0 | | | | | |
| **ironmed** | Iron Medication | categorical | Patient received Iron Medication during admission | | Medications | 0 | | | | | med |
| **isavuconazonium** | Isavuconazonium | categorical | Patient received Isavuconazonium during admission | | Medications | 0 | | | | | med |
| **isoniazid** | Isoniazid | categorical | Patient received Isoniazid during admission | | Medications | 0 | | | | | med |
| **isoproterenol** | Isoproterenol | categorical | Patient received Isoproterenol during admission | | Medications | 0 | | | | | med |
| **isordil** | Isordil | categorical | Patient received Isordil during admission | | Medications | 0 | | | | | med |
| **itraconazole** | Itraconazole | categorical | Patient received Itraconazole during admission | | Medications | 0 | | | | | med |
| **ivpb** | Intravenous Piggyback | categorical | Patient received Intravenous Piggyback during admission | | Medications | 0 | | | | | med |
| **kayexalate** | Kayexalate | categorical | Patient received Kayexalate during admission | | Medications | 0 | | | | | med |
| **kcliv** | Potassium Chloride IV | categorical | Patient received Potassium Chloride during admission | | Medications | 0 | | | | | med |
| **kcltab** | Potassium Chloride Tablet | categorical | Patient received Potassium Chloride Tablet during admission | | Medications | 0 | | | | | med |
| **keppra** | Keppra | categorical | Patient received Keppra during admission | | Medications | 0 | | | | | med |
| **ketamine** | Ketamine | categorical | Patient received Ketamine during admission | | Medications | 0 | | | | | med |
| **ketoconazole** | Ketoconazole | categorical | Patient received Ketoconazole during admission | | Medications | 0 | | | | | |
| **ketorolac** | Ketorolac | categorical | Patient received Ketorolac during admission | | Medications | 0 | | | | | med |
| **kphosiv** | Potassium Phosphate IV | categorical | Patient received Potassium Phosphate IV during admission | | Medications | 0 | | | | | med |
| **kphospo** | Potassium Phosphate | categorical | Patient received Potassium Phosphate during admission | | Medications | 0 | | | | | med |
| **ksparing** | Potassium Sparing Diuretic | categorical | Patient received Potassium Sparing Diuretic during admission | | Medications | 0 | | | | | med |
| **labetalol** | Labetalol | categorical | Patient received Labetalol during admission | | Medications | 0 | | | | | med |
| **lactulose** | Lactulose | categorical | Patient received Lactulose during admission | | Medications | 0 | | | | | med |
| **lamivudine** | Lamivudine | categorical | Patient received Lamivudine during admission | | Medications | 0 | | | | | med |
| **lamotrigine** | Lamotrigine | categorical | Patient received Lamotrigine during admission | | Medications | 0 | | | | | med |
| **lansoprazole** | Lansoprazole | categorical | Patient received Lansoprazole during admission | | Medications | 0 | | | | | med |
| **lanthanum** | Lanthanum | categorical | Patient received Lanthanum during admission | | Medications | 0 | | | | | med |
| **letrozole** | Letrozole | categorical | Patient received Letrozole during admission | | Medications | 0 | | | | | med |
| **levofloxacin** | Levofloxacin | categorical | Patient received Levofloxacin during admission | | Medications | 0 | | | | | med |
| **levothyroxine** | Levothyroxine | categorical | Patient received Levothyroxine during admission | | Medications | 0 | | | | | med |
| **lidocaine** | Lidocaine | categorical | Patient received Lidocaine during admission | | Medications | 0 | | | | | med |
| **linagliptin** | Linagliptin | categorical | Patient received Linagliptin during admission | | Medications | 0 | | | | | med |
| **linezolid** | Linezolid | categorical | Patient received Linezolid during admission | | Medications | 0 | | | | | med |
| **lisinopril** | Lisinopril | categorical | Patient received Lisinopril during admission | | Medications | 0 | | | | | med |
| **lithium** | Lithium | categorical | Patient received Lithium during admission | | Medications | 0 | | | | | med |
| **loopdiuretic** | Loop Diuretic | categorical | Patient received Loop Diuretic during admission | | Medications | 0 | | | | | med |
| **loperamide** | Loperamide | categorical | Patient received Loperamide during admission | | Medications | 0 | | | | | med |
| **lopinavir** | Lopinavir | categorical | Patient received Lopinavir during admission | | Medications | 0 | | | | | |
| **loratadine** | Loratadine | categorical | Patient received Loratadine during admission | | Medications | 0 | | | | | med |
| **lorazepam** | Lorazepam | categorical | Patient received Lorazepam during admission | | Medications | 0 | | | | | med |
| **losartan** | Losartan | categorical | Patient received Losartan during admission | | Medications | 0 | | | | | med |
| **Irbolus** | IR Bolus | categorical | Patient received IR Bolus during admission | | Medications | 0 | | | | | med |
| **Irdrip** | IR Drip | categorical | Patient received IR Drip during admission | | Medications | 0 | | | | | med |
| **magmed** | Magmed | categorical | Patient received Magmed during admission | | Medications | 0 | | | | | med |
| **mannitol** | Mannitol | categorical | Patient received Mannitol during admission | | Medications | 0 | | | | | med |
| **meclizine** | Meclizine | categorical | Patient received Meclizine during admission | | Medications | 0 | | | | | med |
| **melatonin** | Melatonin | categorical | Patient received Melatonin during admission | | Medications | 0 | | | | | med |
| **meloxicam** | Meloxicam | categorical | Patient received Meloxicam during admission | | Medications | 0 | | | | | med |
| **meperidine** | Meperidine | categorical | Patient received Meperidine during admission | | Medications | 0 | | | | | med |
| **mercaptopurine** | Mercaptopurine | categorical | Patient received Mercaptopurine during admission | | Medications | 0 | | | | | med |
| **meropenem** | Meropenem | categorical | Patient received Meropenem during admission | | Medications | 0 | | | | | med |
| **mesalamine** | Mesalamine | categorical | Patient received Mesalamine during admission | | Medications | 0 | | | | | med |
| **metformin** | Metformin | categorical | Patient received Metformin during admission | | Medications | 0 | | | | | med |
| **methadone** | Methadone | categorical | Patient received Methadone during admission | | Medications | 0 | | | | | med |
| **methimazole** | Methimazole | categorical | Patient received Methimazole during admission | | Medications | 0 | | | | | med |

| | | | | | | | | | | | |
|---|---|---|---|---|---|---|---|---|---|---|---|
| **methotrexate** | Methotrexate | categorical | Patient received Methotrexate during admission | | Medications | 0 | | | | | med |
| **methylprednisolone** | Methylprednisolone | categorical | Patient received Methylprednisolone during admission | | Medications | 0 | | | | | med |
| **metoclopramide** | Metoclopramide | categorical | Patient received Metoclopramide during admission | | Medications | 0 | | | | | med |
| **metolazone** | Metolazone | categorical | Patient received Metolazone during admission | | Medications | 0 | | | | | med |
| **metoprolol** | Metoprolol | categorical | Patient received Metoprolol during admission | | Medications | 0 | | | | | med |
| **metronidazole** | Metronidazole | categorical | Patient received Metronidazole during admission | | Medications | 0 | | | | | med |
| **mexiletine** | Mexiletine | categorical | Patient received Mexiletine during admission | | Medications | 0 | | | | | med |
| **midazolam** | Midazolam | categorical | Patient received Midazolam during admission | | Medications | 0 | | | | | med |
| **midodrine** | Midodrine | categorical | Patient received Midodrine during admission | | Medications | 0 | | | | | med |
| **milrinone** | Milrinone | categorical | Patient received Milrinone during admission | | Medications | 0 | | | | | med |
| **minocycline** | Minocycline | categorical | Patient received Minocycline during admission | | Medications | 0 | | | | | med |
| **minoxidil** | Minoxidil | categorical | Patient received Minoxidil during admission | | Medications | 0 | | | | | med |
| **miralax** | Miralax | categorical | Patient received Miralax during admission | | Medications | 0 | | | | | med |
| **mirtazapine** | Mirtazapine | categorical | Patient received Mirtazapine during admission | | Medications | 0 | | | | | med |
| **misoprostol** | Misoprostol | categorical | Patient received Misoprostol during admission | | Medications | 0 | | | | | med |
| **mitomycin** | Mitomycin | categorical | Patient received Mitomycin during admission | | Medications | 0 | | | | | med |
| **modafinil** | Modafinil | categorical | Patient received Modafinil during admission | | Medications | 0 | | | | | med |
| **montelukast** | Montelukast | categorical | Patient received Montelukast during admission | | Medications | 0 | | | | | med |
| **morphine** | Morphine | categorical | Patient received Morphine during admission | | Medications | 0 | | | | | med |
| **moxifloxacin** | Moxifloxacin | categorical | Patient received Moxifloxacin during admission | | Medications | 0 | | | | | med |
| **mycophenolate** | Mycophenolate | categorical | Patient received Mycophenolate during admission | | Medications | 0 | | | | | med |
| **nadolol** | Nadolol | categorical | Patient received Nadolol during admission | | Medications | 0 | | | | | med |
| **naloxone** | Naloxone | categorical | Patient received Naloxone during admission | | Medications | 0 | | | | | med |
| **naltrexone** | Naltrexone | categorical | Patient received Naltrexone during admission | | Medications | 0 | | | | | med |
| **naproxen** | Naproxen | categorical | Patient received Naproxen during admission | | Medications | 0 | | | | | med |
| **narcotic** | Narcotic | categorical | Patient received Narcotic during admission | | Medications | 0 | | | | | med |
| **neomycin** | Neomycin | categorical | Patient received Neomycin during admission | | Medications | 0 | | | | | med |
| **neupogen** | Neupogen | categorical | Patient received Neupogen during admission | | Medications | 0 | | | | | med |
| **niacin** | Niacin | categorical | Patient received Niacin during admission | | Medications | 0 | | | | | med |
| **nicotine** | Nicotine | categorical | Patient received Nicotine during admission | | Medications | 0 | | | | | med |
| **nifedipine** | Nifedipine | categorical | Patient received Nifedipine during admission | | Medications | 0 | | | | | med |
| **nimodipine** | Nimodipine | categorical | Patient received Nimodipine during admission | | Medications | 0 | | | | | med |
| **nitrofurantoin** | Nitrofurantoin | categorical | Patient received Nitrofurantoin during admission | | Medications | 0 | | | | | med |
| **nitroglycerin** | Nitroglycerin | categorical | Patient received Nitroglycerin during admission | | Medications | 0 | | | | | med |
| **nitroprusside** | Nitroprusside | categorical | Patient received Nitroprusside during admission | | Medications | 0 | | | | | med |
| **noac** | New Oral Aticoagulant Drugs | categorical | Patient received New Oral Aticoagulant Drugs during admission | | Medications | 0 | | | | | med |
| **norepinephrine** | Norepinephrine | categorical | Patient received Norepinephrine during admission | | Medications | 0 | | | | | med |
| **nortriptyline** | Nortriptyline | categorical | Patient received Nortriptyline during admission | | Medications | 0 | | | | | med |
| **nsaid** | Non-steroidal Anti-Inflammatory Drugs | categorical | Patient received Non-steroidal Anti-Inflammatory Drugs during admission | | Medications | 0 | | | | | med |
| **nystatin** | Nystatin | categorical | Patient received Nystatin during admission | | Medications | 0 | | | | | med |
| **octreotide** | Octreotide | categorical | Patient received Octreotide during admission | | Medications | 0 | | | | | med |
| **olanzapine** | Olanzapine | categorical | Patient received Olanzapine during admission | | Medications | 0 | | | | | med |
| **olmesartan** | Olmesartan | categorical | Patient received Olmesartan during admission | | Medications | 0 | | | | | med |
| **omeprazole** | Omeprazole | categorical | Patient received Omeprazole during admission | | Medications | 0 | | | | | med |
| **ondansetron** | Ondansetron | categorical | Patient received Ondansetron during admission | | Medications | 0 | | | | | med |
| **oseltamivir** | Oseltamivir | categorical | Patient received Oseltamivir during admission | | Medications | 0 | | | | | med |
| **oxacillin** | Oxacillin | categorical | Patient received Oxacillin during admission | | Medications | 0 | | | | | med |
| **oxaliplatin** | Oxaliplatin | categorical | Patient received Oxaliplatin during admission | | Medications | 0 | | | | | med |
| **oxcarbazepine** | Oxcarbazepine | categorical | Patient received Oxcarbazepine during admission | | Medications | 0 | | | | | med |
| **oxybutynin** | Oxybutynin | categorical | Patient received Oxybutynin during admission | | Medications | 0 | | | | | med |
| **oxycodone** | Oxycodone | categorical | Patient received Oxycodone during admission | | Medications | 0 | | | | | med |
| **paclitaxel** | Paclitaxel | categorical | Patient received Paclitaxel during admission | | Medications | 0 | | | | | |
| **pantoprazole** | Pantoprazole | categorical | Patient received Pantoprazole during admission | | Medications | 0 | | | | | med |
| **paralytic** | Paralytic | categorical | Patient received Paralytic during admission | | Medications | 0 | | | | | med |
| **paricalcitol** | Paricalcitol | categorical | Patient received Paricalcitol during admission | | Medications | 0 | | | | | med |
| **paroxetine** | Paroxetine | categorical | Patient received Paroxetine during admission | | Medications | 0 | | | | | med |
| **pdmed** | Peritoneal Dialysis Medication | categorical | Patient received Peritoneal Dialysis Medication during admission | | Medications | 0 | | | | | med |
| **pemetrexed** | Pemetrexed | categorical | Patient received Pemetrexed during admission | | Medications | 0 | | | | | |
| **penicillin** | Penicillin | categorical | Patient received Penicillin during admission | | Medications | 0 | | | | | med |
| **pentamidine** | Pentamidine | categorical | Patient received Pentamidine during admission | | Medications | 0 | | | | | med |
| **perphenazine** | Perphenazine | categorical | Patient received Perphenazine during admission | | Medications | 0 | | | | | med |
| **phenobarbital** | Phenobarbital | categorical | Patient received Phenobarbital during admission | | Medications | 0 | | | | | med |
| **phenylephrine** | Phenylephrine | categorical | Patient received Phenylephrine during admission | | Medications | 0 | | | | | med |
| **phenytoin** | Phenytoin | categorical | Patient received Phenytoin during admission | | Medications | 0 | | | | | med |
| **phoslo** | Phoslo | categorical | Patient received Phoslo during admission | | Medications | 0 | | | | | med |
| **pioglitazone** | Pioglitazone | categorical | Patient received Pioglitazone during admission | | Medications | 0 | | | | | med |
| **piptazo** | Piptazo | categorical | Patient received Piptazo during admission | | Medications | 0 | | | | | med |
| **posaconazole** | Posaconazole | categorical | Patient received Posaconazole during admission | | Medications | 0 | | | | | med |
| **ppi** | Proton Pump Inhibitor | categorical | Patient received Proton Pump Inhibitor during admission | | Medications | 0 | | | | | med |
| **pramipexole** | Pramipexole | categorical | Patient received Pramipexole during admission | | Medications | 0 | | | | | med |
| **pravastatin** | Pravastatin | categorical | Patient received Pravastatin during admission | | Medications | 0 | | | | | med |
| **prazosin** | Prazosin | categorical | Patient received Prazosin during admission | | Medications | 0 | | | | | med |
| **prednisolone** | Prednisolone | categorical | Patient received Prednisolone during admission | | Medications | 0 | | | | | med |
| **prednisone** | Prednisone | categorical | Patient received Prednisone during admission | | Medications | 0 | | | | | med |
| **pregabalin** | Pregabalin | categorical | Patient received Pregabalin during admission | | Medications | 0 | | | | | med |
| **pressor** | Pressor | categorical | Patient received Pressor during admission | | Medications | 0 | | | | | med |
| **prochlorperazine** | Prochlorperazine | categorical | Patient received Prochlorperazine during admission | | Medications | 0 | | | | | med |
| **propofol** | Propofol | categorical | Patient received Propofol during admission | | Medications | 0 | | | | | med |
| **propranolol** | Propranolol | categorical | Patient received Propranolol during admission | | Medications | 0 | | | | | med |
| **protamine** | Protamine | categorical | Patient received Protamine during admission | | Medications | 0 | | | | | med |
| **pseudoephedrine** | Pseudoephedrine | categorical | Patient received Pseudoephedrine during admission | | Medications | 0 | | | | | med |
| **pyridoxine** | Pyridoxine | categorical | Patient received Pyridoxine during admission | | Medications | 0 | | | | | med |
| **quetiapine** | Quetiapine | categorical | Patient received Quetiapine during admission | | Medications | 0 | | | | | med |
| **quinapril** | Quinapril | categorical | Patient received Quinapril during admission | | Medications | 0 | | | | | |
| **rabeprazole** | Rabeprazole | categorical | Patient received Rabeprazole during admission | | Medications | 0 | | | | | |
| **raloxifene** | Raloxifene | categorical | Patient received Raloxifene during admission | | Medications | 0 | | | | | med |
| **raltegravir** | Raltegravir | categorical | Patient received Raltegravir during admission | | Medications | 0 | | | | | med |
| **ranolazine** | Ranolazine | categorical | Patient received Ranolazine during admission | | Medications | 0 | | | | | med |
| **rasburicase** | Rasburicase | categorical | Patient received Rasburicase during admission | | Medications | 0 | | | | | med |
| **remdesivir** | Remdesivir | categorical | Patient received Remdesivir during admission | | Medications | 0 | | | | | med |
| **remifentanil** | Remifentanil | categorical | Patient received Remifentanil during admission | | Medications | 0 | | | | | med |
| **ribavirin** | Ribavirin | categorical | Patient received Ribavirin during admission | | Medications | 0 | | | | | med |
| **rifampin** | Rifampin | categorical | Patient received Rifampin during admission | | Medications | 0 | | | | | med |
| **rifaximin** | Rifaximin | categorical | Patient received Rifaximin during admission | | Medications | 0 | | | | | med |
| **ringersdrip** | Ringers Drip | categorical | Patient received Ringers Drip during admission | | Medications | 0 | | | | | med |
| **risperidone** | Risperidone | categorical | Patient received Risperidone during admission | | Medications | 0 | | | | | med |
| **ritonavir** | Ritonavir | categorical | Patient received Ritonavir during admission | | Medications | 0 | | | | | med |
| **rituximab** | Rituximab | categorical | Patient received Rituximab during admission | | Medications | 0 | | | | | med |
| **ropinirole** | Ropinirole | categorical | Patient received Ropinirole during admission | | Medications | 0 | | | | | med |
| **rosuvastatin** | Rosuvastatin | categorical | Patient received Rosuvastatin during admission | | Medications | 0 | | | | | med |
| **salinebolus** | Saline Bolus | categorical | Patient received Saline Bolus during admission | | Medications | 0 | | | | | med |
| **salinedrip** | Saline Drip | categorical | Patient received Saline Drip during admission | | Medications | 0 | | | | | med |
| **salttab** | Salt Tablet | categorical | Patient received Salt Tablet during admission | | Medications | 0 | | | | | med |
| **sedative** | Sedative | categorical | Patient received Sedative during admission | | Medications | 0 | | | | | med |
| **selegiline** | Selegiline | categorical | Patient received Selegiline during admission | | Medications | 0 | | | | | med |
| **senna** | Senna | categorical | Patient received Senna during admission | | Medications | 0 | | | | | med |
| **sertraline** | Sertraline | categorical | Patient received Sertraline during admission | | Medications | 0 | | | | | med |
| **sevelamer** | Sevelamer | categorical | Patient received Sevelamer during admission | | Medications | 0 | | | | | med |
| **sildenafil** | Sildenafil | categorical | Patient received Sildenafil during admission | | Medications | 0 | | | | | med |
| **simethicone** | Simethicone | categorical | Patient received Simethicone during admission | | Medications | 0 | | | | | med |
| **simvastatin** | Simvastatin | categorical | Patient received Simvastatin during admission | | Medications | 0 | | | | | med |
| **sinemet** | Sinemet | categorical | Patient received Sinemet during admission | | Medications | 0 | | | | | med |
| **sirolimus** | Sirolimus | categorical | Patient received Sirolimus during admission | | Medications | 0 | | | | | med |
| **sitagliptin** | Sitagliptin | categorical | Patient received Sitagliptin during admission | | Medications | 0 | | | | | med |
| **sodiumbicarb** | Sodium Bicarbonate | categorical | Patient received Sodium Bicarbonate during admission | | Medications | 0 | | | | | med |
| **sotalol** | Sotalol | categorical | Patient received Sotalol during admission | | Medications | 0 | | | | | med |
| **spironolactone** | Spironolactone | categorical | Patient received Spironolactone during admission | | Medications | 0 | | | | | med |
| **statin** | Statin | categorical | Patient received Statin during admission | | Medications | 0 | | | | | med |
| **steroid** | Steroid | categorical | Patient received Steroid during admission | | Medications | 0 | | | | | med |
| **succinylcholine** | Succinylcholine | categorical | Patient received Succinylcholine during admission | | Medications | 0 | | | | | med |
| **sucralfate** | Sucralfate | categorical | Patient received Sucralfate during admission | | Medications | 0 | | | | | med |
| **sulfadiazine** | Sulfadiazine | categorical | Patient received Sulfadiazine during admission | | Medications | 0 | | | | | med |
| **sulfasalazine** | Sulfasalazine | categorical | Patient received Sulfasalazine during admission | | Medications | 0 | | | | | med |
| **tacrolimus** | Tacrolimus | categorical | Patient received Tacrolimus during admission | | Medications | 0 | | | | | med |

| | | | | | | | | | | | |
|---|---|---|---|---|---|---|---|---|---|---|---|
| tadalafil | Tadalafil | categorical | Patient received Tadalafil during admission | | Medications | 0 | | | | | med |
| tamoxifen | Tamoxifen | categorical | Patient received Tamoxifen during admission | | Medications | 0 | | | | | med |
| tamsulosin | Tamsulosin | categorical | Patient received Tamsulosin during admission | | Medications | 0 | | | | | med |
| technetium | Technetium | categorical | Patient received Technetium during admission | | Medications | 0 | | | | | med |
| temazepam | Temazepam | categorical | Patient received Temazepam during admission | | Medications | 0 | | | | | med |
| temsirolimus | Temsirolimus | categorical | Patient received Temsirolimus during admission | | Medications | 0 | | | | | med |
| tenofovir | Tenofovir | categorical | Patient received Tenofovir during admission | | Medications | 0 | | | | | med |
| terazosin | Terazosin | categorical | Patient received Terazosin during admission | | Medications | 0 | | | | | med |
| theophylline | Theophylline | categorical | Patient received Theophylline during admission | | Medications | 0 | | | | | med |
| thiaminemed | Thiamine Medication | categorical | Patient received Thiamine Medication during admission | | Medications | 0 | | | | | med |
| thiazide | Thiazide | categorical | Patient received Thiazide during admission | | Medications | 0 | | | | | med |
| ticagrelor | Ticagrelor | categorical | Patient received Ticagrelor during admission | | Medications | 0 | | | | | med |
| tigecycline | Tigecycline | categorical | Patient received Tigecycline during admission | | Medications | 0 | | | | | med |
| tiotropium | Tiotropium | categorical | Patient received Tiotropium during admission | | Medications | 0 | | | | | med |
| tobramycin | Tobramycin | categorical | Patient received Tobramycin during admission | | Medications | 0 | | | | | med |
| tobramycin_ag | | categorical | | | Medications | 0 | | | | | |
| tocilizumab | Tocilizumab | categorical | Patient received Tocilizumab during admission | | Medications | 0 | | | | | med |
| tolvaptan | Tolvaptan | categorical | Patient received Tolvaptan during admission | | Medications | 0 | | | | | med |
| topiramate | Topiramate | categorical | Patient received Topiramate during admission | | Medications | 0 | | | | | med |
| torsemide | Torsemide | categorical | Patient received Torsemide during admission | | Medications | 0 | | | | | med |
| tpnmed | Total Parenteral Nutrition Medication | categorical | Patient received Total Parenteral Nutrition Medication during admission | | Medications | 0 | | | | | med |
| tramadol | Tramadol | categorical | Patient received Tramadol during admission | | Medications | 0 | | | | | med |
| trazodone | Trazodone | categorical | Patient received Trazodone during admission | | Medications | 0 | | | | | med |
| unasyn | Unasyn | categorical | Patient received Unasyn during admission | | Medications | 0 | | | | | med |
| valacyclovir | Valacyclovir | categorical | Patient received Valacyclovir during admission | | Medications | 0 | | | | | med |
| valganciclovir | Valganciclovir | categorical | Patient received Valganciclovir during admission | | Medications | 0 | | | | | med |
| valproic | Valproic | categorical | Patient received Valproic during admission | | Medications | 0 | | | | | med |
| valsartan | Valsartan | categorical | Patient received Valsartan during admission | | Medications | 0 | | | | | med |
| vancomycin | Vancomycin | categorical | Patient received Vancomycin during admission | | Medications | 0 | | | | | med |
| vancomycinpo | Vancomycinpo | categorical | Patient received Vancomycinpo during admission | | Medications | 0 | | | | | med |
| vasopressin | Vasopressin | categorical | Patient received Vasopressin during admission | | Medications | 0 | | | | | med |
| vasopressor | Vasopressor | categorical | Patient received Vasopressor during admission | | Medications | 0 | | | | | med |
| vecuronium | Vecuronium | categorical | Patient received Vecuronium during admission | | Medications | 0 | | | | | med |
| venlafaxine | Venlafaxine | categorical | Patient received Venlafaxine during admission | | Medications | 0 | | | | | med |
| verapamil | Verapamil | categorical | Patient received Verapamil during admission | | Medications | 0 | | | | | med |
| vincristine | Vincristine | categorical | Patient received Vincristine during admission | | Medications | 0 | | | | | med |
| vitamincmed | Vitamin C Medication | categorical | Patient received Vitamin C Medication during admission | | Medications | 0 | | | | | med |
| vitamindmed | Vitamin D Medication | categorical | Patient received Vitamin D Medication during admission | | Medications | 0 | | | | | med |
| voriconazole | Voriconazole | categorical | Patient received Voriconazole during admission | | Medications | 0 | | | | | med |
| warfarin | Warfarin | categorical | Patient received Warfarin during admission | | Medications | 0 | | | | | med |
| zincmed | Zinc Medication | categorical | Patient received Zinc Medication during admission | | Medications | 0 | | | | | med |
| ziprasidone | Ziprasidone | categorical | Patient received Ziprasidone during admission | | Medications | 0 | | | | | med |
| zoledronic | Zoledronic | categorical | Patient received Zoledronic during admission | | Medications | 0 | | | | | med |
| zolpidem | Zolpidem | categorical | Patient received Zolpidem during admission | | Medications | 0 | | | | | med |
| zonisamide | Zonisamide | categorical | Patient received Zonisamide during admission | | Medications | 0 | | | | | med |
| record_id | Health Record ID | | Patient health record ID | | Metadata | 0 | | | | | |
| arrhythmia | Arrhythmia | categorical | Prior Arrhythmia Diagnosis | | Past Medical History | 0 | | | | | pmhx |
| bloodlossanemia | Blood Loss Anemia | categorical | Prior Diagnosis of Blood Loss Anemia | | Past Medical History | 0 | | | | | pmhx |
| cerebrovasc_disease | | | | | Past Medical History | 0 | | | | | |
| chf | Congestive Heart Failure | categorical | Prior Diagnosis of congestive heart failure | | Past Medical History | 0 | | | | | pmhx |
| ckd | | | | | Past Medical History | 0 | | | | | |
| coagulopathy | Coagulopathy | categorical | Prior experience of Coagulopathy | | Past Medical History | 0 | | | | | pmhx |
| cpd | Cephalopelvic Disproportion | categorical | Prior Diagnonsis of Cephalopelvic Disproportion | | Past Medical History | 0 | | | | | pmhx |
| deficiencyanemia | Anemia Deficiency | categorical | Prior Anemia Deficiency Diagnosis | | Past Medical History | 0 | | | | | pmhx |
| depression | Depression | categorical | Prior Diagnosis of Depression | | Past Medical History | 0 | | | | | pmhx |
| diabetescomp | Diabetes complicated | categorical | Prior diagnosis of complicated diabetes | | Past Medical History | 0 | | | | | pmhx |
| diabetesuncomp | Diabetest uncomplicated | categorical | Prior diagnosis of uncomplicated diabetes | | Past Medical History | 0 | | | | | pmhx |
| drugabuse | Drug Abuse | categorical | Prior Diagnosis of Drug Abuse | | Past Medical History | 0 | | | | | pmhx |
| elx_score | Elixhauser score | continuous | Elixhauser comorbidity score | | Past Medical History | 0 | | | | | pmhx |
| esrd | End-Stage Renal Dialysis | categorical | Prior Diagnosis of End-Stage Renal Dialysis | | Past Medical History | 0 | | | | | pmhx |
| etohabuse | Ethyl Alcohol Abuse | categorical | Prior Diagnosis of Ethyl Alcohol Abuse | | Past Medical History | 0 | | | | | pmhx |
| fluidelecdisorders | Fluidelec Disorders | categorical | Prior Fluidelec Disorders Diagnosis | | Past Medical History | 0 | | | | | pmhx |
| hiv | HIV | categorical | Prior HIV Diagnosis | | Past Medical History | 0 | | | | | pmhx |
| hospital_afib_aflutter | hospital_afib_aflutter | categorical | diagnosis of afib or aflutter during current hospitalization | | Past Medical History | 0 | | | | | |
| hospital_arrhythmia | hospital_arrhythmia | categorical | diagnosis of arrhythmia during current hospitalization | | Past Medical History | 0 | | | | | |
| hospital_bradycardia | hospital_bradycardia | categorical | diagnosis of bradycardia during current hospitalization | | Past Medical History | 0 | | | | | |
| hospital_heart_failure | hospital_heart_failure | categorical | diagnosis of heart failure during current hospitalization | | Past Medical History | 0 | | | | | |
| hospital_intracranial_he | hospital_intracranial_hem | categorical | diagnosis of intracranial hemorrhage during current hospitalization | | Past Medical History | 0 | | | | | |
| hospital_myocarditis | hospital_myocarditis | categorical | diagnosis of myocarditis during current hospitalization | | Past Medical History | 0 | | | | | |
| hospital_pericarditis | hospital_pericarditis | categorical | diagnosis of pericarditis during current hospitalization | | Past Medical History | 0 | | | | | |
| hospital_stroke | hospital_stroke | categorical | diagnosis of stroke during current hospitalization | | Past Medical History | 0 | | | | | |
| hospital_tia | hospital_tia | categorical | diagnosis of TIA during current hospitalization | | Past Medical History | 0 | | | | | |
| htncomp | Hypertension complicated | categorical | Prior Diagnosis of complicated hypertension | | Past Medical History | 0 | | | | | pmhx |
| htnuncomp | Hypertension uncomplicated | categorical | Prior Diagnosis of uncomplicated hypertension | | Past Medical History | 0 | | | | | pmhx |
| hypothyroid | Hypothyroid | categorical | Prior Hypothyroid Diagnosis | | Past Medical History | 0 | | | | | pmhx |
| liverdisease | Liver Disease | categorical | Prior Diagnosis of Liver Disease | | Past Medical History | 0 | | | | | pmhx |
| lymphoma | Lymphoma | categorical | Prior Diagnosis of Lymphoma | | Past Medical History | 0 | | | | | pmhx |
| malignancy | Malignancy | categorical | Prior Diagnosis of Malignancy | | Past Medical History | 0 | | | | | pmhx |
| metastasis | Metastasis | categorical | Prior Diagnosis of Metastasis | | Past Medical History | 0 | | | | | pmhx |
| myocardial_infarction | | | | | Past Medical History | 0 | | | | | |
| neurodz | Neuro Disease | categorical | Prior Neuro Disease Diagnosis | | Past Medical History | 0 | | | | | pmhx |
| obesity | Obesity | categorical | Prior Obesity Diagnosis | | Past Medical History | 0 | | | | | pmhx |
| paralysis | Paralysis | categorical | Prior Paralysis Diagnosis | | Past Medical History | 0 | | | | | pmhx |
| pmhx_afib_aflutter | pmhx_afib_aflutter | categorical | prior diagnosis of afib or aflutter | | Past Medical History | 0 | | | | | |
| pmhx_arrhythmia | pmhx_arrhythmia | categorical | prior diagnosis of arrhythmia | | Past Medical History | 0 | | | | | |
| pmhx_bradycardia | pmhx_bradycardia | categorical | prior diagnosis of bradycardia | | Past Medical History | 0 | | | | | |
| pmhx_heart_failure | pmhx_heart_failure | categorical | prior diagnosis of heart failure | | Past Medical History | 0 | | | | | |
| pmhx_intracranial_hem | pmhx_intracranial_hem | categorical | prior diagnosis of intracranial hemorrhage | | Past Medical History | 0 | | | | | |
| pmhx_myocarditis | pmhx_myocarditis | categorical | prior diagnosis of myocarditis | | Past Medical History | 0 | | | | | |
| pmhx_pericarditis | pmhx_pericarditis | categorical | prior diagnosis of pericarditis | | Past Medical History | 0 | | | | | |
| pmhx_stroke | pmhx_stroke | categorical | prior diagnosis of stroke | | Past Medical History | 0 | | | | | |
| pmhx_tia | pmhx_tia | categorical | prior diagnosis of | | Past Medical History | 0 | | | | | |
| psychosis | Psychosis | categorical | Prior Diagnosis of Psychosis | | Past Medical History | 0 | | | | | pmhx |
| pud | Peptic Ulcer Disease | categorical | Prior Diagnosis Peptic Ulcer Disease Diagnosis | | Past Medical History | 0 | | | | | pmhx |
| pulmcirc | Pulmonary Circulation | categorical | Prior Pulmonary Circulation Disorder | | Past Medical History | 0 | | | | | pmhx |
| pvd | Pulmonary Vascular Disease | categorical | Prior Diagnosis of Pulmonary Vascular Disease | | Past Medical History | 0 | | | | | pmhx |
| renalfailure | Renal Failure | categorical | Prior Renal Failure Diagnosis | | Past Medical History | 0 | | | | | pmhx |
| renaltransplant | Renal Transplant Recipient | categorical | Recipient of prior Renal Transplant procedure | | Past Medical History | 0 | | | | | pmhx |
| rheumatoid | Rheumatoid Arthritis | categorical | Prior Rheumatoid Arthritis Diagnosis | | Past Medical History | 0 | | | | | pmhx |
| transplant | Transplant | categorical | Prior Transplant Recipient | | Past Medical History | 0 | | | | | pmhx |
| valvedz | Heart-Valve Disease | categorical | Prior Diagnosis of Heart-Valve Disease | | Past Medical History | 0 | | | | | pmhx |
| weightloss | Weight Loss | categorical | Prior Weight Loss Diagnosis | | Past Medical History | 0 | | | | | pmhx |
| addictionconsult | Addiction Consult | categorical | Patient received Addiction Medicine Consultation during admission | | Procedures | 0 | | | | | procedure |
| allergyconsult | Allergy Consult | categorical | Patient received Allergy Consultation during admission | | Procedures | 0 | | | | | procedure |
| autopsy | Autopsy | categorical | Patient received Autopsy during admission | | Procedures | 0 | | | | | procedure |
| axr | Abdominal X-Ray | categorical | Patient received Abdominal X-Ray during admission | | Procedures | 0 | | | | | procedure |
| bipap | Bilevel Positive Airway Pressure | categorical | Patient received Bilevel Positive Airway Pressure therapy during admission | | Procedures | 0 | | | | | procedure |
| cardiaccath | Cardiac Catheterization | categorical | Patient underwent Cardiac Catheterization during admission | | Procedures | 0 | | | | | procedure |
| centralline | Central Venus Catheter | categorical | Patient recieved or utilized central line during admission | | Procedures | 0 | | | | | procedure |
| ciwa | Clinical Institute Withdrawal Assessment for Alcohol | categorical | Patient underwent Clinical Institutional Withdrawal Assessment for Alcohol during admission | | Procedures | 0 | | | | | procedure |
| codecomfort | Code Comfort | categorical | Patient utilized Code Comfort during admission | | Procedures | 0 | | | | | procedure |
| codefull | Code Full | categorical | Patient utilized Code Full during admission | | Procedures | 0 | | | | | procedure |
| codelimited | Code Limited | categorical | Patient utilized Code Limited during admission | | Procedures | 0 | | | | | procedure |
| consult_neuro | consult_neuro | categorical | Consult neurology order placed during patient's encounter | | Procedures | 0 | | | | | |
| contactprecaution | Contact Precaution | categorical | Patient labeled for Contact Precaution during admission | | Procedures | 0 | | | | | procedure |
| contrastir | Contrast Interventional Radiology | categorical | Patient received Contrast Interventional Radiology during admission | | Procedures | 0 | | | | | procedure |
| contraststudy | Contrast Study | categorical | Patient underwent Continuous Contrast Study Imaging during admission | | Procedures | 0 | | | | | procedure |
| cpap | Continuous Positive Airway Pressure Therapy | categorical | Patient received Continuous Positive Airway Pressure Therapy during admission | | Procedures | 0 | | | | | procedure |
| crrt | Continuous Positive Airway Pressure Therapy | categorical | Patient received Continuous Positive Airway Pressure Therapy during admission | | Procedures | 0 | | | | | procedure |
| ctabdomen | CT Scan Abdomen | categorical | Patient received Abdominal CT Scan during admission | | Procedures | 0 | | | | | procedure |
| ctchest | CT Scan Chest | categorical | Patient received Chest CT Scan during admission | | Procedures | 0 | | | | | procedure |
| ctextremities | Extremities CT Scan | categorical | Patient received Extremities CT Scan during admission | | Procedures | 0 | | | | | procedure |
| cthead | Head CT Scan | categorical | Patient received Head CT Scan during admission | | Procedures | 0 | | | | | procedure |
| cxr | Chest X-Ray | categorical | Patient received Chest X-Ray during admission | | Procedures | 0 | | | | | procedure |

| | | | | | | | | | | | |
|---|---|---|---|---|---|---|---|---|---|---|---|
| **diabetesconsult** | Diabetes Consult | categorical | Patient received Diabetes consult during admission | | Procedures | 0 | | | | | procedure |
| **ecmo** | Extracorporeal Membrane Oxygenation | categorical | Patient received Extracorporeal Membrane Oxygenation during admission | | Procedures | 0 | | | | | |
| **eeg** | Electroencephalogram | categorical | Patient received Electroencephalogram during admission | | Procedures | 0 | | | | | procedure |
| **facemask** | Face Mask | categorical | Patient utilized Face Mask during admission | | Procedures | 0 | | | | | procedure |
| **fallrisk** | Fall Risk | categorical | Patient labeled for Fall Risk during admission | | Procedures | 0 | | | | | procedure |
| **feedingtube** | Feeding Tube | categorical | Patient utilized Feeding Tube during admission | | Procedures | 0 | | | | | procedure |
| **foley** | Foley Catheter | categorical | Patient utilized Foley Catheter during admission | | Procedures | 0 | | | | | procedure |
| **gadoliniumstudy** | Gadolinium Study | categorical | Patient underwent Gadolinium Study during admission | | Procedures | 0 | | | | | procedure |
| **gemsconsult** | Gems Consult | categorical | Patient received Gems Consultation during admission | | Procedures | 0 | | | | | procedure |
| **hemodialysis** | Hemodialysis | categorical | Patient received Hemodialysis treatment during admission | | Procedures | 0 | | | | | procedure |
| **heparinprotocol** | Heparin Protocol | categorical | Patient revceived Heparin Protocol during admission | | Procedures | 0 | | | | | procedure |
| **highflow** | High Flow Systems | categorical | Patient utilized High Flow device(s) during admission | | Procedures | 0 | | | | | procedure |
| **hospice** | Hospice | categorical | Patient received Hospice Care during admission | | Procedures | 0 | | | | | procedure |
| **iabp** | Intra-aortic Balloon Pump | categorical | Patient utilized Intra-aortic Balloon Pump during admission | | Procedures | 0 | | | | | procedure |
| **mrabdomen** | Abdominal MR Test | categorical | Patient received Abdominal MR Test during admission | | Procedures | 0 | | | | | procedure |
| **mrchest** | Chest MR Test | categorical | Patient received Chest MR Test during admission | | Procedures | 0 | | | | | procedure |
| **mrextremities** | Extremities MR Test | categorical | Patient received Extremities MR Test during admission | | Procedures | 0 | | | | | procedure |
| **mrhead** | Head MR Test | categorical | Patient received Head MR Test during admission | | Procedures | 0 | | | | | procedure |
| **mrspine** | Spinal MR Test | categorical | Patient received Spinal MR Test during admission | | Procedures | 0 | | | | | procedure |
| **nasalcannula** | Nasal Cannula | categorical | Patient utilized Nasal Cannula during admission | | Procedures | 0 | | | | | procedure |
| **consult_neuro** | consult_neuro | categorical | NIHSS scale order placed during patient's encounter | | Procedures | 0 | | | | | |
| **nonrebreather** | Non-rebreather Mask | categorical | Patient utilized Non-rebreather Mask during admission | | Procedures | 0 | | | | | procedure |
| **npoorder** | NPO Order | categorical | Patient received NPO (Nothing by mouth) order during admission | | Procedures | 0 | | | | | procedure |
| **onetone** | One to one | categorical | Patient ordered a """"one to one"""" (sitter) during hospitalization | | Procedures | 0 | | | | | procedure |
| **palliativecareconsult** | Palliative Care Consult | categorical | Patient received Palliative Care Consultation during admission | | Procedures | 0 | | | | | procedure |
| **paracentesis** | Paracentesis | categorical | Patient received Paracentesis treatment during admission | | Procedures | 0 | | | | | procedure |
| **peritonealdial** | Peritoneal Dialysis | categorical | Patient received Peritoneal Dialysis during admission | | Procedures | 0 | | | | | procedure |
| **plasmapheresis** | Plasmapheresis | categorical | Patient received Plasmapheresis during admission | | Procedures | 0 | | | | | procedure |
| **psychconsult** | Psychiatric Consult | categorical | Patient received Psychiatric Consultation during admission | | Procedures | 0 | | | | | procedure |
| **ptot** | Physical Therapy & Occupational Therapy | categorical | Patient received Physical and Occupational Therapy during admission | | Procedures | 0 | | | | | procedure |
| **rectaltube** | Rectal Tube | categorical | Patient utilized Rectal Tube during admission | | Procedures | 0 | | | | | procedure |
| **renaldiet** | Renal Diet | categorical | Patient placed on Renal Diet during admission | | Procedures | 0 | | | | | procedure |
| **restraints** | Medical Restraints | categorical | Patient received Medical Restraints during admission | | Procedures | 0 | | | | | procedure |
| **scdorder** | Sequential Compression Device Order | categorical | Patient received Sequential Compression Device Order during admission | | Procedures | 0 | | | | | procedure |
| **speechswallow** | Speech Swallow | categorical | Patient underwent speech swallow study during admission | | Procedures | 0 | | | | | procedure |
| **tee** | Transesophageal Echocardiogram | categorical | Patient received Transesophageal Echocardiogram during admission | | Procedures | 0 | | | | | procedure |
| **telemetry** | Telemetry | categorical | Patient utilized Telemetry during admission | | Procedures | 0 | | | | | procedure |
| **tpn** | Total Parenteral Nutrition | categorical | Patient utilized Total Parenteral Nutrition during admission | | Procedures | 0 | | | | | procedure |
| **transfuseplasma** | Plasma Transfusion | categorical | Patient received Plasma Transfusion during admission | | Procedures | 0 | | | | | procedure |
| **transfuseplatelet** | Platelet Transfusion | categorical | Patient received Platelet Transfusion during admission | | Procedures | 0 | | | | | procedure |
| **transfuserbc** | Red Blood Cell Transfusion | categorical | Patient received Red Blood Cell Transfusion during admission | | Procedures | 0 | | | | | procedure |
| **tte** | Transthoracic Echocardiogram | categorical | Patient received Transthoracic Echocardiogram during admission | | Procedures | 0 | | | | | procedure |
| **usabdomen** | Abdominal Ultrasound | categorical | Patient received Abdominal Ultrasound during admission | | Procedures | 0 | | | | | procedure |
| **usdvt** | Deep Vein Thrombosis Ultrasound | categorical | Patient received Deep Vein Thrombosis Ultrasound during admission | | Procedures | 0 | | | | | procedure |
| **uslung** | Lung Ultrasound | categorical | Patient received Lung Ultrasound during admission | | Procedures | 0 | | | | | procedure |
| **usrenal** | Renal Ultrasound | categorical | Patient received Renal Ultrasound during admission | | Procedures | 0 | | | | | procedure |
| **ventorder** | Vent Order | categorical | Patient received Vent Order during admission | | Procedures | 0 | | | | | procedure |
| **woundcare** | Wound Care | categorical | Patient received Wound Care during admission | | Procedures | 0 | | | | | procedure |
| **diastolic_25** | Diastolic BP (25th Percentile) | continuous | Diastolic blood pressure at the 25th percentile of this patient's measurements | mmHg | Vitals | 0 | | | | | vitals |
| **diastolic_75** | Diastolic BP (75th Percentile) | continuous | Diastolic blood pressure at the 75th percentile of this patient's measurements | mmHg | Vitals | 0 | | | | | vitals |
| **diastolic_count** | Diastolic BP (Count) | continuous | Count of diastolic blood pressure measurements for this patient | Number of measurements | Vitals | 0 | | | | | vitals |
| **diastolic_first** | Diastolic BP (First) | continuous | First diastolic blood pressure measurement for this patient's hospitalization | mmHg | Vitals | 0 | | | | | vitals |
| **diastolic_last** | Diastolic BP (Last) | continuous | Last diastolic blood pressure measurement for this patient's hospitalization | mmHg | Vitals | 0 | | | | | vitals |
| **diastolic_max** | Diastolic BP (Max) | continuous | Maximum diastolic blood pressure for this patient's hospitalization | mmHg | Vitals | 0 | | | | | vitals |
| **diastolic_mean** | Diastolic BP (Mean) | continuous | Average diastolic blood pressure for this patient's hospitalization | mmHg | Vitals | 0 | | | | | vitals |
| **diastolic_median** | Diastolic BP (Median) | continuous | Median diastolic blood pressure for this patient's hospitalization | mmHg | Vitals | 0 | | | | | vitals |
| **diastolic_min** | Diastolic BP (Min) | continuous | Minimum diastolic blood pressure for this patient's hospitalization | mmHg | Vitals | 0 | | | | | vitals |
| **diastolic_sd** | Diastolic Bp (SD) | continuous | Standard deviation of the diastolic blood pressures during this patient's hospitalization | mmHg | Vitals | 0 | | | | | vitals |
| **pulse_25** | Heart Rate (25th Percentile) | continuous | Heart rate at the 25th percentile of this patient's measurements | Beats per minute | Vitals | 0 | | | | | vitals |
| **pulse_75** | Heart Rate (75th Percentile) | continuous | Heart rate at the 75th percentile of this patient's measurements | Beats per minute | Vitals | 0 | | | | | vitals |
| **pulse_count** | Heart Rate (Count) | continuous | Count of heart rate measurements for this patient | Number of measurements | Vitals | 0 | | | | | vitals |
| **pulse_first** | Heart Rate (First) | continuous | First heart rate measurement for this patient's hospitalization | Beats per minute | Vitals | 0 | | | | | vitals |
| **pulse_last** | Heart Rate (Last) | continuous | Last heart rate measurement for this patient's hospitalization | Beats per minute | Vitals | 0 | | | | | vitals |
| **pulse_max** | Heart Rate (Max) | continuous | Maximum heart rate for this patient's hospitalization | Beats per minute | Vitals | 0 | | | | | vitals |
| **pulse_mean** | Heart Rate (Mean) | continuous | Average heart rate for this patient's hospitalization | Beats per minute | Vitals | 0 | | | | | vitals |
| **pulse_median** | Heart Rate (Median) | continuous | Median heart rate of this patient's hospitalization | Beats per minute | Vitals | 0 | | | | | vitals |
| **pulse_min** | Heart Rate (Minimum) | continuous | Minimum heart rate for this patient's hospitalization | Beats per minute | Vitals | 0 | | | | | vitals |
| **pulse_sd** | Heart Rate (SD) | continuous | Standard deviation of the heart rate measurements during this patient's hospitalization | Beats per minute | Vitals | 0 | | | | | vitals |
| **resp_25** | Respiratory Rate (25th Percentile) | continuous | Respiratory rate at the 25th percentile of this patient's measurements | Beats per minute | Vitals | 0 | | | | | vitals |
| **resp_75** | Respiratory Rate (75th Percentile) | continuous | Respiratory rate at the 75th percentile of this patient's measurements | Beats per minute | Vitals | 0 | | | | | vitals |
| **resp_count** | Respiratory Rate (Count) | continuous | Count of respiratory rate measurements for this patient | Number of measurements | Vitals | 0 | | | | | vitals |
| **resp_first** | Respiratory Rate (First) | continuous | First respiratory rate measurement for this patient's hospitalization | Beats per minute | Vitals | 0 | | | | | vitals |
| **resp_last** | Respiratory Rate (Last) | continuous | Last respiratory rate measurement for this patient's hospitalization | Beats per minute | Vitals | 0 | | | | | vitals |
| **resp_max** | Respiratory Rate (Max) | continuous | Maximum respiratory rate for this patient's hospitalization | Beats per minute | Vitals | 0 | | | | | vitals |
| **resp_mean** | Respiratory Rate (Mean) | continuous | Average respiratory rate for this patient's hospitalization | Beats per minute | Vitals | 0 | | | | | vitals |
| **resp_median** | Respiratory Rate (Median) | continuous | Median respiratory rate for this patient's hospitalization | Beats per minute | Vitals | 0 | | | | | vitals |
| **resp_min** | Respiratory Rate (Min) | continuous | Minimum respiratory rate for this patient's hospitalization | Beats per minute | Vitals | 0 | | | | | vitals |
| **resp_sd** | Respiratory Rate (SD) | continuous | Standard deviation of the respiratory rate measurements during this patient's hospitalization | Beats per minute | Vitals | 0 | | | | | vitals |
| **spo2_25** | Oxygen Saturation Ratio (25th Percentile)? | continuous | Oxygen Saturation Ratio at the 25th percentile of this patient's measurements | Percent? | Vitals | 0 | | | | | vitals |
| **spo2_75** | Oxygen Saturation Ratio (75th Percentile) | continuous | Oxygen Saturation Ratio at the 75th percentile of this patient's measurements | Percent | Vitals | 0 | | | | | vitals |
| **spo2_count** | Oxygen Saturation Ratio (Count) | continuous | Count of oxygen saturation ratio measurements for this patient | Number of measurements | Vitals | 0 | | | | | vitals |
| **spo2_first** | Oxygen Saturation Ratio (First) | continuous | First oxygen saturation ratio measurement for this patient's hospitalization | Percent | Vitals | 0 | | | | | vitals |
| **spo2_last** | Oxygen Saturation Ratio (Last) | continuous | Last oxygen saturation ratio measurement for this patient's hospitalization | Percent | Vitals | 0 | | | | | vitals |
| **spo2_max** | Oxygen Saturation Ratio (Max) | continuous | Maximum oxygen saturation ratio for this patient's hospitalization | Percent | Vitals | 0 | | | | | vitals |
| **spo2_mean** | Oxygen Saturation Ratio (Mean) | continuous | Average oxygen saturation ratio for this patient's hospitalization | Percent | Vitals | 0 | | | | | vitals |
| **spo2_median** | Oxygen Saturation Ratio (Median) | continuous | Median oxygen saturation ratio for this patient's hospitalization | Percent | Vitals | 0 | | | | | vitals |
| **spo2_min** | Oxygen Saturation Ratio (Min) | continuous | Minimum oxygen saturation ratio for this patient's hospitalization | Percent | Vitals | 0 | | | | | vitals |
| **spo2_sd** | Oxygen Saturation Ratio (SD) | continuous | Standard deviation of the measured oxygen saturation ratios during this patient's hospitalization | Percent | Vitals | 0 | | | | | vitals |
| **systolic_25** | Sytolic BP (25th Percentile) | continuous | Systolic blood pressure at the 25th percentile of this patient's measurements | mmHg | Vitals | 0 | | | | | vitals |
| **systolic_75** | Sytolic BP (75th Percentile) | continuous | Systolic blood pressure at the 75th percentile of this patient's measurements | mmHg | Vitals | 0 | | | | | vitals |
| **systolic_count** | Systolic BP (Count) | continuous | Count of systolic blood pressure measurements for this patient | Number of measurements | Vitals | 0 | | | | | vitals |
| **systolic_first** | Systolic BP (First) | continuous | First systolic blood pressure measurement for this patient's hospitalization | mmHg | Vitals | 0 | | | | | vitals |
| **systolic_last** | Systolic BP (Last) | continuous | Last systolic blood pressure measurement for this patient's hospitalization | mmHg | Vitals | 0 | | | | | vitals |
| **systolic_max** | Systolic BP (Max) | continuous | Maximum systolic blood pressure for this patient's hospitalization | mmHg | Vitals | 0 | | | | | vitals |
| **systolic_mean** | Systolic BP (Mean) | continuous | Average systolic blood pressure for this patient's hospitalization | mmHg | Vitals | 0 | | | | | vitals |
| **systolic_median** | Systolic BP (Median) | continuous | Median systolic blood pressure for this patient's hospitalization | mmHg | Vitals | 0 | | | | | vitals |
| **systolic_min** | Systolic BP (Min) | continuous | Minimum systolic blood pressure for this patient's hospitalization | mmHg | Vitals | 0 | | | | | vitals |
| **systolic_sd** | Systolic Bp (SD) | continuous | Standard deviation of the systolic blood pressures during this patient's hospitalization | mmHg | Vitals | 0 | | | | | vitals |
